# Harnessing methods, data analysis, and near-real-time wastewater monitoring for enhanced public health response using high throughput sequencing

**DOI:** 10.1101/2024.12.10.24318772

**Authors:** Padmini Ramachandran, Tunc Kayikcioglu, Tamara Walsky, Kathryn Judy, Jasmine Amirzadegan, Candace Hope Bias, Bereket Tesfaldet, Maria Balkey, Dietrich EppSchmidt, Hugh Rand, James Pettengill, Sandra Tallent, Eric Brown, Tina Pfefer, Ruth Timme, Amanda Windsor, Christopher Grim, Maria Hoffmann

## Abstract

Wastewater-based analysis has emerged as a pivotal method for monitoring SARS-CoV-2 (SC2). Leveraging high-throughput sequencing on wastewater samples facilitates a comprehensive, population-level assessment of circulating and emerging SC2 variants within a community. This study meticulously evaluates the detection performance, variant calling accuracy, and the time taken from sample collection to public data release for wastewater SC2 monitoring. We employed two different SC2 target enrichment panels on Illumina MiSeq and Oxford Nanopore Technologies (ONT) GridION sequencing platforms for a robust analysis. Daily collection of routine raw grab and composite samples took place at a wastewater treatment plant (WWTP) site in Maryland, USA (MD) from mid-January 2022 to the end of June 2022. Total Nucleic Acid (TNA) was extracted from samples and target enrichment was executed using QIAseq DIRECT and NEBNext VarSkip Short amplicon kits, with subsequent sequencing on MiSeq or ONT GridION platforms, respectively. Obtained sequences was analyzed using our custom CFSAN Wastewater Analysis Pipeline (C-WAP). Raw sequence data and detailed metadata were submitted to NCBI (BioProject PRJNA757291) as it became available. Our wastewater data successfully detected the onset of new variants BA.2, BA.2.12, BA.4.6, and BA.5 to the observed population. Notably, Omicron sub-variants were identified approximately a week ahead of publicly available clinical data at the MD ZIP-code level. Variation in quality metrics paralleled the rise and fall of BA waves, underscoring the impact of viral load on sequencing quality. Regular updates of estimated variant proportions were made available on the FDA-CFSAN “Wastewater Surveillance for SARS-CoV-2 Variants” website. In contrast to the median 28-day turnaround for our samples, the lead time from sample collection to public release of raw sequence data via NCBI was remarkably swift, accomplished within a mere 57 hours in this specific exercise. Our processing, sequencing, and analysis methods empowered the swift and accurate detection of SC2 trends and circulating variants within a community, offering insights for public health decision-making.

## Introduction

Clinical variant determination of SARS-CoV-2 (SC2) relies on the detection of genomic elements of SARS-CoV-2 by reverse transcription-quantitative polymerase chain reaction (RT-qPCR) based methods from individual patients (1, 2). Clinical analyses are now also being complemented with antibody-based assays that provide an indication of current or previous exposure to SC2 (3). High-throughput sequencing technologies are being used to sequence the SC2 genome from a subset of the infected population (4, 5). This has resulted in a large number of published genomes, and has provided insight into its origins, spread, evolution, and diversity via computational approaches in genomic epidemiology (1). However, inferring lineage prevalence by clinical testing is infeasible at scale, especially in areas with limited testing and/or sequencing capacity or limited community participation, both of which can introduce sampling biases (4).

Utilizing wastewater samples to identify pathogenic human viruses, including SC2, has gained attention as a method for understanding population-level trends in infections (3). SARS-CoV-2 RNA concentration in wastewater has been shown to convey regional infection dynamics and provides less biased abundance estimates than clinical testing (1, 3). As SC2 continues to spread and evolve, early detection of emerging variants is critical for orchestrating public health interventions.

There are many factors limiting wastewater-based genomic surveillance of pathogens. For example, despite the promising success of a few studies (6, 7), it is still challenging to understand how well wastewater-based epidemiology can identify the genetic diversity of SARS-CoV-2 in each population and how this relates to known viral diversity of clinical cases. This is especially important as new variants emerge (1). Many challenges also lie in the accurate identification of SC2 variants and estimation of their abundance in mixed population samples such as wastewater. While deep sequencing without prior amplification may be efficient for viruses that reside in high quantity in plasma, in a mixed population of variants and a complex environmental matrix like wastewater, sequencing without amplification is impractical (8). For wastewater-based viral detection, implementation of a multiplexed target enrichment panel is necessary. However, there are challenges in the analysis of sequencing reads from target enrichment panels for accurate identification of multiple variants in a single sample. Here we use an analysis pipeline, the CFSAN Wastewater Analysis Pipeline (C-WAP, https://github.com/CFSAN-Biostatistics/C-WAP), for identification and abundance estimation of circulating variants. As SC2 evolves and new variants arise, continued monitoring for emerging variants will be an important component of public health efforts (9). As the virus evolves, enrichment kits, databases of variants, and detection methods will also need to evolve and be re-evaluated to ensure the best possible methods are in use.

The application of high-throughput sequencing for the rapid identification and surveillance of pathogens has recently become common place in public health systems (10–12). The SC2 pandemic has helped bring WBE to the forefront of community-scale pathogen surveillance (7, 13). Here, we describe how monitoring wastewater from urban areas can be used to detect the arrival of emerging SC2 variants of SC2 in a chosen sewer shed. This study adapted two different target enrichment panels, initially designed for clinical samples, for application in wastewater samples. Additionally, both protocols were optimized and published to protocols.io (14). The study established critical quality control checkpoints within the laboratory workflow and on the analysis of sequence data derived from a mixed population of wastewater samples. The evaluation encompassed the detection performance and variant calling accuracy using two different SC2 target enrichment panels, covering 99.16 +/− 0.58% of the SC2 genome conducted on both Illumina MiSeq and Oxford Nanopore Technologies (ONT) GridION sequencing platforms. A crucial aspect explored was the turn-around-time from sample collection to public data release, recognizing its significant public health impact. The findings demonstrated the effectiveness of high-throughput sequencing of SC2 genomes from wastewater. The incorporation of precision analysis tools, such as C-WAP, facilitated the early detection of emerging variants of concern in wastewater samples.

In summary, this study emphasizes the need for community-based surveillance, complemented by rigorous and comprehensive analysis and investigation. The thorough analysis when assessing the performance of enrichment panels employed here, considering the interplay between the enrichment panel itself and the specific sequencing technology, is extremely valuable. This study utilized a comprehensive wastewater analysis pipeline, crafted for real-time surveillance of mixed population samples that took into consideration several aspects of targeted amplicon-based sequencing, implemented strict QC measures to avoid false positives, critically assessed emerging variants by looking at the characteristic mutations, and evaluated several genome coverage metrics across the genome. The insights gained from this study offer valuable lessons applicable to wastewater surveillance methodologies targeting emerging pathogens.

## Materials and Methods

### Sample collection

Wastewater samples were collected from a wastewater treatment plant in Maryland from January 12, 2022, to June 28^th^, 2022. The raw wastewater was collected daily from January 12^th^ to February 28^th^, 2022 as two 1-liter raw grabs (2 biological reps) in Nalgene bottles and stored at 4°C. The samples were retrieved weekly and subsequently stored at 4°C until use. From January 12 to February 28th, 2022, raw wastewater grab samples were extracted and analyzed as individual samples with 2 biological replicates for each day resulting in 14 samples per week.

Composites were collected as 1L grabs from Feb23^rd^ 2022 till June 28^th^, 2022. Composites were aliquoted from the auto sampler (auto sampler has a collection interval of 30 mins for 24hrs) as two-1L grabs in Nalgene bottles and stored at 4°C until use. Each biological replicate per week after pooling was then split into three 40mL technical replicates, resulting in six samples per week (Supplementary table 1, Schematic Fig 1). The complete workflow for this study is described in detail in Fig 1.

**Figure 1:**
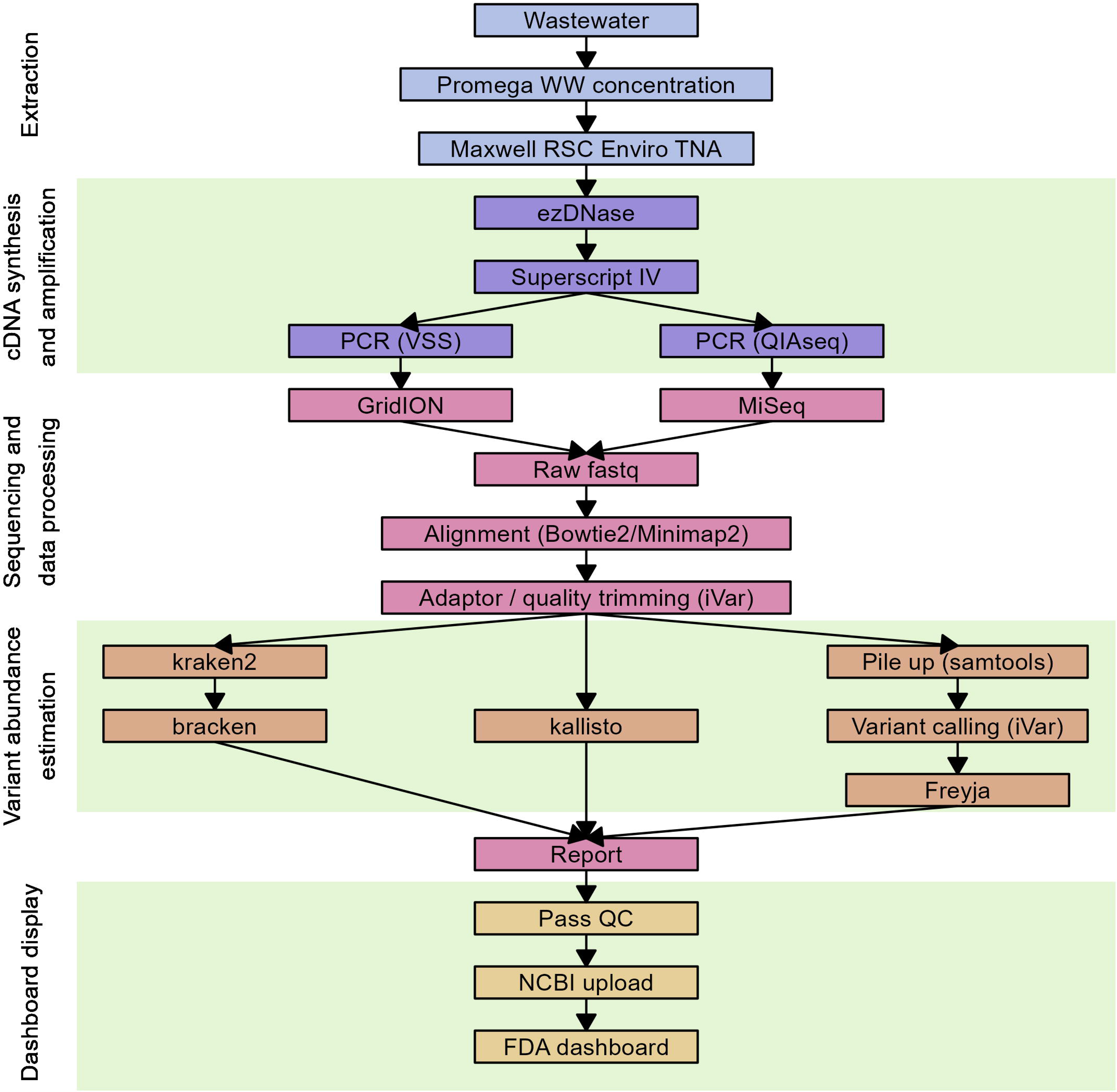
Flowchart of wastewater processing and analysis workflow: Wastewater samples collected from the treatment plant were pooled by week and the total nucleic acid (TNA) was extracted using Promega Enviro TNA kit. Genomic DNA was digested with ezDNase prior to cDNA synthesis using Superscript IV Reverse Transcriptase. The cDNA was amplified using either NEBNext VarSkip Short (VSS) primers for long-read (∼550bp) sequencing on the Oxford Nanopore Technologies GridION or QIAseq DIRECT (QIAseq) primers for short-read (∼250bp) sequencing on the Illumina MiSeq. All sequences were processed through the CFSAN Wastewater Analysis Pipeline, producing a report that included QC metrics as well as relative abundance of variants. Sequences that pass QC are submitted to NCBI and the variant calls were published in the FDA dashboard.

**Schematic Fig 1:**
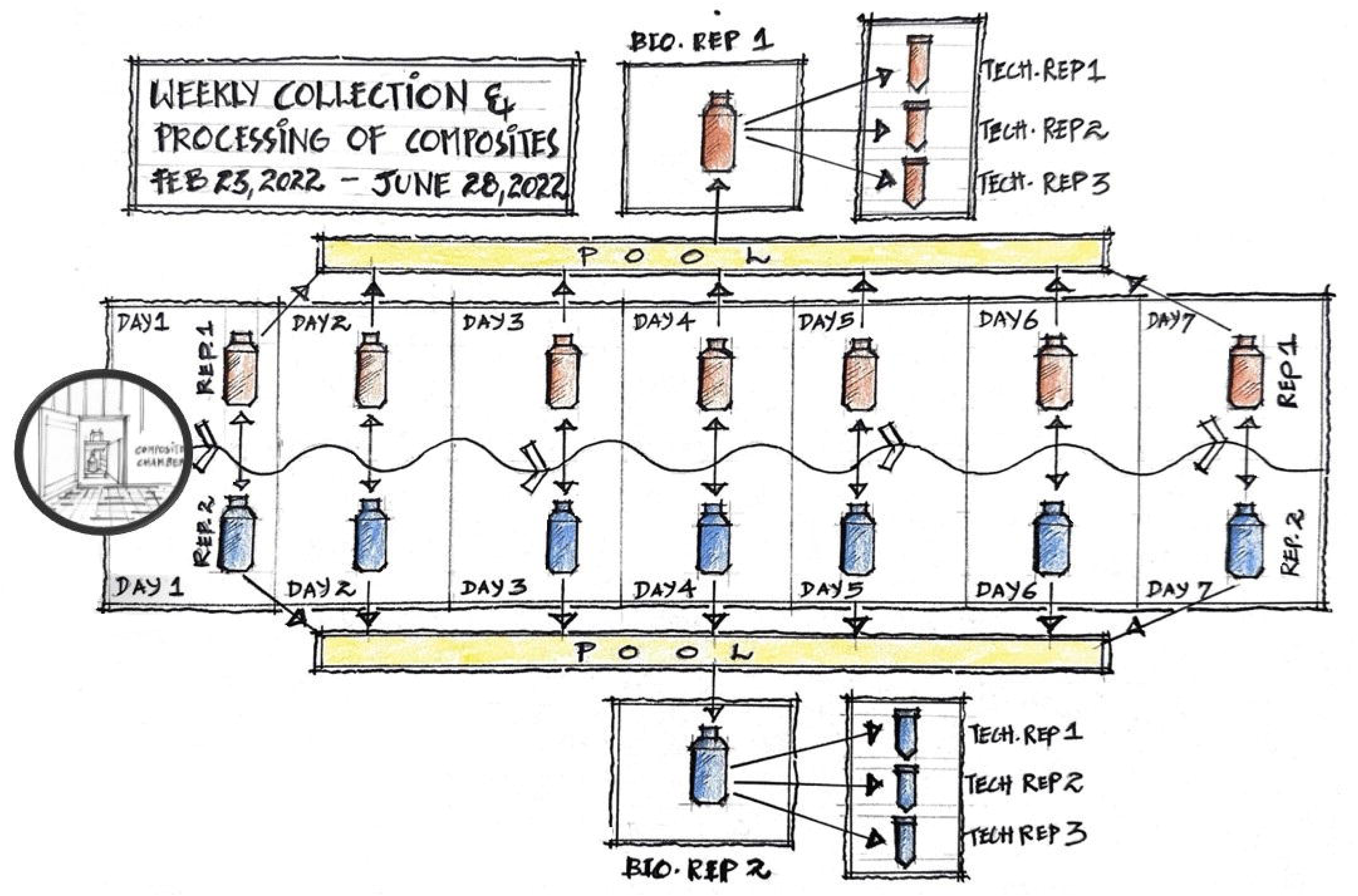
Weekly collection and processing of composite samples: From February 23rd through June 30, 2022, daily composite samples were pooled into a single sample of two biological replicates per week. Each biological replicate was then split into three 40mL technical replicates, resulting in six samples per week that was taken into further processing.

### Total nucleic acid extraction and SARS-CoV-2 RT-qPCR detection

Sample concentration and total nucleic acid extraction was performed on the pooled wastewater using Maxwell RSC Enviro TNA Kit (Promega, Madison, WI USA) following manufacturer’s recommendations. The extracted total nucleic acid was stored at −20 C for short term (a week) to process for the cDNA generation and RT-qPCR. After the necessary amount was aliquoted, the TNA was stored at −80 C for long term storage. The detailed protocol has been published in <u>dx.doi.org/10.17504/protocols.io.rm7vzy52xlx1/v1</u>.

The quantification of SARS-CoV-2 in wastewater was performed utilizing the Promega GoTaq® Enviro Wastewater SARS-CoV-2 System, which targets the N1 gene (Promega, Madison, WI USA) following manufacturer’s recommendations. This includes guidance for converting RT-qPCR output into calculated viral genome copies per liter.

### DNA digestion and cDNA synthesis

Genomic DNA was digested with ezDNase (Invitrogen, Waltham, MA USA) with protocol modifications. Reaction cocktails consisted of 10uL template TNA, 2µL nuclease free water, 1µL ezDNase™ buffer, and 1 µL ezDNase™ enzyme. Reactions were then digested at 37°C for 5 minutes. Digestion was stopped by addition of 1µL 10mM DTT and a 5-minute incubation at 55°C. Reactions were chilled on ice for at least 1 minute before proceeding to cDNA synthesis.

cDNA was synthesized from the purified RNA with the Superscript IV Reverse Transcriptase (Invitrogen, Waltham, MA, USA) with modifications to the manufacturer protocol. Primer annealing was carried out with 1µL 50µM random hexamers, 1uL 10mM dNTP mix, 1µL DEPC-treated water, and 10µL template RNA. Reactions were incubated at 65°C for 5 minutes, then incubated on ice for at least 1 minute. The Reverse Transcriptase reaction mix was prepared with 4µL 5x SSIV Buffer, 1 µL 100mM DTT, 1µL RNaseOUT™ Recombinant RNase Inhibitor, and 1uL SuperScript IV Reverse Transcriptase (200U/µL) and then added to the chilled primer-annealed RNA. Reactions were then incubated at 23°C for 10 minutes, 50°C for 30 minutes, then 80°C for 10 min. cDNA was stored at −20°C until use.

### Selection of enrichment panels for this study

October through December 2021, we evaluated some commercially available SC2 enrichment panels. Enrichment panels for assessment were chosen based on multiple criteria. Our main objective was to ensure even coverage across the genome; therefore, we chose kits with tiled amplicon across the entire SC2 genome and simple library preparation of these amplicons to achieve rapid, simple, and cost-effective sequencing. Read lengths of the amplicon to adapt to both short read sequencing chemistry and long read sequencing chemistry were also considered. The commercial enrichment panels that were assessed initially included Midnight FREED primers (1200 bp long amplicons with Oxford Nanopore library and sequencing) (15), SWIFT SNAP additional genome coverage panel (150 bp long amplicons with Illumina MiSeq sequencing) (16), NEBNext VarSkip Short primers (560 bp long, with possibility of sequencing in both Illumina and ONT platform) (17), and QIAseq DIRECT enrichment panel (250 bp amplicons with Illumina MiSeq sequencing). Based on the ease of generating the amplicons, ease of library preparation and adaptability of the amplicons with different sequencing platforms we chose two enrichment panels for this study that are discussed in detail in this manuscript, QIAseq DIRECT and NEBNext VarSkip Short (NEB VSS). The QIAseq DIRECT panel has 222 primer pairs with average amplicon length of 250 bp while the NEB VSS panel has 74 primer pairs with an amplicon length of about 560 bp, on average.

### QIAseq DIRECT SARS-CoV-2 enrichment panel

cDNA amplification was performed using QIAseq DIRECT SARS-CoV-2 enrichment panel following manufacturer’s protocols. After the emergence of variant BA.5, (June 13^th^, 2022), the manufacturer distributed an auxiliary primer set as a spike-in to the existing protocol. The samples from May 17^th^ and onward were amplified using the spike-in to the enrichment panel. The detailed protocol is publicly available at **dx.doi.org/10.17504/protocols.io.rm7vzy39rlx1/v4**

### NEB VSS SARS-CoV-2 enrichment panel

cDNA amplification was performed utilizing the NEB VSS SARS-CoV-2 enrichment panel following manufacturer’s protocol. Due to the emergence of BA.2.12, there was a change in the primer version from VSS v1a to VSS v2a starting from the week of March 1st. There was also a spike-in introduced in the primer pairs as an update to the kit due to the emergence of BA.5. The samples from the week of May 11^th^ and onward were amplified using the enrichment panel with spike-in. The detailed protocol is publicly available at **dx.doi.org/10.17504/protocols.io.3byl4bwervo5/v2**

### High throughput Sequencing

QIAseq DIRECT amplicons were sequenced using Illumina MiSeq (Illumina, San Diego, CA). NEB VSS amplicons were sequenced using Oxford Nanopore GridION sequencing (Oxford Nanopore Technologies, Oxford, UK). Every sample (n = 182) was amplified using both QIAseq DIRECT SARS-CoV-2 enrichment panel and NEB VSS SARS-CoV-2 panel (n=364). The libraries of NEB VSS amplicons were prepared using ligation sequencing kit (Oxford Nanopore Technologies Ligation Sequencing Kit (SQK-LSK109), Cambridge, UK) as described in the publicly available protocol **dx.doi.org/10.17504/protocols.io.3byl4bwervo5/v2** run on a R.9.4 flow cell (R9.4.1) for 72Qh.

### Data analysis, quality metrics and variant calling

To address the many challenges in accurate identification of multiple variants, we further customized our analysis tool CFSAN Wastewater Analysis Pipeline (C-WAP)(9). The latest version of C-WAP is now available on GitHub https://github.com/CFSAN-Biostatistics/C-WAP. Briefly, sequences are aligned to the reference genome (Wuhan-Hu-1, NCBI assembly ASM985889v3, NC_045512.2) using Bowtie2 (v2.4.5) and Minimap2 (v2.24), followed by adapter trimming and quality filtering in iVar (v1.3.1). When processing Illumina data, the iVar trim function was configured to discard the adaptor sequences; filter sequences below a quality score of 20 (default); filter reads that are shorter than 50% of the average length of the first 1000 reads (default); and use a 4 bp sliding window (default). For ONT data, iVar trimming is configured the same, with the exception that all sequences are retained (quality score filter of 1), effectively preserving the filtering of reads to a quality score of 7 performed by Guppy, the default intrinsic base caller onboard ONT MinKNOW sequencing software.

C-WAP provides three methods for variant calling: Kraken2 (v2.1.2) / bracken (v2.7), kallisto (v0.48), and a Samtools (v1.15) pileup piped into Freyja (v1.4.4). Bracken is a Bayesian abundance estimator based on Kraken2 sequence calls. A comprehensive list of dependencies for C-WAP is available in the GitHub readme.

C-WAP takes into account the number of raw reads that align to the reference sequence and pass filter, *i.e.*, read lengths after adaptor trimming ≥30 and minimum read quality ≥20 within a sliding window of width 4 bases. C-WAP outputs a comprehensive report along with several distinct quality metrics (Table 1). These include the percent of reads aligned to SARS-CoV-2 genome, coverage depth of the sequencing reads at every genomic coordinate of the SARS-CoV-2 genome, read quality, coverage depth over the entire breadth of the SARS-CoV-2 genome, absolute counts of genomic coordinates with less than 10X coverage or no coverage, scaled counts of genomic coordinates that with less than 10X coverage or no coverage, and variant abundance estimation using Freyja, kallisto, and Kraken.

**Table 1:** QC metrics monitored for data submission and further analysis.

Characteristic mutations for several variants, including variants B.1.617.2, BA.1, BA.2, BA.3, BA.4, and BA.5 (18), the variants of concern most relevant to the timeframe of this study, were extracted from the C-WAP report (Supplementary table 2). The table is generated under the assumption that the presence of a variant requires the detection of all mutations of a particular variant. The characteristic mutations which support the presence of a particular variant are indicated in the respective column of the table. A detected mutation table with detection frequency and p-values for every alternate base in each position is generated where only the genomic coordinates with at least 10X coverage were considered.

### Concordance correlation coefficient

A concordance correlation coefficient (CCC) with Euclidean distance was used to evaluate the levels of pairwise agreement among the abundance estimation methods for both amplicon panels, NEB VSS and QIAseq DIRECT. It was adapted by Cui et al. (19) from Lin’s concordance correlation coefficient for agreement studies with microbiome compositional data. CCC has values between −1 and 1, where −1 indicates a perfect disagreement, 1 indicates a perfect agreement, and 0 indicates a complete absence of agreement between the two abundance estimation methods. Bayes-Laplace Bayesian-multiplicative replacement method (20) is used to impute zero relative abundances on the entire dataset before computing CCC values. A bootstrap method with sample size of 5,000 is used to build a 95% confidence interval for each CCC estimate.

To understand the agreement between the variant callers on real world data, in the amplicon panels used in this study, for the CCC analysis a subset of (n= 11) samples were chosen. The sample chosen for this analysis were samples collected on the week of May 5^th^, 2022 (6 samples) and the week of June 28^th^, 2022 (5 samples). The 11 samples/22 sequences (with each sample being amplified with NEB VSS and QIAseq DIRECT and sequenced in ONT and Illumina MiSeq respectively) were chosen based on sequence reads having uniform coverage across the genome, good breadth versus depth metric and higher percent covid hits. For the week of May 5th samples, the amplicon panels lacked the spike-in primers. In this instance, the basic panel successfully identified the circulating variants at the time. For the June 28th samples, both amplicon panels were equipped with updated spike-in primers capable of detecting the prevailing variant circulating at that time.

### Dashboard display

Sequencing data that passed QC assessment (category A and B, Table 1) were submitted in real time to National Center for Biotechnology Information (NCBI) for active monitoring for circulating variants. To facilitate the sharing of information about progress on this sequencing effort, FDA developed a dashboard that graphically presented findings from this project (21). Active support and updates to the dashboard ended on June 30, 2023, the final state of the dashboard remains available at https://www.fda.gov/food/whole-genome-sequencing-wgs-program/wastewater-surveillance-sars-cov-2-variants.

### Data visualization

Downstream data analysis and visualization were carried out in RStudio (v.1.3.1093) using the following R packages: ggplot2 (v3.4.1), dplyr (v1.1.0), reshape2 (1.4.4), ggh4x, and stringr (v1.5.0).

### Data availability

Data generated in this study were deposited in the NCBI BioProject database under the BioProject accession number PRJNA757291.

## Results

### SARS-CoV-2 viral load and relationship to clinical cases

The viral load of SARS-CoV-2 in wastewater was assessed throughout the time course of this study to understand the impact of viral burden levels and dynamics on amplicon sequencing success and output. Computed viral load, calculated from RT-qPCR on samples taken from the wastewater treatment plant, ranged from 350 genome copies/L to 99,840 genome copies/L, with fluctuations that mirrored variant waves (Fig 2). To understand the relationship of wastewater-based surveillance burden estimations and actual clinical reporting, clinical data were plotted against the viral genome copies/L over time (Fig 2, Supplementary table 3). The rolling 7-day average of new clinical SARS-CoV-2 cases for the two ZIP codes (21157 and 21158) served by the wastewater treatment plant was extracted from MD department of Health website, https://opendata.maryland.gov/Health-and-Human-Services/MD-COVID-19-Cases-by-ZIP-Code/ntd2-dqpx/data_preview. When compared to the reported clinical cases, which ranged anywhere between 1 case per week to 866 cases per week, we observed a seemingly lower rate of clinical testing from March 2022 onward (Fig 2).

**Figure 2:**
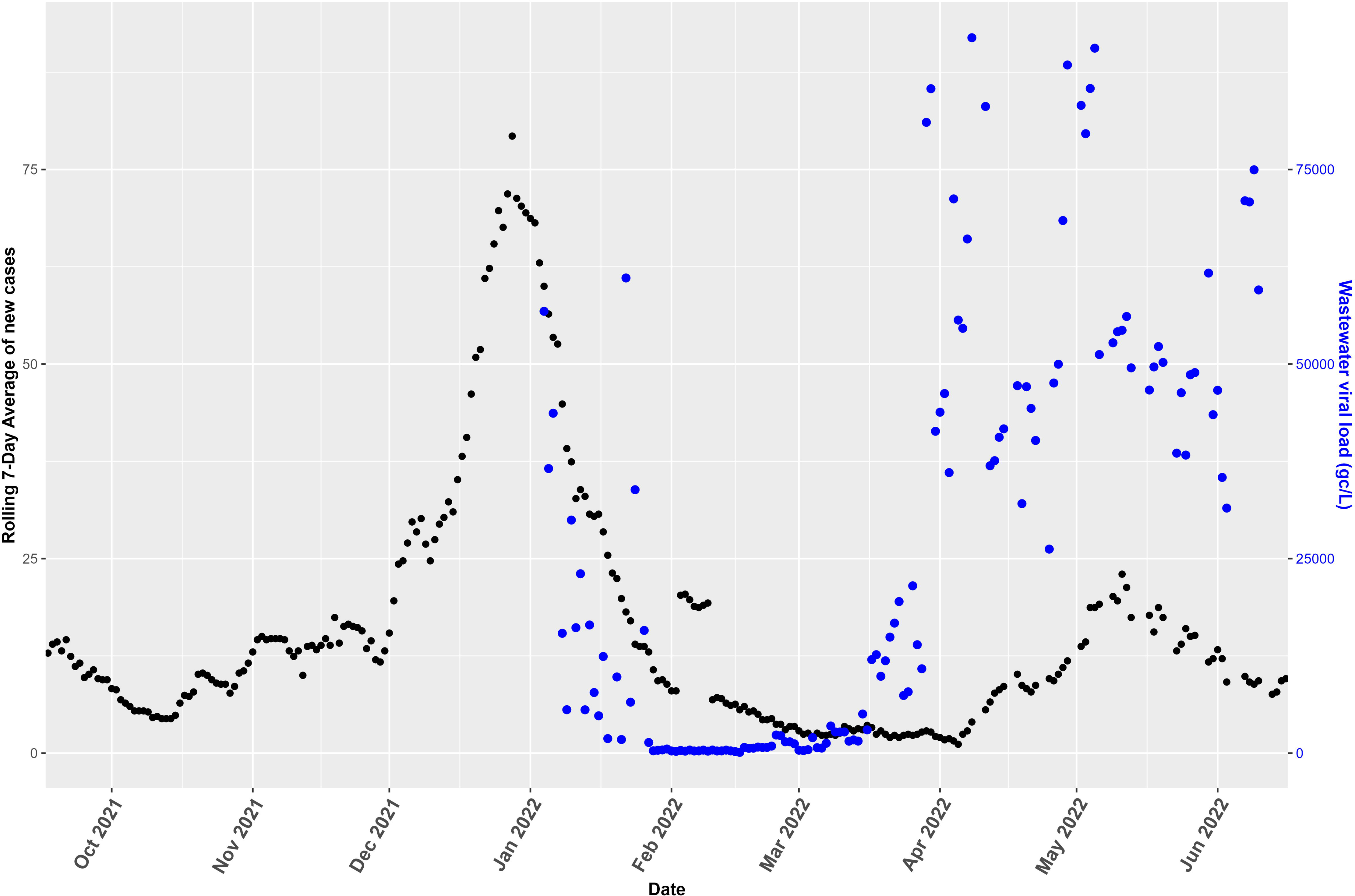
SARS-CoV-2 viral load and relationship to clinical cases: The relationship of wastewater-based surveillance burden estimations and actual clinical reporting, with clinical data plotted against the viral genome copies/L, over time. The rolling 7-day average of new clinical SARS-CoV-2 cases (black) in the sewer shed graphed alongside the viral load over time (blue), calculated from RT-qPCR on raw wastewater and 24h composite samples taken from the wastewater treatment plant. Number of cases is indicated on the right-hand axis, while genome copies/L is marked on the left.

### Percent reads aligned vs genome copies of SARS-CoV-2

Weekly average percent reads aligned to the SC2 genome using both enrichment panels (n=364) were compared against the weekly average SC2 genome copies per liter of wastewater detected by RT-qPCR (Fig 3). The average percent reads aligned to the SARS-CoV-2 genome from weekly sampling roughly corresponds to the genome copies/L in wastewater from the corresponding time period, emphasizing the dependence of high quality SC2 genome amplicon sequencing on circulating viral load. The lowest percent reads aligned were observed during February and March 2022 when SARS-CoV-2 was relatively scarce, but percent reads aligned increased beginning in April, with the surge of the Omicron variants, through the end of the monitoring period for this study, June 2022.

**Figure 3:**
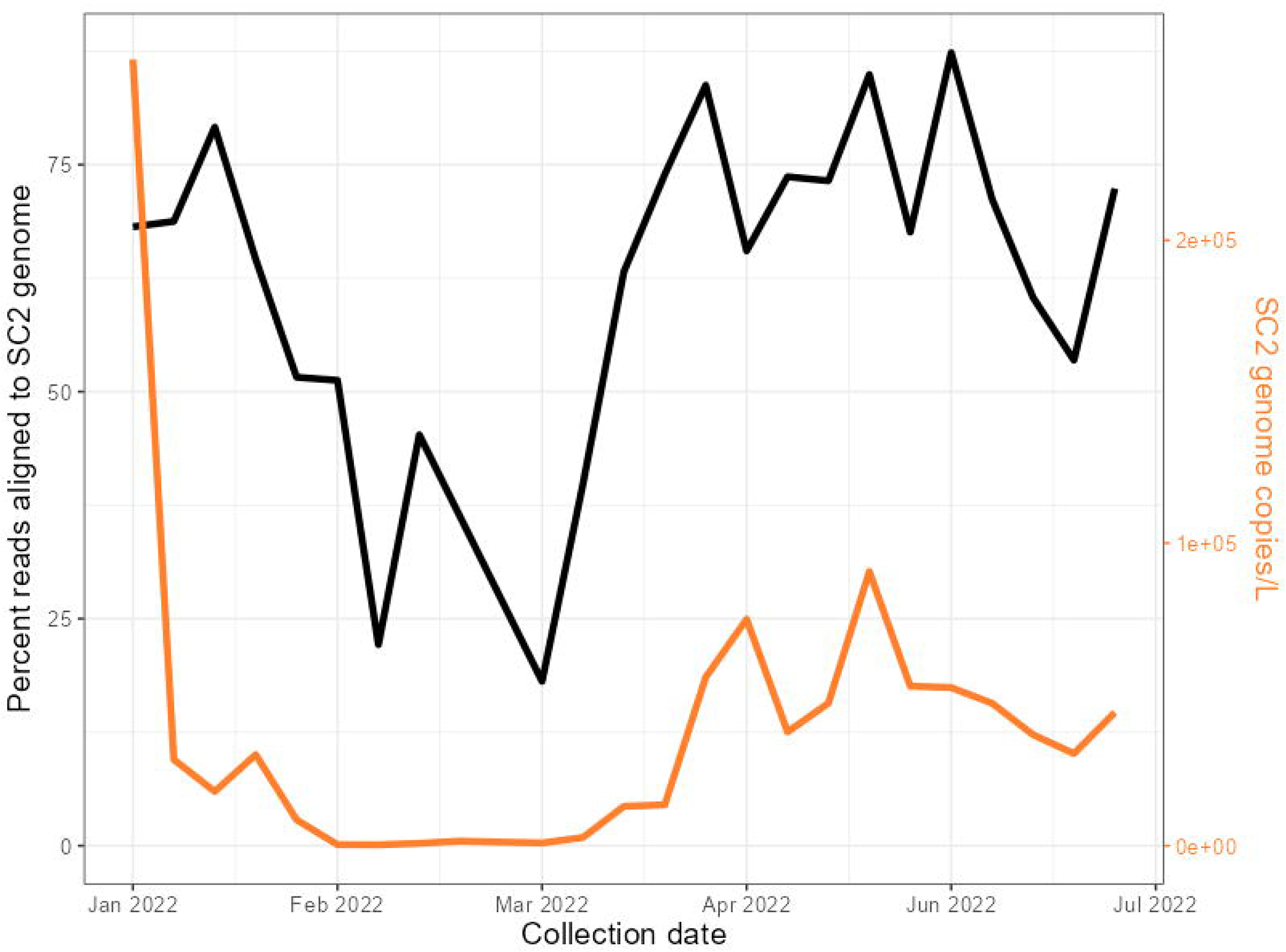
Percentage of aligned reads vs. wastewater viral load : Weekly average percent reads aligned to the SC2 genome (black) and the weekly average SC2 viral load (gc/L) detected by RT-qPCR from wastewater (orange). Sequencing runs/data from both enrichment panels (NEB VSS and QIAseq DIRECT, n=364) included in the averaging to calculate the weekly percent reads aligned were quality filtered to < 20% sites with 0x coverage and < 40% sites covered <10x after alignment.

### Comprehensive QC metric analysis

To characterize sequencing performance, C-WAP outputs a comprehensive report with several distinct quality metrics including the aforementioned percent reads aligned to the SC2 genome, coverage depth of the sequencing reads at every genomic coordinate of the SC2 genome, read quality, read counts for each primer pair of the amplicon panel, coverage depth over the entire breadth of the SC2 genome, absolute counts of genomic coordinates with less than 10X coverage or no coverage, scaled counts of genomic coordinates that with less than 10X coverage or no coverage, and variant abundance estimation using Freyja, kallisto, and Kraken (Supplementary report 1, 2). The following sections includes detailed results of the metrics considered by C-WAP for the samples discussed in the study.

### Coverage depth vs coverage breadth

We compared sequencing depth versus breadth of the SC2 genome for both amplicon kits, QIAseq DIRECT and NEB VSS. The ideal sequencing run would have the maximum sequencing depth (greatest number of reads aligned) over the entire breadth (100%) of the genome, indicating even coverage over all genomic coordinates. Across the time course of this study, the coverage depth versus breadth for each amplicon kit used for each month of wastewater surveillance (Fig 4) improved as we optimized our sample collection and sequencing methods, with values for the metric converged towards the ideal in June compared to January. Comparison of the amplicon panels shows that the NEB VSS panel has lower genome coverage when compared to the QIAseq DIRECT panel for January and February, but by June the breadth versus depth for all sequencing runs are similar between amplicon kits.

**Figure 4:**
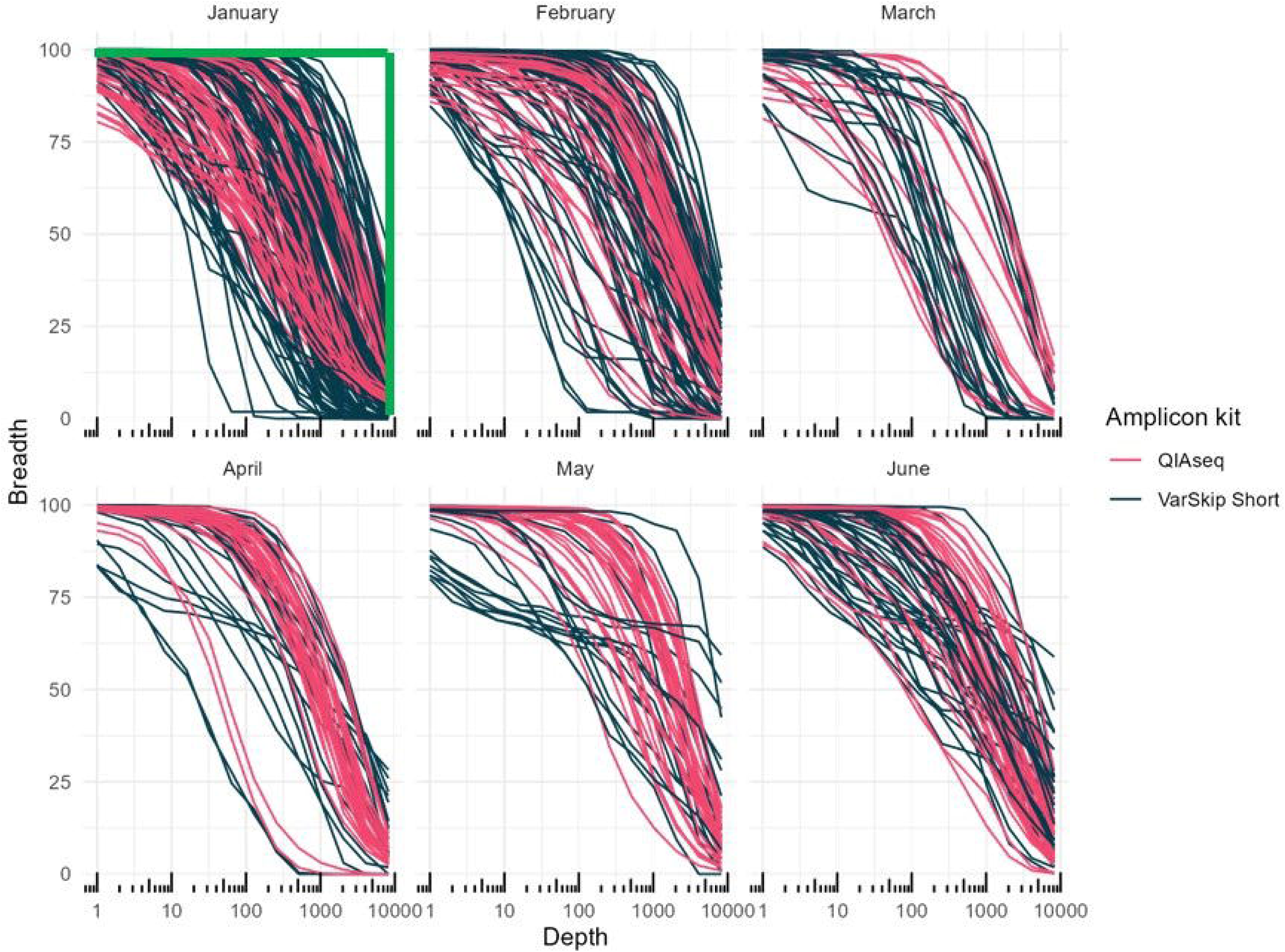
Coverage depth vs coverage breadth: Sequencing depth versus breadth of the SARS-CoV-2 genome covered for each amplicon kit and every month of wastewater surveillance. Any point along a given line indicates the percentage (breadth, y-axis) of the SARS-CoV-2 genome covered to the corresponding sequence depth (x-axis) for that single sample. The green bar on the first panel of January 2022, shows an ideal sequencing run with maximum depth covering the entire genome. Each line indicates one sequenced sample that resulted in <20% sites along the genome covered 0x and <40% sites covered <10x after alignment.

### Genome coverage across ORF1ab, S, M, and N genes of SC2 genome

Calculations of absolute and scaled coverage for each nucleotide site within all SC2 genes (5’ UTR, ORF1ab, S, ORF3a, E, M, ORF6, ORF7ab, ORF8, N, ORF10, 3’UTR) was extracted from the C-WAP report. Genome coverage for four key genes, namely ORF1ab, S, M and N genes, was used to detect characteristic mutations of the variants from January through June 2022 (Fig 5) during this study. Both NEB VSS and QIAseq DIRECT enrichment panels had more under-covered (<10x) sites within the S gene when compared to coverage of the remaining three key genes. 255 samples enriched by QIAseq DIRECT showed under-coverage in the S gene, versus an average of 166 samples for the other three key genes, while 109 samples enriched by NEB VSS showed S gene under-coverage, versus an average 92.3 samples? for the other three genes. An overall poor performance of QIAseq DIRECT on the M gene was observed across all months of sampling, when compared to NEB VSS, with the average scaled M gene under-coverage across all QIAseq DIRECT samples 117% greater than that of NEB. In contrast, the NEB VSS amplicon panel was observed to have more under-covered sites on ORF1ab and N genes across all months of sampling—78% and 13% greater average scaled under-coverage, respectively.

**Figure 5:**
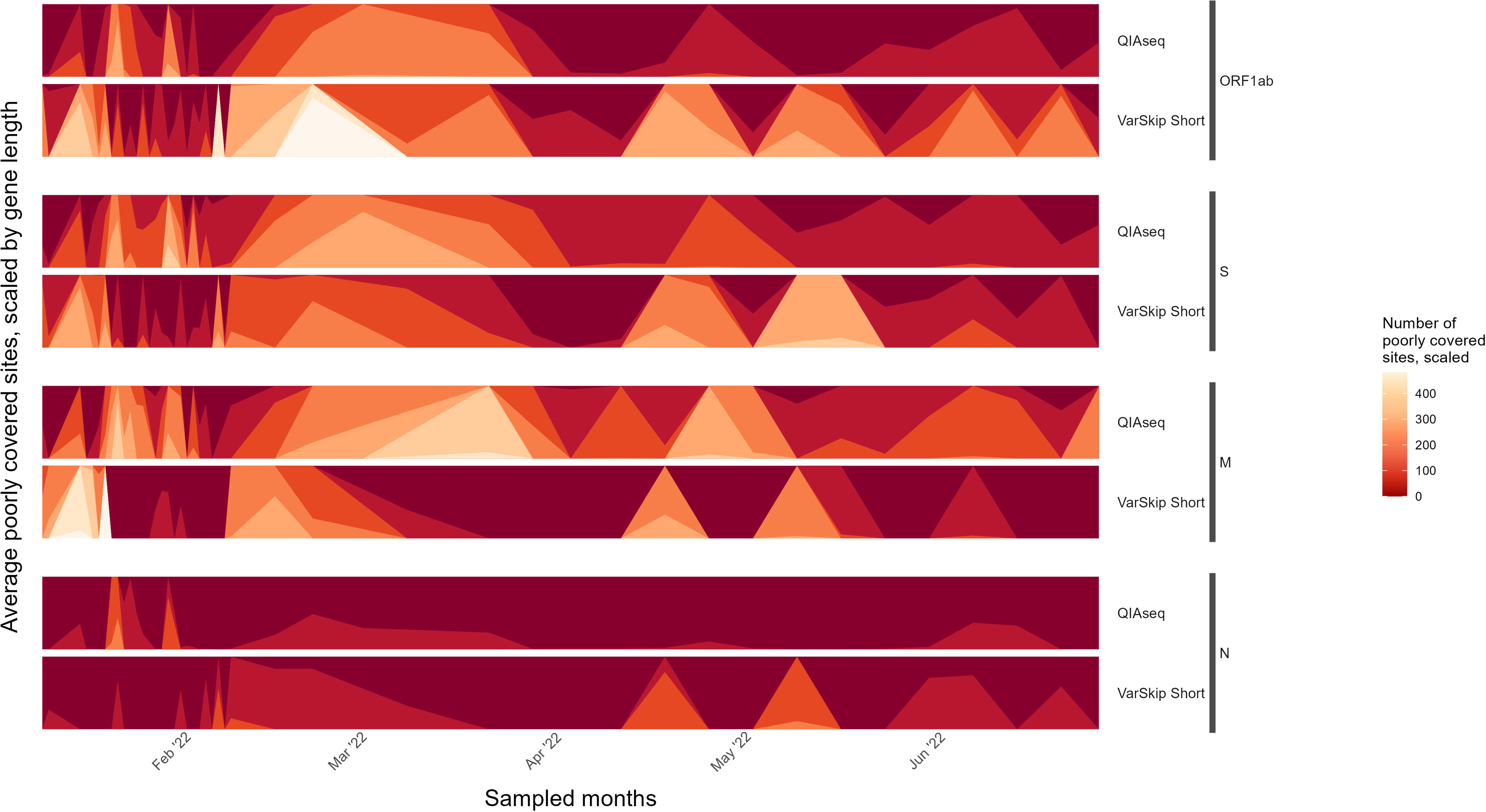
Genome coverage across ORF1ab, S, M, and N genes of SC2 genome: Genome coverage by amplicon kit for key genes used to detect SARS-CoV-2 variants from January through June 2022. The averaged number of under-covered (fewer than 10 quality-filtered and trimmed reads aligned) for each gene and amplicon kit over time is indicated in red, with worse coverage (poorer sequencing performance) in lighter colors.

### Concordant Correlation Coefficient

To further assess performance and agreement between the two amplicon panels and the variant callers used in this study, we computed concordant correlation coefficients (CCC) to compare the performance of the top two variant callers (9), Freyja and kallisto, using a subset of high-quality sequenced samples, determined by having good QC metrics with uniform coverage across the genome (Fig 6). The CCC was positive for each kit, though under 0.25 for each, indicating mild agreement between the variant callers (Fig 6). The CCC between the variant callers was relatively higher for QIAseq DIRECT samples (0.2113 as opposed to 0.0102 for NEB VSS) and had a 95% confidence interval that did not span 0, indicating a statistically significant agreement. In contrast, the CCC for NEB VSS samples had a 95% confidence interval spanning 0, indicating there is likely no relationship between the variant detection calls by Freyja and kallisto. Accordingly, this suggests that for NEB VSS samples with GridION sequencing, one variant caller may be a better choice than the other to produce the most accurate results, while for QIAseq DIRECT samples with MiSeq sequencing, the choice of variant caller may be less important.

**Figure 6:**
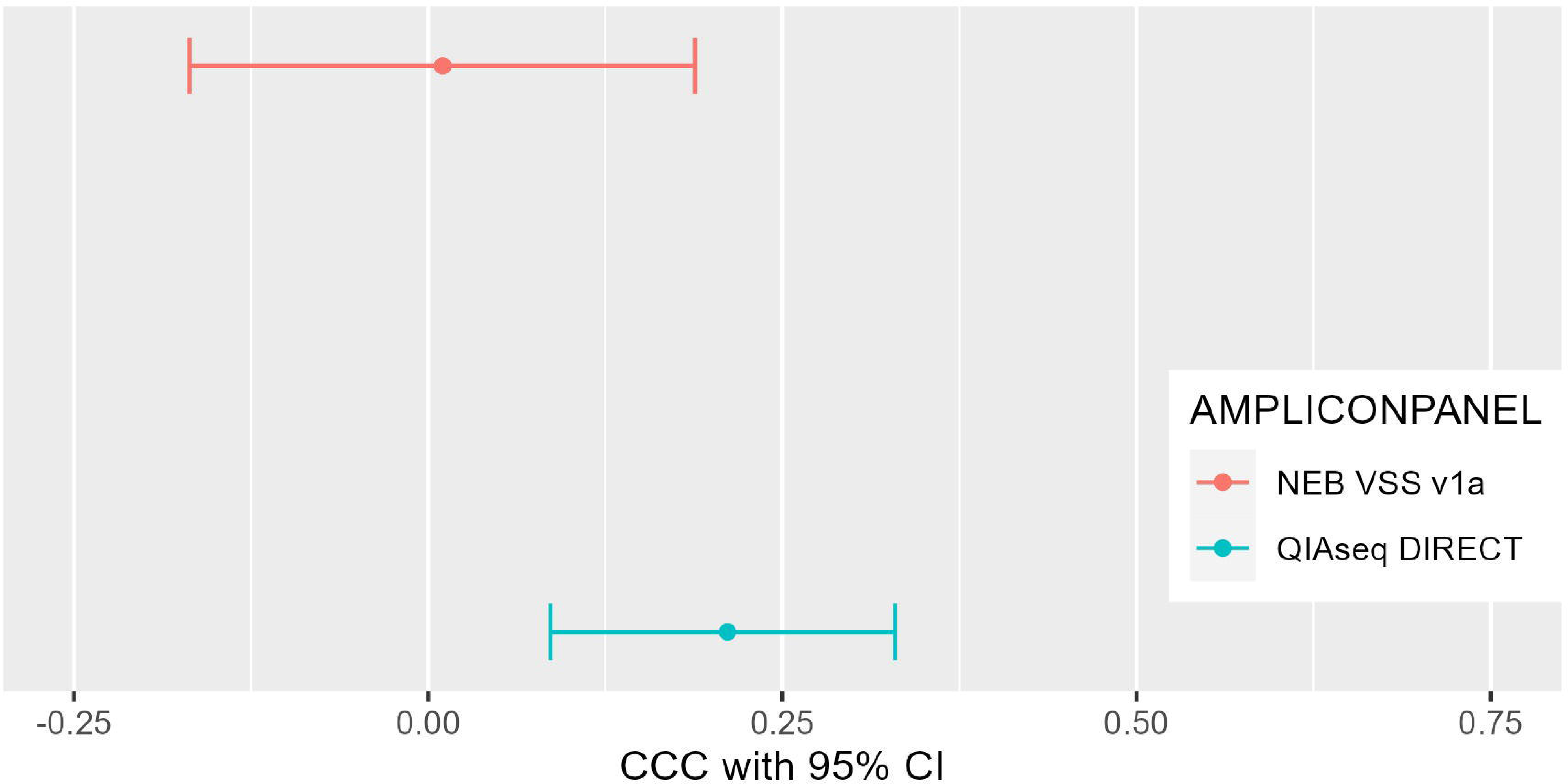
Concordant Correlation Coefficient (CCC) analysis: concordant correlation coefficient analysis on the variant calling performance of Freyja and kallisto on eleven samples between May and June 2022 sequenced with both NEB VarSkip Short and QIAseq DIRECT primers. A CCC of +1 indicates perfect agreement, −1 indicates perfect disagreement, and 0 indicates no agreement.

### Circulating variant estimation

During the sampling period, our surveillance system not only captured the major and minor variants circulating within the community but also detected the emergence of new variants. We combined the variant results from all 364 sequences from 182 samples, agnostic to the kit and sequencing platform used, to observe the general trend of circulating variants using Freyja as its database is updated more frequently. The MD wastewater samples clearly identified the emerging variants of concern like BA.1, BA.2.12, BA.4.6 and BA.5 from January 2022 through June 2022 (Fig 7, Supplementary Fig 1). Omicron BA.1 was the dominant circulating variant when the wastewater sampling began in January 2022. January 29^th^ samples showed signs of emergence of BA.2, and dominant detection of BA.2 was observed from March 30^th^, 2022. BA.2.12 was detected starting from end of March 2022, followed by BA.4 and BA.5 in mid-May, and dominantly detected starting end of May 2022. We also observed traces of BA.4.6 in mid to late June, agreeing with the onset of BA.4.6 in clinical cases in Maryland.

**Figure 7:**
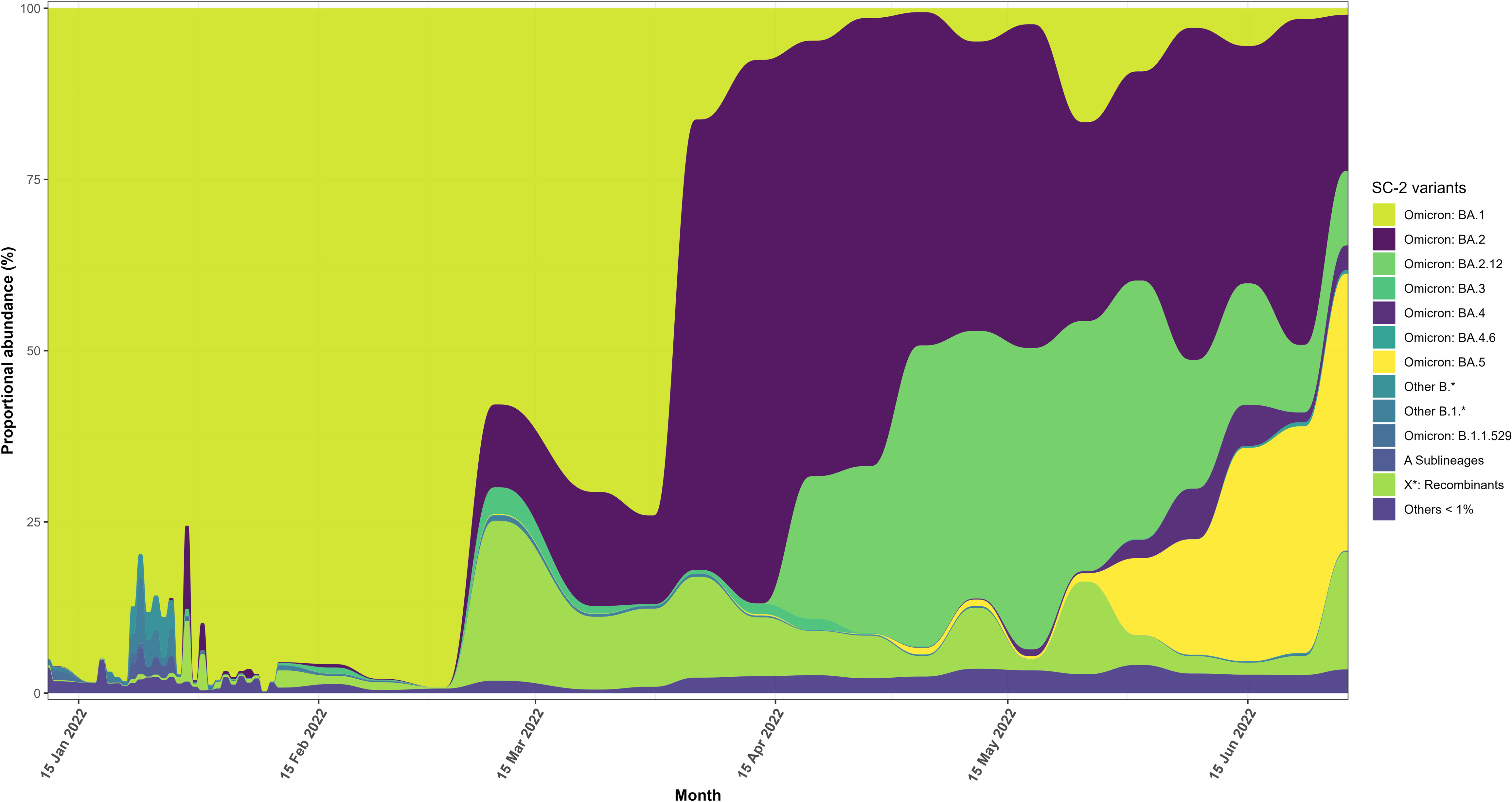
Proportional abundance of SARS-CoV-2 variants detected over time: Proportional abundance of SARS-CoV-2 variants detected over time, as identified by the Freyja variant caller on sequence data from both the enrichment panels (NEB VSS and QIAseq DIRECT, n=364). Variants always detected in a proportion of less than 1% were grouped together as “Others <1%”.

### Early detection of BA.2

The BA.2 lineage was first detected in samples collected on January 29^th^, 2022, and again on January 31, 2022 with both the NEB VSS and QIAseq DIRECT enrichment panels. The first clinical reporting of the BA.2 lineage in the entire state of MD by the CDC was on January 27^th^, 2022 (variant data is not available per zip code). To verify the early detection of BA.2 in these wastewater samples, every mutation identified using both enrichment panels was examined for six days (January 27^th^– February 1^st^) surrounding the first purported appearance of BA.2 (Fig 8A). Detected mutations for each day were categorized based on their presence/absence in BA.1 and BA.2 lineage sequences. Mutations shared between BA.1 and BA.2 in the characteristic mutation table generated by C-WAP were marked as “BA.1 or BA.2” mutations. Identified mutations that are not present in BA.1 were categorized as “Incompatible with BA.1” and mutations only present in BA.2 were categorized as “BA.2 diagnostic mutations.” All mutations detected were considered statistically significant (p-value < 0.05) and present in at least 10 sequencing reads.

**Figure 8:**
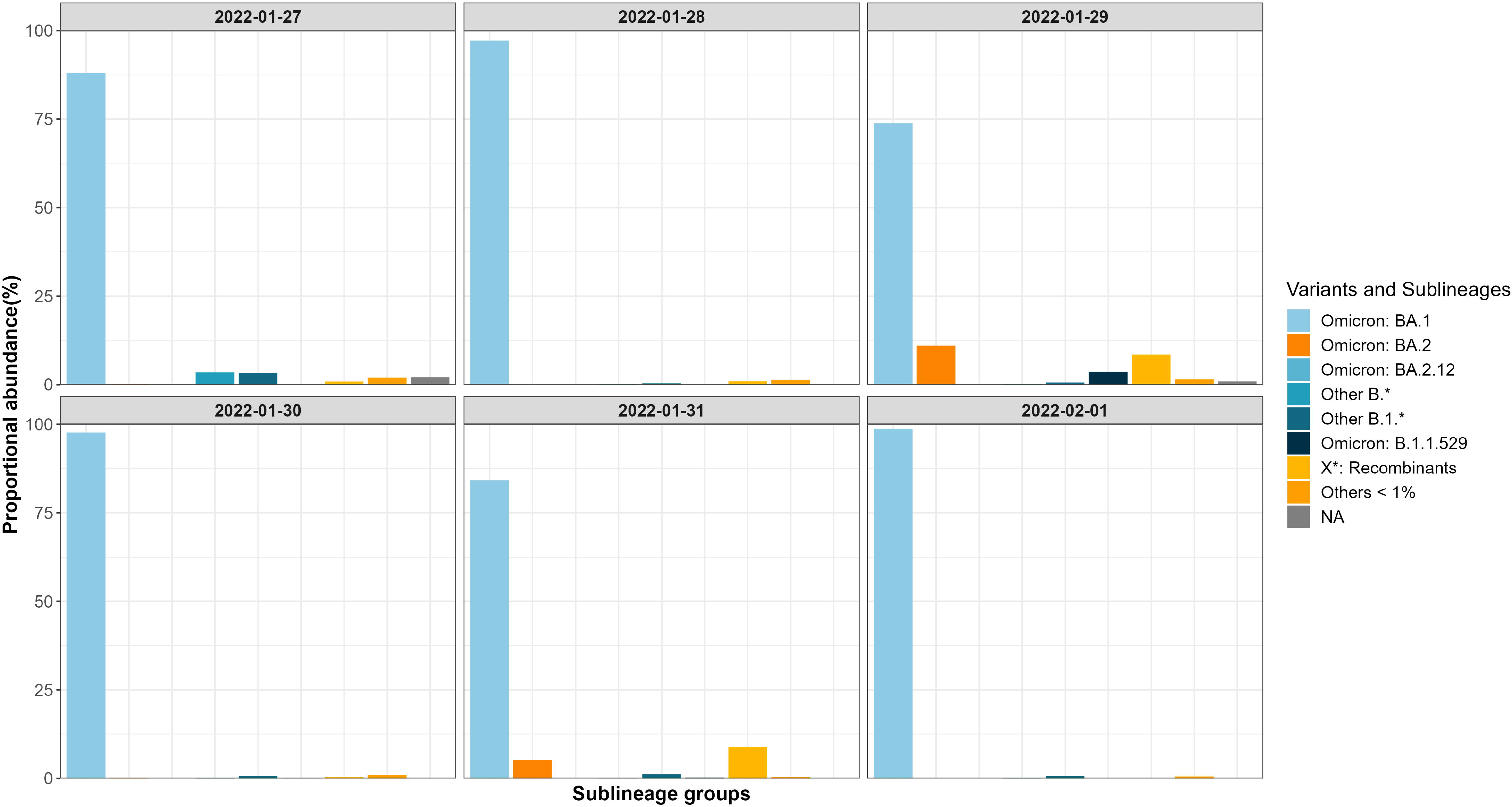
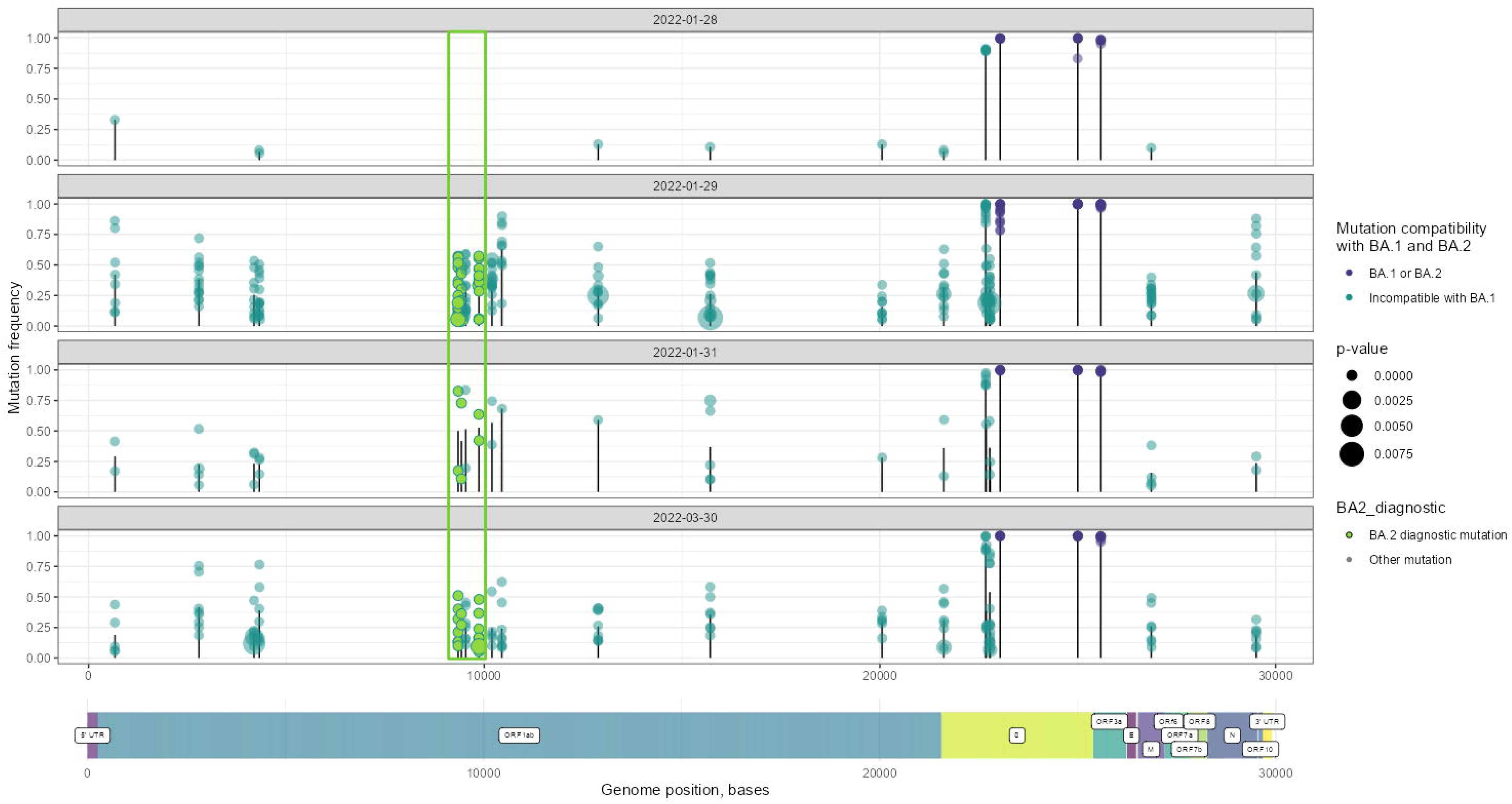
Verification of the BA.2 variant detection in wastewater on January 29^th^, 2022, using both the enrichment panel (NEB VSS and QIAseq DIRECT): 8A. Proportional abundance of SARS-CoV-2 variants by day, January 27^th^ – February 1^st^ 2022. 8b. Frequency of BA.1 and BA.2 –characteristic mutations over four dates. Mutations typically incompatible with BA.1 (teal), characteristic for BA.1 or BA.2 (purple), and diagnostic for BA.2 (green, in a highlighted box) are shown at their location in the SARS-CoV-2 genome. All depicted mutations are significant (p-value < 0.05), but smaller circles indicate lower p-values. BA.2 was detected in the bottom three date panels.

The prevalence of these three mutation categories on both enrichment panels on each of four days in January and in March 2022, when BA.2 was well established, was visualized by showing the frequency of each mutation and its place in the SARS-CoV-2 genome (Fig 8B). The landscape of relevant mutations on both early BA.2-detection days (January 29^th^ and 31^st^) is highly similar to the pattern of mutation prevalence and detection in March, when BA.2 was consistently present in both wastewater and clinical samples. BA.2 diagnostic mutations were detected with frequencies up to at least 0.5 in some samples, while most detected BA.1-incompatible mutations were present with frequency of 0.25 or greater.

In contrast, the mutations detected on January 28 include no BA.2 diagnostic mutations and BA.1-incompatible mutations are largely present in low frequency. This shows the presence and frequency of BA.2 diagnostic mutations and the overall profile of mutations observed at the early detection dates, January 29^th^ and 31^st^, mimic that from a later sample, from March 30^th^, when BA.2 was known to be present in the WWTP service area.

### Realtime submission of surveillance data

After the sequencing protocol was optimized and QC metrics were in place, we made an effort to quantify the minimum time from sample collection to sequence analysis and release. Composite grabs for the week of June 28^th^, 2022, were collected at 6 am from the treatment plant and the time to process these samples was recorded. Total nucleic acid extraction, cDNA synthesis, amplification using NEB VSS enrichment panel, and loading of the libraries on to Oxford Nanopore GridION took 30hrs. After 16hrs, we had collected sufficient sequence data that passed QC to detect all the characteristic and diagnostic mutations for BA.5. Data analysis using C-WAP, upload of this data to NCBI, and display on the dashboard took 11 hrs. The total turnaround time from sample collection to dashboard display was 57hrs.

## Discussion

Six months of wastewater surveillance, made possible through the partnership with a wastewater treatment plant in Maryland, detected new variants as they emerged along with major and minor variants circulating in the community. From a clinical perspective, this level of resolution of community disease dynamics could underpin traditionally monitored clinical testing and help public health officials to serve communities more efficiently. A wastewater surveillance system fills data gaps for areas where clinical testing and surveillance may be limited, including underserved communities with limited access to testing sites.

The laboratory and bioinformatic methods employed in this study confidently detected multiple circulating SARS-CoV-2 (SC2) variants in wastewater samples using amplicon-based targeted enrichment panels. Several enrichment panel kits were evaluated, but only two, the QIAseq DIRECT and NEB VSS panels were pursued as the best options for wastewater samples.

The viral load in wastewater, when compared to the clinical cases, indicated a low rate of clinical testing as the pandemic began to wane (Fig 2). In other words, there were higher levels of SC2 in the community, based on wastewater analysis, than were being estimated based on reports from clinical testing. This is perhaps because at-home test kits became widely available within the community around the same time as this study commenced. From wastewater data, it is evident that clinical testing did not fully capture the wave of infections during early April to June 2022 (Fig 2). A limit of detection with genome copies/L to the SC2 hits was not established and so all samples were sequenced regardless of RT-qPCR result (22). Having strict QC measure in place helped us to overcome this lack of a robust RT-qPCR LOD threshold and eliminate some poorly performing samples attributed to the lower genome copies/L.

A main goal of wastewater surveillance is to track emerging and circulating variants at a community level and hopefully provide an early warning signal of emerging and increasing infections without the need for community clinical information. For example, we detected BA.2 ahead of the clinical findings for the state. It is important to note that the variant calls are published only at the state level by CDC, not at the ZIP code level. Here we detected BA.2 at the community level covering two ZIP codes 14 days ahead of BA.2 variant calls published by CDC at the state level for Maryland. This emphasizes an ability of community level surveillance using wastewater: enabling a more focused public health local response during a pandemic.

In SC2 variants, mutations across the S-gene play a crucial role in variant calling. Research is ongoing to elucidate the mechanisms involved but the mutations may boost infectivity and transmissibility through increased spike density, enhanced cleavage and host cell uptake, and increased viral load and ability to evade host immune responses (23). Mutations may also interfere with laboratory testing and complicate epidemiological monitoring by interfering with detection in sequence-based tests. If a mutation occurs in the part of the viral genome assessed by a PCR test, the sample may result in gene “dropout.” We observed a significant drop in coverage of the S gene with the NEB VSS panel on the week of May 11. For this same week, RT-qPCR of these samples had 90,587 genome copies/L indicating a high SC2 viral load in the community (the range of genome copies for the duration of this study was from 350 genome copies/L to 99840 genome copies/L). The significant drop in S gene coverage raised concerns and the vendor was contacted and spike-in to the primer pool 1 was obtained to overcome the dropout due to the rise of BA.5 sub variants. When we repeated the amplification and sequencing of samples from the week of May 11 using the primer spike-in (NEB VSS), we observed improved coverage on the S gene because of the three characteristic mutations (S:S371F, S:T376A, S:D405N) detected with the spike-in that were not detected with the original primer panel.

A diagnostic mutation of BA.5 in the M gene (M:D3N) was first detected the week of May 11 with QIAseq DIRECT enrichment panel. This same mutation was not detected until the week of June 1 using NEB VSS enrichment panel. A spike-in for the QIAseq DIRECT enrichment panel was introduced from the week of June 13. The spike-in primers in the QIAseq DIRECT panel had a slightly higher concentration of primers compared to its original panel across this region (23082-23144 bp). While there was not a full dropout in this region (23082-23144 bp), the observed coverage was lower, and the concentration of the primers (QIAseq_170_RIGHT, QIAseq_172_LEFT, and QIAseq170-2_RIGHT) were increased to provide more uniform coverage within that gene. Since mutations may affect detection, treatment, and prevention, it is important to identify strains correctly to direct public health containment strategies (5). To address this type of limitation, there are studies that incorporate targeted sequencing of only variable regions of interest in the genome, particularly those regions that contain mutations unique to specific variants of concern (5). In addition, there are studies that apply PCR amplifying, cloning, and sequencing a 1.5Qkb region of the spike protein gene to confirm the linkage of mutations of interest to understand all the circulating variants in a complex sample (24). While these approaches provide valuable information on a very targeted genomic area where mutations are known to occur, mutations at other genomic coordinates, which might be indicative of emergent variants and variants, will not be obtained.

In practice, when a diagnostic or surveillance test is based on targeted amplification, for it to be able to detect variations in the genome of pathogens, it is necessary to regularly update the primer pairs and/or alter the target regions used in the detection systems or use several target genes of the infectious agent in the same test reaction to minimize the risk of false negative results (5). Both enrichment panels evaluated in this study consistently updated the primer pairs for the detection of circulating variants. In an ARTIC primer approach where entire primer versions are updated, the emergence of new variants renders any remaining primers obsolete (25, 26). This stands in contrast to the spike-in approach, where the spike-in is introduced to the original pools, minimizing the regeneration of old primers and revitalizing their performance. Many enrichment panels contain degenerate primer designs of polymorphic regions of the genome to allow more robust amplification of variable strains. Also, strategically designed primer pairs for consistent coverage are essential for highly evolving genomes like SC2 (5). Keeping the challenges of uniform coverage within the targeted site, primer binding efficiency, and adaptation to several sequencing approaches in mind, choosing a suitable target enrichment panel and pairing it with the suitable library preparation kit based on the sequencing platform may play a critical role in identifying the circulating and emerging variants.

When we assessed genome coverage metrics for four key genes (ORF1ab, S, M and N genes), both QIAseq DIRECT enrichment panel and NEB VSS enrichment panel had substantial under-covered regions on the S gene when compared to the other three genes. In the NEB VSS SARS-CoV-2 enrichment panel, there are 10 primer pairs targeting the S gene (genome coordinates:21563-25384) with an overlapping primer pair included to enable detection of BA.5 specific mutations. The S gene is the most mutation-prone region of SC2 genome, which means that primer binding sites for this gene can play a major role in variant calling performance on a mixed sample like wastewater. For the M gene, the QIAseq DIRECT enrichment panel had lower coverage across all months when compared to NEB VSS enrichment panel (Fig 5). Two characteristic mutations within the M gene used to assess all Omicron sub variants correspond to primer binding regions in the QIAseq DIRECT enrichment panel, while neither mutation is in a primer binding region for NEB VSS. The QIAseq DIRECT enrichment panel has more amplicons with more primer pairs (222 primer pairs total, whereas NEB VSS panel has 74 primer pairs) and is more suitable for short read sequencing (∼250bp amplicon length) which results in high coverage across the genome. But there is a greater chance of mutations occurring in a primer binding site, especially for rapidly evolving genes, such as the S gene. The NEB VSS primer panel somewhat mitigates this with longer amplicon length (∼560bp) and fewer primer pairs. Continuous evaluation of current and emerging target enrichment panels is essential to accurately detect existing and evolving SC2 variants from wastewater.

During the development of C-WAP, a critical performance assessment of various variant callers on *in silico* reads of mixed variants revealed that Freyja was the most suitable variant caller on short read data and kallisto was the most suitable variant caller for the long read data (9). The concordant coefficient analysis on our real-world data agreed with those previous findings (Fig 6) However, to solidify these conclusions, further investigations are warranted, particularly with a larger sample size. Additionally, considering the evolving nature of viruses, future studies should explore the impact of different amplification kits on variant calling accuracy.

We tested the feasibility of the real time submission of data to support the public health response. Real time submission and publication to the dashboard within 57 hours was possible because of real-time long read sequencing through Oxford Nanopore GridION. After just 16 hrs of sequencing, there was enough accumulated sequencing depth data that passed QC to provide high confidence in the accuracy of variant calling. Even though the run time in GridION was chosen to be 72hrs, most of the data were collected in the initial 20hrs after sequencing started. However, to ensure high quality data, we multiplexed only 9 samples per flow cell on GridION, compared to 28-32 possible samples on Illumina MiSeq. The Illumina MiSeq, despite its inherent speed limitations, presents promising prospects for near-real-time applications when Read1 data is being used. While just the Read1 data can successfully pass all quality control (QC) metrics, there is a subsequent delay of up to 24 hours for uploading the sequence data from a paired-end run, which might be advantageous for maximizing quality scores. It is important to understand the complexity associated with multiplexing samples per flow cell, a factor that may necessitate adjustments based on the viral load. Our team has established best practices for the efficient collection, organization and surveillance of sequence data (21). Analyzing the turnaround time from sample collection to NCBI data release revealed a median duration of approximately 28 days for the samples in this study. However, this exercise has illuminated the potential for real-time data submission, enabling immediate surveillance insights. Regardless of the sequencing technology, near real time data submission is possible through wastewater surveillance which will support community level decision making easier during pandemic. Incorporation of automated liquid handling robots could further reduce time from sample receipt to data publication.

Due to the dynamic and swiftly evolving nature of SARS-CoV-2, our dedication to real-time dashboard updates and the dissemination of optimized protocols to the wastewater surveillance community, this study has some limitations. One limitation was the inability to delve into the genuine distinctions in raw wastewater and composites beyond a one-week sample analysis period where we explored both raw wastewater grabs (February 23^rd^ to February 28^th^) and composite grabs (February 23^rd^ to February 28^th^). We needed to investigate the feasibility of weekly sample pooling as a strategy (February 28^th^, 2022, and onwards) to reduce the processing workload in our laboratory, and when we did not find any differences in the viral load and other QC metrics when the sequences, regardless of enrichment panel, were analyzed from that week, we proceeded with weekly sample pooling. Additionally, we were constrained in exploring the potential benefits of multiplexing varying sample quantities on GridION flow cells to comprehend sequencing depth. This study is also limited by not including an evaluation of other commercially available enrichment panels, such as ARTIC V4 or Resende (26, 27). Moreover, the study was hindered in its capacity to investigate spike-in studies with known concentration of the virus in various matrices, which are essential for establishing a true limit of detection.

The two amplicon panel kits demonstrated robust efficacy in detecting both circulating and emerging variants in wastewater samples. The two panels used in this study underwent sequencing on two distinct platforms, and to ensure accurate assessment, we established customized quality control (QC) metrics using C-WAP. Regarding variant calling, both enrichment panels successfully detected the presence of BA.2 on January 29^th^, 2022, samples and consistently detected other circulating variants throughout the study duration. The comprehensive analysis employed in this study emphasizes the importance of evaluating the performance of any enrichment panel in conjunction with the chosen sequencing technology.

## Conclusion

We supported the public health response effectively during the pandemic by constant method optimization, updates to the wet lab protocol, updates to the databases for the variant calling, comprehensive analysis, pairing up the enrichment panel with suitable sequencing technology, appropriate variant callers used throughout the surveillance, and the efficient updates of a dashboard. Finally, as seen here, our findings highlight the value of extensive data analysis metrics, method optimization, and a near-real-time public health response. These insights can serve as valuable considerations for decision-making in the establishment of new surveillance initiatives, especially those involving mixed samples and complex sample types such as wastewater. In essence, this study provides a blueprint for thoughtful consideration of various metrics when conducting surveillance using target enrichment panels, offering valuable lessons that can inform the development of future surveillance studies.

## Supporting information

Supplementary report 1

Supplementary report 2

## Abbreviations

SC2: SARS-CoV-2
WWTP: Wastewater treatment plant
TNA: Total Nucleic Acid
C-WAP: CFSAN Wastewater Analysis Pipeline

## Acknowledgements

We gratefully acknowledge the wastewater treatment plant for collecting samples for this study and sharing it with us. We acknowledge Dr. Leena Malayil and Dr. Amy Sapkota, School of Public Health, University of Maryland for introducing us to the WWTP for sample collection. We acknowledge CovidTrakr working groups at the Center for Food Safety and Applied Nutrition (CFSAN) for discussion that facilitated this manuscript. We also acknowledge the support of CFSAN high performance computing engineers G. Engelbach, J. Payne, K. Konganti, and M. Hammond. We acknowledge all data contributors for generating the genetic sequence and metadata and sharing via the GISAID Initiative, on which this research is partly based.

## Author Contributions

PR performed sample collection, lab experiments, data interpretations and wrote and edited the manuscript. TK developed C-WAP and performed data analysis and interpretation. PR, AW, CG and MH conceived and designed the study and performed laboratory experiments. KJ performed lab experiments and data interpretation. CB, TP, MB, ST and RT developed the dashboard. EB retrieved samples from WWTP. BT performed concordant correlation coefficient analysis. JA, DE, JP and HR performed data analysis. All authors reviewed and edited the manuscript and approved the final version.

---

Supplementary table 1: Sample collection and pooling strategies employed in this study.

Supplementary table 2: Characteristic mutation table by C-WAP.

Supplementary table 3:Rolling 7 day average of new cases for zip codes monitored in this study along with the wastewater viral load (gc/L).

Supplementary Figure 1: SC2 population trend by sampling date using both the enrichment panel and Freyja as the variant caller.

**Supplementary reports:**
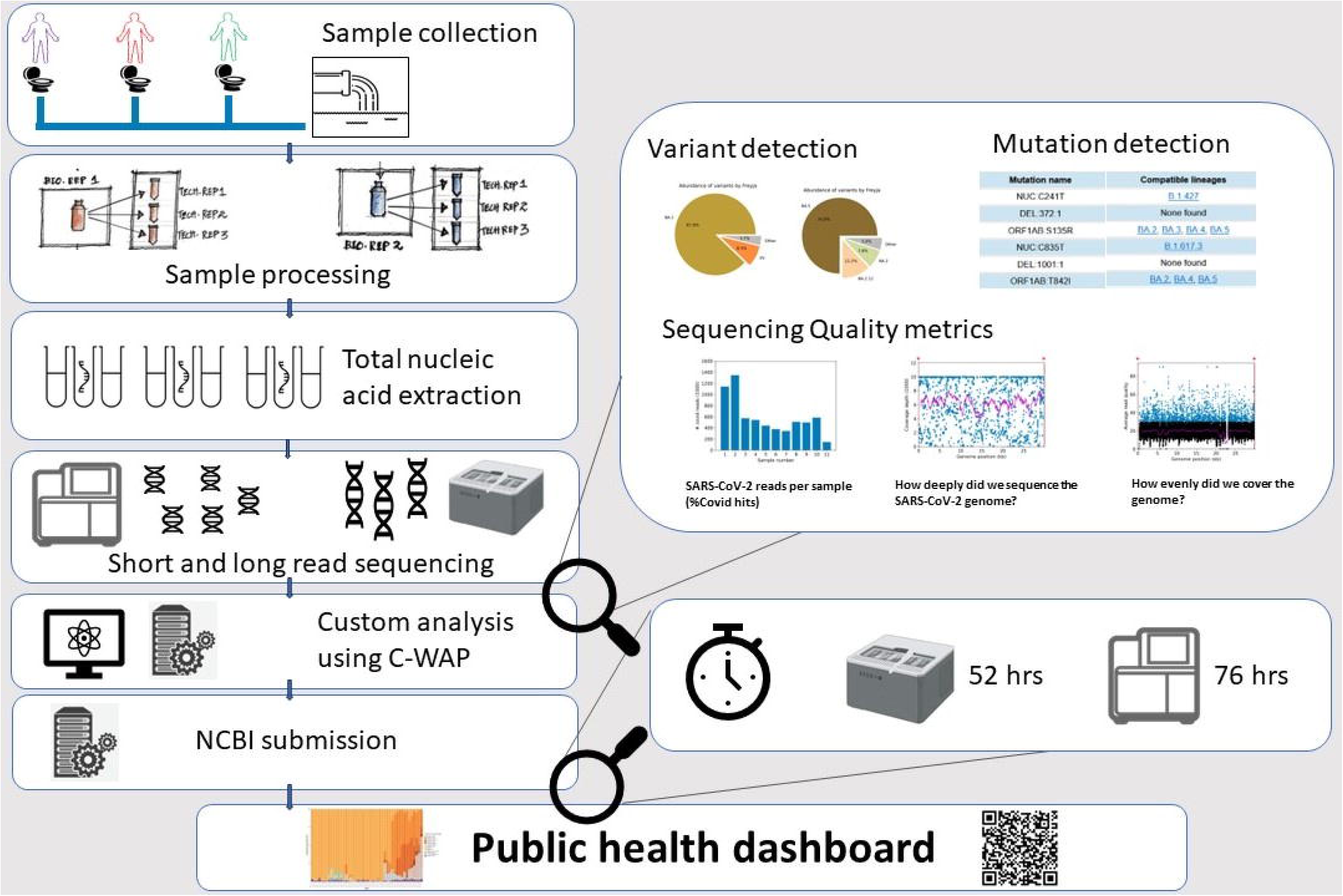
Example reports of the comprehensive analysis produced by C-WAP.

