## Supplementary report 1 for "Harnessing methods, data analysis, and near-real-time wastewater monitoring for enhanced public health response using high throughput sequencing"

U.S. FOOD & DRUG ADMINISTRATION

CFSAN/OAO
BIOSTATISTICS AND BIOINFORMATICS STAFF

### WASTEWATER SARS-COV2 ANALYSIS REPORT

#### Summary

| Sample# | Sample name | Total #reads | Reads aligned PF* | Genomic coordinates 0X | Genomic coordinates <10X |
|---|---|---|---|---|---|
| 1 | CFSANSMP000117854 | 2967188 | 2408377 (81%) | 119nt (0%) | 184nt (0%) |
| 2 | CFSANSMP000117855 | 2834566 | 2515428 (88%) | 94nt (0%) | 180nt (0%) |
| 3 | CFSANSMP000118326 | 4062754 | 3632580 (89%) | 65nt (0%) | 157nt (0%) |
| 4 | CFSANSMP000118328 | 1905298 | 1724073 (90%) | 97nt (0%) | 270nt (0%) |
| 5 | CFSANSMP000118506 | 1300104 | 1097393 (84%) | 119nt (0%) | 159nt (0%) |
| 6 | CFSANSMP000118507 | 2106332 | 1848070 (87%) | 119nt (0%) | 214nt (0%) |
| 7 | CFSANSMP000118583 | 2338632 | 2140098 (91%) | 94nt (0%) | 248nt (0%) |
| 8 | CFSANSMP000118584 | 2526212 | 2345410 (92%) | 119nt (0%) | 410nt (1%) |
| 9 | CFSANSMP000118585 | 2829500 | 2527472 (89%) | 119nt (0%) | 324nt (1%) |
| 10 | CFSANSMP000118586 | 2750930 | 2432859 (88%) | 145nt (0%) | 471nt (1%) |
| 11 | CFSANSMP000118587 | 2531278 | 1323983 (52%) | 121nt (0%) | 523nt (1%) |
| 12 | CFSANSMP000119220 | 2955272 | 1493268 (50%) | 119nt (0%) | 393nt (1%) |
| 13 | CFSANSMP000119222 | 2214544 | 1635964 (73%) | 144nt (0%) | 392nt (1%) |
| 14 | CFSANSMP000119223 | 4089396 | 3182172 (77%) | 92nt (0%) | 504nt (1%) |
| 15 | CFSANSMP000119224 | 6440258 | 2062962 (32%) | 127nt (0%) | 879nt (2%) |
| 16 | CFSANSMP000119624 | 3821038 | 3303434 (86%) | 94nt (0%) | 174nt (0%) |
| 17 | CFSANSMP000119626 | 3761430 | 1843202 (49%) | 122nt (0%) | 277nt (0%) |
| 18 | CFSANSMP000119627 | 3257848 | 1756675 (53%) | 178nt (0%) | 464nt (1%) |
| 19 | CFSANSMP000119629 | 3144978 | 1390136 (44%) | 369nt (1%) | 998nt (3%) |
|  |  |  | 2691770 |  |  |

| | | | | | |
|---|---|---|---|---|---|
| 20 | Undetermined | 4361554 | (61%) | 119nt (0%) | 126nt (0%) |
| 21 | Water | 45196 | 1515 (3%) | 19387nt (64%) | 25466nt (85%) |

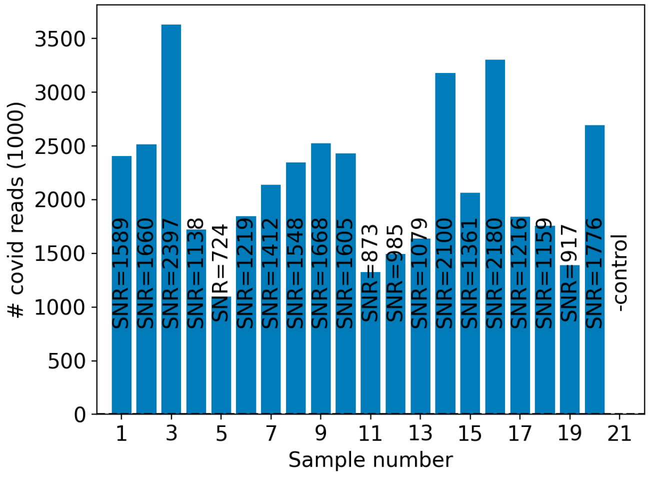


*Quantity of raw reads that align to the reference sequence and pass filter, i.e. the read length after adaptor trimming ≥30 and minimum read quality ≥20 within a sliding window of width 4. SNR refers to the ratio of SC2-mapping reads aligned that pass filter in the sample vs. that in the auto-detected negative control samples (if any). The dashed line represents the baseline level of covid reads detected from the negative control or their average if multiple negative controls we included.

#### QC-bot (Experimental)

| QC category | Subjective definition | Objective metrics |
|---|---|---|
| A | No QC issues evident | 0x coordinates <1%<br>10x coordinates <5%<br>average coverage > 1000X<br>average quality score >35 for Illumina, >15 if ONT, >70 if PacBio HiFi<br>most abundant taxon is coronovirinae |
| B | Some QC issues, but accurate variant calling possible | 0x coordinates <20%<br>10X coordinates < 40%<br>>80% of diverse SNPs covered<br>average coverage > 100X<br>average quality score >35 for Illumina<br>>15 if ONT, >70 if PacBio HiFi |
| C | Some QC issues, and accurate variant calling impossible | 0x coordinates <99%<br>10X coordinates <95% |
| F | Significant QC/study design issues | Contamination (SNR<50)<br>No/negligible coverage (< 1X)<br>Biological/technical replicates' results are irreconcileable. |

| Sample Number | Suggested category | Suggested QC flags |
|---|---|---|
| 1 | A | None |
| 2 | A | None |
| 3 | A | None |
| 4 | A | None |
| 5 | A | None |
| 6 | A | None |

| | | |
|---|---|---|
| 7 | A | None |
| 8 | A | None |
| 9 | A | None |
| 10 | A | None |
| 11 | A | None |
| 12 | A | None |
| 13 | A | None |
| 14 | A | None |
| 15 | A | None |
| 16 | A | None |
| 17 | A | None |
| 18 | A | None |
| 19 | B/C | low_coverage_breadth |
| 20 | A | None |
| 21 | F | sample_contamination |

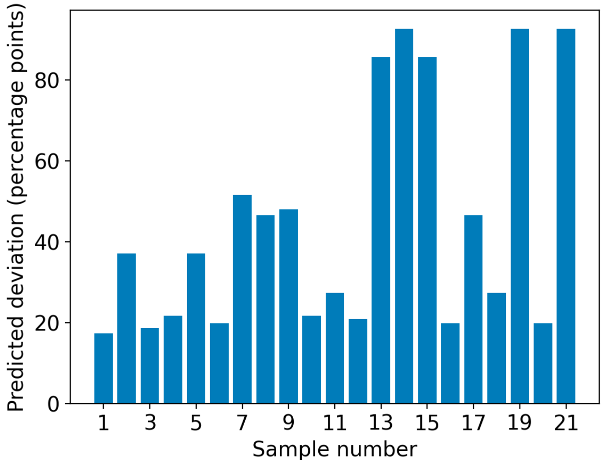


Machine-learning based prediction of the SC2 variant calling accuracy of Freyja of this dataset. The model is a random forest trained on FDA/CFSAN's experimental wastewater WGS data obtained in January 2022 and aims to assess the impact of the potential coverage gaps on the variant abundance estimates. The plotted values represent the predicted deviation of the omicron percentage points from the value that would have been obtained if the coverage was near-complete.

CFSANSMP000117854

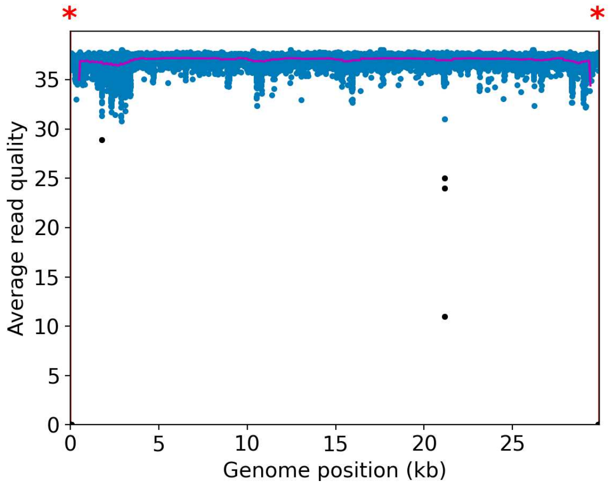


CFSANSMP000117855

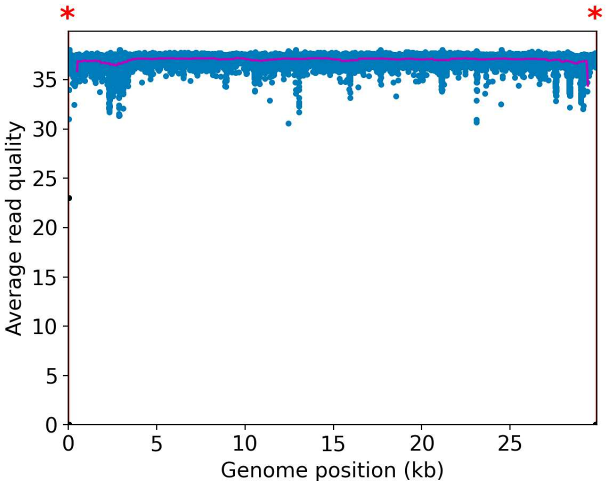

CFSANSMP000118326

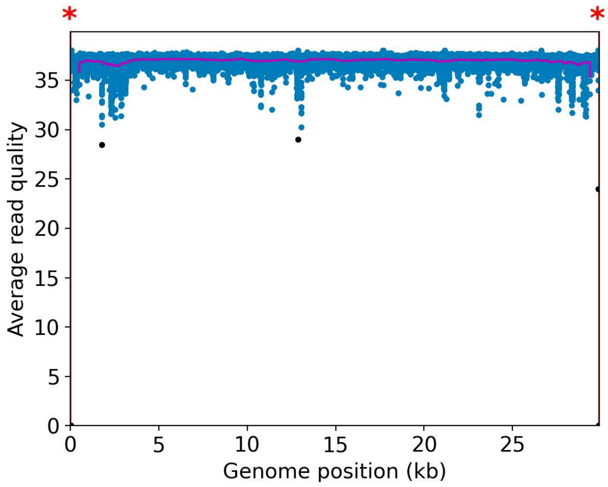


CFSANSMP000118328

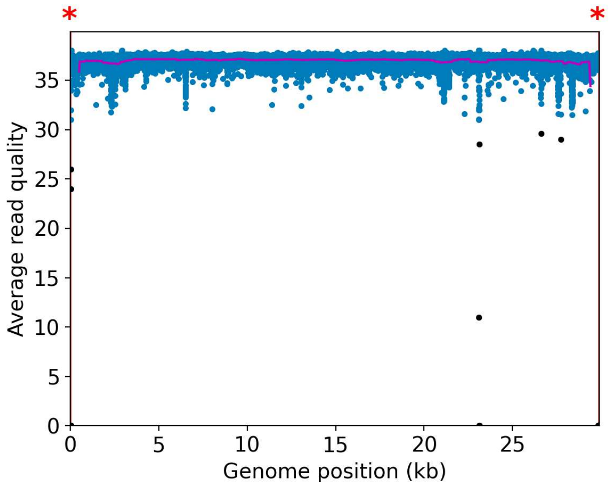


CFSANSMP000118506

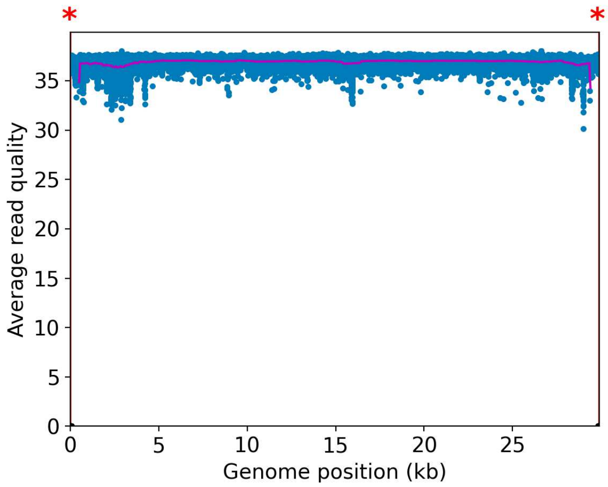


CFSANSMP000118507

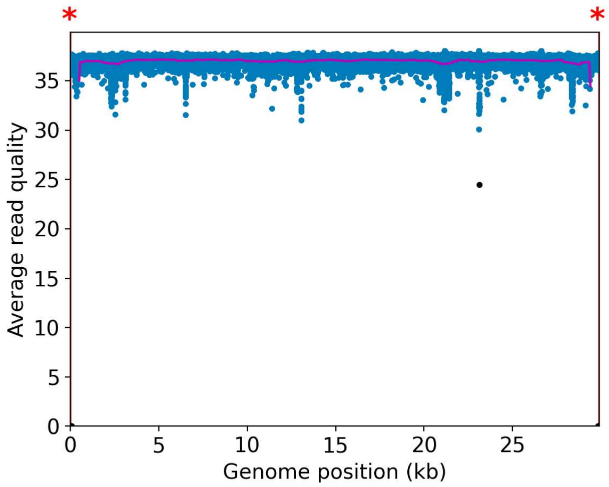


CFSANSMP000118583

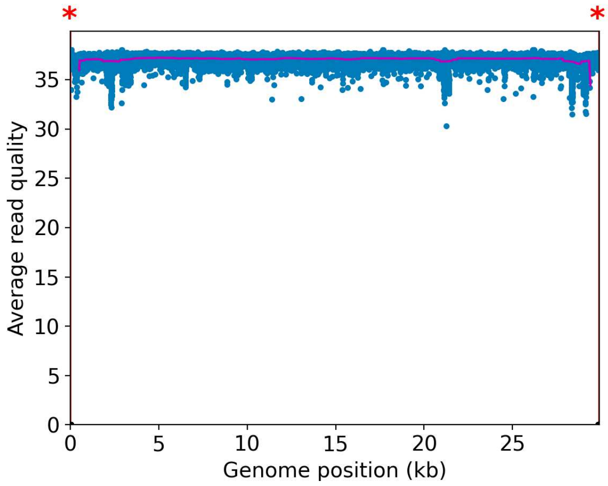


CFSANSMP000118584

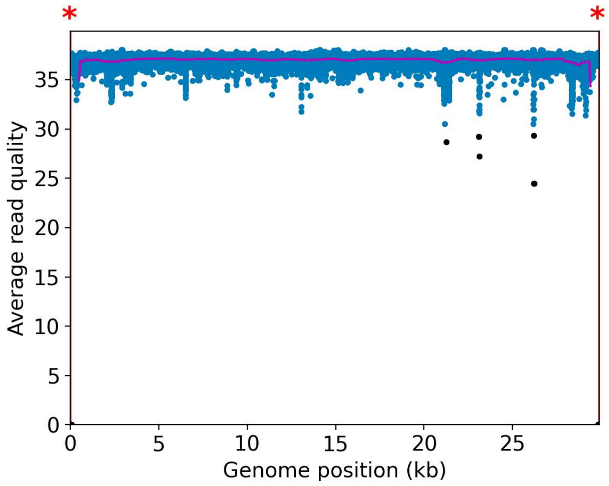


CFSANSMP000118585

CFSANSMP000118586

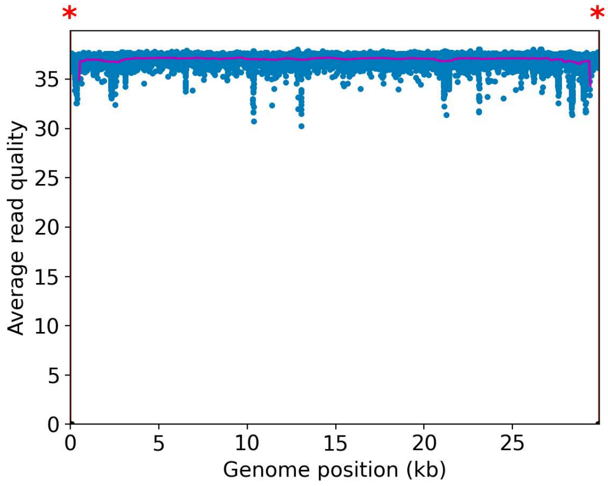


CFSANSMP000118587

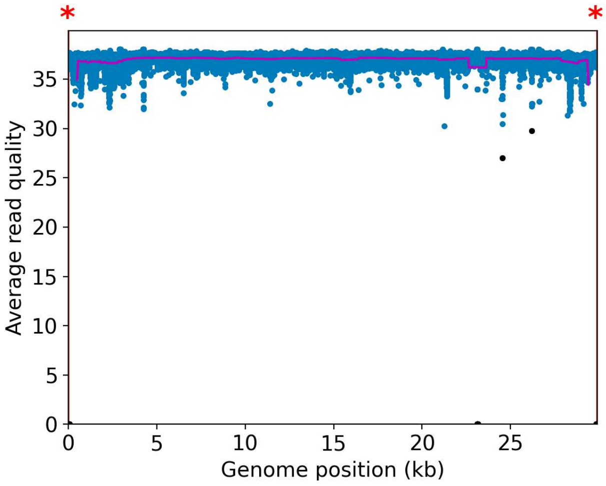


CFSANSMP000119220

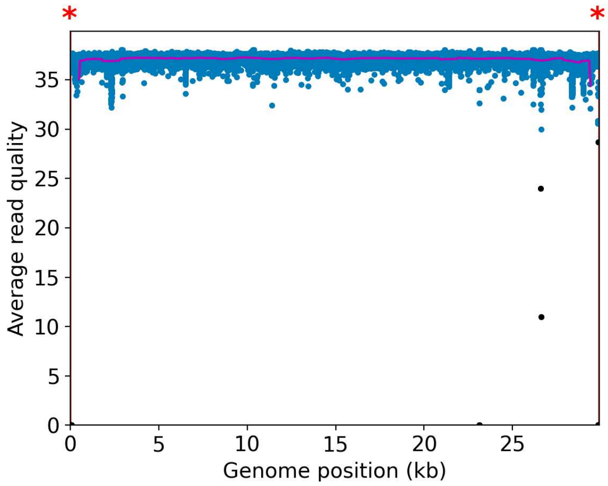


CFSANSMP000119222

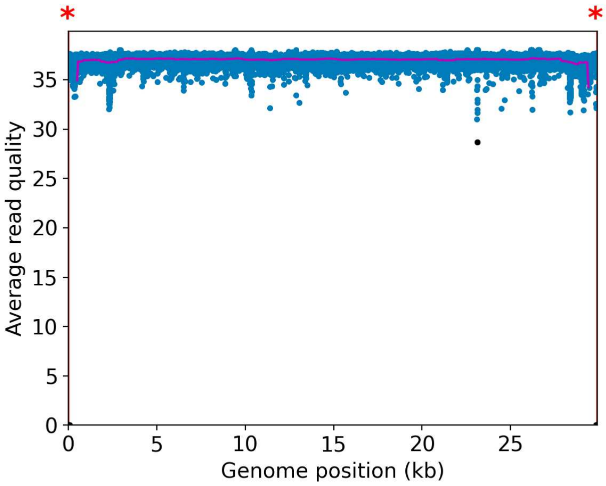


CFSANSMP000119223

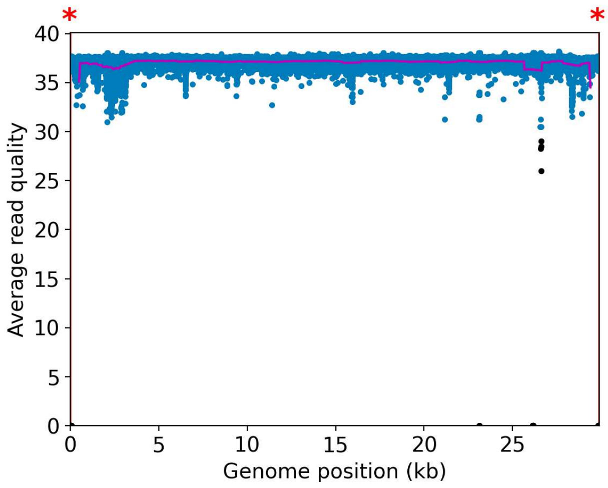


CFSANSMP000119224

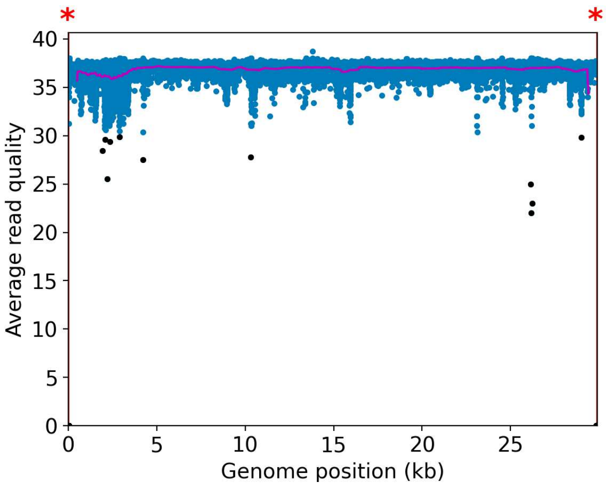


CFSANSMP000119624

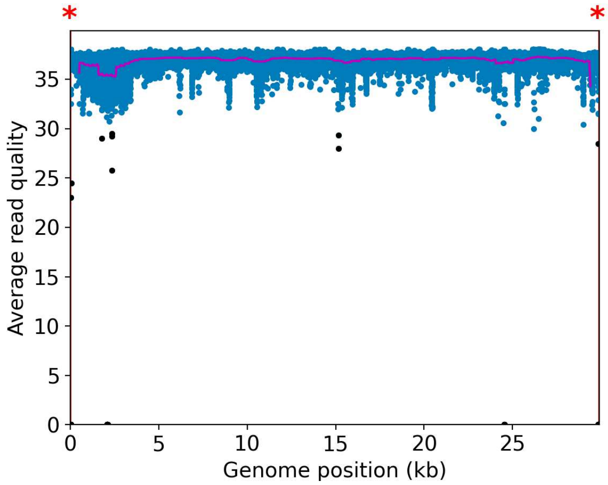


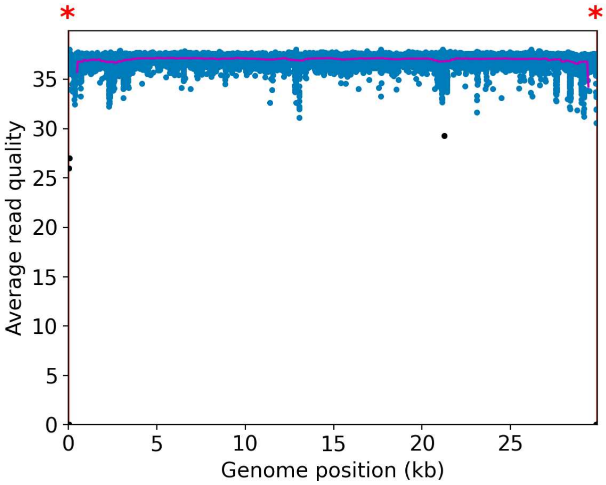


CFSANSMP000119626

CFSANSMP000119627

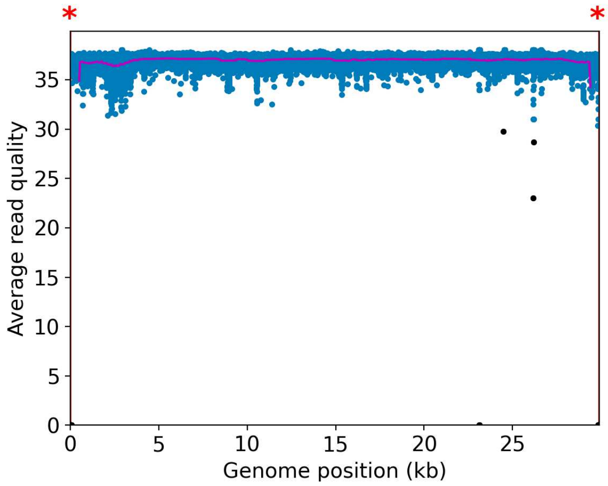


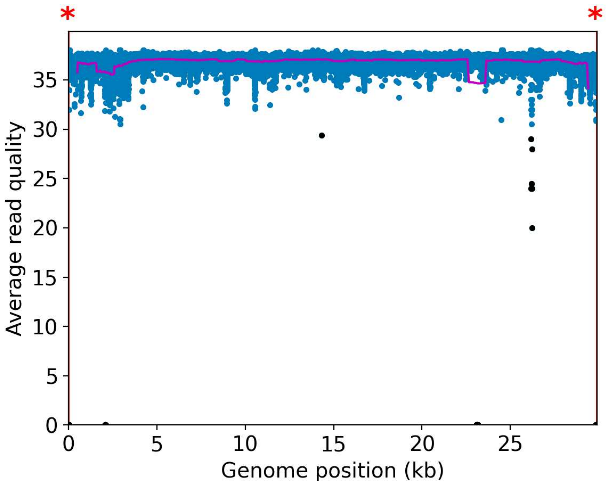


CFSANSMP000119629

Undetermined

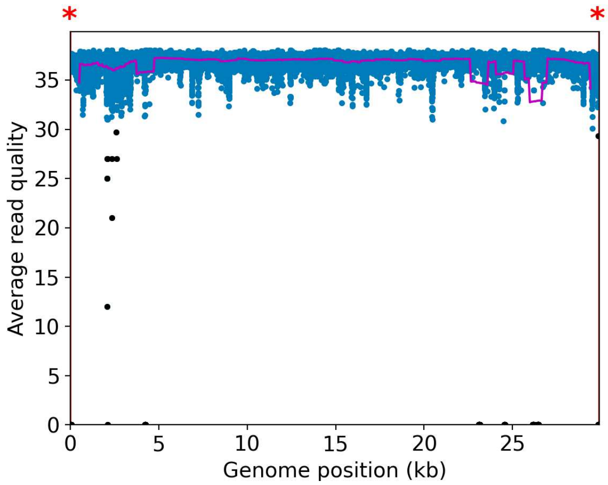


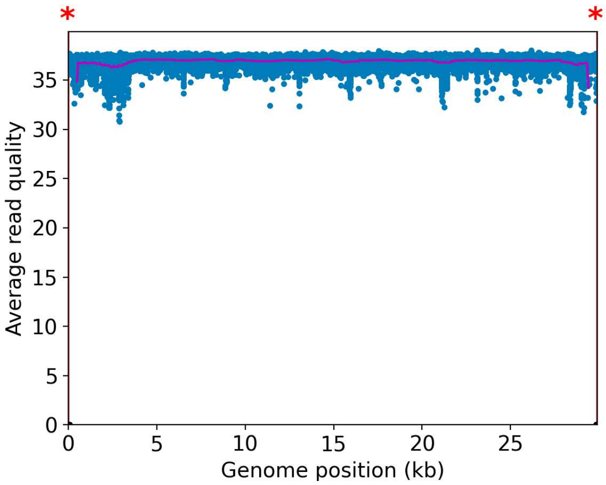


Water

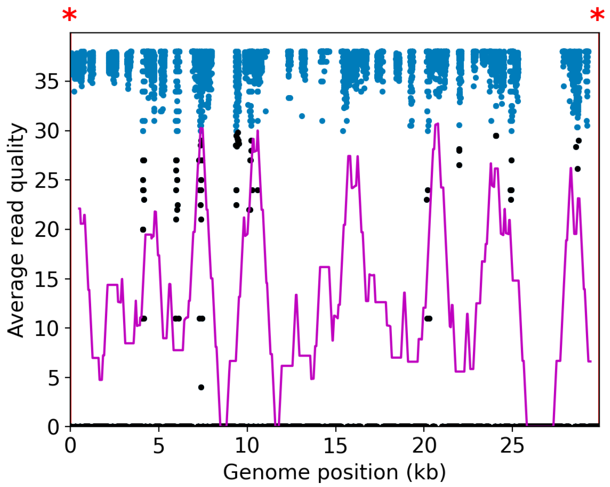

*
*
Average read quality
Genome position (kb)
0
5
10
15
20
25
30
35

U.S. FOOD & DRUG ADMINISTRATION

**CFSAN/OAO**
**BIOSTATISTICS AND BIOINFORMATICS STAFF**

### WASTEWATER SARS-COV2 ANALYSIS REPORT

| | |
|---|---|
| Sample name: | CFSANSMP000117854 |
| Date generated: | 2023-02-01, 20:08:32 EST |
| Timestamp of C-WAP version used: | Tue Jan 31 11:49:22 2023 -0500 |
| Executed by: | Jasmine Amirzadegan |
| Executed on: | 172.20.44.121 (aka n121.raven.cfsan) |

#### Sequencing summary

| | |
|---|---|
| Sequencing chemistry: | Missing with Missing |
| Source site: | Missing (?,?) |
| Sampling date: | Missing |
| Collected by: | Missing |
| Sequenced by: | Missing |
| Total number of reads: | 2967188 |
| Reads aligned: | 2501230 (84%) |
| Average read quality: | 36.7 |
| Average read length: | 149 |
| Reads passing filter: | 2408377 (81%) |
| Average read quality passing filter: | 37.0 |
| Average read length passing filter: | 149 |
| Average coverage passing filter: | 12000X |

A read passes filter if the read length after adaptor trimming ≥ 30 and minimum read quality ≥ 20 within a sliding window of width 4.

#### Overall sequence characteristics

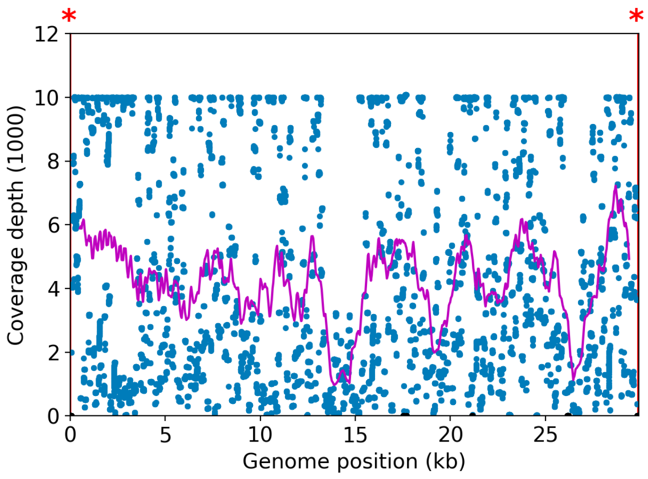


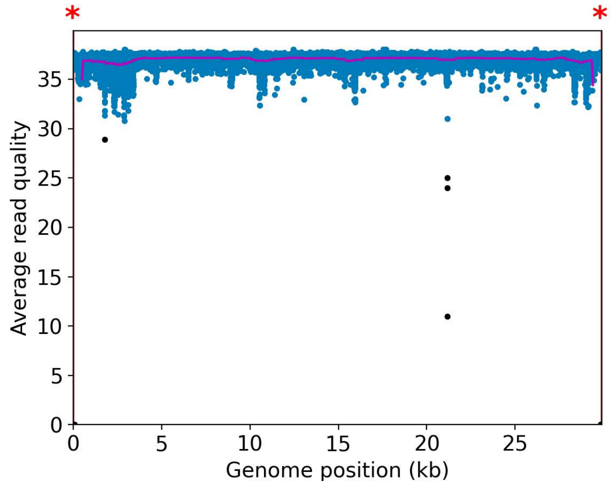


NOTE: The red shaded areas marked with a (*) are not covered by the design of the library preparation kit and hence excluded from analyses. Magenta curves represent moving average with a window width of 1kb.

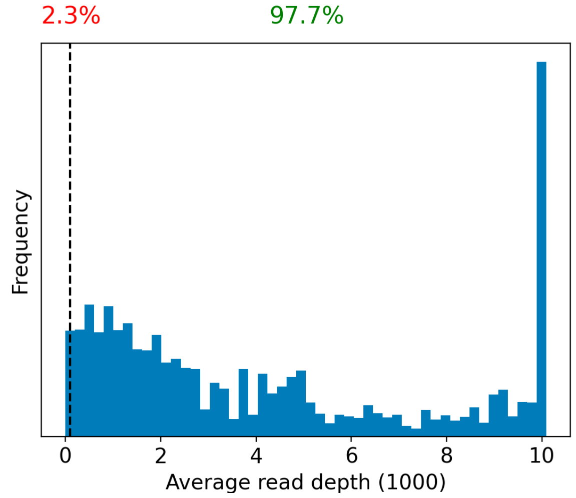


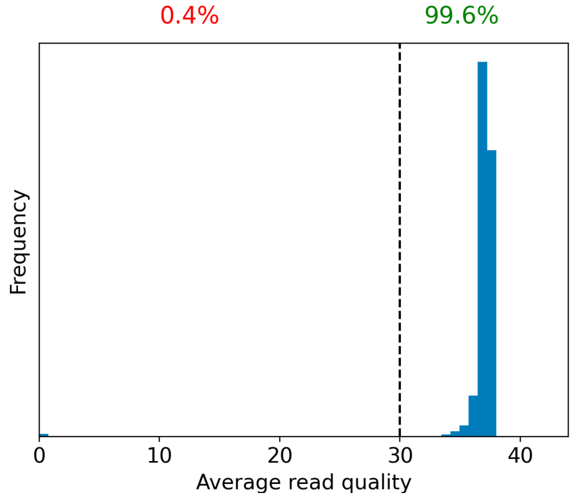


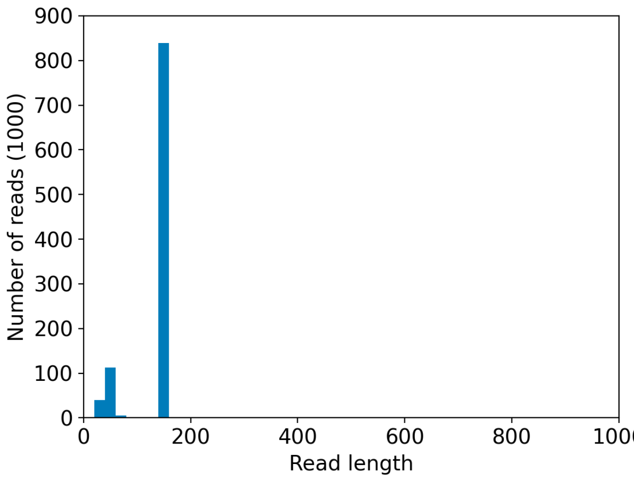


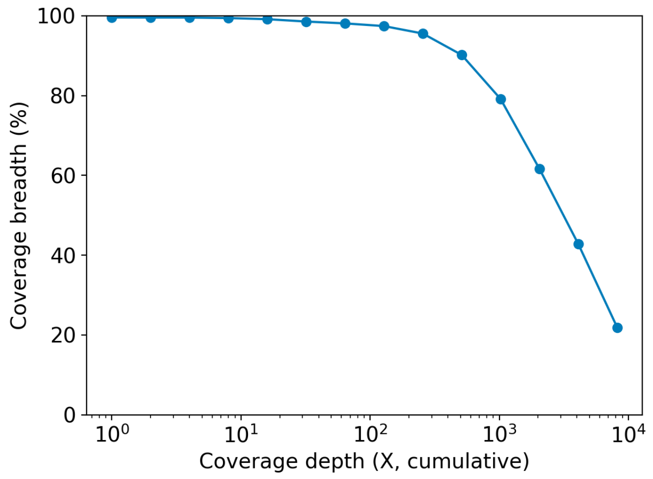

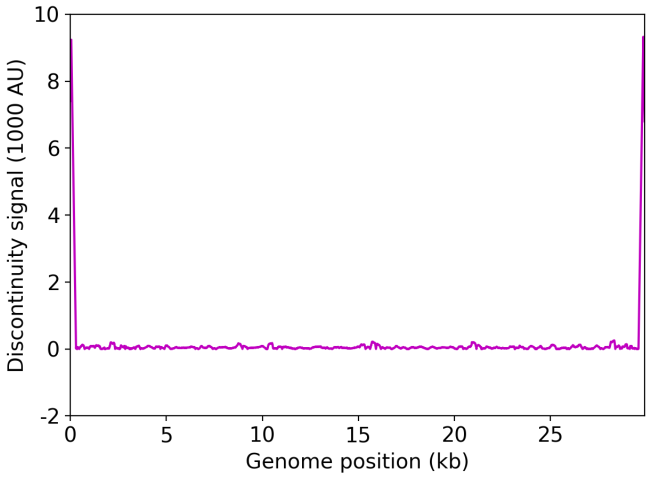


| | Uncovered coordinates (0X) | Poorly covered coordinates (<10X) |
|---|---|---|
| # Inaccessible genomic coordinates by kit design: | 121nt (0%) | 121nt (0%) |
| All genomic coordinates: | 119nt (0%) | 184nt (0%) |
| Common SNPs: | 0nt (0%) | 0nt (0%) |
| Diverse SNPs: | 29nt (12%) | 29nt (12%) |
| Rare SNPs: | 10nt (1%) | 10nt (1%) |

SNPs refer to the polymorphic sites currently in circulation that were detected out of recent GISAID entries. The sites that differ from the SC2 reference sequence are denoted as "common" if [90%, 100%] of the submissions carry this mutation, whereas those that are prevalent in [0%,10%] of the submissions are grouped under the "rare" category. The population is still diverse at the mutation sites that are observed in (10%,90%) of the entries and these coordinates are grouped under the "diverse" category.

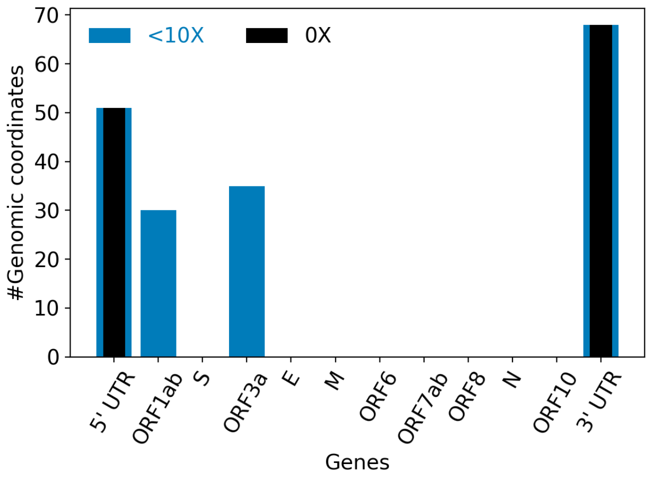


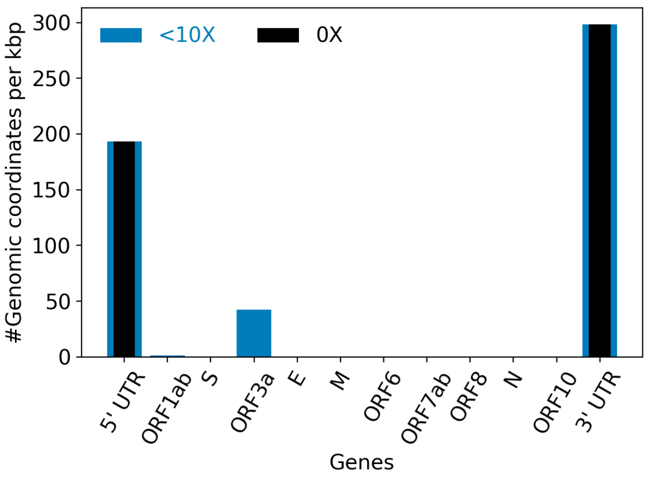


| | |
|---|---|
| Hits to SARS-Cov2 genome (kraken2): | 1282992 reads (86.48%) |
| Hits to human genome (kraken2): | 733 reads (0.05%) |
| Hits to synthetic sequences (kraken2, taxid 28384): | 238 reads (0.02%) |
| Most abundant organisms (kraken2, family level): | Coronaviridae (86.48%)<br>Hominidae (0.05%)<br>Staphylococcaceae (0.02%) |

| | |
|---|---|
| | Arcobacteraceae (0.02%)<br>Enterobacteriaceae (0.01%) |

#### Detected variants (Experimental)

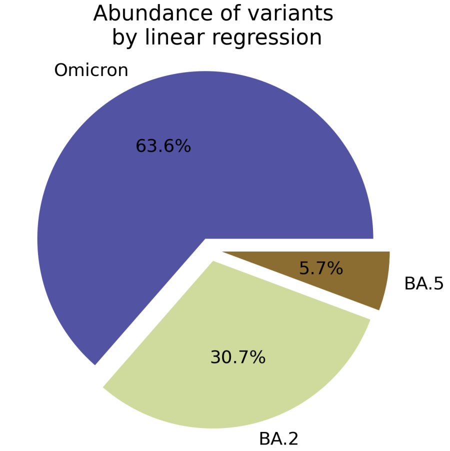


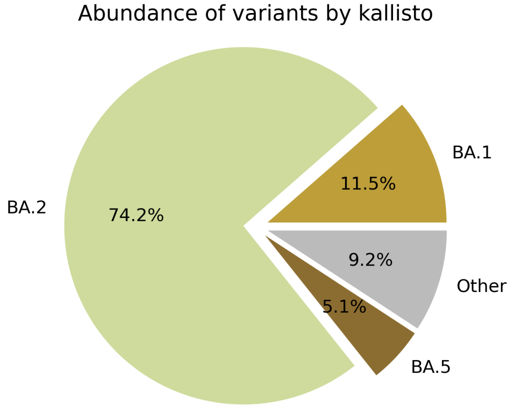


Based on deconvolution, B.1.1.529 is estimated to constitute 63.58% of the viral particles and hence is the most abundant variant in the sample. The $R^2$ for the linear regression was 0.62. Variants that were detected less than 5% were grouped under "Other"

Based on the consensus sequence of the observed reads, the "ensemble-averaged sequence" most closely resembles the BA.2 lineage. If this is a sample consisting of a single source of pathogens or an overwhelming majority of the different sources are infected with the same variant, the sample is dominated by this variant.

Based on mapping individual reads to the variant consensus sequences in the reference database, kallisto predicts that the sample is dominated by BA.2 lineage. Accuracy of this measure is expected to improve if the input data consists of long reads as opposed to convolution.

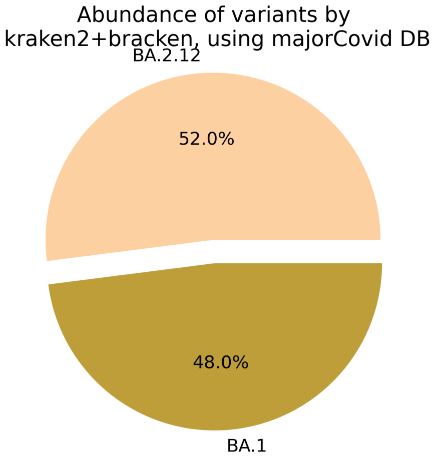


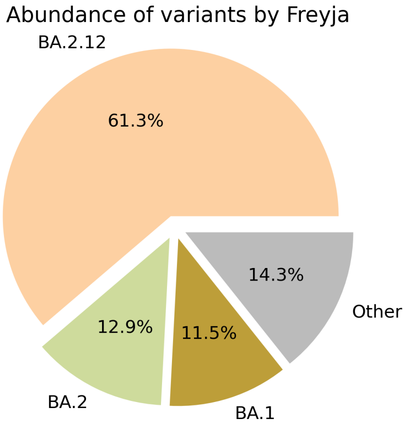

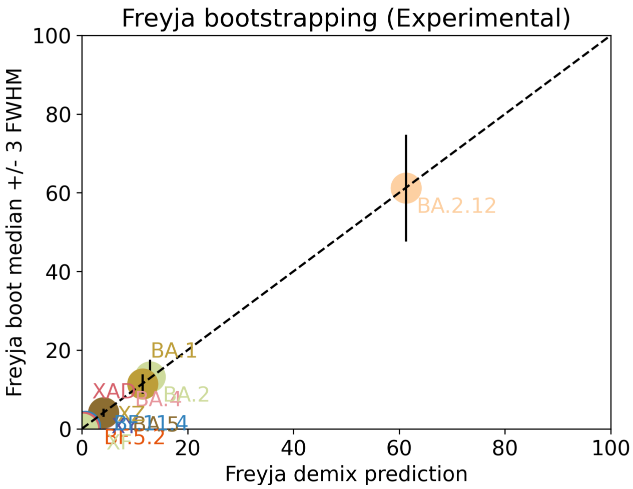


Under the assumption that the presence of a variant requires the detection of all respective mutations of the variant, the characteric mutations which support the presence of the respective variant are indicated in the respective column of the table. Numbers show the number of mutations detected, if any, and the number of mutations expected to be present based on the variant definitions.

| VOC | B.1.617.2 | BA.1 | BA.2 | BA.3 | BA.4 | BA.5 |
|---|---|---|---|---|---|---|
| Characteristic mutations detected | (3 of 13)<br>S:G142D<br>S:L452R<br>S:T478K | (17 of 26)<br>M:D3G<br>NUC:C15240T<br>NUC:C25000T<br>NUC:C25584T<br>NUC:T13195C<br>NUC:T5386G<br>ORF1AB:A2710T<br>ORF1AB:I3758V<br>ORF1AB:K856R<br>S:A67V<br>S:G446S<br>S:G496S<br>S:L981F<br>S:N856K<br>S:Q493R<br>S:T547K<br>S:T95I | (23 of 31)<br>N:S413R<br>NUC:A20055G<br>NUC:A9424G<br>NUC:C10198T<br>NUC:C12880T<br>NUC:C15714T<br>NUC:C25000T<br>NUC:C25584T<br>NUC:C26858T<br>NUC:C4321T<br>NUC:G10447A<br>ORF1AB:G1307S<br>ORF1AB:L3027F<br>ORF1AB:L3201F<br>ORF1AB:S135R<br>ORF1AB:T3090I<br>ORF1AB:T842I<br>S:D405N<br>S:Q493R<br>S:R408S<br>S:S371F<br>S:T19I<br>S:T376A | (13 of 21)<br>N:S413R<br>NUC:C12880T<br>NUC:C15714T<br>NUC:C26858T<br>NUC:G10447A<br>ORF1AB:G1307S<br>ORF1AB:S135R<br>ORF1AB:T3090I<br>S:A67V<br>S:D405N<br>S:G446S<br>S:Q493R<br>S:S371F | (24 of 31)<br>N:P151S<br>N:S413R<br>NUC:A20055G<br>NUC:C10198T<br>NUC:C12880T<br>NUC:C15714T<br>NUC:C25000T<br>NUC:C25584T<br>NUC:C26858T<br>NUC:C4321T<br>NUC:G10447A<br>NUC:G12160A<br>NUC:G27788T<br>ORF1AB:G1307S<br>ORF1AB:S135R<br>ORF1AB:T3090I<br>ORF1AB:T842I<br>S:D405N<br>S:F486V<br>S:L452R<br>S:S371F<br>S:T19I<br>S:T376A<br>S:V213G | (22 of 28)<br>M:D3N<br>N:S413R<br>NUC:A20055G<br>NUC:C10198T<br>NUC:C12880T<br>NUC:C15714T<br>NUC:C25000T<br>NUC:C25584T<br>NUC:C4321T<br>NUC:G10447A<br>NUC:G12160A<br>ORF1AB:G1307S<br>ORF1AB:S135R<br>ORF1AB:T3090I<br>ORF1AB:T842I<br>S:D405N<br>S:F486V<br>S:L452R<br>S:S371F<br>S:T19I<br>S:T376A<br>S:V213G |

Jaccard Index is a measure of similarity between two sets A and B, reaching the maximum value of 1 if A=B and minimum value of 0 if A ∩ B = {}. In the c(d) representation below, c represents the Jaccard index of the set of mutations that were experimentally detected for this sample as listed above, whereas d refers to the ideal value of the Jaccard index expected from complete genome coverage without any sequencing errors.

| | B.1.617.2 | BA.1 | BA.2 | BA.3 | BA.4 | BA.5 |
|---|---|---|---|---|---|---|
| B.1.617.2 | 1.00 (1.00) | 0.00 (0.00) | 0.00 (0.00) | 0.00 (0.00) | 0.04 (0.02) | 0.04 (0.03) |
| BA.1 | 0.00 (0.00) | 1.00 (1.00) | 0.08 (0.10) | 0.11 (0.21) | 0.05 (0.08) | 0.05 (0.08) |
| BA.2 | 0.00 (0.00) | 0.08 (0.10) | 1.00 (1.00) | 0.44 (0.33) | 0.62 (0.63) | 0.61 (0.59) |
| BA.3 | 0.00 (0.00) | 0.11 (0.21) | 0.44 (0.33) | 1.00 (1.00) | 0.37 (0.30) | 0.35 (0.29) |

| | | | | | | |
|---|---|---|---|---|---|---|
| BA.4 | 0.04 (0.02) | 0.05 (0.08) | 0.62 (0.63) | 0.37 (0.30) | 1.00 (1.00) | 0.84 (0.84) |
| BA.5 | 0.04 (0.03) | 0.05 (0.08) | 0.61 (0.59) | 0.35 (0.29) | 0.84 (0.84) | 1.00 (1.00) |

#### Detected mutations

Excluded from this pdf version due to file size limitations.

CFSAN/OAO
BIOSTATISTICS AND BIOINFORMATICS STAFF

### WASTEWATER SARS-COV2 ANALYSIS REPORT

| | |
|---|---|
| Sample name: | CFSANSMP000117855 |
| Date generated: | 2023-02-01, 20:10:30 EST |
| Timestamp of C-WAP version used: | Tue Jan 31 11:49:22 2023 -0500 |
| Executed by: | Jasmine Amirzadegan |
| Executed on: | 172.20.44.108 (aka n108.raven.cfsan) |

#### Sequencing summary

| | |
|---|---|
| Sequencing chemistry: | Missing with Missing |
| Source site: | Missing (?,?) |
| Sampling date: | Missing |
| Collected by: | Missing |
| Sequenced by: | Missing |
| Total number of reads: | 2834566 |
| Reads aligned: | 2620595 (92%) |
| Average read quality: | 36.7 |
| Average read length: | 149 |
| Reads passing filter: | 2515428 (88%) |
| Average read quality passing filter: | 36.9 |
| Average read length passing filter: | 149 |
| Average coverage passing filter: | 12533X |

A read passes filter if the read length after adaptor trimming ≥ 30 and minimum read quality ≥ 20 within a sliding window of width 4.

#### Overall sequence characteristics

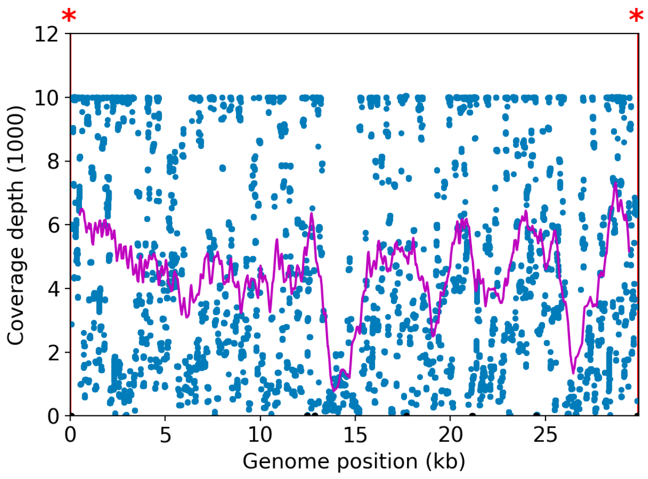

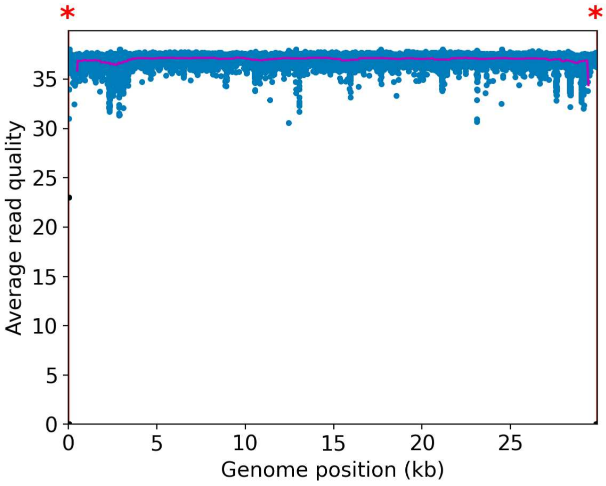


NOTE: The red shaded areas marked with a (*) are not covered by the design of the library preparation kit and hence excluded from analyses. Magenta curves represent moving average with a window width of 1kb.

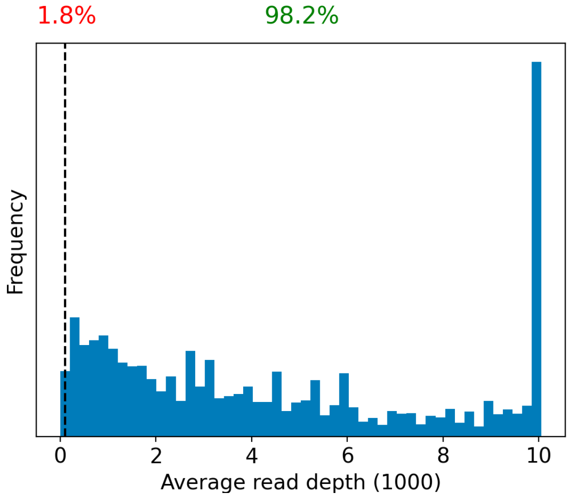

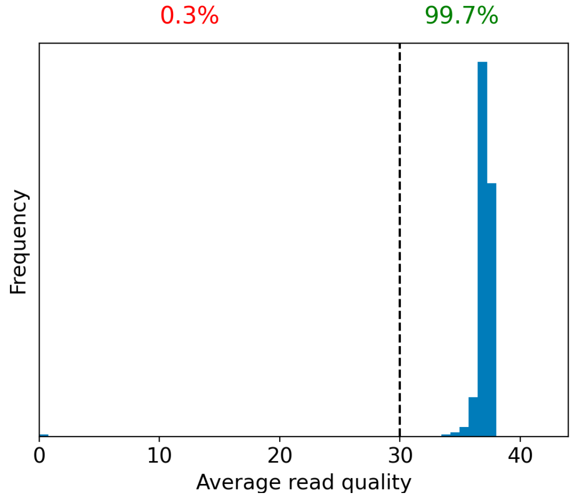

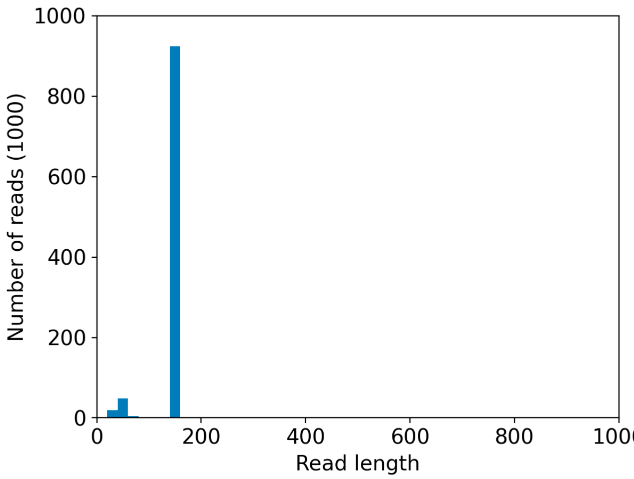

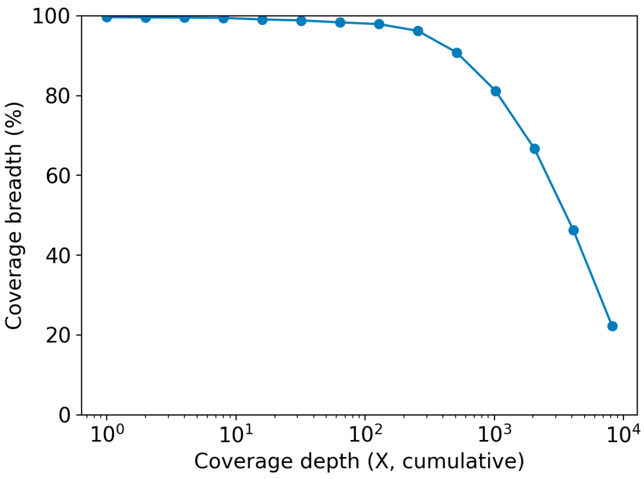

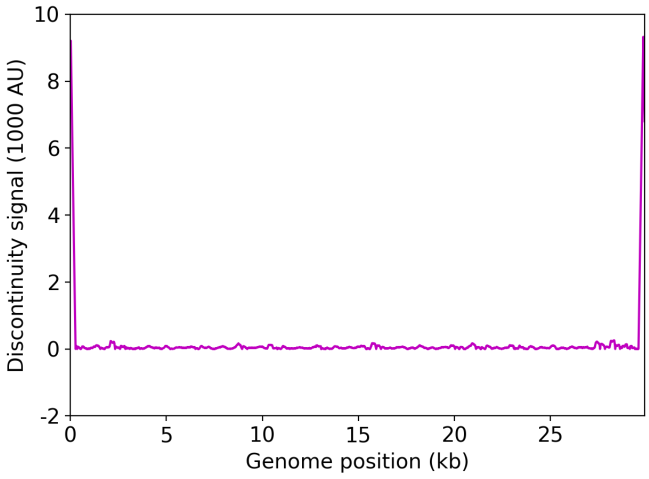


| | Uncovered coordinates (0X) | Poorly covered coordinates (<10X) |
|---|---|---|
| # Inaccessible genomic coordinates by kit design: | 121nt (0%) | 121nt (0%) |
| All genomic coordinates: | 94nt (0%) | 180nt (0%) |
| Common SNPs: | 0nt (0%) | 0nt (0%) |
| Diverse SNPs: | 28nt (12%) | 29nt (12%) |
| Rare SNPs: | 9nt (0%) | 10nt (1%) |

SNPs refer to the polymorphic sites currently in circulation that were detected out of recent GISAID entries. The sites that differ from the SC2 reference sequence are denoted as "common" if [90%, 100%] of the submissions carry this mutation, whereas those that are prevalent in [0%,10%] of the submissions are grouped under the "rare" category. The population is still diverse at the mutation sites that are observed in (10%,90%) of the entries and these coordinates are grouped under the "diverse" category.

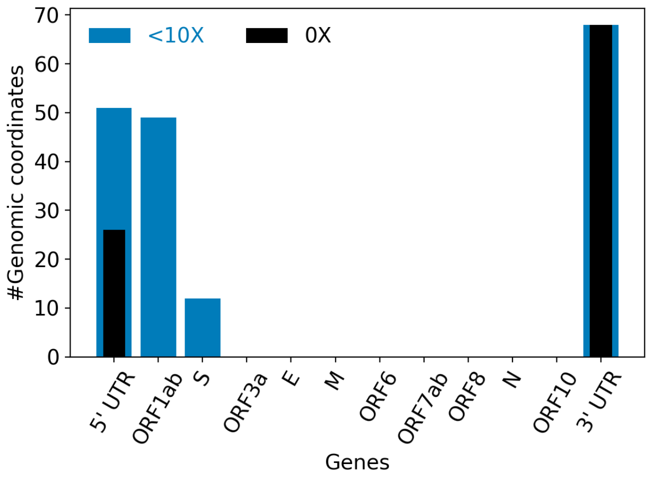


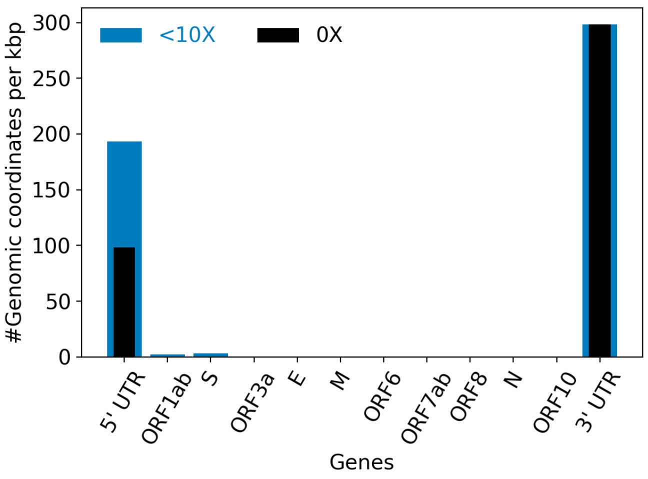


| | |
|---|---|
| Hits to SARS-Cov2 genome (kraken2): | 1332746 reads (94.04%) |
| Hits to human genome (kraken2): | 151 reads (0.01%) |
| Hits to synthetic sequences (kraken2, taxid 28384): | 65 reads (0.00%) |
| Most abundant organisms (kraken2, family level): | Coronaviridae (94.04%)<br>Bacteroidaceae (0.07%)<br>Hominidae (0.01%) |

| | |
|---|---|
| | Staphylococcaceae (0.01%)<br>Arcobacteraceae (0.00%) |

### Detected variants (Experimental)

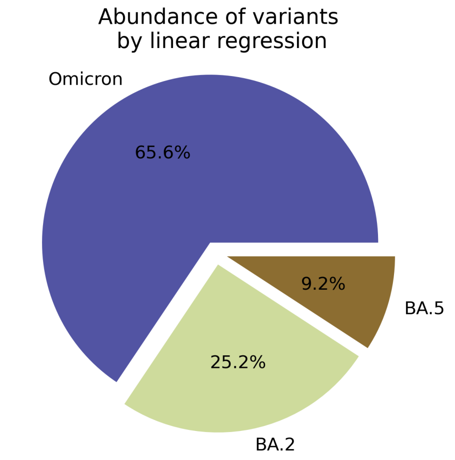


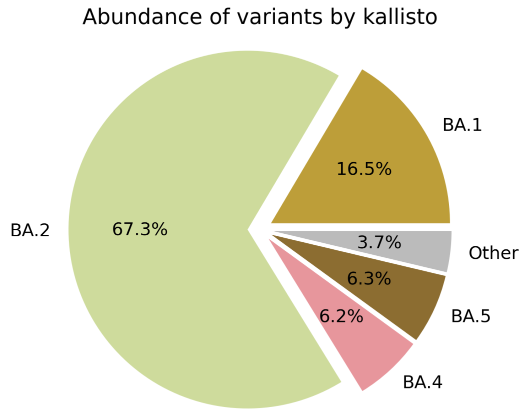


Based on deconvolution, B.1.1.529 is estimated to constitute 65.56% of the viral particles and hence is the most abundant variant in the sample. The $R^2$ for the linear regression was 0.63. Variants that were detected less than 5% were grouped under "Other"

Based on the consensus sequence of the observed reads, the "ensemble-averaged sequence" most closely resembles the BA.2 lineage. If this is a sample consisting of a single source of pathogens or an overwhelming majority of the different sources are infected with the same variant, the sample is dominated by this variant.

Based on mapping individual reads to the variant consensus sequences in the reference database, kallisto predicts that the sample is dominated by BA.2 lineage. Accuracy of this measure is expected to improve if the input data consists of long reads as opposed to convolution.

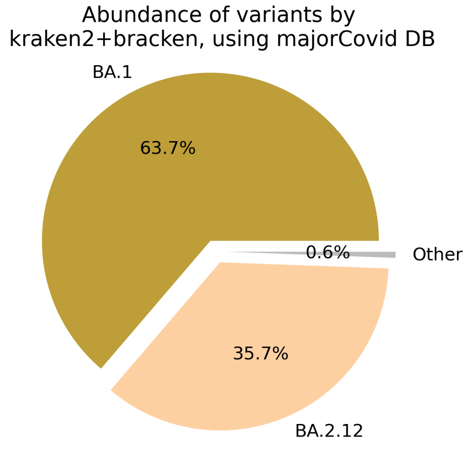


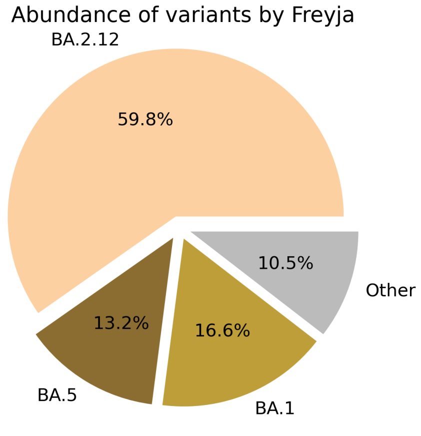

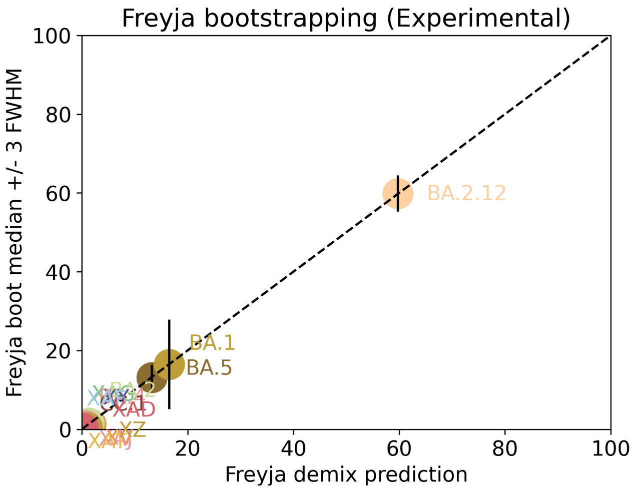


Under the assumption that the presence of a variant requires the detection of all respective mutations of the variant, the characteric mutations which support the presence of the respective variant are indicated in the respective column of the table. Numbers show the number of mutations detected, if any, and the number of mutations expected to be present based on the variant definitions.

| VOC | [B.1.617.2]() | [BA.1]() | [BA.2]() | [BA.3]() | [BA.4]() | [BA.5]() |
|---|---|---|---|---|---|---|
| Characteristic mutations detected | (3 of 13)<br>S:G142D<br>S:L452R<br>S:T478K | (16 of 26)<br>M:D3G<br>NUC:C15240T<br>NUC:C25000T<br>NUC:C25584T<br>NUC:T13195C<br>NUC:T5386G<br>ORF1AB:A2710T<br>ORF1AB:I3758V<br>ORF1AB:K856R<br>S:G446S<br>S:G496S<br>S:L981F<br>S:N856K<br>S:Q493R<br>S:T547K<br>S:T95I | (23 of 31)<br>N:S413R<br>NUC:A20055G<br>NUC:A9424G<br>NUC:C10198T<br>NUC:C12880T<br>NUC:C15714T<br>NUC:C25000T<br>NUC:C25584T<br>NUC:C26858T<br>NUC:C4321T<br>NUC:G10447A<br>ORF1AB:G1307S<br>ORF1AB:L3027F<br>ORF1AB:L3201F<br>ORF1AB:S135R<br>ORF1AB:T3090I<br>ORF1AB:T842I<br>S:D405N<br>S:Q493R<br>S:R408S<br>S:S371F<br>S:T19I<br>S:T376A | (12 of 21)<br>N:S413R<br>NUC:C12880T<br>NUC:C15714T<br>NUC:C26858T<br>NUC:G10447A<br>ORF1AB:G1307S<br>ORF1AB:S135R<br>ORF1AB:T3090I<br>S:D405N<br>S:G446S<br>S:Q493R<br>S:S371F | (24 of 31)<br>N:P151S<br>N:S413R<br>NUC:A20055G<br>NUC:C10198T<br>NUC:C12880T<br>NUC:C15714T<br>NUC:C25000T<br>NUC:C25584T<br>NUC:C26858T<br>NUC:C4321T<br>NUC:G10447A<br>NUC:G12160A<br>NUC:G27788T<br>ORF1AB:G1307S<br>ORF1AB:S135R<br>ORF1AB:T3090I<br>ORF1AB:T842I<br>S:D405N<br>S:F486V<br>S:L452R<br>S:S371F<br>S:T19I<br>S:T376A<br>S:V213G | (22 of 28)<br>M:D3N<br>N:S413R<br>NUC:A20055G<br>NUC:C10198T<br>NUC:C12880T<br>NUC:C15714T<br>NUC:C25000T<br>NUC:C25584T<br>NUC:C4321T<br>NUC:G10447A<br>NUC:G12160A<br>ORF1AB:G1307S<br>ORF1AB:S135R<br>ORF1AB:T3090I<br>ORF1AB:T842I<br>S:D405N<br>S:F486V<br>S:L452R<br>S:S371F<br>S:T19I<br>S:T376A<br>S:V213G |

[Jaccard Index]() is a measure of similarity between two sets A and B, reaching the maximum value of 1 if A=B and minimum value of 0 if A ∩ B = {}. In the c(d) representation below, c represents the Jaccard index of the set of mutations that were experimentally detected for this sample as listed above, whereas d refers to the ideal value of the Jaccard index expected from complete genome coverage without any sequencing errors.

| | B.1.617.2 | BA.1 | BA.2 | BA.3 | BA.4 | BA.5 |
|---|---|---|---|---|---|---|
| B.1.617.2 | 1.00 ([1.00]()) | 0.00 ([0.00]()) | 0.00 ([0.00]()) | 0.00 ([0.00]()) | 0.04 ([0.02]()) | 0.04 ([0.03]()) |
| BA.1 | 0.00 ([0.00]()) | 1.00 ([1.00]()) | 0.08 ([0.10]()) | 0.08 ([0.21]()) | 0.05 ([0.08]()) | 0.06 ([0.08]()) |
| BA.2 | 0.00 ([0.00]()) | 0.08 ([0.10]()) | 1.00 ([1.00]()) | 0.46 ([0.33]()) | 0.62 ([0.63]()) | 0.61 ([0.59]()) |
| BA.3 | 0.00 ([0.00]()) | 0.08 ([0.21]()) | 0.46 ([0.33]()) | 1.00 ([1.00]()) | 0.38 ([0.30]()) | 0.36 ([0.29]()) |

| | | | | | | |
|---|---|---|---|---|---|---|
| BA.4 | 0.04 (0.02) | 0.05 (0.08) | 0.62 (0.63) | 0.38 (0.30) | 1.00 (1.00) | 0.84 (0.84) |
| BA.5 | 0.04 (0.03) | 0.06 (0.08) | 0.61 (0.59) | 0.36 (0.29) | 0.84 (0.84) | 1.00 (1.00) |

#### Detected mutations

Excluded from this pdf version due to file size limitations.

CFSAN/OAO
BIOSTATISTICS AND BIOINFORMATICS STAFF

### WASTEWATER SARS-COV2 ANALYSIS REPORT

| | |
|---|---|
| Sample name: | CFSANSMP000118326 |
| Date generated: | 2023-02-01, 20:15:34 EST |
| Timestamp of C-WAP version used: | Tue Jan 31 11:49:22 2023 -0500 |
| Executed by: | Jasmine Amirzadegan |
| Executed on: | 172.20.44.108 (aka n108.raven.cfsan) |

#### Sequencing summary

| | |
|---|---|
| Sequencing chemistry: | Missing with Missing |
| Source site: | Missing (?,?) |
| Sampling date: | Missing |
| Collected by: | Missing |
| Sequenced by: | Missing |
| Total number of reads: | 4062754 |
| Reads aligned: | 3798618 (93%) |
| Average read quality: | 36.6 |
| Average read length: | 149 |
| Reads passing filter: | 3632580 (89%) |
| Average read quality passing filter: | 36.9 |
| Average read length passing filter: | 149 |
| Average coverage passing filter: | 18100X |

A read passes filter if the read length after adaptor trimming ≥ 30 and minimum read quality ≥ 20 within a sliding window of width 4.

#### Overall sequence characteristics

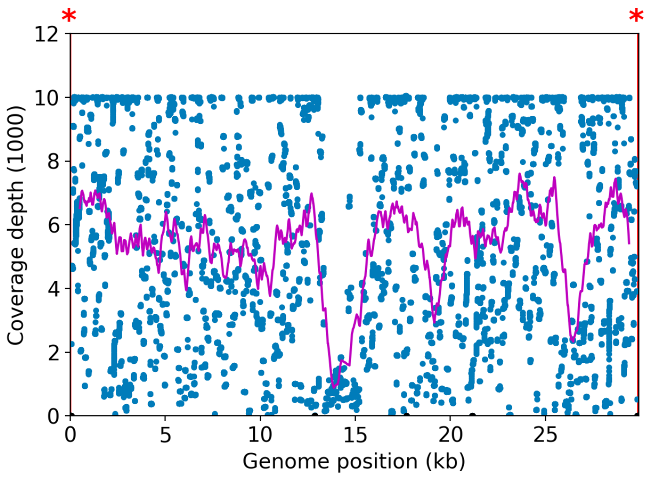


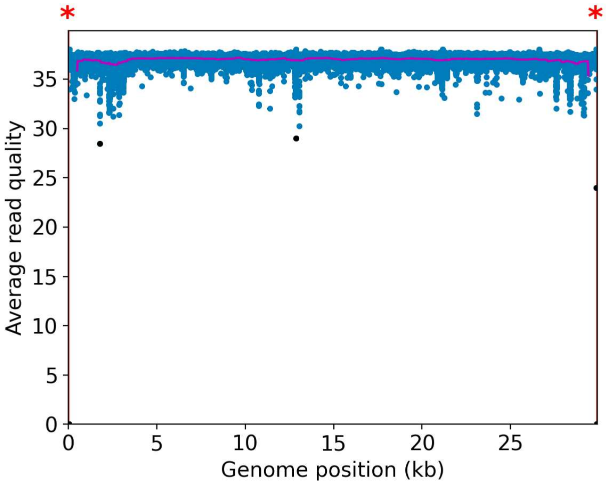


NOTE: The red shaded areas marked with a (*) are not covered by the design of the library preparation kit and hence excluded from analyses. Magenta curves represent moving average with a window width of 1kb.

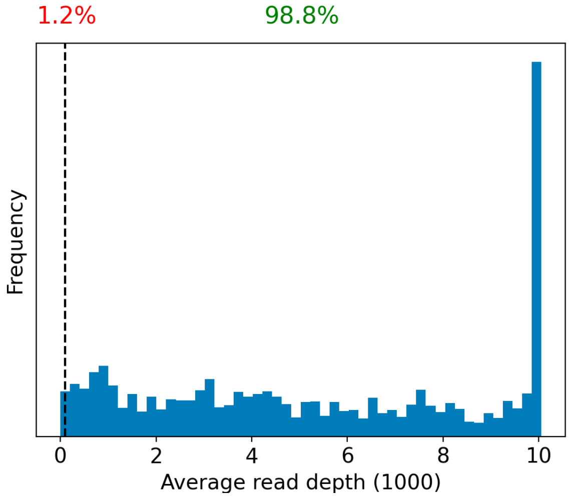


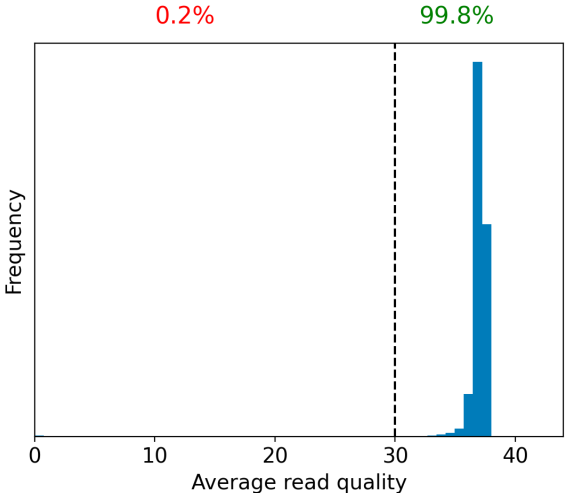


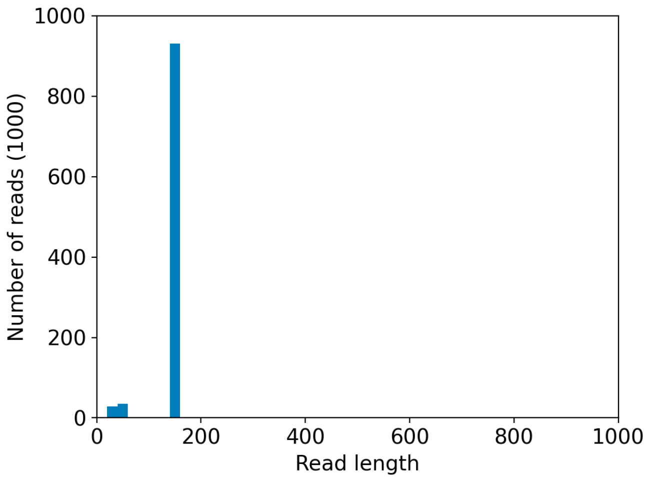


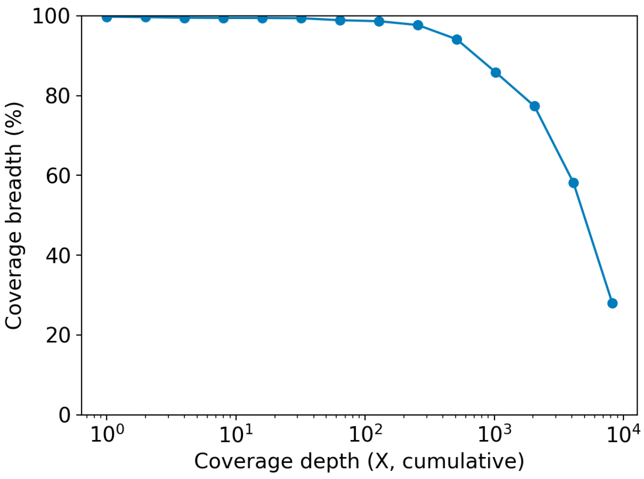

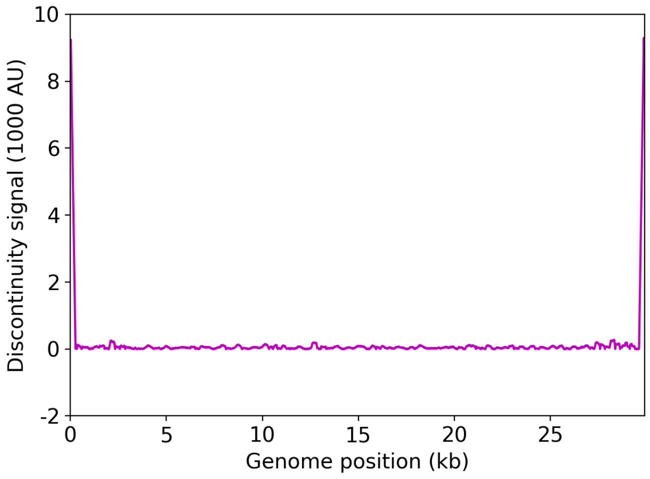


| | Uncovered coordinates (0X) | Poorly covered coordinates (<10X) |
|---|---|---|
| # Inaccessible genomic coordinates by kit design: | 121nt (0%) | 121nt (0%) |
| All genomic coordinates: | 65nt (0%) | 157nt (0%) |
| Common SNPs: | 0nt (0%) | 0nt (0%) |
| Diverse SNPs: | 7nt (3%) | 29nt (12%) |
| Rare SNPs: | 1nt (0%) | 10nt (1%) |

SNPs refer to the polymorphic sites currently in circulation that were detected out of recent GISAID entries. The sites that differ from the SC2 reference sequence are denoted as "common" if [90%, 100%] of the submissions carry this mutation, whereas those that are prevalent in [0%,10%] of the submissions are grouped under the "rare" category. The population is still diverse at the mutation sites that are observed in (10%,90%) of the entries and these coordinates are grouped under the "diverse" category.

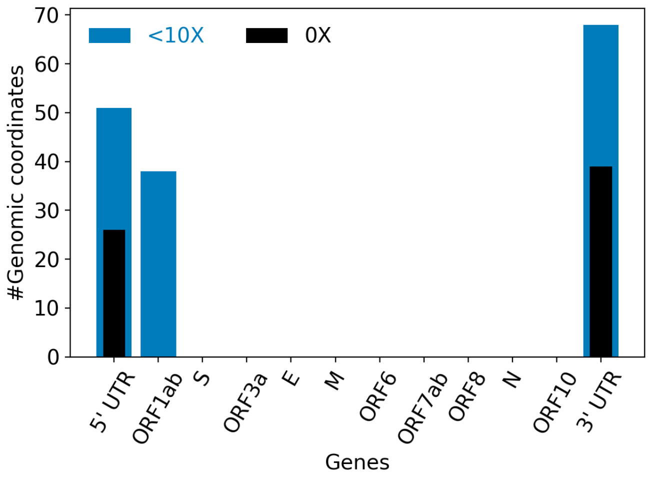


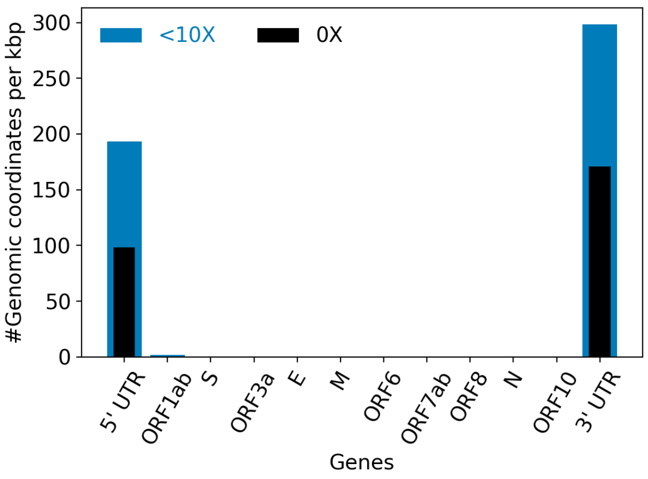


| | |
|---|---|
| Hits to SARS-Cov2 genome (kraken2): | 1925235 reads (94.77%) |
| Hits to human genome (kraken2): | 224 reads (0.01%) |
| Hits to synthetic sequences (kraken2, taxid 28384): | 51 reads (0.00%) |
| Most abundant organisms (kraken2, family level): | Coronaviridae (94.77%)<br>Staphylococcaceae (0.08%)<br>Hominidae (0.01%) |

| | |
|---|---|
| | Mycobacteriaceae (0.00%)<br>Pseudomonadaceae (0.00%) |

### Detected variants (Experimental)

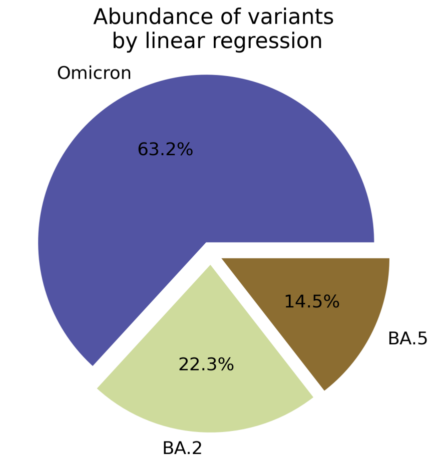


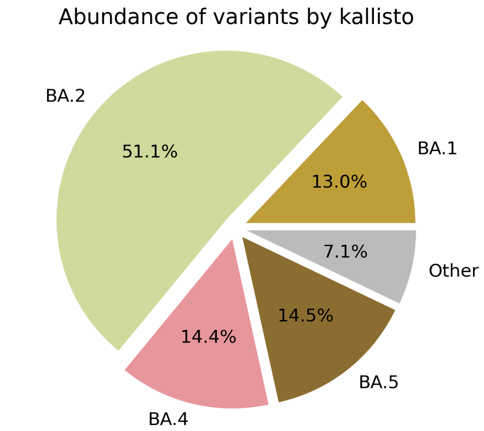


Based on deconvolution, B.1.1.529 is estimated to constitute 63.15% of the viral particles and hence is the most abundant variant in the sample. The $R^2$ for the linear regression was 0.62. Variants that were detected less than 5% were grouped under "Other"

Based on the consensus sequence of the observed reads, the "ensemble-averaged sequence" most closely resembles the BA.2 lineage. If this is a sample consisting of a single source of pathogens or an overwhelming majority of the different sources are infected with the same variant, the sample is dominated by this variant.

Based on mapping individual reads to the variant consensus sequences in the reference database, kallisto predicts that the sample is dominated by BA.2 lineage. Accuracy of this measure is expected to improve if the input data consists of long reads as opposed to convolution.

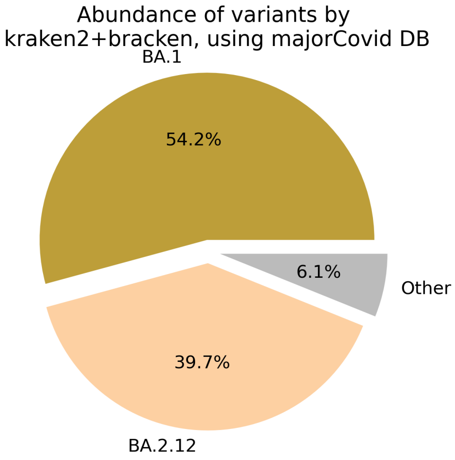


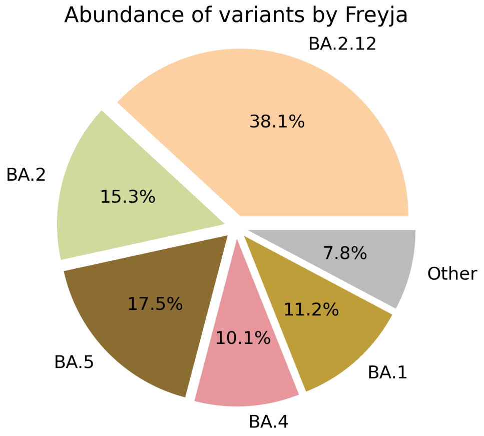

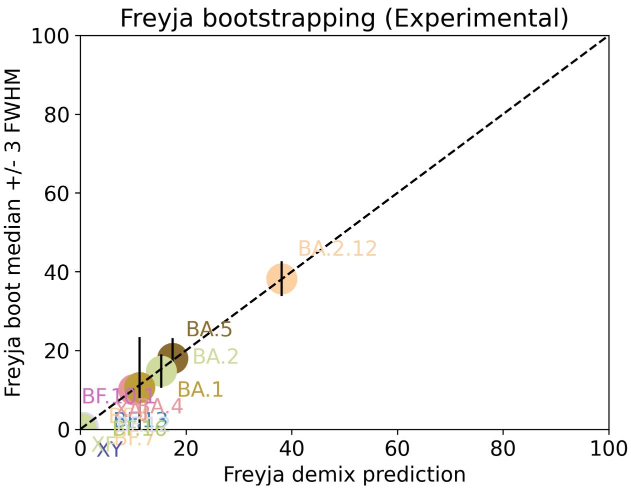


Under the assumption that the presence of a variant requires the detection of all respective mutations of the variant, the characteric mutations which support the presence of the respective variant are indicated in the respective column of the table. Numbers show the number of mutations detected, if any, and the number of mutations expected to be present based on the variant definitions.

| VOC | B.1.617.2 | BA.1 | BA.2 | BA.3 | BA.4 | BA.5 |
|---|---|---|---|---|---|---|
| Characteristic mutations detected | (3 of 13)<br>S:G142D<br>S:L452R<br>S:T478K | (15 of 26)<br>NUC:C15240T<br>NUC:C25000T<br>NUC:C25584T<br>NUC:T13195C<br>NUC:T5386G<br>ORF1AB:A2710T<br>ORF1AB:I3758V<br>ORF1AB:K856R<br>S:G446S<br>S:G496S<br>S:L981F<br>S:N856K<br>S:Q493R<br>S:T547K<br>S:T95I | (23 of 31)<br>N:S413R<br>NUC:A20055G<br>NUC:A9424G<br>NUC:C10198T<br>NUC:C12880T<br>NUC:C15714T<br>NUC:C25000T<br>NUC:C25584T<br>NUC:C26858T<br>NUC:C4321T<br>NUC:G10447A<br>ORF1AB:G1307S<br>ORF1AB:L3027F<br>ORF1AB:L3201F<br>ORF1AB:S135R<br>ORF1AB:T3090I<br>ORF1AB:T842I<br>S:D405N<br>S:Q493R<br>S:R408S<br>S:S371F<br>S:T19I<br>S:T376A | (12 of 21)<br>N:S413R<br>NUC:C12880T<br>NUC:C15714T<br>NUC:C26858T<br>NUC:G10447A<br>ORF1AB:G1307S<br>ORF1AB:S135R<br>ORF1AB:T3090I<br>S:D405N<br>S:G446S<br>S:Q493R<br>S:S371F | (24 of 31)<br>N:P151S<br>N:S413R<br>NUC:A20055G<br>NUC:C10198T<br>NUC:C12880T<br>NUC:C15714T<br>NUC:C25000T<br>NUC:C25584T<br>NUC:C26858T<br>NUC:C4321T<br>NUC:G10447A<br>NUC:G12160A<br>NUC:G27788T<br>ORF1AB:G1307S<br>ORF1AB:S135R<br>ORF1AB:T3090I<br>ORF1AB:T842I<br>S:D405N<br>S:F486V<br>S:L452R<br>S:S371F<br>S:T19I<br>S:T376A<br>S:V213G | (22 of 28)<br>M:D3N<br>N:S413R<br>NUC:A20055G<br>NUC:C10198T<br>NUC:C12880T<br>NUC:C15714T<br>NUC:C25000T<br>NUC:C25584T<br>NUC:C4321T<br>NUC:G10447A<br>NUC:G12160A<br>ORF1AB:G1307S<br>ORF1AB:S135R<br>ORF1AB:T3090I<br>ORF1AB:T842I<br>S:D405N<br>S:F486V<br>S:L452R<br>S:S371F<br>S:T19I<br>S:T376A<br>S:V213G |

Jaccard Index is a measure of similarity between two sets A and B, reaching the maximum value of 1 if A=B and minimum value of 0 if A ∩ B = {}. In the c(d) representation below, c represents the Jaccard index of the set of mutations that were experimentally detected for this sample as listed above, whereas d refers to the ideal value of the Jaccard index expected from complete genome coverage without any sequencing errors.

| | B.1.617.2 | BA.1 | BA.2 | BA.3 | BA.4 | BA.5 |
|---|---|---|---|---|---|---|
| B.1.617.2 | 1.00 (1.00) | 0.00 (0.00) | 0.00 (0.00) | 0.00 (0.00) | 0.04 (0.02) | 0.04 (0.03) |
| BA.1 | 0.00 (0.00) | 1.00 (1.00) | 0.09 (0.10) | 0.08 (0.21) | 0.05 (0.08) | 0.06 (0.08) |
| BA.2 | 0.00 (0.00) | 0.09 (0.10) | 1.00 (1.00) | 0.46 (0.33) | 0.62 (0.63) | 0.61 (0.59) |
| BA.3 | 0.00 (0.00) | 0.08 (0.21) | 0.46 (0.33) | 1.00 (1.00) | 0.38 (0.30) | 0.36 (0.29) |

| | | | | | | |
|---|---|---|---|---|---|---|
| BA.4 | 0.04 (0.02) | 0.05 (0.08) | 0.62 (0.63) | 0.38 (0.30) | 1.00 (1.00) | 0.84 (0.84) |
| BA.5 | 0.04 (0.03) | 0.06 (0.08) | 0.61 (0.59) | 0.36 (0.29) | 0.84 (0.84) | 1.00 (1.00) |

#### Detected mutations

Excluded from this pdf version due to file size limitations.

CFSAN/OAO
BIOSTATISTICS AND BIOINFORMATICS STAFF

### WASTEWATER SARS-COV2 ANALYSIS REPORT

| | |
|---|---|
| Sample name: | CFSANSMP000118328 |
| Date generated: | 2023-02-01, 20:06:45 EST |
| Timestamp of C-WAP version used: | Tue Jan 31 11:49:22 2023 -0500 |
| Executed by: | Jasmine Amirzadegan |
| Executed on: | 172.20.44.137 (aka n137.raven.cfsan) |

#### Sequencing summary

| | |
|---|---|
| Sequencing chemistry: | Missing with Missing |
| Source site: | Missing (?,?) |
| Sampling date: | Missing |
| Collected by: | Missing |
| Sequenced by: | Missing |
| Total number of reads: | 1905298 |
| Reads aligned: | 1798522 (94%) |
| Average read quality: | 36.6 |
| Average read length: | 149 |
| Reads passing filter: | 1724073 (90%) |
| Average read quality passing filter: | 36.9 |
| Average read length passing filter: | 149 |
| Average coverage passing filter: | 8590X |

A read passes filter if the read length after adaptor trimming ≥ 30 and minimum read quality ≥ 20 within a sliding window of width 4.

#### Overall sequence characteristics

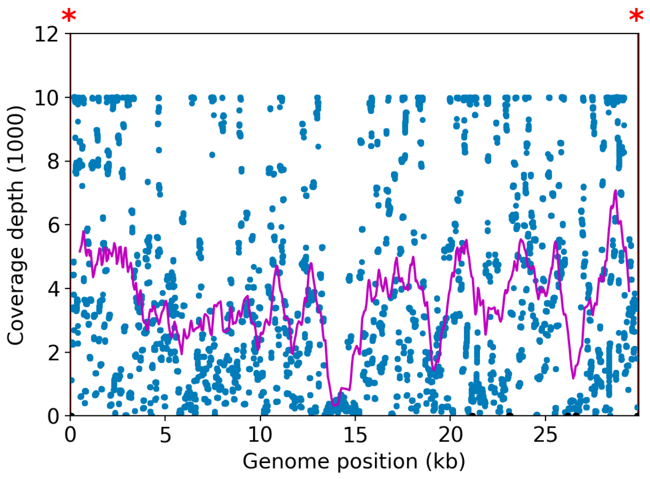


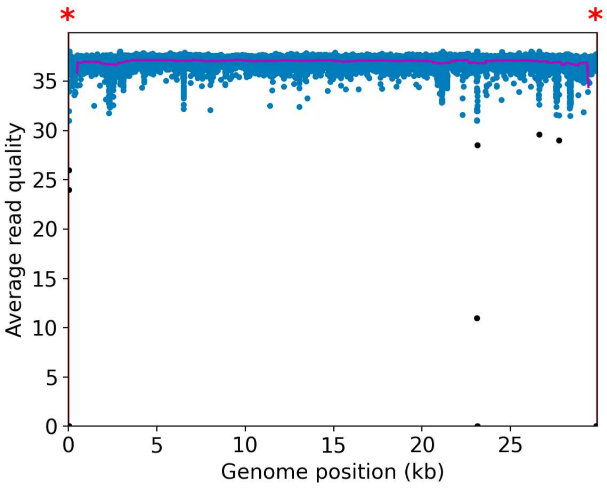


NOTE: The red shaded areas marked with a (*) are not covered by the design of the library preparation kit and hence excluded from analyses. Magenta curves represent moving average with a window width of 1kb.

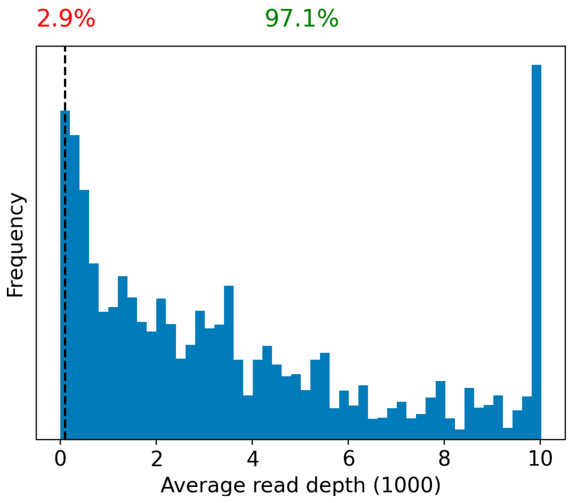


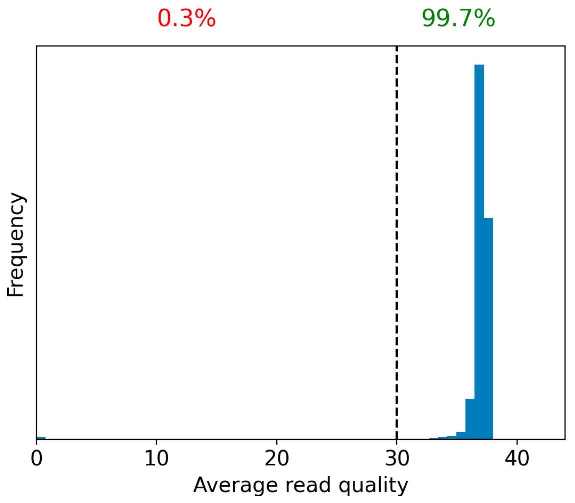


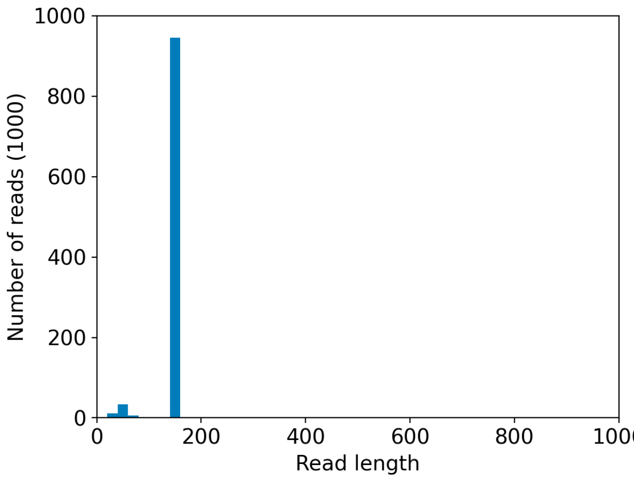


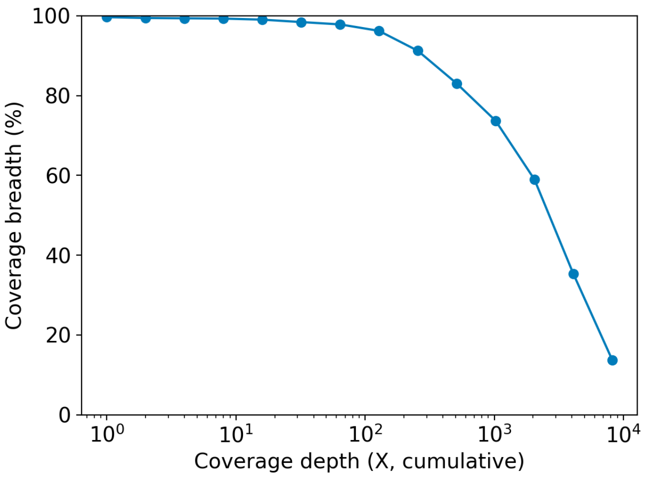

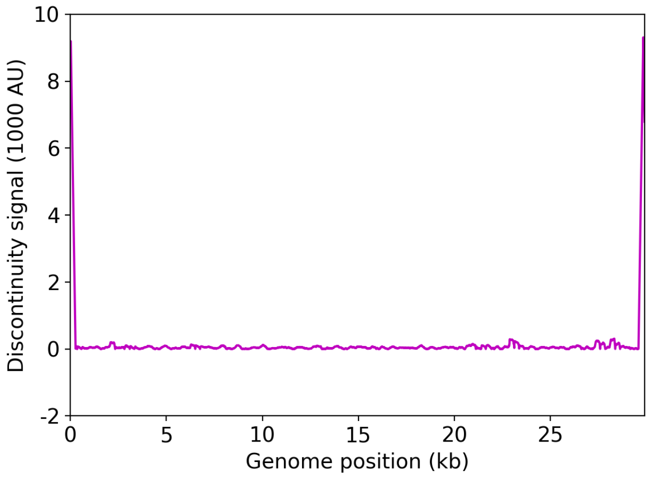


| | Uncovered coordinates (0X) | Poorly covered coordinates (<10X) |
|---|---|---|
| # Inaccessible genomic coordinates by kit design: | 121nt (0%) | 121nt (0%) |
| All genomic coordinates: | 97nt (0%) | 270nt (0%) |
| Common SNPs: | 0nt (0%) | 1nt (2%) |
| Diverse SNPs: | 28nt (12%) | 29nt (12%) |
| Rare SNPs: | 9nt (0%) | 47nt (5%) |

SNPs refer to the polymorphic sites currently in circulation that were detected out of recent GISAID entries. The sites that differ from the SC2 reference sequence are denoted as "common" if [90%, 100%] of the submissions carry this mutation, whereas those that are prevalent in [0%,10%] of the submissions are grouped under the "rare" category. The population is still diverse at the mutation sites that are observed in (10%,90%) of the entries and these coordinates are grouped under the "diverse" category.

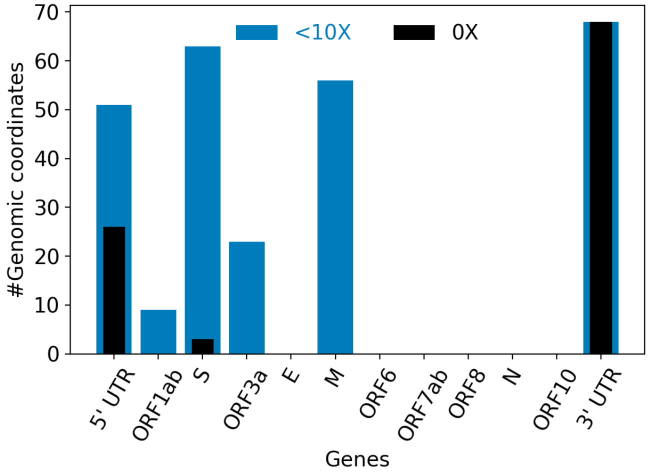


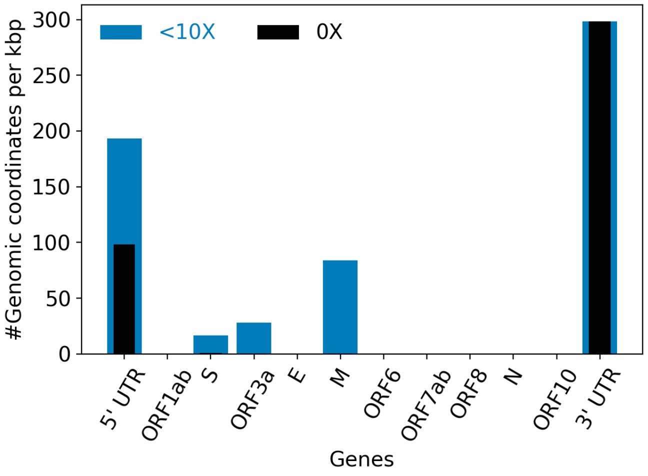


| | |
|---|---|
| Hits to SARS-Cov2 genome (kraken2): | 908271 reads (95.34%) |
| Hits to human genome (kraken2): | 94 reads (0.01%) |
| Hits to synthetic sequences (kraken2, taxid 28384): | 15 reads (0.00%) |
| Most abundant organisms (kraken2, family level): | Coronaviridae (95.34%)<br>Arcobacteraceae (0.04%)<br>Hominidae (0.01%) |

| | |
|---|---|
| | Staphylococcaceae (0.01%)<br>Flavobacteriaceae (0.00%) |

### Detected variants (Experimental)

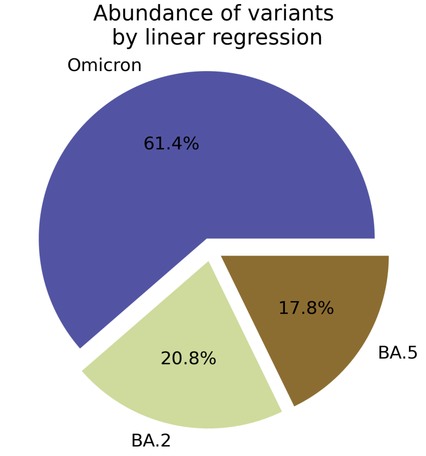


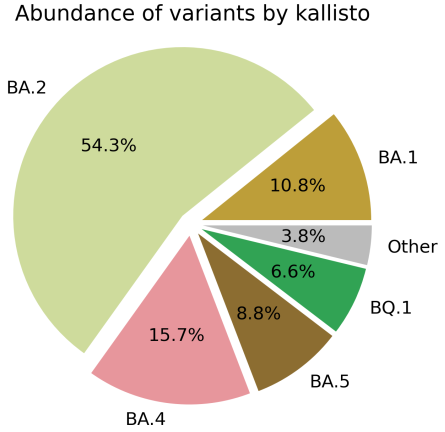


Based on deconvolution, B.1.1.529 is estimated to constitute 61.41% of the viral particles and hence is the most abundant variant in the sample. The $R^2$ for the linear regression was 0.60. Variants that were detected less than 5% were grouped under "Other"

Based on the consensus sequence of the observed reads, the "ensemble-averaged sequence" most closely resembles the BA.2 lineage. If this is a sample consisting of a single source of pathogens or an overwhelming majority of the different sources are infected with the same variant, the sample is dominated by this variant.

Based on mapping individual reads to the variant consensus sequences in the reference database, kallisto predicts that the sample is dominated by BA.2 lineage. Accuracy of this measure is expected to improve if the input data consists of long reads as opposed to convolution.

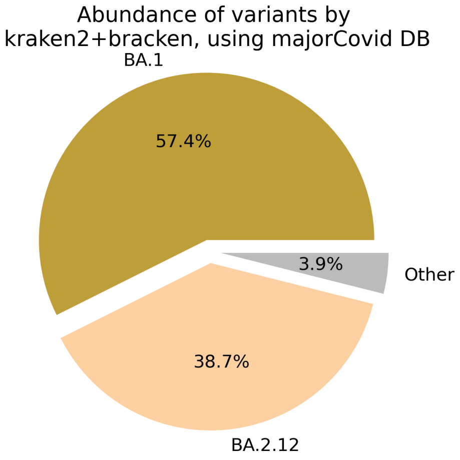


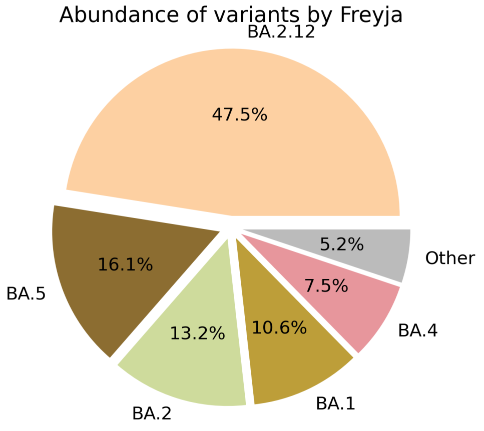

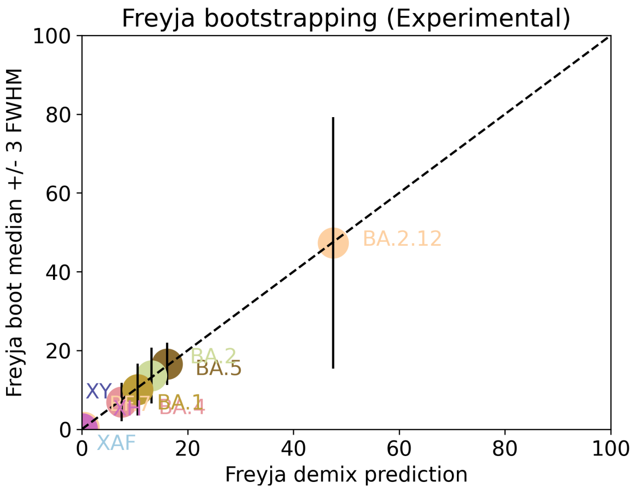


Under the assumption that the presence of a variant requires the detection of all respective mutations of the variant, the characteric mutations which support the presence of the respective variant are indicated in the respective column of the table. Numbers show the number of mutations detected, if any, and the number of mutations expected to be present based on the variant definitions.

| VOC | B.1.617.2 | BA.1 | BA.2 | BA.3 | BA.4 | BA.5 |
|---|---|---|---|---|---|---|
| Characteristic mutations detected | (3 of 13)<br>S:G142D<br>S:L452R<br>S:T478K | (13 of 26)<br>NUC:C15240T<br>NUC:C25000T<br>NUC:C25584T<br>NUC:T13195C<br>NUC:T5386G<br>ORF1AB:A2710T<br>ORF1AB:K856R<br>S:G446S<br>S:G496S<br>S:L981F<br>S:N856K<br>S:Q493R<br>S:T95I | (22 of 31)<br>N:S413R<br>NUC:A20055G<br>NUC:C10198T<br>NUC:C12880T<br>NUC:C15714T<br>NUC:C25000T<br>NUC:C25584T<br>NUC:C26858T<br>NUC:C4321T<br>NUC:G10447A<br>ORF1AB:G1307S<br>ORF1AB:L3027F<br>ORF1AB:L3201F<br>ORF1AB:S135R<br>ORF1AB:T3090I<br>ORF1AB:T842I<br>S:D405N<br>S:Q493R<br>S:R408S<br>S:S371F<br>S:T19I<br>S:T376A | (12 of 21)<br>N:S413R<br>NUC:C12880T<br>NUC:C15714T<br>NUC:C26858T<br>NUC:G10447A<br>ORF1AB:G1307S<br>ORF1AB:S135R<br>ORF1AB:T3090I<br>S:D405N<br>S:G446S<br>S:Q493R<br>S:S371F | (24 of 31)<br>N:P151S<br>N:S413R<br>NUC:A20055G<br>NUC:C10198T<br>NUC:C12880T<br>NUC:C15714T<br>NUC:C25000T<br>NUC:C25584T<br>NUC:C26858T<br>NUC:C4321T<br>NUC:G10447A<br>NUC:G12160A<br>NUC:G27788T<br>ORF1AB:G1307S<br>ORF1AB:S135R<br>ORF1AB:T3090I<br>ORF1AB:T842I<br>S:D405N<br>S:F486V<br>S:L452R<br>S:S371F<br>S:T19I<br>S:T376A<br>S:V213G | (22 of 28)<br>M:D3N<br>N:S413R<br>NUC:A20055G<br>NUC:C10198T<br>NUC:C12880T<br>NUC:C15714T<br>NUC:C25000T<br>NUC:C25584T<br>NUC:C4321T<br>NUC:G10447A<br>NUC:G12160A<br>ORF1AB:G1307S<br>ORF1AB:S135R<br>ORF1AB:T3090I<br>ORF1AB:T842I<br>S:D405N<br>S:F486V<br>S:L452R<br>S:S371F<br>S:T19I<br>S:T376A<br>S:V213G |

Jaccard Index is a measure of similarity between two sets A and B, reaching the maximum value of 1 if A=B and minimum value of 0 if A ∩ B = {}. In the c(d) representation below, c represents the Jaccard index of the set of mutations that were experimentally detected for this sample as listed above, whereas d refers to the ideal value of the Jaccard index expected from complete genome coverage without any sequencing errors.

| | B.1.617.2 | BA.1 | BA.2 | BA.3 | BA.4 | BA.5 |
|---|---|---|---|---|---|---|
| B.1.617.2 | 1.00 (1.00) | 0.00 (0.00) | 0.00 (0.00) | 0.00 (0.00) | 0.04 (0.02) | 0.04 (0.03) |
| BA.1 | 0.00 (0.00) | 1.00 (1.00) | 0.09 (0.10) | 0.09 (0.21) | 0.06 (0.08) | 0.06 (0.08) |
| BA.2 | 0.00 (0.00) | 0.09 (0.10) | 1.00 (1.00) | 0.48 (0.33) | 0.64 (0.63) | 0.63 (0.59) |
| BA.3 | 0.00 (0.00) | 0.09 (0.21) | 0.48 (0.33) | 1.00 (1.00) | 0.38 (0.30) | 0.36 (0.29) |

| | | | | | | |
|---|---|---|---|---|---|---|
| BA.4 | 0.04 (0.02) | 0.06 (0.08) | 0.64 (0.63) | 0.38 (0.30) | 1.00 (1.00) | 0.84 (0.84) |
| BA.5 | 0.04 (0.03) | 0.06 (0.08) | 0.63 (0.59) | 0.36 (0.29) | 0.84 (0.84) | 1.00 (1.00) |

### Detected mutations

Excluded from this pdf version due to file size limitations.

CFSAN/OAO
BIOSTATISTICS AND BIOINFORMATICS STAFF

### WASTEWATER SARS-COV2 ANALYSIS REPORT

| | |
|---|---|
| Sample name: | CFSANSMP000118506 |
| Date generated: | 2023-02-01, 20:05:44 EST |
| Timestamp of C-WAP version used: | Tue Jan 31 11:49:22 2023 -0500 |
| Executed by: | Jasmine Amirzadegan |
| Executed on: | 172.20.44.127 (aka n127.raven.cfsan) |

#### Sequencing summary

| | |
|---|---|
| Sequencing chemistry: | Missing with Missing |
| Source site: | Missing (?,?) |
| Sampling date: | Missing |
| Collected by: | Missing |
| Sequenced by: | Missing |
| Total number of reads: | 1300104 |
| Reads aligned: | 1160847 (89%) |
| Average read quality: | 36.4 |
| Average read length: | 149 |
| Reads passing filter: | 1097393 (84%) |
| Average read quality passing filter: | 36.7 |
| Average read length passing filter: | 149 |
| Average coverage passing filter: | 5468X |

A read passes filter if the read length after adaptor trimming ≥ 30 and minimum read quality ≥ 20 within a sliding window of width 4.

#### Overall sequence characteristics

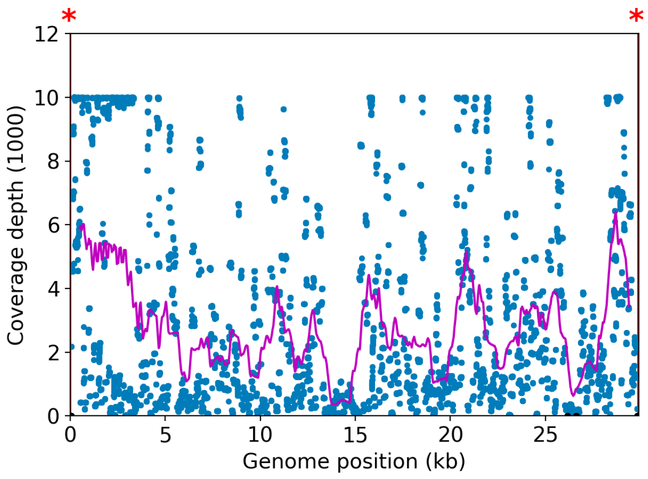


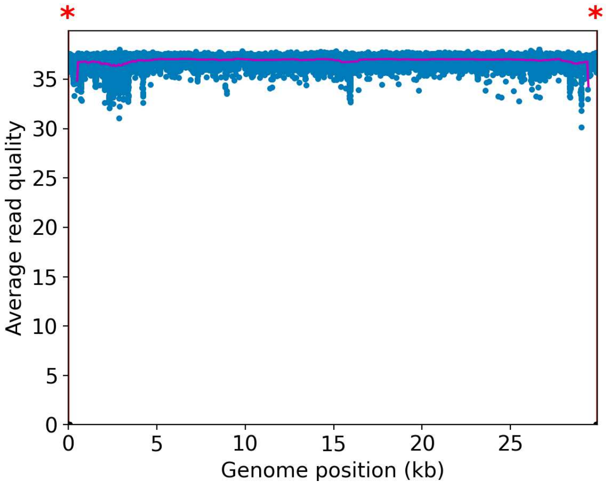


NOTE: The red shaded areas marked with a (*) are not covered by the design of the library preparation kit and hence excluded from analyses. Magenta curves represent moving average with a window width of 1kb.

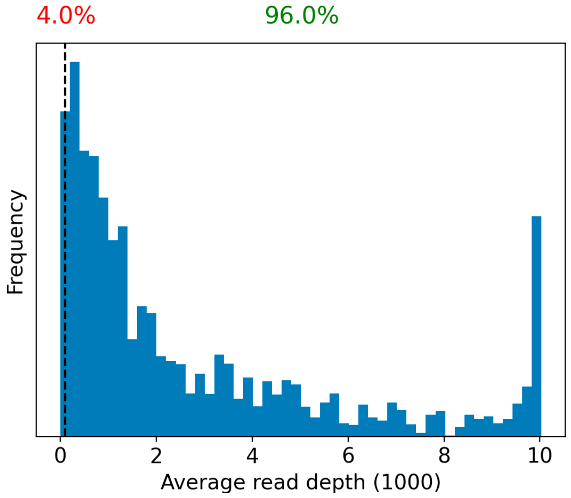


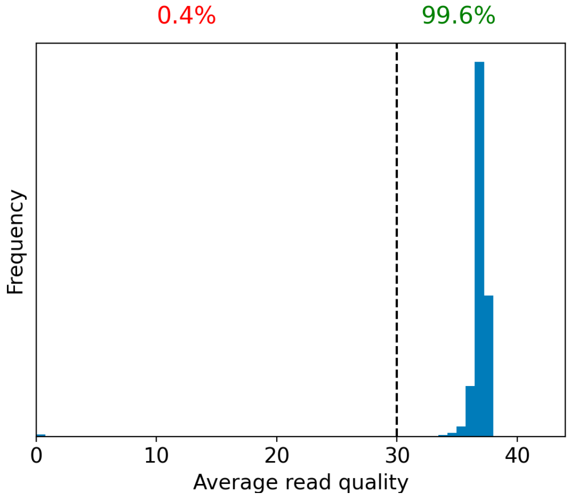


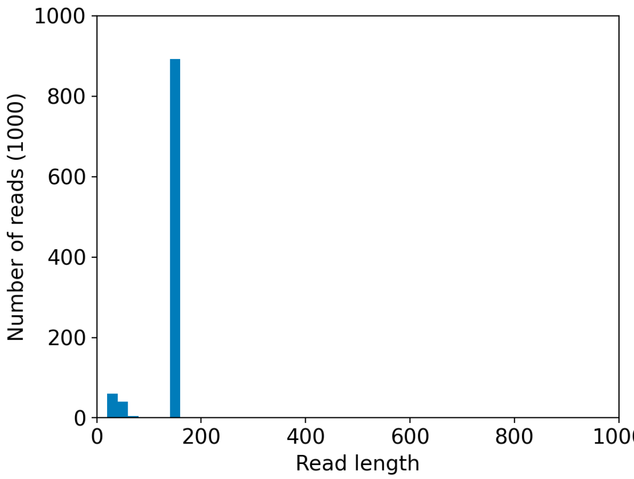


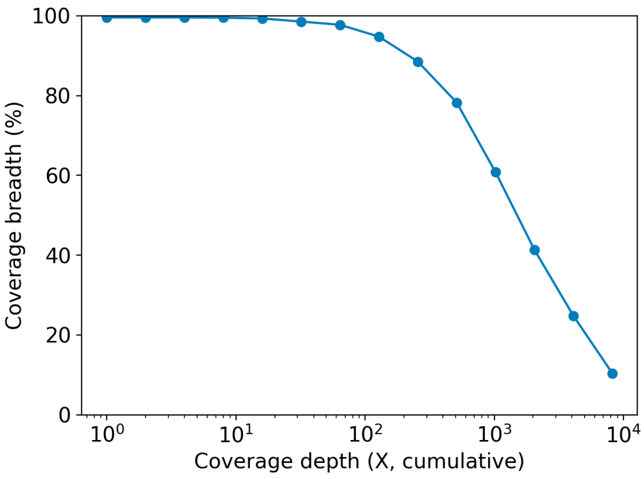

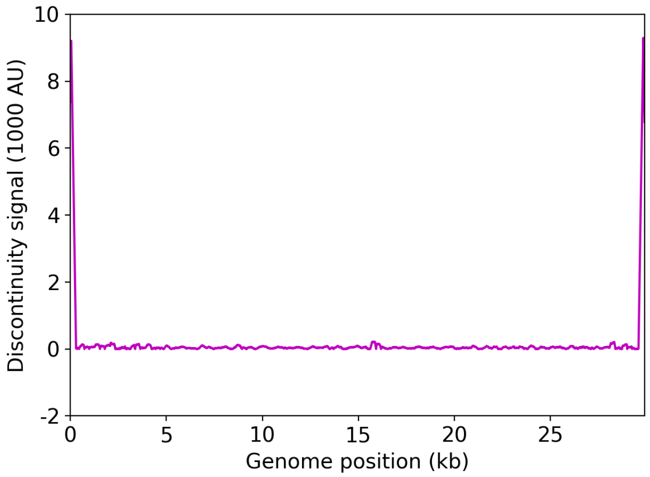


| | Uncovered coordinates (0X) | Poorly covered coordinates (<10X) |
|---|---|---|
| # Inaccessible genomic coordinates by kit design: | 121nt (0%) | 121nt (0%) |
| All genomic coordinates: | 119nt (0%) | 159nt (0%) |
| Common SNPs: | 0nt (0%) | 0nt (0%) |
| Diverse SNPs: | 29nt (12%) | 29nt (12%) |
| Rare SNPs: | 10nt (1%) | 10nt (1%) |

SNPs refer to the polymorphic sites currently in circulation that were detected out of recent GISAID entries. The sites that differ from the SC2 reference sequence are denoted as "common" if [90%, 100%] of the submissions carry this mutation, whereas those that are prevalent in [0%,10%] of the submissions are grouped under the "rare" category. The population is still diverse at the mutation sites that are observed in (10%,90%) of the entries and these coordinates are grouped under the "diverse" category.

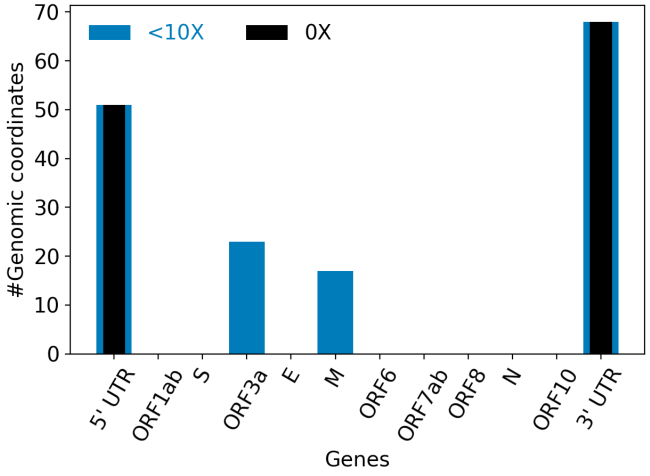


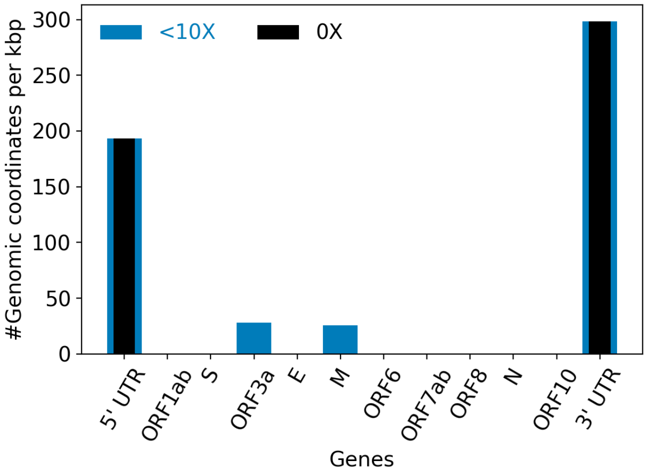


| | |
|---|---|
| Hits to SARS-Cov2 genome (kraken2): | 593765 reads (91.34%) |
| Hits to human genome (kraken2): | 140 reads (0.02%) |
| Hits to synthetic sequences (kraken2, taxid 28384): | 83 reads (0.01%) |
| Most abundant organisms (kraken2, family level): | Coronaviridae (91.34%)<br>Enterobacteriaceae (0.06%)<br>Hominidae (0.02%) |

| | |
|---|---|
| | Staphylococcaceae (0.01%)<br>Fusobacteriaceae (0.00%) |

### Detected variants (Experimental)

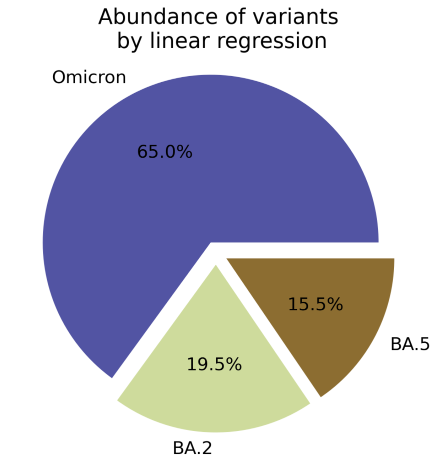


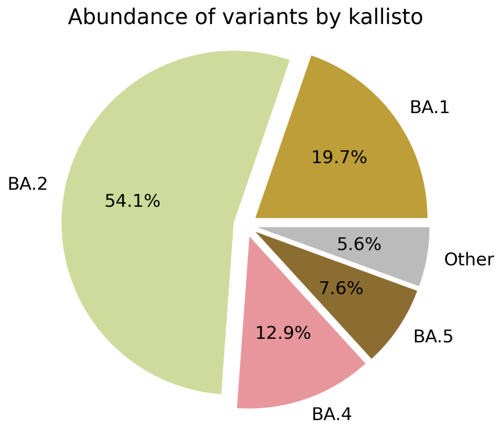


Based on deconvolution, B.1.1.529 is estimated to constitute 64.97% of the viral particles and hence is the most abundant variant in the sample. The $R^2$ for the linear regression was 0.63. Variants that were detected less than 5% were grouped under "Other"

Based on the consensus sequence of the observed reads, the "ensemble-averaged sequence" most closely resembles the BA.2 lineage. If this is a sample consisting of a single source of pathogens or an overwhelming majority of the different sources are infected with the same variant, the sample is dominated by this variant.

Based on mapping individual reads to the variant consensus sequences in the reference database, kallisto predicts that the sample is dominated by BA.2 lineage. Accuracy of this measure is expected to improve if the input data consists of long reads as opposed to convolution.

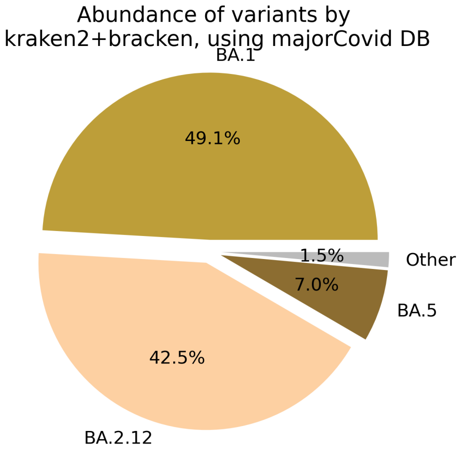


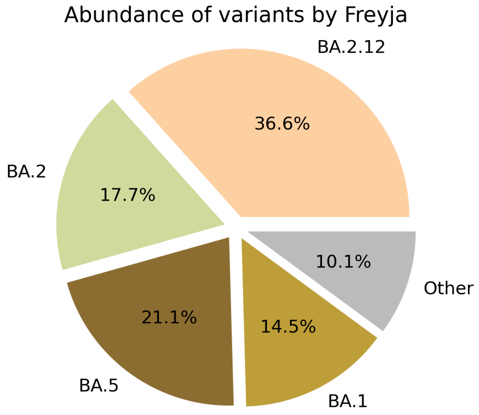

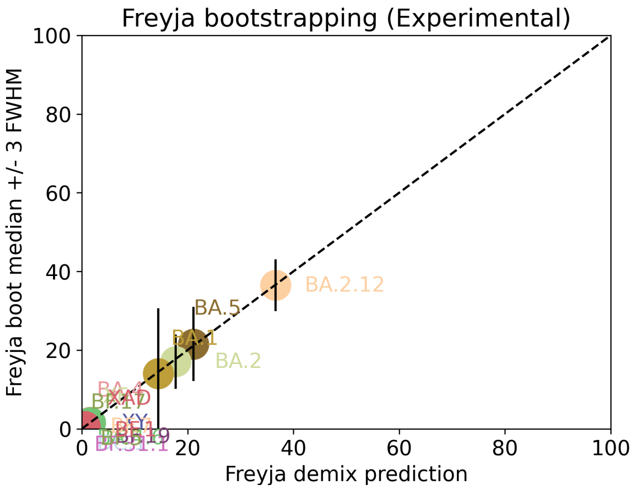


Under the assumption that the presence of a variant requires the detection of all respective mutations of the variant, the characteric mutations which support the presence of the respective variant are indicated in the respective column of the table. Numbers show the number of mutations detected, if any, and the number of mutations expected to be present based on the variant definitions.

| VOC | B.1.617.2 | BA.1 | BA.2 | BA.3 | BA.4 | BA.5 |
|---|---|---|---|---|---|---|
| Characteristic mutations detected | (3 of 13)<br>S:G142D<br>S:L452R<br>S:T478K | (14 of 26)<br>NUC:C15240T<br>NUC:C25000T<br>NUC:C25584T<br>NUC:T13195C<br>NUC:T5386G<br>ORF1AB:A2710T<br>ORF1AB:I3758V<br>S:G446S<br>S:G496S<br>S:L981F<br>S:N856K<br>S:Q493R<br>S:T547K<br>S:T95I | (23 of 31)<br>N:S413R<br>NUC:A20055G<br>NUC:A9424G<br>NUC:C10198T<br>NUC:C12880T<br>NUC:C15714T<br>NUC:C25000T<br>NUC:C25584T<br>NUC:C26858T<br>NUC:C4321T<br>NUC:G10447A<br>ORF1AB:G1307S<br>ORF1AB:L3027F<br>ORF1AB:L3201F<br>ORF1AB:S135R<br>ORF1AB:T3090I<br>ORF1AB:T842I<br>S:D405N<br>S:Q493R<br>S:R408S<br>S:S371F<br>S:T19I<br>S:T376A | (12 of 21)<br>N:S413R<br>NUC:C12880T<br>NUC:C15714T<br>NUC:C26858T<br>NUC:G10447A<br>ORF1AB:G1307S<br>ORF1AB:S135R<br>ORF1AB:T3090I<br>S:D405N<br>S:G446S<br>S:Q493R<br>S:S371F | (24 of 31)<br>N:P151S<br>N:S413R<br>NUC:A20055G<br>NUC:C10198T<br>NUC:C12880T<br>NUC:C15714T<br>NUC:C25000T<br>NUC:C25584T<br>NUC:C26858T<br>NUC:C4321T<br>NUC:G10447A<br>NUC:G12160A<br>NUC:G27788T<br>ORF1AB:G1307S<br>ORF1AB:S135R<br>ORF1AB:T3090I<br>ORF1AB:T842I<br>S:D405N<br>S:F486V<br>S:L452R<br>S:S371F<br>S:T19I<br>S:T376A<br>S:V213G | (22 of 28)<br>M:D3N<br>N:S413R<br>NUC:A20055G<br>NUC:C10198T<br>NUC:C12880T<br>NUC:C15714T<br>NUC:C25000T<br>NUC:C25584T<br>NUC:C4321T<br>NUC:G10447A<br>NUC:G12160A<br>ORF1AB:G1307S<br>ORF1AB:S135R<br>ORF1AB:T3090I<br>ORF1AB:T842I<br>S:D405N<br>S:F486V<br>S:L452R<br>S:S371F<br>S:T19I<br>S:T376A<br>S:V213G |

Jaccard Index is a measure of similarity between two sets A and B, reaching the maximum value of 1 if A=B and minimum value of 0 if A ∩ B = {}. In the c(d) representation below, c represents the Jaccard index of the set of mutations that were experimentally detected for this sample as listed above, whereas d refers to the ideal value of the Jaccard index expected from complete genome coverage without any sequencing errors.

| | B.1.617.2 | BA.1 | BA.2 | BA.3 | BA.4 | BA.5 |
|---|---|---|---|---|---|---|
| B.1.617.2 | 1.00 (1.00) | 0.00 (0.00) | 0.00 (0.00) | 0.00 (0.00) | 0.04 (0.02) | 0.04 (0.03) |
| BA.1 | 0.00 (0.00) | 1.00 (1.00) | 0.09 (0.10) | 0.08 (0.21) | 0.06 (0.08) | 0.06 (0.08) |
| BA.2 | 0.00 (0.00) | 0.09 (0.10) | 1.00 (1.00) | 0.46 (0.33) | 0.62 (0.63) | 0.61 (0.59) |
| BA.3 | 0.00 (0.00) | 0.08 (0.21) | 0.46 (0.33) | 1.00 (1.00) | 0.38 (0.30) | 0.36 (0.29) |

| | | | | | | |
|---|---|---|---|---|---|---|
| BA.4 | 0.04 (0.02) | 0.06 (0.08) | 0.62 (0.63) | 0.38 (0.30) | 1.00 (1.00) | 0.84 (0.84) |
| BA.5 | 0.04 (0.03) | 0.06 (0.08) | 0.61 (0.59) | 0.36 (0.29) | 0.84 (0.84) | 1.00 (1.00) |

#### Detected mutations

Excluded from this pdf version due to file size limitations.

#### CFSAN/OAO
#### BIOSTATISTICS AND BIOINFORMATICS STAFF

### WASTEWATER SARS-COV2 ANALYSIS REPORT

| | |
|---|---|
| Sample name: | CFSANSMP000118507 |
| Date generated: | 2023-02-01, 20:09:26 EST |
| Timestamp of C-WAP version used: | Tue Jan 31 11:49:22 2023 -0500 |
| Executed by: | Jasmine Amirzadegan |
| Executed on: | 172.20.44.108 (aka n108.raven.cfsan) |

#### Sequencing summary

| | |
|---|---|
| Sequencing chemistry: | Missing with Missing |
| Source site: | Missing (?,?) |
| Sampling date: | Missing |
| Collected by: | Missing |
| Sequenced by: | Missing |
| Total number of reads: | 2106332 |
| Reads aligned: | 1930920 (91%) |
| Average read quality: | 36.6 |
| Average read length: | 149 |
| Reads passing filter: | 1848070 (87%) |
| Average read quality passing filter: | 36.9 |
| Average read length passing filter: | 149 |
| Average coverage passing filter: | 9208X |

A read passes filter if the read length after adaptor trimming ≥ 30 and minimum read quality ≥ 20 within a sliding window of width 4.

#### Overall sequence characteristics

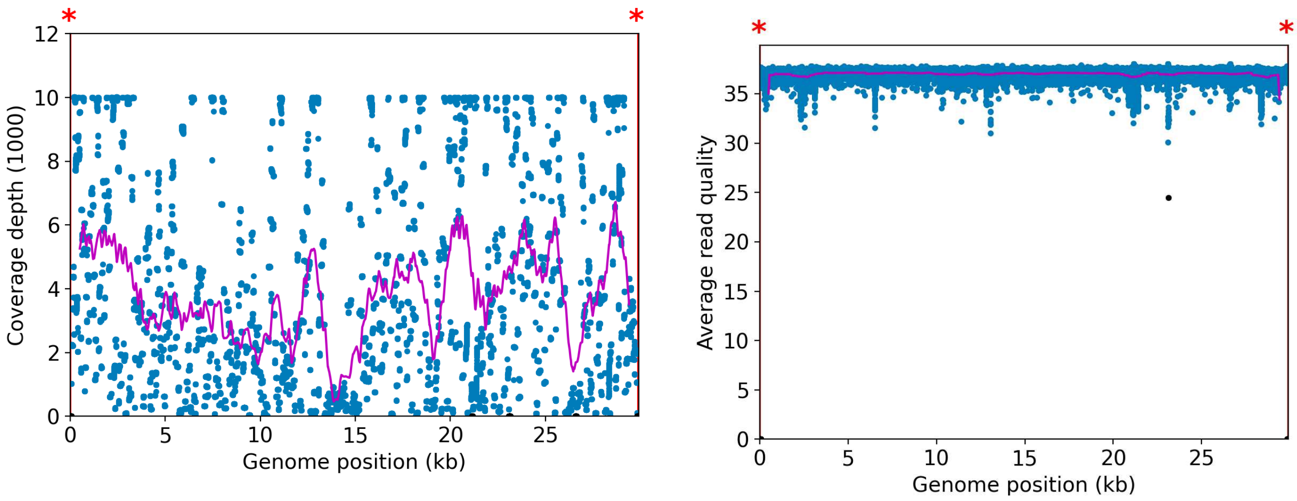


NOTE: The red shaded areas marked with a (*) are not covered by the design of the library preparation kit and hence excluded from analyses. Magenta curves represent moving average with a window width of 1kb.

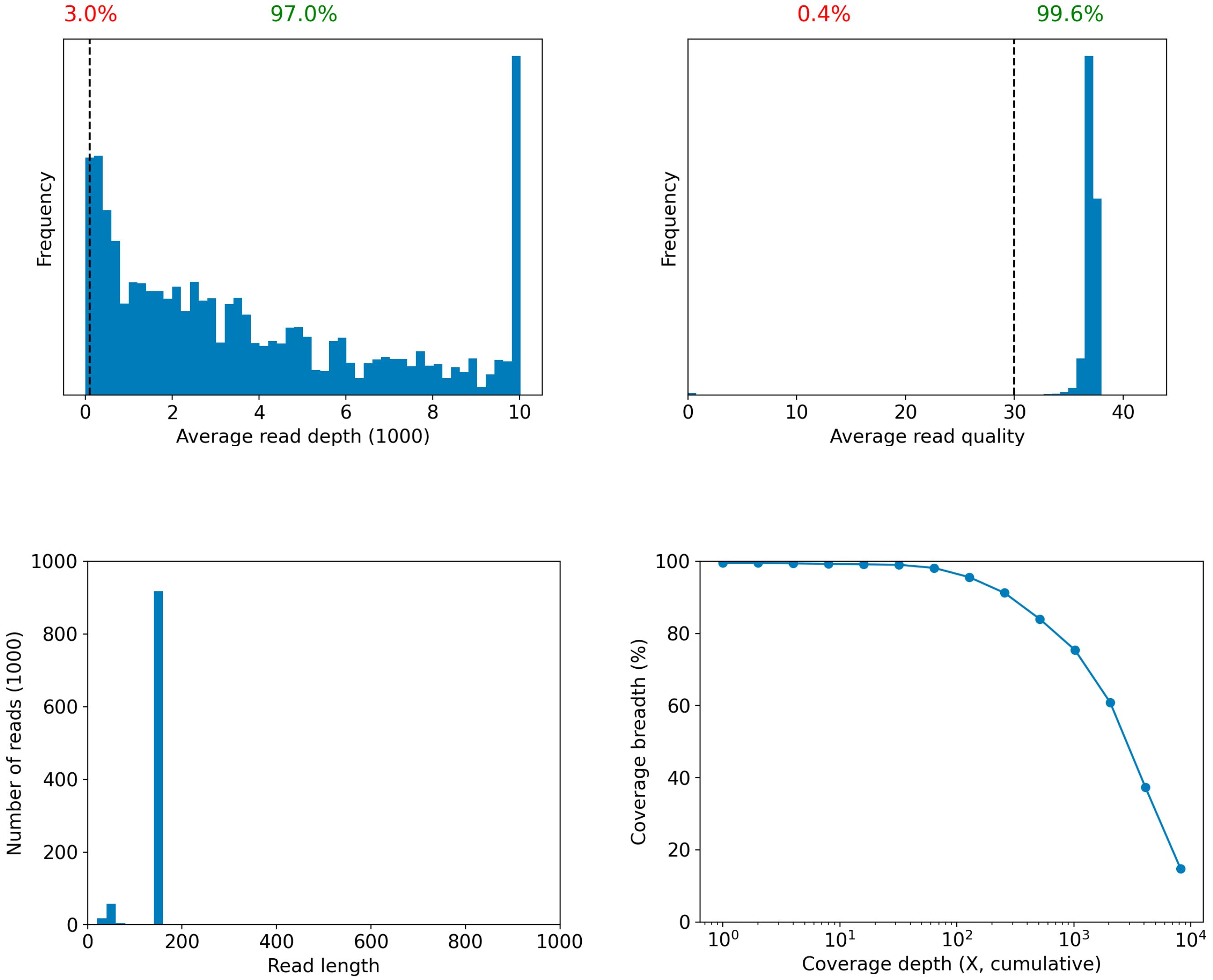

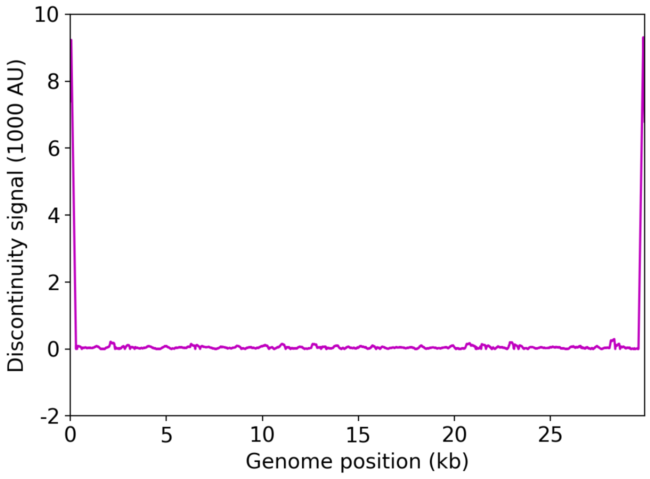


| | Uncovered coordinates (0X) | Poorly covered coordinates (<10X) |
|---|---|---|
| # Inaccessible genomic coordinates by kit design: | 121nt (0%) | 121nt (0%) |
| All genomic coordinates: | 119nt (0%) | 214nt (0%) |
| Common SNPs: | 0nt (0%) | 0nt (0%) |
| Diverse SNPs: | 29nt (12%) | 29nt (12%) |
| Rare SNPs: | 10nt (1%) | 46nt (4%) |

SNPs refer to the polymorphic sites currently in circulation that were detected out of recent GISAID entries. The sites that differ from the SC2 reference sequence are denoted as "common" if [90%, 100%] of the submissions carry this mutation, whereas those that are prevalent in [0%,10%] of the submissions are grouped under the "rare" category. The population is still diverse at the mutation sites that are observed in (10%,90%) of the entries and these coordinates are grouped under the "diverse" category.

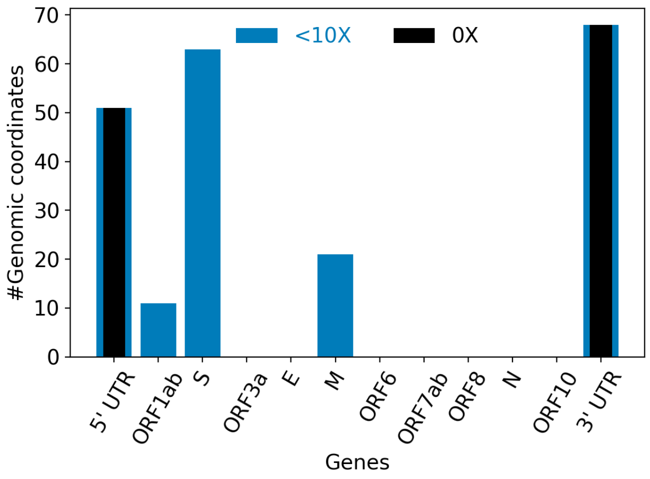


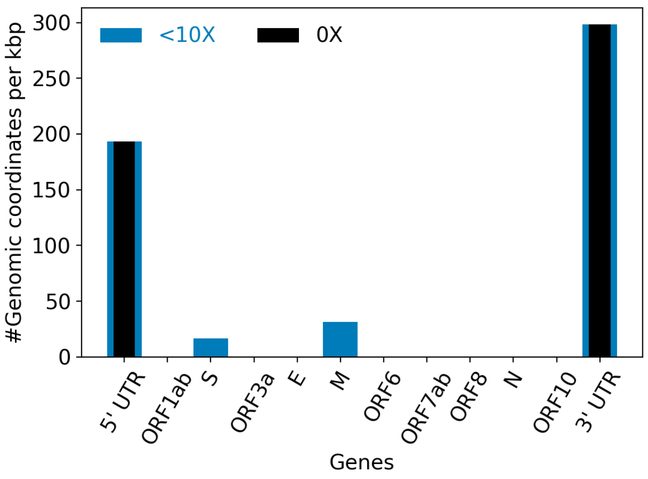


| | |
|---|---|
| Hits to SARS-Cov2 genome (kraken2): | 982969 reads (93.33%) |
| Hits to human genome (kraken2): | 192 reads (0.02%) |
| Hits to synthetic sequences (kraken2, taxid 28384): | 0 reads (0.00%) |
| Most abundant organisms (kraken2, family level): | Coronaviridae (93.33%)<br>Hominidae (0.02%)<br>Staphylococcaceae (0.01%) |

| | |
|---|---|
| | Lachnospiraceae (0.01%)<br>Enterobacteriaceae (0.00%) |

### Detected variants (Experimental)

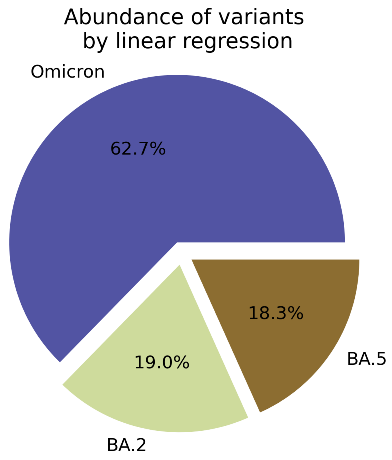


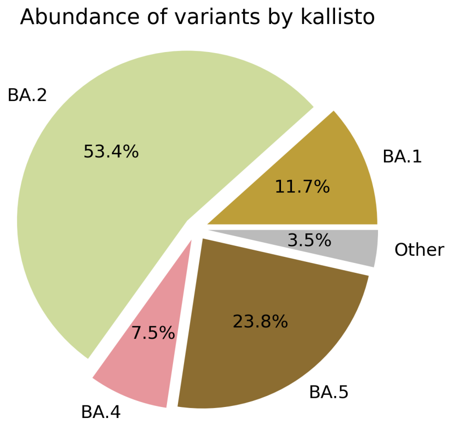


Based on deconvolution, B.1.1.529 is estimated to constitute 62.68% of the viral particles and hence is the most abundant variant in the sample. The $R^2$ for the linear regression was 0.61. Variants that were detected less than 5% were grouped under "Other"

Based on the consensus sequence of the observed reads, the "ensemble-averaged sequence" most closely resembles the BA.2 lineage. If this is a sample consisting of a single source of pathogens or an overwhelming majority of the different sources are infected with the same variant, the sample is dominated by this variant.

Based on mapping individual reads to the variant consensus sequences in the reference database, kallisto predicts that the sample is dominated by BA.2 lineage. Accuracy of this measure is expected to improve if the input data consists of long reads as opposed to convolution.

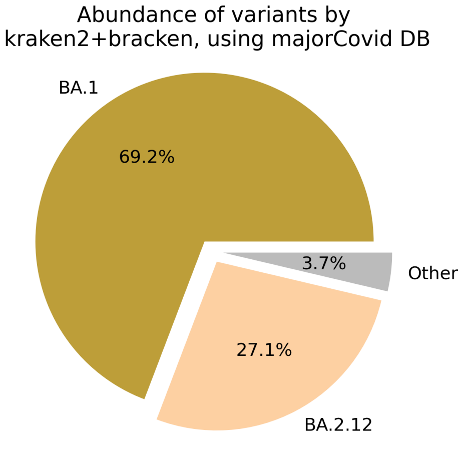


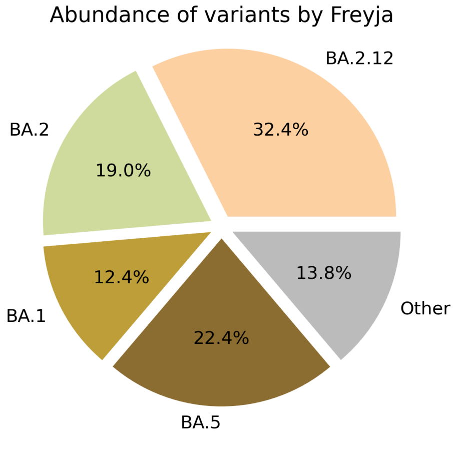

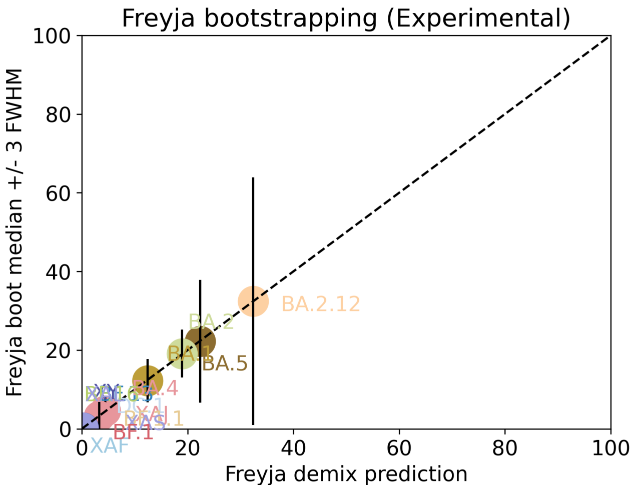


Under the assumption that the presence of a variant requires the detection of all respective mutations of the variant, the characteric mutations which support the presence of the respective variant are indicated in the respective column of the table. Numbers show the number of mutations detected, if any, and the number of mutations expected to be present based on the variant definitions.

| VOC | B.1.617.2 | BA.1 | BA.2 | BA.3 | BA.4 | BA.5 |
|---|---|---|---|---|---|---|
| Characteristic mutations detected | (3 of 13)<br>S:G142D<br>S:L452R<br>S:T478K | (14 of 26)<br>NUC:C15240T<br>NUC:C25000T<br>NUC:C25584T<br>NUC:T13195C<br>NUC:T5386G<br>ORF1AB:A2710T<br>ORF1AB:I3758V<br>S:A67V<br>S:G446S<br>S:G496S<br>S:L981F<br>S:N856K<br>S:Q493R<br>S:T95I | (23 of 31)<br>N:S413R<br>NUC:A20055G<br>NUC:A9424G<br>NUC:C10198T<br>NUC:C12880T<br>NUC:C15714T<br>NUC:C25000T<br>NUC:C25584T<br>NUC:C26858T<br>NUC:C4321T<br>NUC:G10447A<br>ORF1AB:G1307S<br>ORF1AB:L3027F<br>ORF1AB:L3201F<br>ORF1AB:S135R<br>ORF1AB:T3090I<br>ORF1AB:T842I<br>S:D405N<br>S:Q493R<br>S:R408S<br>S:S371F<br>S:T19I<br>S:T376A | (13 of 21)<br>N:S413R<br>NUC:C12880T<br>NUC:C15714T<br>NUC:C26858T<br>NUC:G10447A<br>ORF1AB:G1307S<br>ORF1AB:S135R<br>ORF1AB:T3090I<br>S:A67V<br>S:D405N<br>S:G446S<br>S:Q493R<br>S:S371F | (24 of 31)<br>N:P151S<br>N:S413R<br>NUC:A20055G<br>NUC:C10198T<br>NUC:C12880T<br>NUC:C15714T<br>NUC:C25000T<br>NUC:C25584T<br>NUC:C26858T<br>NUC:C4321T<br>NUC:G10447A<br>NUC:G12160A<br>NUC:G27788T<br>ORF1AB:G1307S<br>ORF1AB:S135R<br>ORF1AB:T3090I<br>ORF1AB:T842I<br>S:D405N<br>S:F486V<br>S:L452R<br>S:S371F<br>S:T19I<br>S:T376A<br>S:V213G | (22 of 28)<br>M:D3N<br>N:S413R<br>NUC:A20055G<br>NUC:C10198T<br>NUC:C12880T<br>NUC:C15714T<br>NUC:C25000T<br>NUC:C25584T<br>NUC:C4321T<br>NUC:G10447A<br>NUC:G12160A<br>ORF1AB:G1307S<br>ORF1AB:S135R<br>ORF1AB:T3090I<br>ORF1AB:T842I<br>S:D405N<br>S:F486V<br>S:L452R<br>S:S371F<br>S:T19I<br>S:T376A<br>S:V213G |

Jaccard Index is a measure of similarity between two sets A and B, reaching the maximum value of 1 if A=B and minimum value of 0 if A ∩ B = {}. In the c(d) representation below, c represents the Jaccard index of the set of mutations that were experimentally detected for this sample as listed above, whereas d refers to the ideal value of the Jaccard index expected from complete genome coverage without any sequencing errors.

| | B.1.617.2 | BA.1 | BA.2 | BA.3 | BA.4 | BA.5 |
|---|---|---|---|---|---|---|
| B.1.617.2 | 1.00 (1.00) | 0.00 (0.00) | 0.00 (0.00) | 0.00 (0.00) | 0.04 (0.02) | 0.04 (0.03) |
| BA.1 | 0.00 (0.00) | 1.00 (1.00) | 0.09 (0.10) | 0.12 (0.21) | 0.06 (0.08) | 0.06 (0.08) |
| BA.2 | 0.00 (0.00) | 0.09 (0.10) | 1.00 (1.00) | 0.44 (0.33) | 0.62 (0.63) | 0.61 (0.59) |
| BA.3 | 0.00 (0.00) | 0.12 (0.21) | 0.44 (0.33) | 1.00 (1.00) | 0.37 (0.30) | 0.35 (0.29) |

| | | | | | | |
|---|---|---|---|---|---|---|
| BA.4 | 0.04 (0.02) | 0.06 (0.08) | 0.62 (0.63) | 0.37 (0.30) | 1.00 (1.00) | 0.84 (0.84) |
| BA.5 | 0.04 (0.03) | 0.06 (0.08) | 0.61 (0.59) | 0.35 (0.29) | 0.84 (0.84) | 1.00 (1.00) |

#### Detected mutations

Excluded from this pdf version due to file size limitations.

U.S. FOOD & DRUG ADMINISTRATION

#### CFSAN/OAO
#### BIOSTATISTICS AND BIOINFORMATICS STAFF

### WASTEWATER SARS-COV2 ANALYSIS REPORT

| | |
|---|---|
| Sample name: | CFSANSMP000118583 |
| Date generated: | 2023-02-01, 20:07:40 EST |
| Timestamp of C-WAP version used: | Tue Jan 31 11:49:22 2023 -0500 |
| Executed by: | Jasmine Amirzadegan |
| Executed on: | 172.20.44.132 (aka n132.raven.cfsan) |

#### Sequencing summary

| | |
|---|---|
| Sequencing chemistry: | Missing with Missing |
| Source site: | Missing (?,?) |
| Sampling date: | Missing |
| Collected by: | Missing |
| Sequenced by: | Missing |
| Total number of reads: | 2338632 |
| Reads aligned: | 2216589 (94%) |
| Average read quality: | 36.8 |
| Average read length: | 149 |
| Reads passing filter: | 2140098 (91%) |
| Average read quality passing filter: | 37.0 |
| Average read length passing filter: | 149 |
| Average coverage passing filter: | 10663X |

A read passes filter if the read length after adaptor trimming ≥ 30 and minimum read quality ≥ 20 within a sliding window of width 4.

#### Overall sequence characteristics

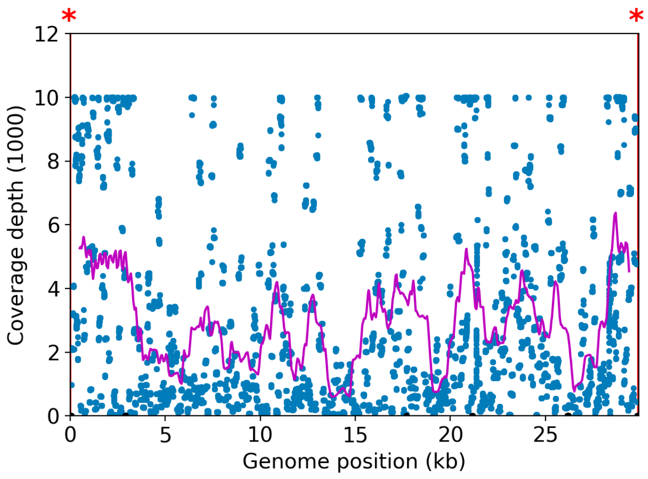


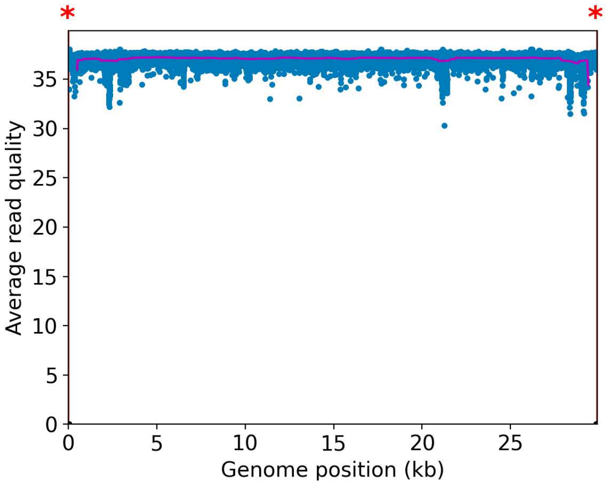


NOTE: The red shaded areas marked with a (*) are not covered by the design of the library preparation kit and hence excluded from analyses. Magenta curves represent moving average with a window width of 1kb.

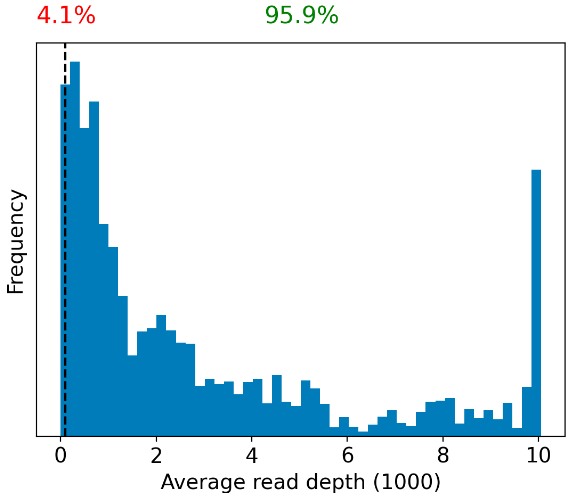


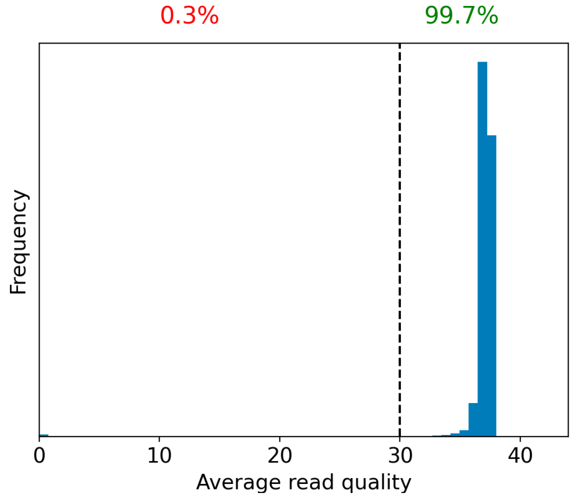


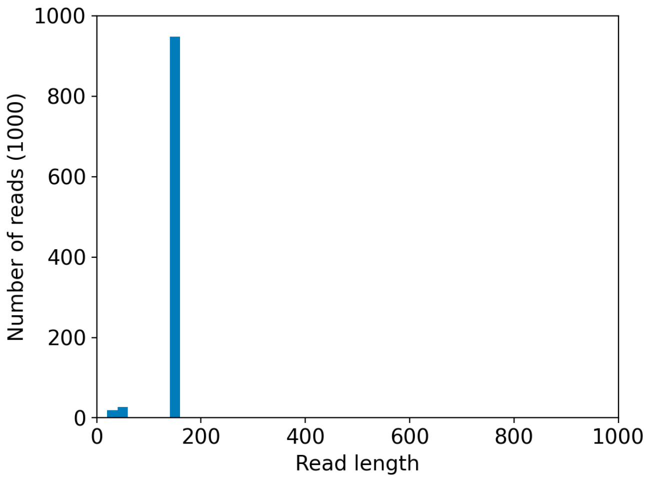


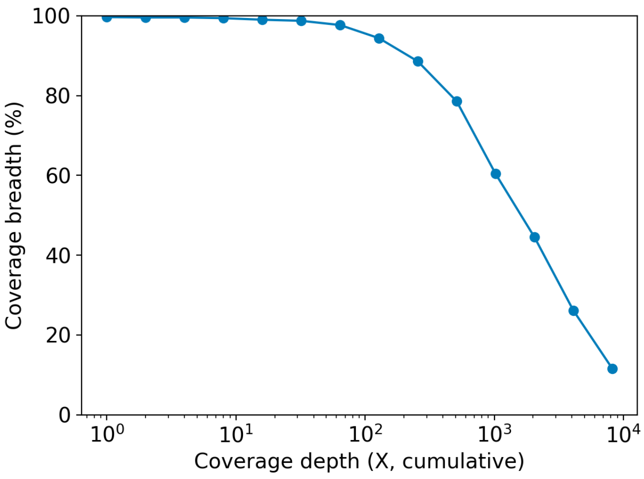

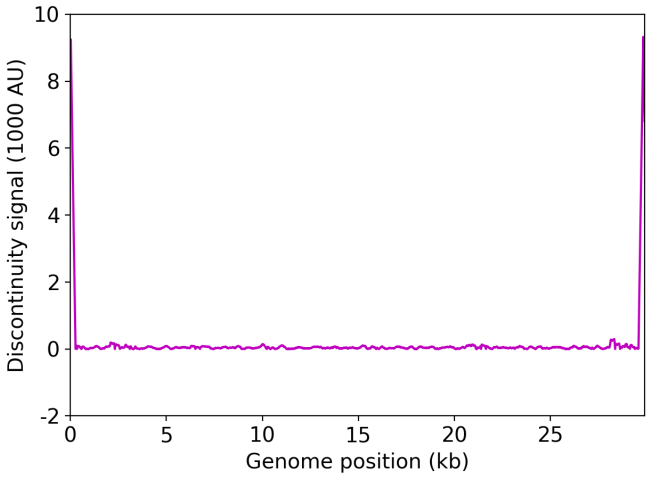


| | Uncovered coordinates (0X) | Poorly covered coordinates (<10X) |
|---|---|---|
| # Inaccessible genomic coordinates by kit design: | 121nt (0%) | 121nt (0%) |
| All genomic coordinates: | 94nt (0%) | 248nt (0%) |
| Common SNPs: | 0nt (0%) | 0nt (0%) |
| Diverse SNPs: | 28nt (12%) | 29nt (12%) |
| Rare SNPs: | 9nt (0%) | 10nt (1%) |

SNPs refer to the polymorphic sites currently in circulation that were detected out of recent GISAID entries. The sites that differ from the SC2 reference sequence are denoted as "common" if [90%, 100%] of the submissions carry this mutation, whereas those that are prevalent in [0%,10%] of the submissions are grouped under the "rare" category. The population is still diverse at the mutation sites that are observed in (10%,90%) of the entries and these coordinates are grouped under the "diverse" category.

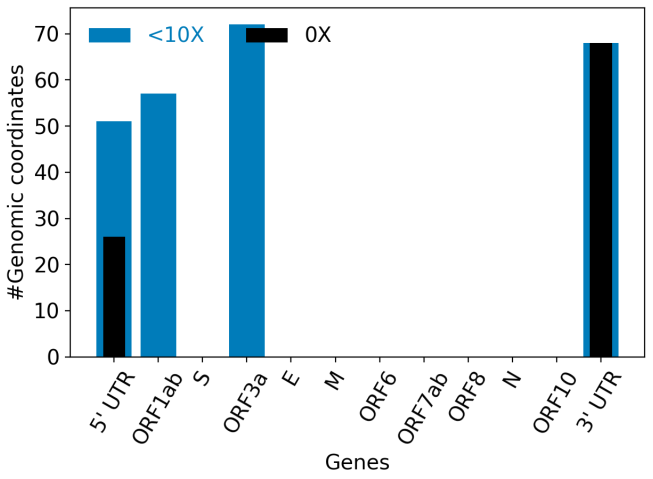


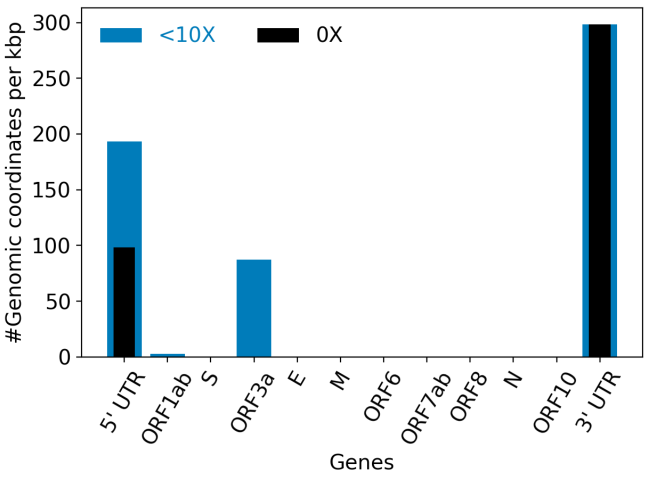


| | |
|---|---|
| Hits to SARS-Cov2 genome (kraken2): | 1117100 reads (95.53%) |
| Hits to human genome (kraken2): | 111 reads (0.01%) |
| Hits to synthetic sequences (kraken2, taxid 28384): | 6 reads (0.00%) |
| Most abundant organisms (kraken2, family level): | Coronaviridae (95.53%)<br>Arcobacteraceae (0.05%)<br>Staphylococcaceae (0.01%) |

| | |
|---|---|
| | Hominidae (0.01%)<br>Moraxellaceae (0.00%) |

### Detected variants (Experimental)

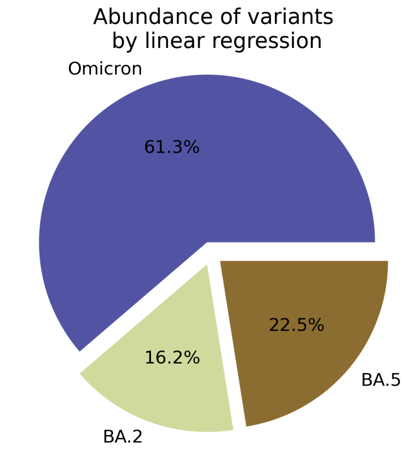


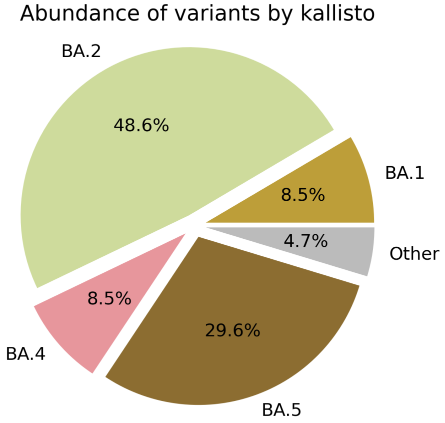


Based on deconvolution, B.1.1.529 is estimated to constitute 61.26% of the viral particles and hence is the most abundant variant in the sample. The $R^2$ for the linear regression was 0.62. Variants that were detected less than 5% were grouped under "Other"

Based on the consensus sequence of the observed reads, the "ensemble-averaged sequence" most closely resembles the BA.2 lineage. If this is a sample consisting of a single source of pathogens or an overwhelming majority of the different sources are infected with the same variant, the sample is dominated by this variant.

Based on mapping individual reads to the variant consensus sequences in the reference database, kallisto predicts that the sample is dominated by BA.2 lineage. Accuracy of this measure is expected to improve if the input data consists of long reads as opposed to convolution.

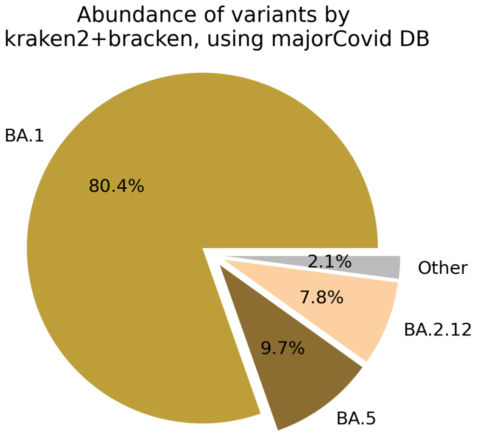


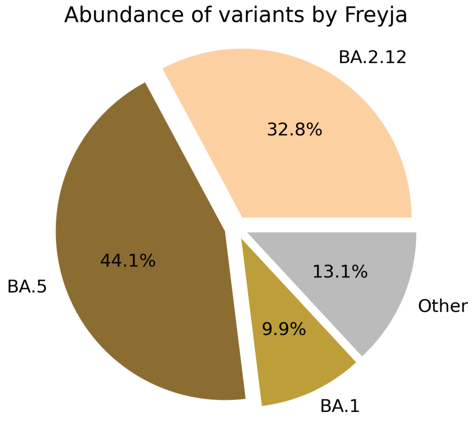

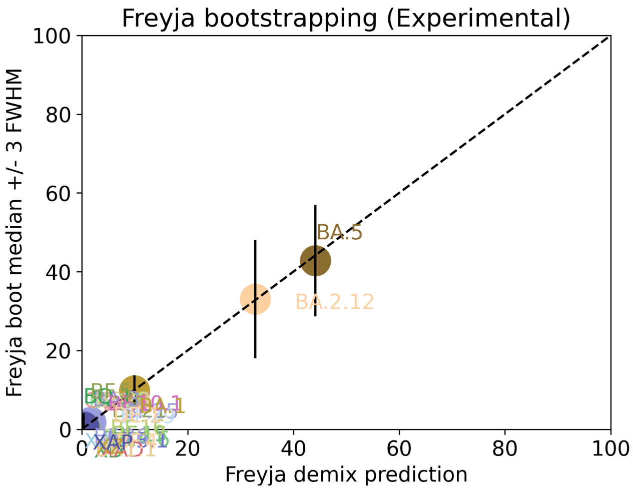


Under the assumption that the presence of a variant requires the detection of all respective mutations of the variant, the characteric mutations which support the presence of the respective variant are indicated in the respective column of the table. Numbers show the number of mutations detected, if any, and the number of mutations expected to be present based on the variant definitions.

| VOC | B.1.617.2 | BA.1 | BA.2 | BA.3 | BA.4 | BA.5 |
|---|---|---|---|---|---|---|
| Characteristic mutations detected | (4 of 13)<br>N:D377Y<br>S:G142D<br>S:L452R<br>S:T478K | (11 of 26)<br>NUC:C25000T<br>NUC:C25584T<br>NUC:T13195C<br>ORF1AB:A2710T<br>S:G446S<br>S:G496S<br>S:L981F<br>S:N856K<br>S:Q493R<br>S:T547K<br>S:T95I | (23 of 31)<br>N:S413R<br>NUC:A20055G<br>NUC:A9424G<br>NUC:C10198T<br>NUC:C12880T<br>NUC:C15714T<br>NUC:C25000T<br>NUC:C25584T<br>NUC:C26858T<br>NUC:C4321T<br>NUC:G10447A<br>ORF1AB:G1307S<br>ORF1AB:L3027F<br>ORF1AB:L3201F<br>ORF1AB:S135R<br>ORF1AB:T3090I<br>ORF1AB:T842I<br>S:D405N<br>S:Q493R<br>S:R408S<br>S:S371F<br>S:T19I<br>S:T376A | (12 of 21)<br>N:S413R<br>NUC:C12880T<br>NUC:C15714T<br>NUC:C26858T<br>NUC:G10447A<br>ORF1AB:G1307S<br>ORF1AB:S135R<br>ORF1AB:T3090I<br>S:D405N<br>S:G446S<br>S:Q493R<br>S:S371F | (24 of 31)<br>N:P151S<br>N:S413R<br>NUC:A20055G<br>NUC:C10198T<br>NUC:C12880T<br>NUC:C15714T<br>NUC:C25000T<br>NUC:C25584T<br>NUC:C26858T<br>NUC:C4321T<br>NUC:G10447A<br>NUC:G12160A<br>NUC:G27788T<br>ORF1AB:G1307S<br>ORF1AB:S135R<br>ORF1AB:T3090I<br>ORF1AB:T842I<br>S:D405N<br>S:F486V<br>S:L452R<br>S:S371F<br>S:T19I<br>S:T376A<br>S:V213G | (22 of 28)<br>M:D3N<br>N:S413R<br>NUC:A20055G<br>NUC:C10198T<br>NUC:C12880T<br>NUC:C15714T<br>NUC:C25000T<br>NUC:C25584T<br>NUC:C4321T<br>NUC:G10447A<br>NUC:G12160A<br>ORF1AB:G1307S<br>ORF1AB:S135R<br>ORF1AB:T3090I<br>ORF1AB:T842I<br>S:D405N<br>S:F486V<br>S:L452R<br>S:S371F<br>S:T19I<br>S:T376A<br>S:V213G |

Jaccard Index is a measure of similarity between two sets A and B, reaching the maximum value of 1 if A=B and minimum value of 0 if A ∩ B = {}. In the c(d) representation below, c represents the Jaccard index of the set of mutations that were experimentally detected for this sample as listed above, whereas d refers to the ideal value of the Jaccard index expected from complete genome coverage without any sequencing errors.

| | B.1.617.2 | BA.1 | BA.2 | BA.3 | BA.4 | BA.5 |
|---|---|---|---|---|---|---|
| B.1.617.2 | 1.00 (1.00) | 0.00 (0.00) | 0.00 (0.00) | 0.00 (0.00) | 0.04 (0.02) | 0.04 (0.03) |
| BA.1 | 0.00 (0.00) | 1.00 (1.00) | 0.10 (0.10) | 0.10 (0.21) | 0.06 (0.08) | 0.06 (0.08) |
| BA.2 | 0.00 (0.00) | 0.10 (0.10) | 1.00 (1.00) | 0.46 (0.33) | 0.62 (0.63) | 0.61 (0.59) |
| BA.3 | 0.00 (0.00) | 0.10 (0.21) | 0.46 (0.33) | 1.00 (1.00) | 0.38 (0.30) | 0.36 (0.29) |

| | | | | | | |
|---|---|---|---|---|---|---|
| BA.4 | 0.04 (0.02) | 0.06 (0.08) | 0.62 (0.63) | 0.38 (0.30) | 1.00 (1.00) | 0.84 (0.84) |
| BA.5 | 0.04 (0.03) | 0.06 (0.08) | 0.61 (0.59) | 0.36 (0.29) | 0.84 (0.84) | 1.00 (1.00) |

#### Detected mutations

Excluded from this pdf version due to file size limitations.

CFSAN/OAO
BIOSTATISTICS AND BIOINFORMATICS STAFF

### WASTEWATER SARS-COV2 ANALYSIS REPORT

| | |
|---|---|
| Sample name: | CFSANSMP000118584 |
| Date generated: | 2023-02-01, 20:10:05 EST |
| Timestamp of C-WAP version used: | Tue Jan 31 11:49:22 2023 -0500 |
| Executed by: | Jasmine Amirzadegan |
| Executed on: | 172.20.44.108 (aka n108.raven.cfsan) |

#### Sequencing summary

| | |
|---|---|
| Sequencing chemistry: | Missing with Missing |
| Source site: | Missing (?,?) |
| Sampling date: | Missing |
| Collected by: | Missing |
| Sequenced by: | Missing |
| Total number of reads: | 2526212 |
| Reads aligned: | 2451480 (97%) |
| Average read quality: | 36.6 |
| Average read length: | 149 |
| Reads passing filter: | 2345410 (92%) |
| Average read quality passing filter: | 36.9 |
| Average read length passing filter: | 149 |
| Average coverage passing filter: | 11686X |

A read passes filter if the read length after adaptor trimming ≥ 30 and minimum read quality ≥ 20 within a sliding window of width 4.

#### Overall sequence characteristics

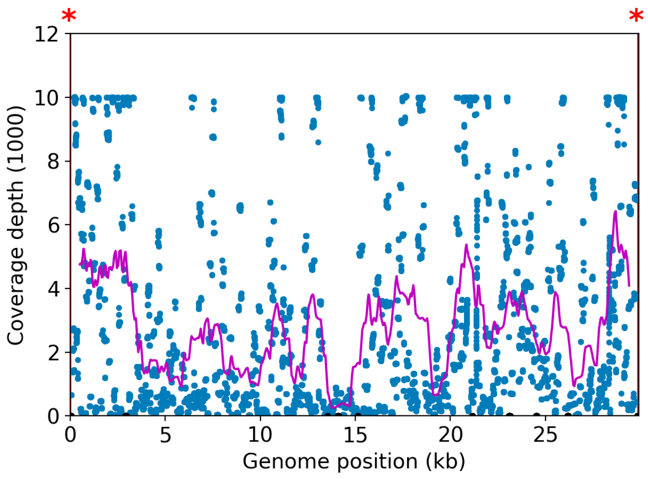


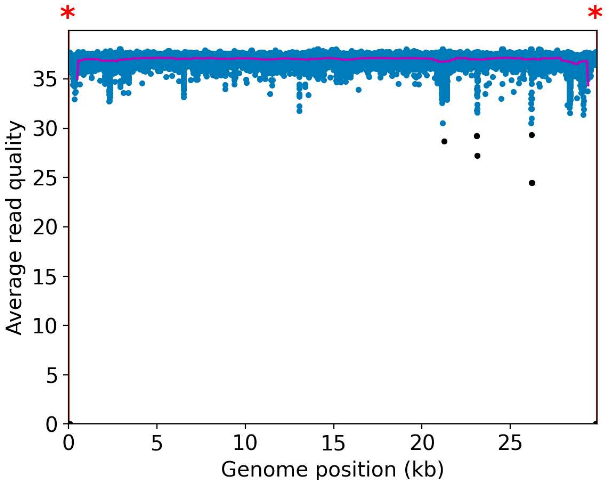


NOTE: The red shaded areas marked with a (*) are not covered by the design of the library preparation kit and hence excluded from analyses. Magenta curves represent moving average with a window width of 1kb.

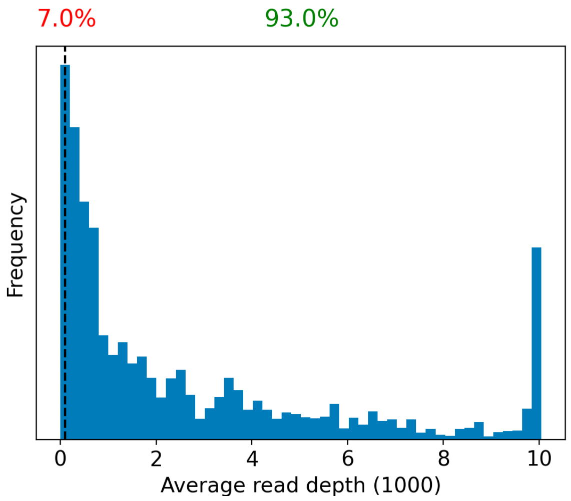


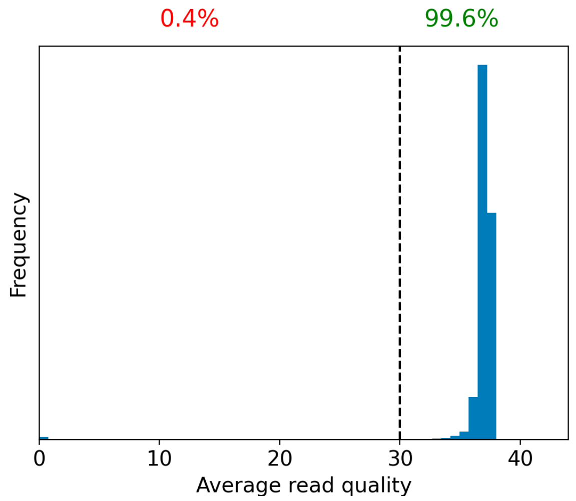


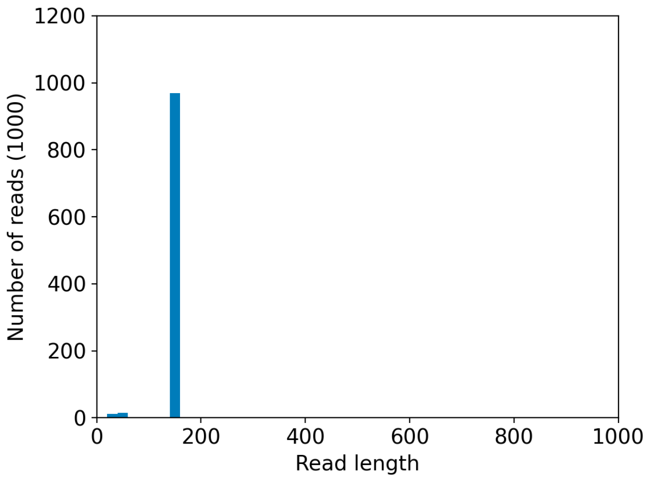


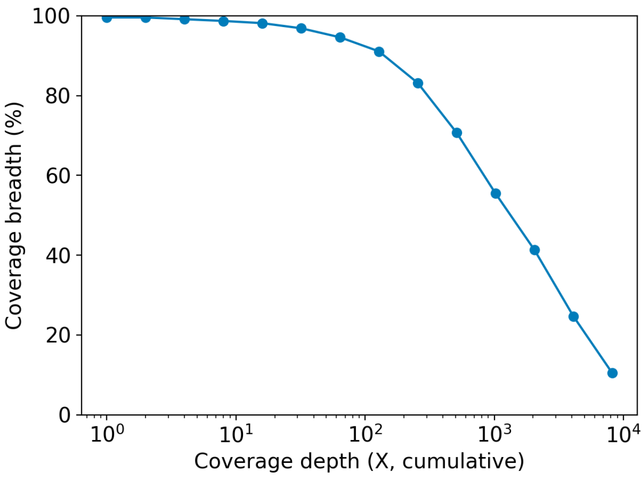

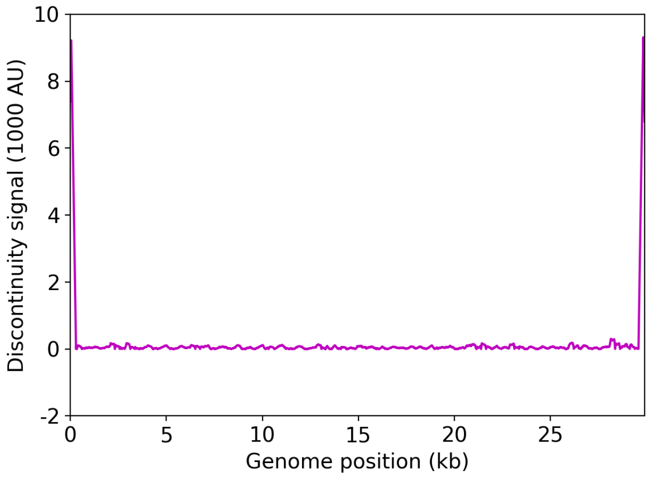


| | Uncovered coordinates (0X) | Poorly covered coordinates (<10X) |
|---|---|---|
| # Inaccessible genomic coordinates by kit design: | 121nt (0%) | 121nt (0%) |
| All genomic coordinates: | 119nt (0%) | 410nt (1%) |
| Common SNPs: | 0nt (0%) | 0nt (0%) |
| Diverse SNPs: | 29nt (12%) | 29nt (12%) |
| Rare SNPs: | 10nt (1%) | 46nt (4%) |

SNPs refer to the polymorphic sites currently in circulation that were detected out of recent GISAID entries. The sites that differ from the SC2 reference sequence are denoted as "common" if [90%, 100%] of the submissions carry this mutation, whereas those that are prevalent in [0%,10%] of the submissions are grouped under the "rare" category. The population is still diverse at the mutation sites that are observed in (10%,90%) of the entries and these coordinates are grouped under the "diverse" category.

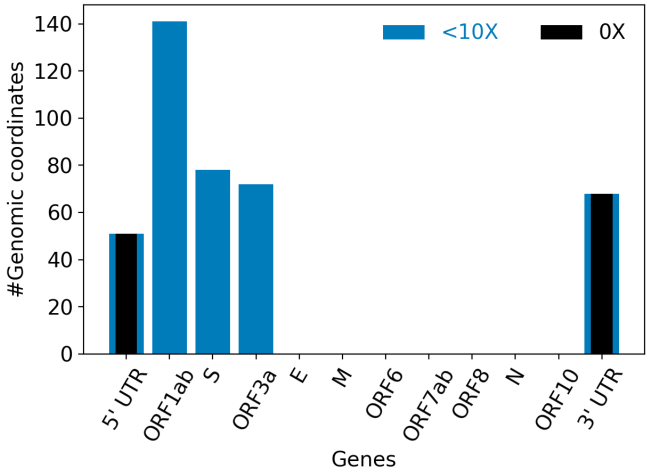


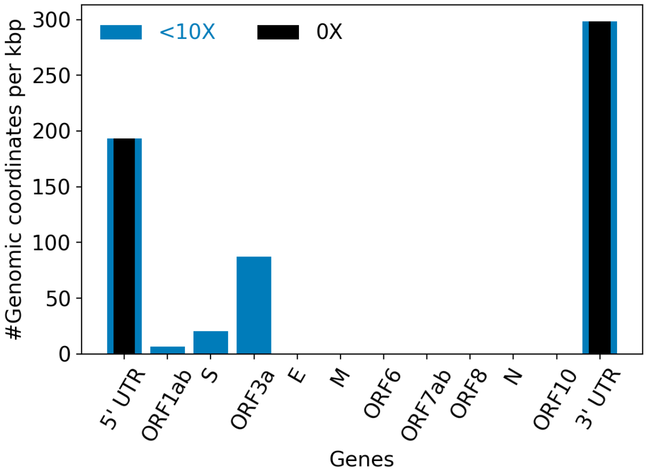


| | |
|---|---|
| Hits to SARS-Cov2 genome (kraken2): | 1232483 reads (97.58%) |
| Hits to human genome (kraken2): | 90 reads (0.01%) |
| Hits to synthetic sequences (kraken2, taxid 28384): | 19 reads (0.00%) |
| Most abundant organisms (kraken2, family level): | Coronaviridae (97.58%)<br>Mycobacteriaceae (0.01%)<br>Hominidae (0.01%) |

| | |
|---|---|
| | Sphaerotilaceae (0.00%)<br>Staphylococcaceae (0.00%) |

### Detected variants (Experimental)

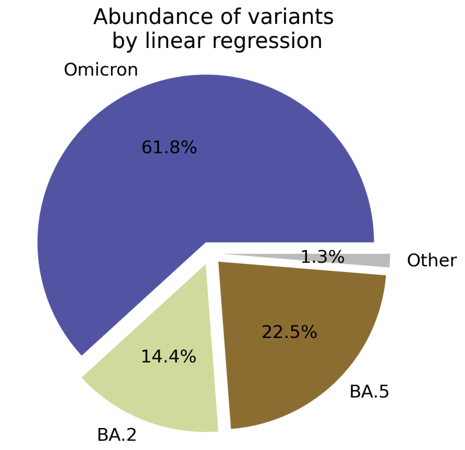


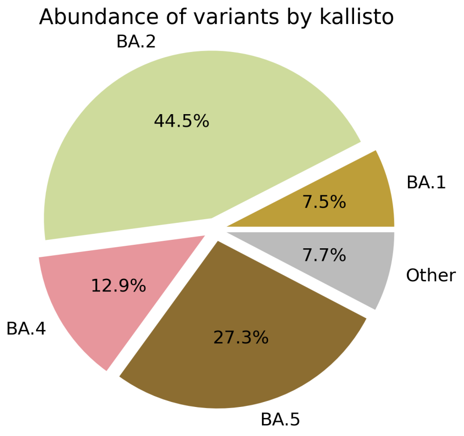


Based on deconvolution, B.1.1.529 is estimated to constitute 61.82% of the viral particles and hence is the most abundant variant in the sample. The $R^2$ for the linear regression was 0.62. Variants that were detected less than 5% were grouped under "Other"

Based on the consensus sequence of the observed reads, the "ensemble-averaged sequence" most closely resembles the BA.2 lineage. If this is a sample consisting of a single source of pathogens or an overwhelming majority of the different sources are infected with the same variant, the sample is dominated by this variant.

Based on mapping individual reads to the variant consensus sequences in the reference database, kallisto predicts that the sample is dominated by BA.2 lineage. Accuracy of this measure is expected to improve if the input data consists of long reads as opposed to convolution.

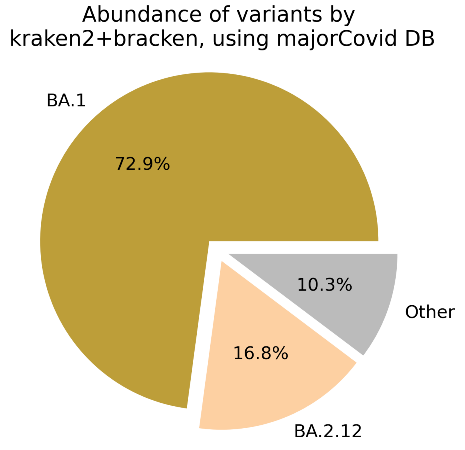


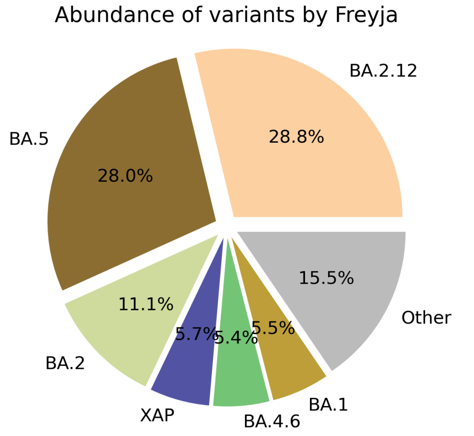

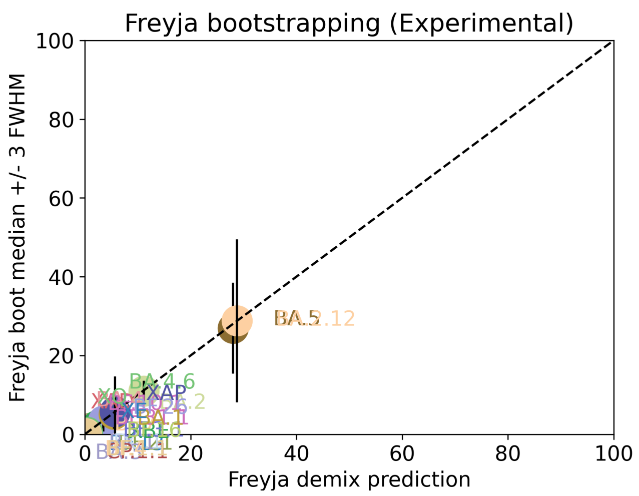


Under the assumption that the presence of a variant requires the detection of all respective mutations of the variant, the characteric mutations which support the presence of the respective variant are indicated in the respective column of the table. Numbers show the number of mutations detected, if any, and the number of mutations expected to be present based on the variant definitions.

| VOC | B.1.617.2 | BA.1 | BA.2 | BA.3 | BA.4 | BA.5 |
|---|---|---|---|---|---|---|
| Characteristic mutations detected | (4 of 13)<br>N:D377Y<br>S:G142D<br>S:L452R<br>S:T478K | (11 of 26)<br>NUC:C25000T<br>NUC:C25584T<br>NUC:T13195C<br>NUC:T5386G<br>S:G446S<br>S:G496S<br>S:L981F<br>S:N856K<br>S:Q493R<br>S:T547K<br>S:T95I | (22 of 31)<br>N:S413R<br>NUC:A20055G<br>NUC:C10198T<br>NUC:C12880T<br>NUC:C15714T<br>NUC:C25000T<br>NUC:C25584T<br>NUC:C26858T<br>NUC:C4321T<br>NUC:G10447A<br>ORF1AB:G1307S<br>ORF1AB:L3027F<br>ORF1AB:L3201F<br>ORF1AB:S135R<br>ORF1AB:T3090I<br>ORF1AB:T842I<br>S:D405N<br>S:Q493R<br>S:R408S<br>S:S371F<br>S:T19I<br>S:T376A | (12 of 21)<br>N:S413R<br>NUC:C12880T<br>NUC:C15714T<br>NUC:C26858T<br>NUC:G10447A<br>ORF1AB:G1307S<br>ORF1AB:S135R<br>ORF1AB:T3090I<br>S:D405N<br>S:G446S<br>S:Q493R<br>S:S371F | (24 of 31)<br>N:P151S<br>N:S413R<br>NUC:A20055G<br>NUC:C10198T<br>NUC:C12880T<br>NUC:C15714T<br>NUC:C25000T<br>NUC:C25584T<br>NUC:C26858T<br>NUC:C4321T<br>NUC:G10447A<br>NUC:G12160A<br>NUC:G27788T<br>ORF1AB:G1307S<br>ORF1AB:S135R<br>ORF1AB:T3090I<br>ORF1AB:T842I<br>S:D405N<br>S:F486V<br>S:L452R<br>S:S371F<br>S:T19I<br>S:T376A<br>S:V213G | (22 of 28)<br>M:D3N<br>N:S413R<br>NUC:A20055G<br>NUC:C10198T<br>NUC:C12880T<br>NUC:C15714T<br>NUC:C25000T<br>NUC:C25584T<br>NUC:C4321T<br>NUC:G10447A<br>NUC:G12160A<br>ORF1AB:G1307S<br>ORF1AB:S135R<br>ORF1AB:T3090I<br>ORF1AB:T842I<br>S:D405N<br>S:F486V<br>S:L452R<br>S:S371F<br>S:T19I<br>S:T376A<br>S:V213G |

Jaccard Index is a measure of similarity between two sets A and B, reaching the maximum value of 1 if A=B and minimum value of 0 if A ∩ B = {}. In the c(d) representation below, c represents the Jaccard index of the set of mutations that were experimentally detected for this sample as listed above, whereas d refers to the ideal value of the Jaccard index expected from complete genome coverage without any sequencing errors.

| | B.1.617.2 | BA.1 | BA.2 | BA.3 | BA.4 | BA.5 |
|---|---|---|---|---|---|---|
| B.1.617.2 | 1.00 (1.00) | 0.00 (0.00) | 0.00 (0.00) | 0.00 (0.00) | 0.04 (0.02) | 0.04 (0.03) |
| BA.1 | 0.00 (0.00) | 1.00 (1.00) | 0.10 (0.10) | 0.10 (0.21) | 0.06 (0.08) | 0.06 (0.08) |
| BA.2 | 0.00 (0.00) | 0.10 (0.10) | 1.00 (1.00) | 0.48 (0.33) | 0.64 (0.63) | 0.63 (0.59) |
| BA.3 | 0.00 (0.00) | 0.10 (0.21) | 0.48 (0.33) | 1.00 (1.00) | 0.38 (0.30) | 0.36 (0.29) |

| | | | | | | |
|---|---|---|---|---|---|---|
| BA.4 | 0.04 (0.02) | 0.06 (0.08) | 0.64 (0.63) | 0.38 (0.30) | 1.00 (1.00) | 0.84 (0.84) |
| BA.5 | 0.04 (0.03) | 0.06 (0.08) | 0.63 (0.59) | 0.36 (0.29) | 0.84 (0.84) | 1.00 (1.00) |

#### Detected mutations

Excluded from this pdf version due to file size limitations.

CFSAN/OAO
BIOSTATISTICS AND BIOINFORMATICS STAFF

### WASTEWATER SARS-COV2 ANALYSIS REPORT

| | |
|---|---|
| Sample name: | CFSANSMP000118585 |
| Date generated: | 2023-02-01, 20:10:44 EST |
| Timestamp of C-WAP version used: | Tue Jan 31 11:49:22 2023 -0500 |
| Executed by: | Jasmine Amirzadegan |
| Executed on: | 172.20.44.108 (aka n108.raven.cfsan) |

#### Sequencing summary

| | |
|---|---|
| Sequencing chemistry: | Missing with Missing |
| Source site: | Missing (?,?) |
| Sampling date: | Missing |
| Collected by: | Missing |
| Sequenced by: | Missing |
| Total number of reads: | 2829500 |
| Reads aligned: | 2627859 (92%) |
| Average read quality: | 36.7 |
| Average read length: | 149 |
| Reads passing filter: | 2527472 (89%) |
| Average read quality passing filter: | 36.9 |
| Average read length passing filter: | 149 |
| Average coverage passing filter: | 12593X |

A read passes filter if the read length after adaptor trimming ≥ 30 and minimum read quality ≥ 20 within a sliding window of width 4.

#### Overall sequence characteristics

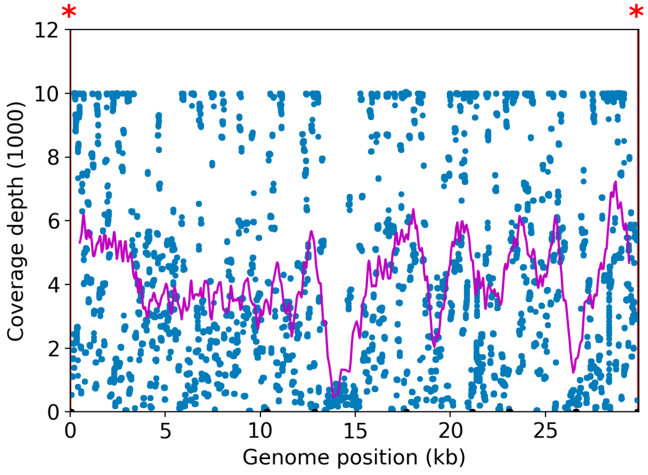


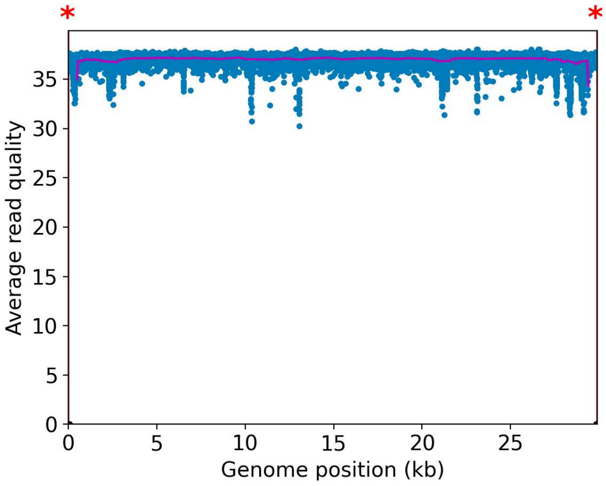


NOTE: The red shaded areas marked with a (*) are not covered by the design of the library preparation kit and hence excluded from analyses. Magenta curves represent moving average with a window width of 1kb.

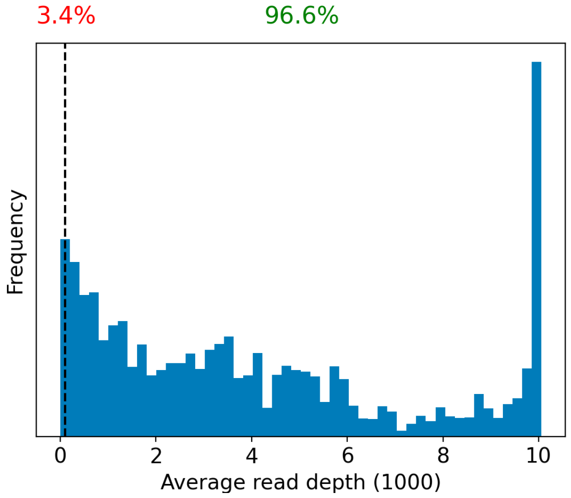


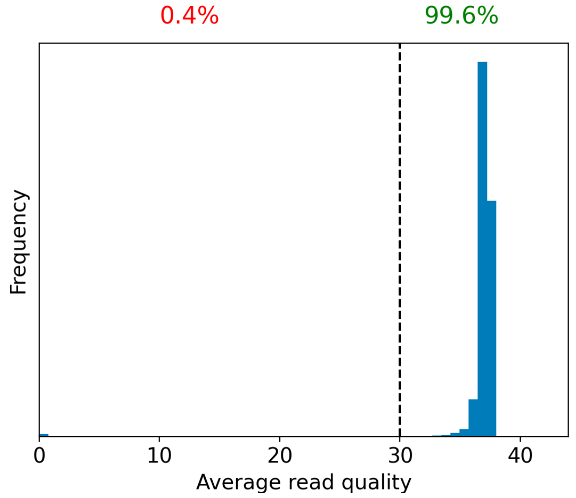


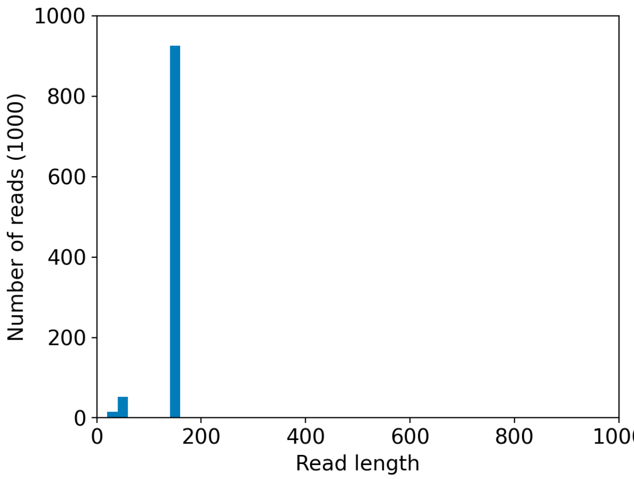


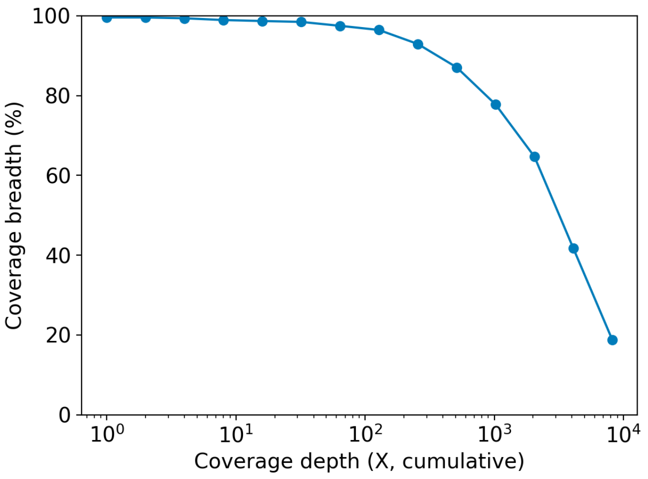

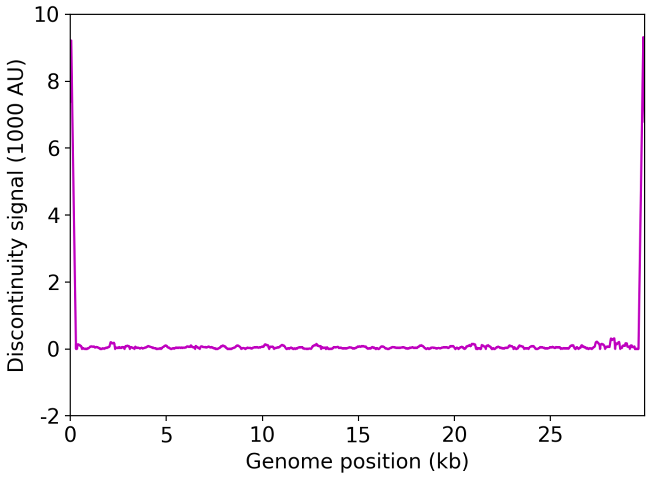


| | Uncovered coordinates (0X) | Poorly covered coordinates (<10X) |
|---|---|---|
| # Inaccessible genomic coordinates by kit design: | 121nt (0%) | 121nt (0%) |
| All genomic coordinates: | 119nt (0%) | 324nt (1%) |
| Common SNPs: | 0nt (0%) | 1nt (2%) |
| Diverse SNPs: | 29nt (12%) | 29nt (12%) |
| Rare SNPs: | 10nt (1%) | 47nt (5%) |

SNPs refer to the polymorphic sites currently in circulation that were detected out of recent GISAID entries. The sites that differ from the SC2 reference sequence are denoted as "common" if [90%, 100%] of the submissions carry this mutation, whereas those that are prevalent in [0%,10%] of the submissions are grouped under the "rare" category. The population is still diverse at the mutation sites that are observed in (10%,90%) of the entries and these coordinates are grouped under the "diverse" category.

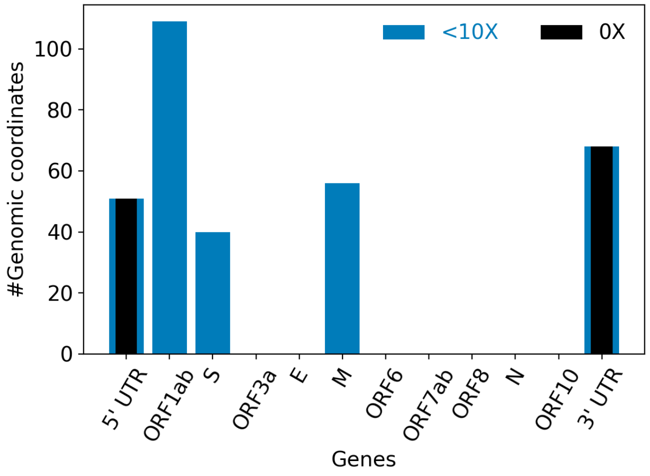


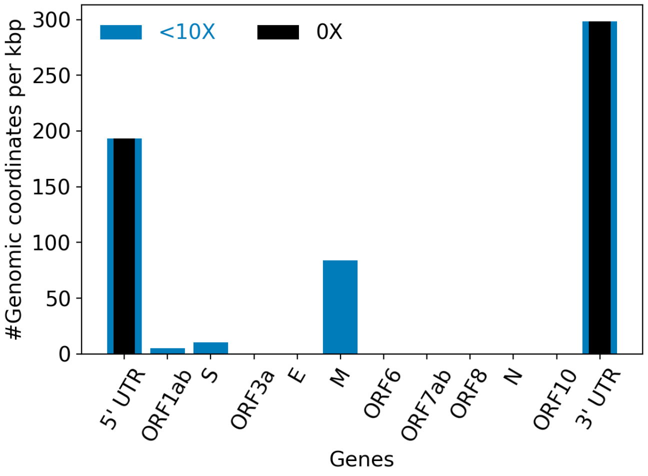


| | |
|---|---|
| Hits to SARS-Cov2 genome (kraken2): | 1329189 reads (93.95%) |
| Hits to human genome (kraken2): | 438 reads (0.03%) |
| Hits to synthetic sequences (kraken2, taxid 28384): | 46 reads (0.00%) |
| Most abundant organisms (kraken2, family level): | Coronaviridae (93.95%)<br>Staphylococcaceae (0.09%)<br>Hominidae (0.03%) |

| | |
|---|---|
| | Arcobacteraceae (0.02%)<br>Lachnospiraceae (0.00%) |

#### Detected variants (Experimental)

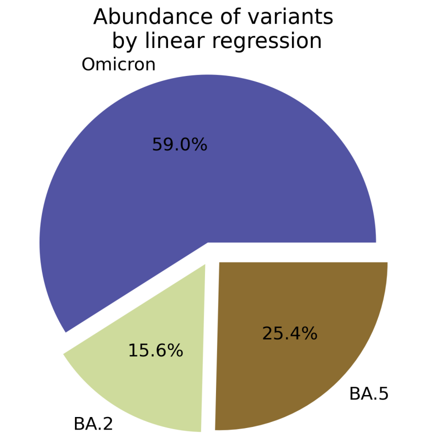


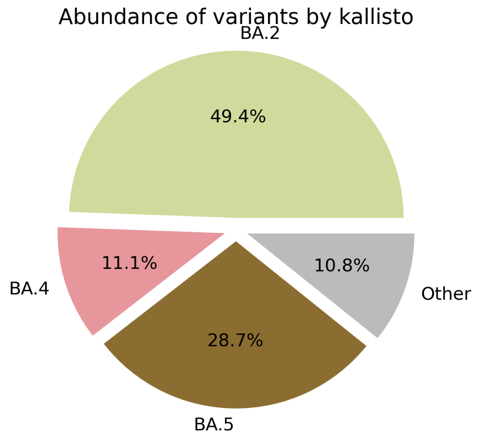


Based on deconvolution, B.1.1.529 is estimated to constitute 58.98% of the viral particles and hence is the most abundant variant in the sample. The $R^2$ for the linear regression was 0.60. Variants that were detected less than 5% were grouped under "Other"

Based on the consensus sequence of the observed reads, the "ensemble-averaged sequence" most closely resembles the BA.2 lineage. If this is a sample consisting of a single source of pathogens or an overwhelming majority of the different sources are infected with the same variant, the sample is dominated by this variant.

Based on mapping individual reads to the variant consensus sequences in the reference database, kallisto predicts that the sample is dominated by BA.2 lineage. Accuracy of this measure is expected to improve if the input data consists of long reads as opposed to convolution.

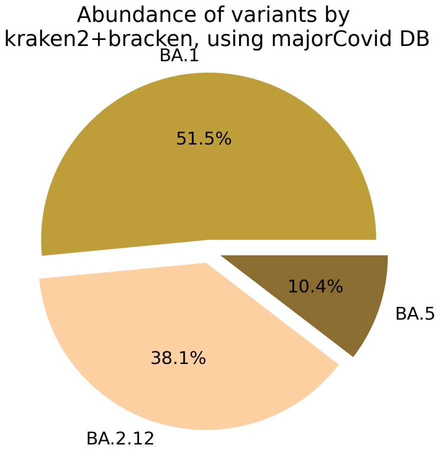


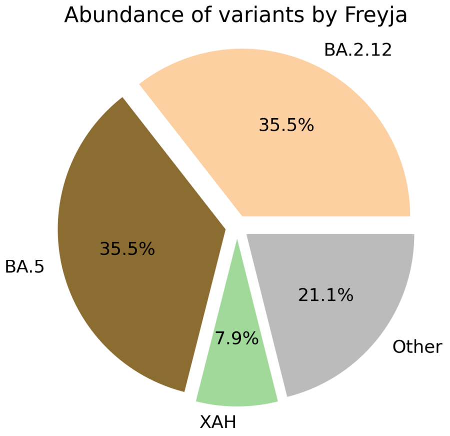

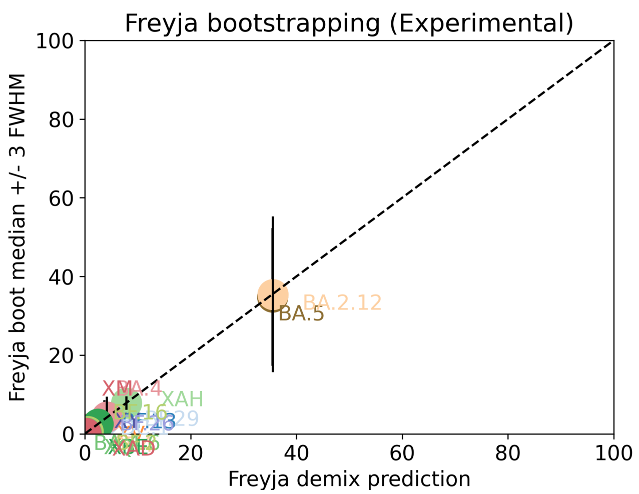


Under the assumption that the presence of a variant requires the detection of all respective mutations of the variant, the characteric mutations which support the presence of the respective variant are indicated in the respective column of the table. Numbers show the number of mutations detected, if any, and the number of mutations expected to be present based on the variant definitions.

| VOC | B.1.617.2 | BA.1 | BA.2 | BA.3 | BA.4 | BA.5 |
|---|---|---|---|---|---|---|
| Characteristic mutations detected | (4 of 13)<br>N:D377Y<br>S:G142D<br>S:L452R<br>S:T478K | (9 of 26)<br>NUC:C25000T<br>NUC:C25584T<br>NUC:T13195C<br>ORF1AB:A2710T<br>ORF1AB:I3758V<br>ORF1AB:K856R<br>S:L981F<br>S:N856K<br>S:Q493R | (22 of 31)<br>N:S413R<br>NUC:A20055G<br>NUC:C10198T<br>NUC:C12880T<br>NUC:C15714T<br>NUC:C25000T<br>NUC:C25584T<br>NUC:C26858T<br>NUC:C4321T<br>NUC:G10447A<br>ORF1AB:G1307S<br>ORF1AB:L3027F<br>ORF1AB:L3201F<br>ORF1AB:S135R<br>ORF1AB:T3090I<br>ORF1AB:T842I<br>S:D405N<br>S:Q493R<br>S:R408S<br>S:S371F<br>S:T19I<br>S:T376A | (11 of 21)<br>N:S413R<br>NUC:C12880T<br>NUC:C15714T<br>NUC:C26858T<br>NUC:G10447A<br>ORF1AB:G1307S<br>ORF1AB:S135R<br>ORF1AB:T3090I<br>S:D405N<br>S:Q493R<br>S:S371F | (24 of 31)<br>N:P151S<br>N:S413R<br>NUC:A20055G<br>NUC:C10198T<br>NUC:C12880T<br>NUC:C15714T<br>NUC:C25000T<br>NUC:C25584T<br>NUC:C26858T<br>NUC:C4321T<br>NUC:G10447A<br>NUC:G12160A<br>NUC:G27788T<br>ORF1AB:G1307S<br>ORF1AB:S135R<br>ORF1AB:T3090I<br>ORF1AB:T842I<br>S:D405N<br>S:F486V<br>S:L452R<br>S:S371F<br>S:T19I<br>S:T376A<br>S:V213G | (22 of 28)<br>M:D3N<br>N:S413R<br>NUC:A20055G<br>NUC:C10198T<br>NUC:C12880T<br>NUC:C15714T<br>NUC:C25000T<br>NUC:C25584T<br>NUC:C4321T<br>NUC:G10447A<br>NUC:G12160A<br>ORF1AB:G1307S<br>ORF1AB:S135R<br>ORF1AB:T3090I<br>ORF1AB:T842I<br>S:D405N<br>S:F486V<br>S:L452R<br>S:S371F<br>S:T19I<br>S:T376A<br>S:V213G |

Jaccard Index is a measure of similarity between two sets A and B, reaching the maximum value of 1 if A=B and minimum value of 0 if A ∩ B = {}. In the c(d) representation below, c represents the Jaccard index of the set of mutations that were experimentally detected for this sample as listed above, whereas d refers to the ideal value of the Jaccard index expected from complete genome coverage without any sequencing errors.

| | B.1.617.2 | BA.1 | BA.2 | BA.3 | BA.4 | BA.5 |
|---|---|---|---|---|---|---|
| B.1.617.2 | 1.00 (1.00) | 0.00 (0.00) | 0.00 (0.00) | 0.00 (0.00) | 0.04 (0.02) | 0.04 (0.03) |
| BA.1 | 0.00 (0.00) | 1.00 (1.00) | 0.11 (0.10) | 0.05 (0.21) | 0.06 (0.08) | 0.07 (0.08) |
| BA.2 | 0.00 (0.00) | 0.11 (0.10) | 1.00 (1.00) | 0.50 (0.33) | 0.64 (0.63) | 0.63 (0.59) |
| BA.3 | 0.00 (0.00) | 0.05 (0.21) | 0.50 (0.33) | 1.00 (1.00) | 0.40 (0.30) | 0.38 (0.29) |

| | | | | | | |
|---|---|---|---|---|---|---|
| BA.4 | 0.04 (0.02) | 0.06 (0.08) | 0.64 (0.63) | 0.40 (0.30) | 1.00 (1.00) | 0.84 (0.84) |
| BA.5 | 0.04 (0.03) | 0.07 (0.08) | 0.63 (0.59) | 0.38 (0.29) | 0.84 (0.84) | 1.00 (1.00) |

#### Detected mutations

Excluded from this pdf version due to file size limitations.

CFSAN/OAO
BIOSTATISTICS AND BIOINFORMATICS STAFF

### WASTEWATER SARS-COV2 ANALYSIS REPORT

| | |
|---|---|
| Sample name: | CFSANSMP000118586 |
| Date generated: | 2023-02-01, 20:05:19 EST |
| Timestamp of C-WAP version used: | Tue Jan 31 11:49:22 2023 -0500 |
| Executed by: | Jasmine Amirzadegan |
| Executed on: | 172.20.44.127 (aka n127.raven.cfsan) |

#### Sequencing summary

| | |
|---|---|
| Sequencing chemistry: | Missing with Missing |
| Source site: | Missing (?,?) |
| Sampling date: | Missing |
| Collected by: | Missing |
| Sequenced by: | Missing |
| Total number of reads: | 2750930 |
| Reads aligned: | 2533050 (92%) |
| Average read quality: | 36.7 |
| Average read length: | 149 |
| Reads passing filter: | 2432859 (88%) |
| Average read quality passing filter: | 37.0 |
| Average read length passing filter: | 149 |
| Average coverage passing filter: | 12122X |

A read passes filter if the read length after adaptor trimming ≥ 30 and minimum read quality ≥ 20 within a sliding window of width 4.

#### Overall sequence characteristics

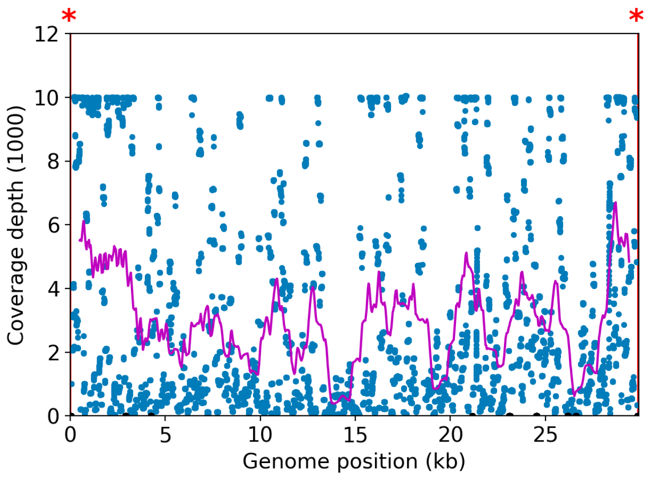


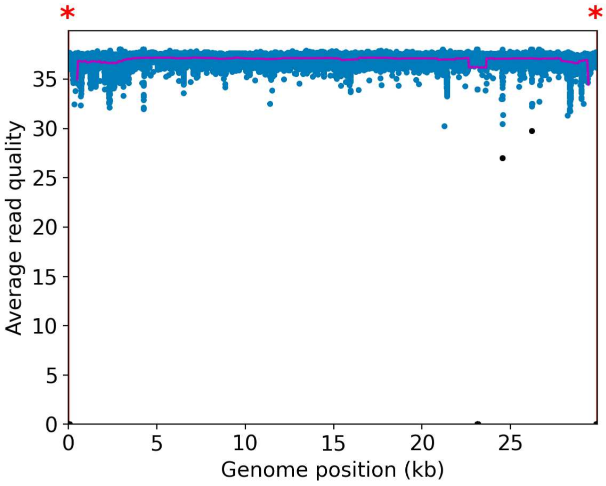


NOTE: The red shaded areas marked with a (*) are not covered by the design of the library preparation kit and hence excluded from analyses. Magenta curves represent moving average with a window width of 1kb.

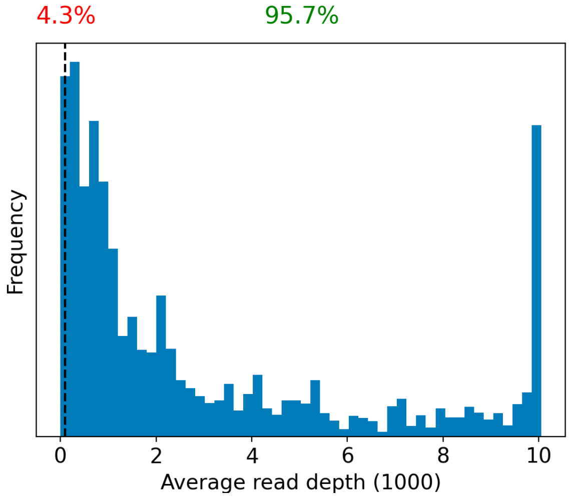


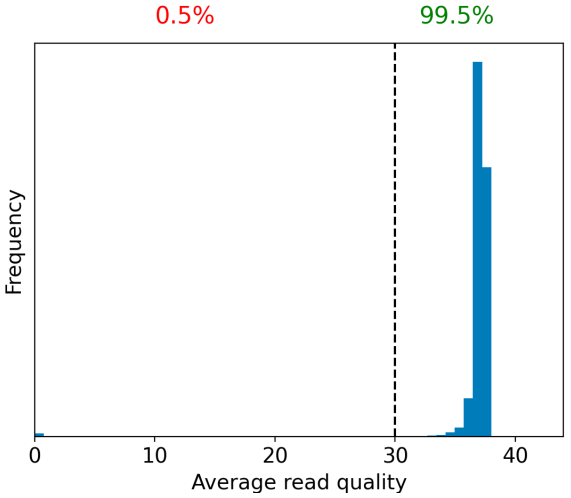


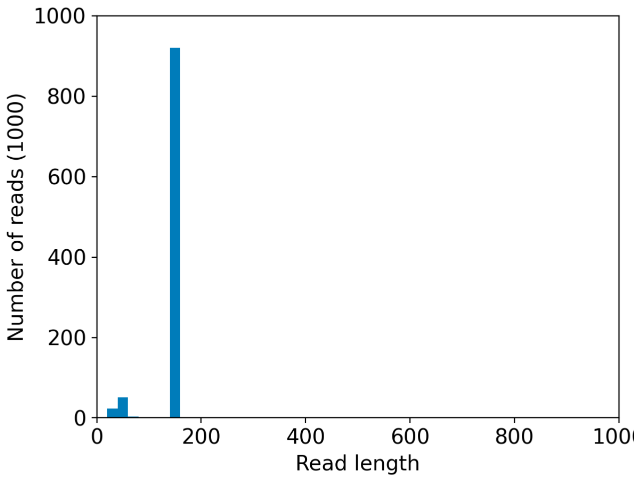


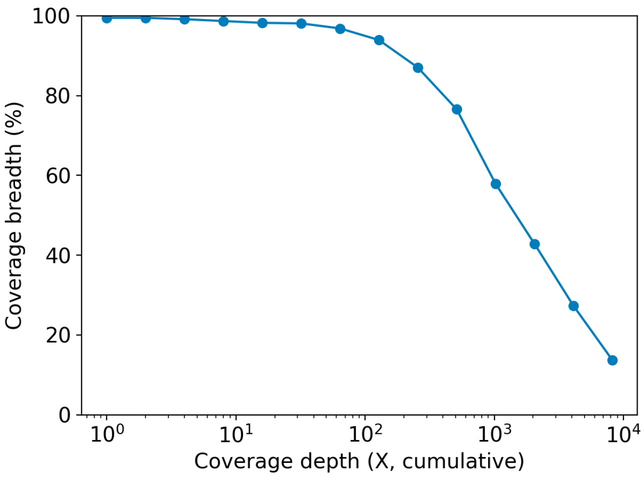

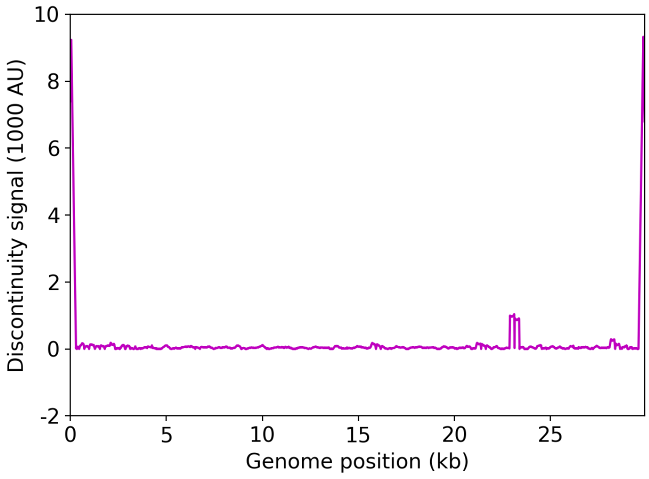


| | Uncovered coordinates (0X) | Poorly covered coordinates (<10X) |
|---|---|---|
| # Inaccessible genomic coordinates by kit design: | 121nt (0%) | 121nt (0%) |
| All genomic coordinates: | 145nt (0%) | 471nt (1%) |
| Common SNPs: | 0nt (0%) | 0nt (0%) |
| Diverse SNPs: | 29nt (12%) | 29nt (12%) |
| Rare SNPs: | 10nt (1%) | 46nt (4%) |

SNPs refer to the polymorphic sites currently in circulation that were detected out of recent GISAID entries. The sites that differ from the SC2 reference sequence are denoted as "common" if [90%, 100%] of the submissions carry this mutation, whereas those that are prevalent in [0%,10%] of the submissions are grouped under the "rare" category. The population is still diverse at the mutation sites that are observed in (10%,90%) of the entries and these coordinates are grouped under the "diverse" category.

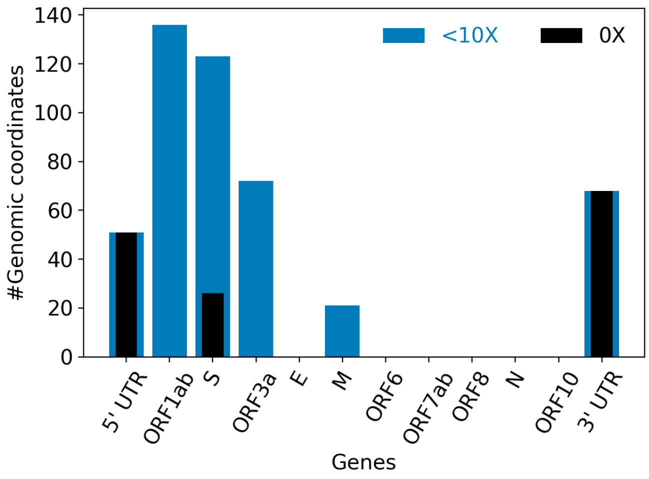


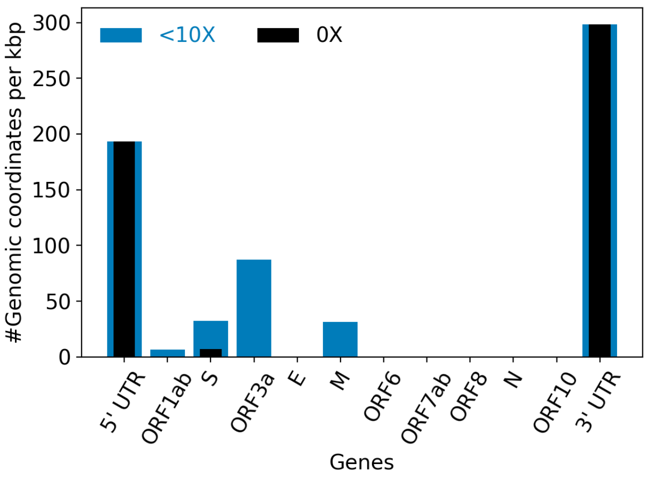


| | |
|---|---|
| Hits to SARS-Cov2 genome (kraken2): | 1285745 reads (93.48%) |
| Hits to human genome (kraken2): | 126 reads (0.01%) |
| Hits to synthetic sequences (kraken2, taxid 28384): | 24 reads (0.00%) |
| Most abundant organisms (kraken2, family level): | Coronaviridae (93.48%)<br>Enterobacteriaceae (0.08%)<br>Mycobacteriaceae (0.01%) |

| | |
|---|---|
| | Hominidae (0.01%)<br>Comamonadaceae (0.00%) |

### Detected variants (Experimental)

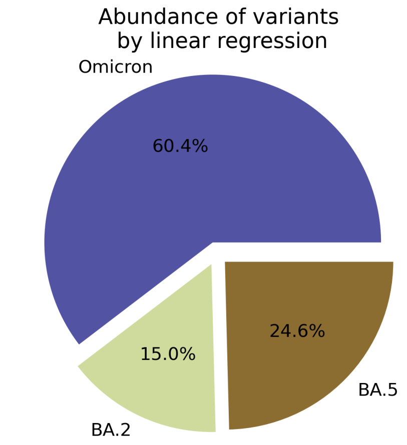


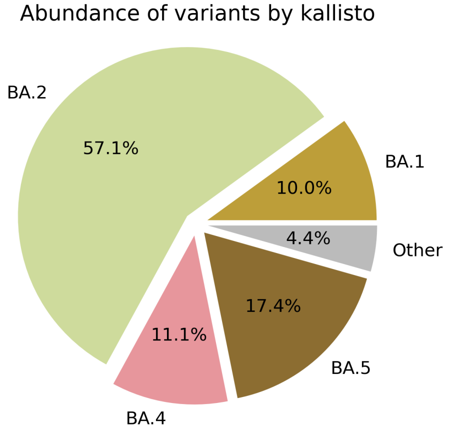


Based on deconvolution, B.1.1.529 is estimated to constitute 60.37% of the viral particles and hence is the most abundant variant in the sample. The $R^2$ for the linear regression was 0.61. Variants that were detected less than 5% were grouped under "Other"

Based on the consensus sequence of the observed reads, the "ensemble-averaged sequence" most closely resembles the BA.2 lineage. If this is a sample consisting of a single source of pathogens or an overwhelming majority of the different sources are infected with the same variant, the sample is dominated by this variant.

Based on mapping individual reads to the variant consensus sequences in the reference database, kallisto predicts that the sample is dominated by BA.2 lineage. Accuracy of this measure is expected to improve if the input data consists of long reads as opposed to convolution.

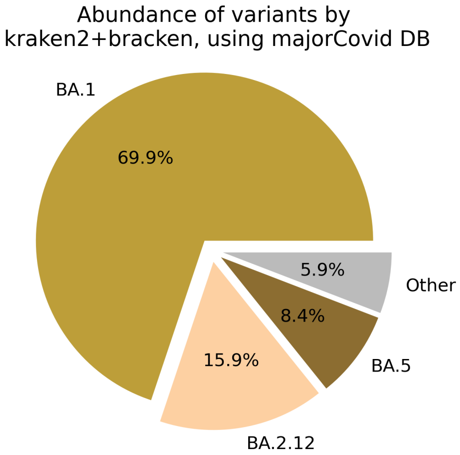


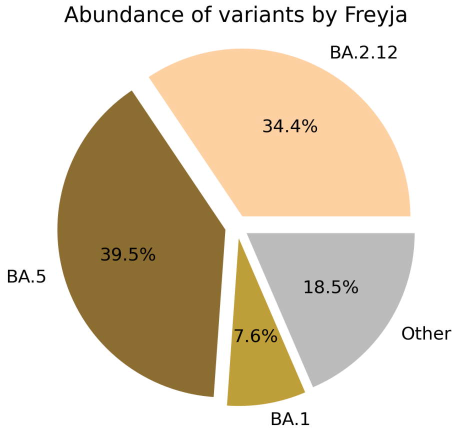

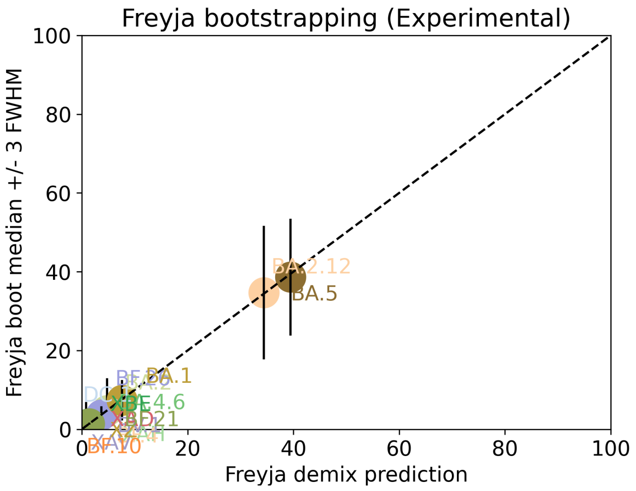


Under the assumption that the presence of a variant requires the detection of all respective mutations of the variant, the characteric mutations which support the presence of the respective variant are indicated in the respective column of the table. Numbers show the number of mutations detected, if any, and the number of mutations expected to be present based on the variant definitions.

| VOC | B.1.617.2 | BA.1 | BA.2 | BA.3 | BA.4 | BA.5 |
|---|---|---|---|---|---|---|
| Characteristic mutations detected | (4 of 13)<br>N:D377Y<br>S:G142D<br>S:L452R<br>S:T478K | (10 of 26)<br>NUC:C25000T<br>NUC:C25584T<br>NUC:T13195C<br>ORF1AB:A2710T<br>S:G446S<br>S:G496S<br>S:L981F<br>S:N856K<br>S:Q493R<br>S:T95I | (22 of 31)<br>N:S413R<br>NUC:A20055G<br>NUC:C10198T<br>NUC:C12880T<br>NUC:C15714T<br>NUC:C25000T<br>NUC:C25584T<br>NUC:C26858T<br>NUC:C4321T<br>NUC:G10447A<br>ORF1AB:G1307S<br>ORF1AB:L3027F<br>ORF1AB:L3201F<br>ORF1AB:S135R<br>ORF1AB:T3090I<br>ORF1AB:T842I<br>S:D405N<br>S:Q493R<br>S:R408S<br>S:S371F<br>S:T19I<br>S:T376A | (12 of 21)<br>N:S413R<br>NUC:C12880T<br>NUC:C15714T<br>NUC:C26858T<br>NUC:G10447A<br>ORF1AB:G1307S<br>ORF1AB:S135R<br>ORF1AB:T3090I<br>S:D405N<br>S:G446S<br>S:Q493R<br>S:S371F | (24 of 31)<br>N:P151S<br>N:S413R<br>NUC:A20055G<br>NUC:C10198T<br>NUC:C12880T<br>NUC:C15714T<br>NUC:C25000T<br>NUC:C25584T<br>NUC:C26858T<br>NUC:C4321T<br>NUC:G10447A<br>NUC:G12160A<br>NUC:G27788T<br>ORF1AB:G1307S<br>ORF1AB:S135R<br>ORF1AB:T3090I<br>ORF1AB:T842I<br>S:D405N<br>S:F486V<br>S:L452R<br>S:S371F<br>S:T19I<br>S:T376A<br>S:V213G | (22 of 28)<br>M:D3N<br>N:S413R<br>NUC:A20055G<br>NUC:C10198T<br>NUC:C12880T<br>NUC:C15714T<br>NUC:C25000T<br>NUC:C25584T<br>NUC:C4321T<br>NUC:G10447A<br>NUC:G12160A<br>ORF1AB:G1307S<br>ORF1AB:S135R<br>ORF1AB:T3090I<br>ORF1AB:T842I<br>S:D405N<br>S:F486V<br>S:L452R<br>S:S371F<br>S:T19I<br>S:T376A<br>S:V213G |

Jaccard Index is a measure of similarity between two sets A and B, reaching the maximum value of 1 if A=B and minimum value of 0 if A ∩ B = {}. In the c(d) representation below, c represents the Jaccard index of the set of mutations that were experimentally detected for this sample as listed above, whereas d refers to the ideal value of the Jaccard index expected from complete genome coverage without any sequencing errors.

| | B.1.617.2 | BA.1 | BA.2 | BA.3 | BA.4 | BA.5 |
|---|---|---|---|---|---|---|
| B.1.617.2 | 1.00 (1.00) | 0.00 (0.00) | 0.00 (0.00) | 0.00 (0.00) | 0.04 (0.02) | 0.04 (0.03) |
| BA.1 | 0.00 (0.00) | 1.00 (1.00) | 0.10 (0.10) | 0.10 (0.21) | 0.06 (0.08) | 0.07 (0.08) |
| BA.2 | 0.00 (0.00) | 0.10 (0.10) | 1.00 (1.00) | 0.48 (0.33) | 0.64 (0.63) | 0.63 (0.59) |
| BA.3 | 0.00 (0.00) | 0.10 (0.21) | 0.48 (0.33) | 1.00 (1.00) | 0.38 (0.30) | 0.36 (0.29) |

| | | | | | | |
|---|---|---|---|---|---|---|
| BA.4 | 0.04 (0.02) | 0.06 (0.08) | 0.64 (0.63) | 0.38 (0.30) | 1.00 (1.00) | 0.84 (0.84) |
| BA.5 | 0.04 (0.03) | 0.07 (0.08) | 0.63 (0.59) | 0.36 (0.29) | 0.84 (0.84) | 1.00 (1.00) |

#### Detected mutations

Excluded from this pdf version due to file size limitations.

CFSAN/OAO
BIOSTATISTICS AND BIOINFORMATICS STAFF

### WASTEWATER SARS-COV2 ANALYSIS REPORT

| | |
|---|---|
| Sample name: | CFSANSMP000118587 |
| Date generated: | 2023-02-01, 20:04:21 EST |
| Timestamp of C-WAP version used: | Tue Jan 31 11:49:22 2023 -0500 |
| Executed by: | Jasmine Amirzadegan |
| Executed on: | 172.20.44.127 (aka n127.raven.cfsan) |

#### Sequencing summary

| | |
|---|---|
| Sequencing chemistry: | Missing with Missing |
| Source site: | Missing (?,?) |
| Sampling date: | Missing |
| Collected by: | Missing |
| Sequenced by: | Missing |
| Total number of reads: | 2531278 |
| Reads aligned: | 1373578 (54%) |
| Average read quality: | 36.8 |
| Average read length: | 149 |
| Reads passing filter: | 1323983 (52%) |
| Average read quality passing filter: | 37.0 |
| Average read length passing filter: | 149 |
| Average coverage passing filter: | 6597X |

A read passes filter if the read length after adaptor trimming ≥ 30 and minimum read quality ≥ 20 within a sliding window of width 4.

#### Overall sequence characteristics

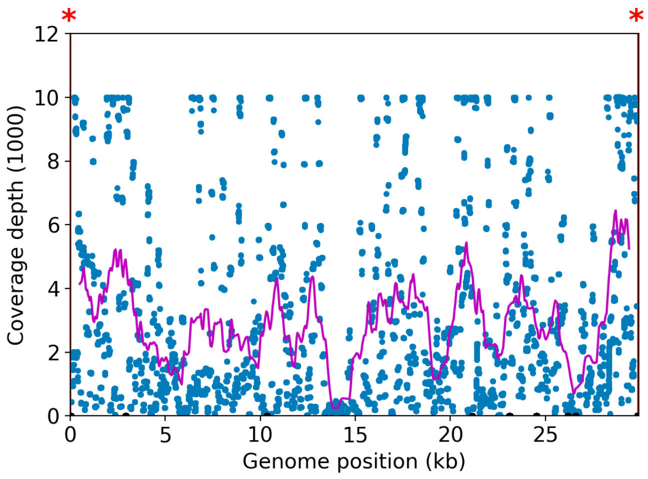


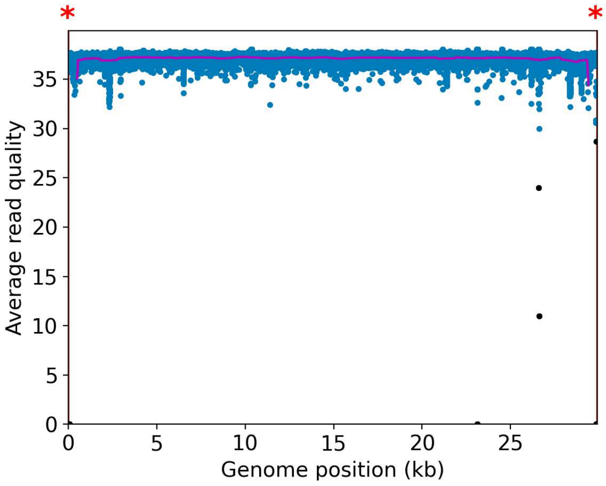


NOTE: The red shaded areas marked with a (*) are not covered by the design of the library preparation kit and hence excluded from analyses. Magenta curves represent moving average with a window width of 1kb.

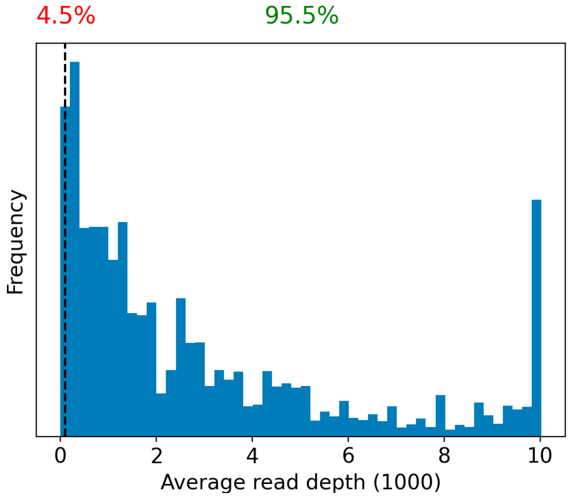


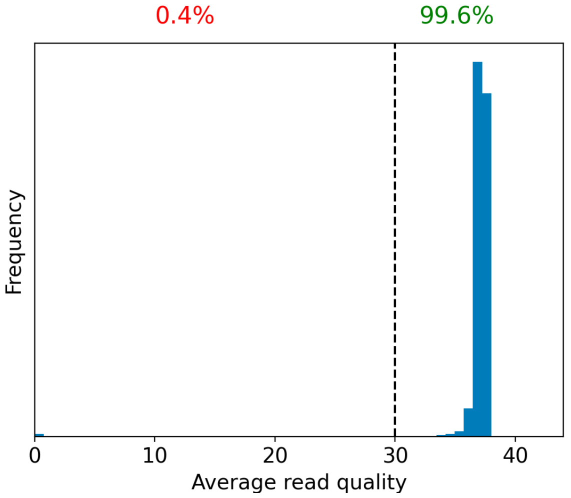


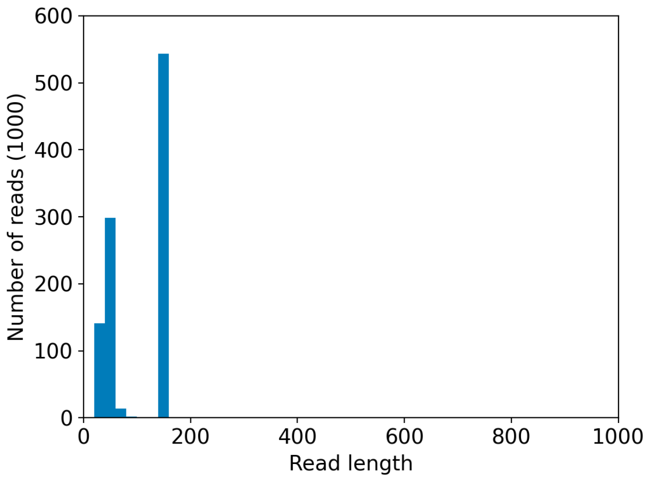


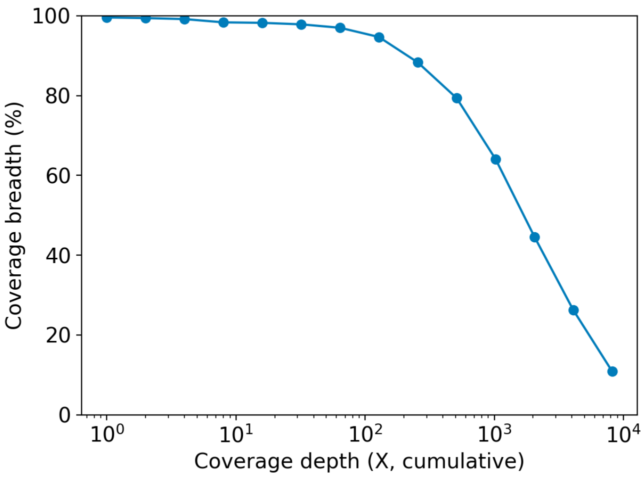

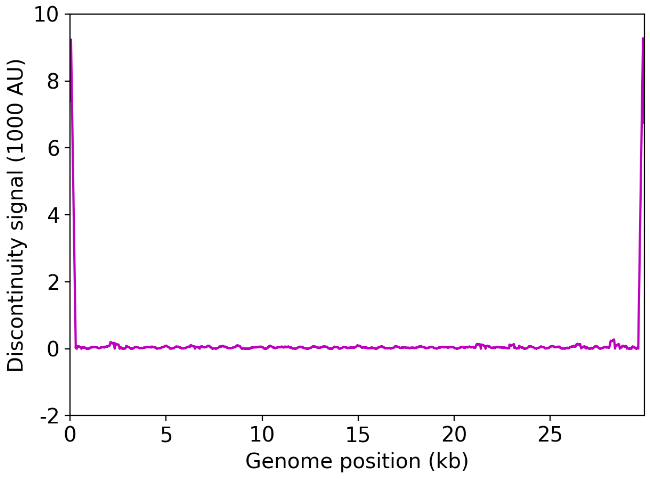


| | Uncovered coordinates (0X) | Poorly covered coordinates (<10X) |
|---|---|---|
| # Inaccessible genomic coordinates by kit design: | 121nt (0%) | 121nt (0%) |
| All genomic coordinates: | 121nt (0%) | 523nt (1%) |
| Common SNPs: | 0nt (0%) | 1nt (2%) |
| Diverse SNPs: | 29nt (12%) | 29nt (12%) |
| Rare SNPs: | 10nt (1%) | 47nt (5%) |

SNPs refer to the polymorphic sites currently in circulation that were detected out of recent GISAID entries. The sites that differ from the SC2 reference sequence are denoted as "common" if [90%, 100%] of the submissions carry this mutation, whereas those that are prevalent in [0%,10%] of the submissions are grouped under the "rare" category. The population is still diverse at the mutation sites that are observed in (10%,90%) of the entries and these coordinates are grouped under the "diverse" category.

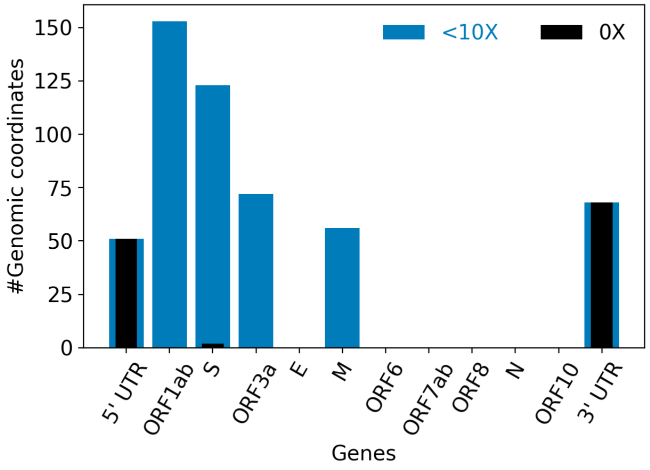


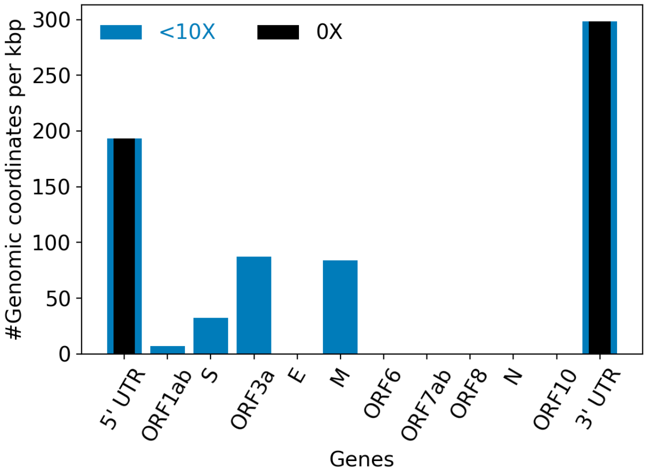


| | |
|---|---|
| Hits to SARS-Cov2 genome (kraken2): | 774679 reads (61.21%) |
| Hits to human genome (kraken2): | 734 reads (0.06%) |
| Hits to synthetic sequences (kraken2, taxid 28384): | 1357 reads (0.11%) |
| Most abundant organisms (kraken2, family level): | Coronaviridae (61.21%)<br>Enterobacteriaceae (0.46%)<br>Flavobacteriaceae (0.08%) |

| | |
|---|---|
| | Staphylococcaceae (0.07%)<br>Hominidae (0.06%) |

### Detected variants (Experimental)

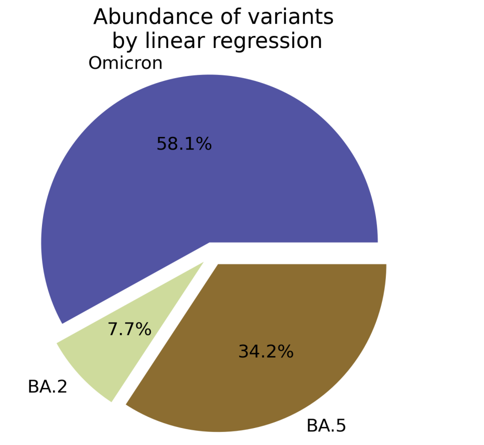


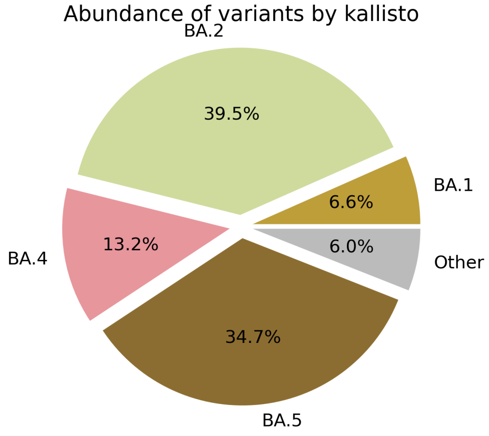


Based on deconvolution, B.1.1.529 is estimated to constitute 58.03% of the viral particles and hence is the most abundant variant in the sample. The $R^2$ for the linear regression was 0.60. Variants that were detected less than 5% were grouped under "Other"

Based on the consensus sequence of the observed reads, the "ensemble-averaged sequence" most closely resembles the BA.2 lineage. If this is a sample consisting of a single source of pathogens or an overwhelming majority of the different sources are infected with the same variant, the sample is dominated by this variant.

Based on mapping individual reads to the variant consensus sequences in the reference database, kallisto predicts that the sample is dominated by BA.2 lineage. Accuracy of this measure is expected to improve if the input data consists of long reads as opposed to convolution.

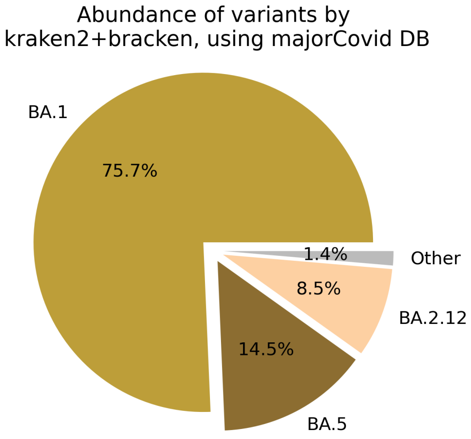


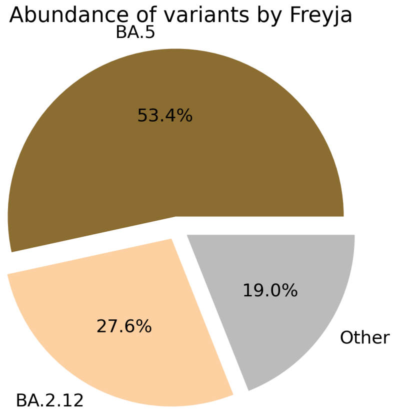

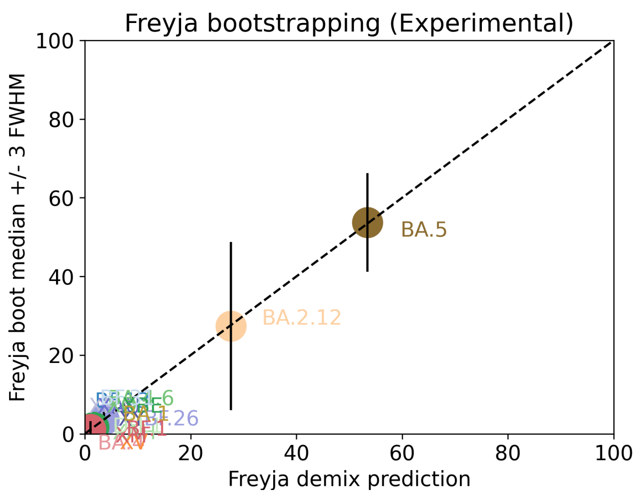


Under the assumption that the presence of a variant requires the detection of all respective mutations of the variant, the characteric mutations which support the presence of the respective variant are indicated in the respective column of the table. Numbers show the number of mutations detected, if any, and the number of mutations expected to be present based on the variant definitions.

| VOC | [B.1.617.2]() | [BA.1]() | [BA.2]() | [BA.3]() | [BA.4]() | [BA.5]() |
|---|---|---|---|---|---|---|
| Characteristic mutations detected | (4 of 13)<br>N:D377Y<br>S:G142D<br>S:L452R<br>S:T478K | (10 of 26)<br>NUC:C25000T<br>NUC:C25584T<br>NUC:T13195C<br>ORF1AB:A2710T<br>ORF1AB:I3758V<br>ORF1AB:K856R<br>S:G446S<br>S:L981F<br>S:N856K<br>S:Q493R | (22 of 31)<br>N:S413R<br>NUC:A20055G<br>NUC:C10198T<br>NUC:C12880T<br>NUC:C15714T<br>NUC:C25000T<br>NUC:C25584T<br>NUC:C26858T<br>NUC:C4321T<br>NUC:G10447A<br>ORF1AB:G1307S<br>ORF1AB:L3027F<br>ORF1AB:L3201F<br>ORF1AB:S135R<br>ORF1AB:T3090I<br>ORF1AB:T842I<br>S:D405N<br>S:Q493R<br>S:R408S<br>S:S371F<br>S:T19I<br>S:T376A | (12 of 21)<br>N:S413R<br>NUC:C12880T<br>NUC:C15714T<br>NUC:C26858T<br>NUC:G10447A<br>ORF1AB:G1307S<br>ORF1AB:S135R<br>ORF1AB:T3090I<br>S:D405N<br>S:G446S<br>S:Q493R<br>S:S371F | (24 of 31)<br>N:P151S<br>N:S413R<br>NUC:A20055G<br>NUC:C10198T<br>NUC:C12880T<br>NUC:C15714T<br>NUC:C25000T<br>NUC:C25584T<br>NUC:C26858T<br>NUC:C4321T<br>NUC:G10447A<br>NUC:G12160A<br>NUC:G27788T<br>ORF1AB:G1307S<br>ORF1AB:S135R<br>ORF1AB:T3090I<br>ORF1AB:T842I<br>S:D405N<br>S:F486V<br>S:L452R<br>S:S371F<br>S:T19I<br>S:T376A<br>S:V213G | (22 of 28)<br>M:D3N<br>N:S413R<br>NUC:A20055G<br>NUC:C10198T<br>NUC:C12880T<br>NUC:C15714T<br>NUC:C25000T<br>NUC:C25584T<br>NUC:C4321T<br>NUC:G10447A<br>NUC:G12160A<br>ORF1AB:G1307S<br>ORF1AB:S135R<br>ORF1AB:T3090I<br>ORF1AB:T842I<br>S:D405N<br>S:F486V<br>S:L452R<br>S:S371F<br>S:T19I<br>S:T376A<br>S:V213G |

[Jaccard Index]() is a measure of similarity between two sets A and B, reaching the maximum value of 1 if A=B and minimum value of 0 if A ∩ B = {}. In the c(d) representation below, c represents the Jaccard index of the set of mutations that were experimentally detected for this sample as listed above, whereas d refers to the ideal value of the Jaccard index expected from complete genome coverage without any sequencing errors.

| | B.1.617.2 | BA.1 | BA.2 | BA.3 | BA.4 | BA.5 |
|---|---|---|---|---|---|---|
| B.1.617.2 | 1.00 ([1.00]()) | 0.00 ([0.00]()) | 0.00 ([0.00]()) | 0.00 ([0.00]()) | 0.04 ([0.02]()) | 0.04 ([0.03]()) |
| BA.1 | 0.00 ([0.00]()) | 1.00 ([1.00]()) | 0.10 ([0.10]()) | 0.10 ([0.21]()) | 0.06 ([0.08]()) | 0.07 ([0.08]()) |
| BA.2 | 0.00 ([0.00]()) | 0.10 ([0.10]()) | 1.00 ([1.00]()) | 0.48 ([0.33]()) | 0.64 ([0.63]()) | 0.63 ([0.59]()) |
| BA.3 | 0.00 ([0.00]()) | 0.10 ([0.21]()) | 0.48 ([0.33]()) | 1.00 ([1.00]()) | 0.38 ([0.30]()) | 0.36 ([0.29]()) |

| | | | | | | |
|---|---|---|---|---|---|---|
| BA.4 | 0.04 (0.02) | 0.06 (0.08) | 0.64 (0.63) | 0.38 (0.30) | 1.00 (1.00) | 0.84 (0.84) |
| BA.5 | 0.04 (0.03) | 0.07 (0.08) | 0.63 (0.59) | 0.36 (0.29) | 0.84 (0.84) | 1.00 (1.00) |

#### Detected mutations

Excluded from this pdf version due to file size limitations.

U.S. FOOD & DRUG ADMINISTRATION

CFSAN/OAO
BIOSTATISTICS AND BIOINFORMATICS STAFF

### WASTEWATER SARS-COV2 ANALYSIS REPORT

| | |
|---|---|
| Sample name: | CFSANSMP000119220 |
| Date generated: | 2023-02-01, 20:06:45 EST |
| Timestamp of C-WAP version used: | Tue Jan 31 11:49:22 2023 -0500 |
| Executed by: | Jasmine Amirzadegan |
| Executed on: | 172.20.44.127 (aka n127.raven.cfsan) |

#### Sequencing summary

| | |
|---|---|
| Sequencing chemistry: | Missing with Missing |
| Source site: | Missing (?,?) |
| Sampling date: | Missing |
| Collected by: | Missing |
| Sequenced by: | Missing |
| Total number of reads: | 2955272 |
| Reads aligned: | 1563410 (52%) |
| Average read quality: | 36.6 |
| Average read length: | 149 |
| Reads passing filter: | 1493268 (50%) |
| Average read quality passing filter: | 36.9 |
| Average read length passing filter: | 149 |
| Average coverage passing filter: | 7440X |

A read passes filter if the read length after adaptor trimming ≥ 30 and minimum read quality ≥ 20 within a sliding window of width 4.

#### Overall sequence characteristics

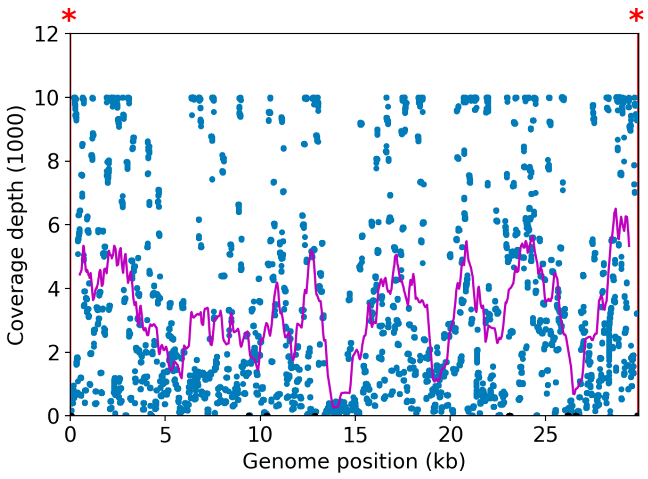


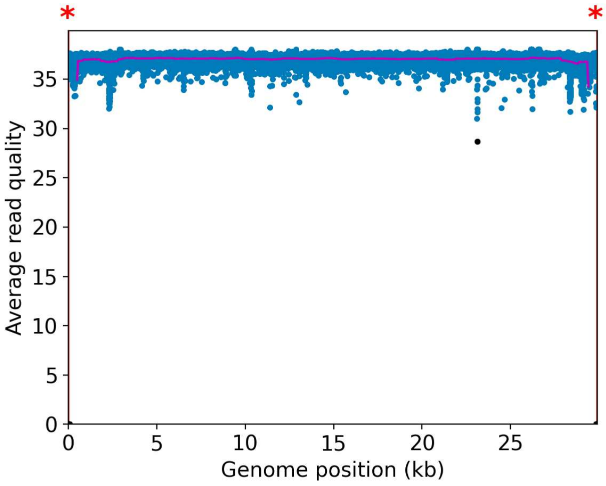


NOTE: The red shaded areas marked with a (*) are not covered by the design of the library preparation kit and hence excluded from analyses. Magenta curves represent moving average with a window width of 1kb.

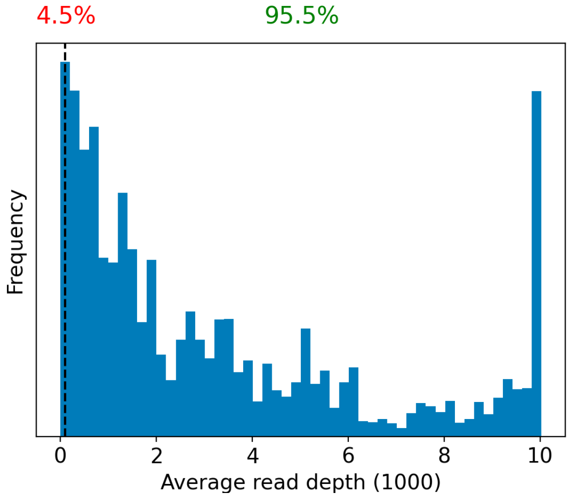


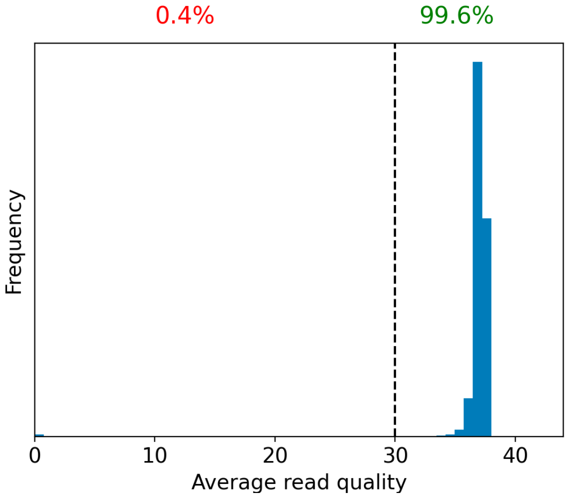


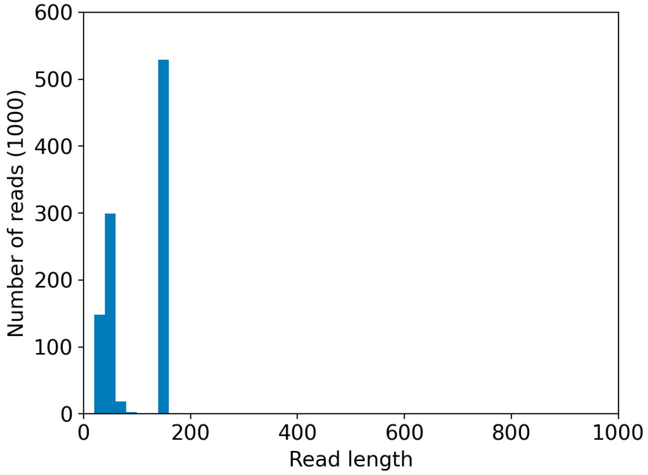


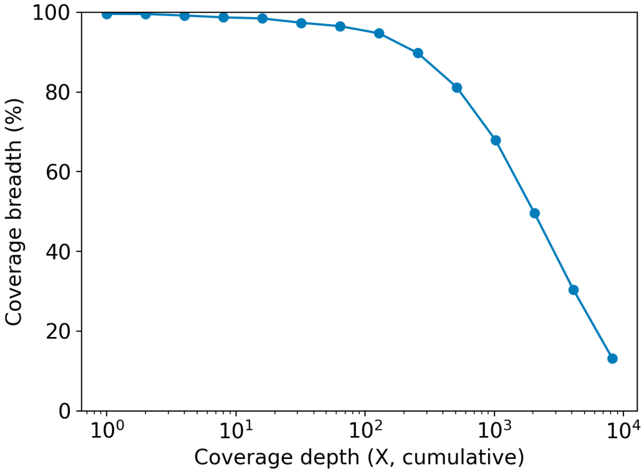

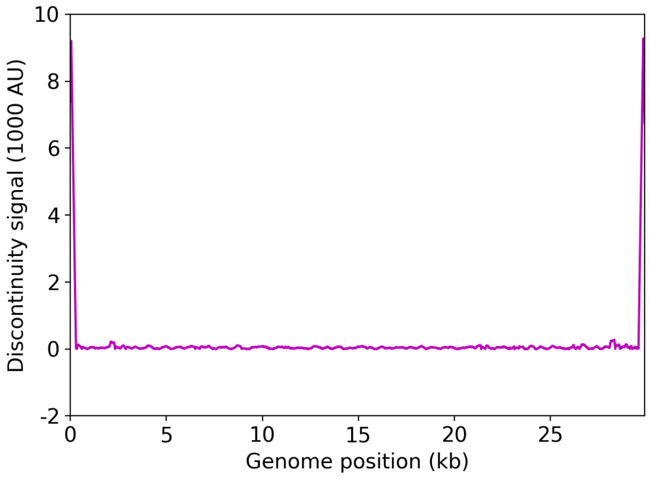


| | Uncovered coordinates (0X) | Poorly covered coordinates (<10X) |
|---|---|---|
| # Inaccessible genomic coordinates by kit design: | 121nt (0%) | 121nt (0%) |
| All genomic coordinates: | 119nt (0%) | 393nt (1%) |
| Common SNPs: | 0nt (0%) | 1nt (2%) |
| Diverse SNPs: | 29nt (12%) | 29nt (12%) |
| Rare SNPs: | 10nt (1%) | 47nt (5%) |

SNPs refer to the polymorphic sites currently in circulation that were detected out of recent GISAID entries. The sites that differ from the SC2 reference sequence are denoted as "common" if [90%, 100%] of the submissions carry this mutation, whereas those that are prevalent in [0%,10%] of the submissions are grouped under the "rare" category. The population is still diverse at the mutation sites that are observed in (10%,90%) of the entries and these coordinates are grouped under the "diverse" category.

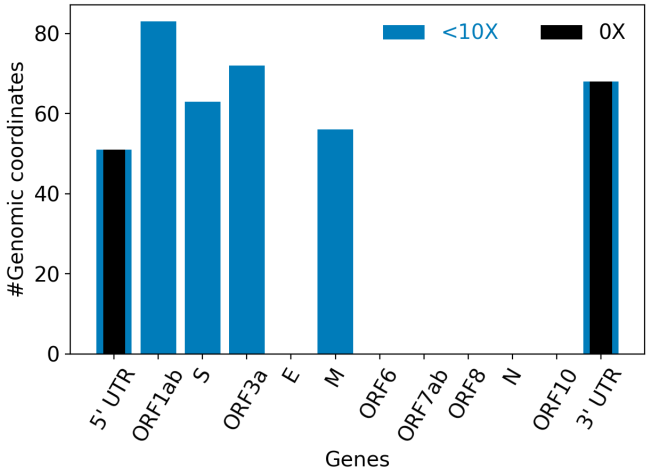


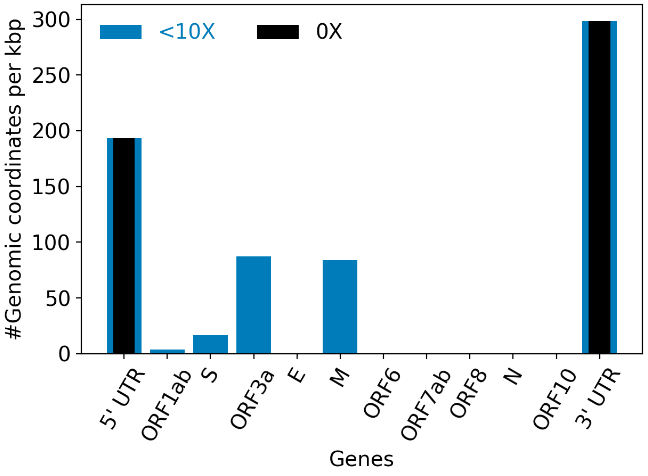


| | |
|---|---|
| Hits to SARS-Cov2 genome (kraken2): | 886306 reads (59.98%) |
| Hits to human genome (kraken2): | 1255 reads (0.08%) |
| Hits to synthetic sequences (kraken2, taxid 28384): | 165 reads (0.01%) |
| Most abundant organisms (kraken2, family level): | Coronaviridae (59.98%)<br>Enterococcaceae (0.61%)<br>Hominidae (0.08%) |

| | |
|---|---|
| | Arcobacteraceae (0.07%)<br>Burkholderiaceae (0.04%) |

### Detected variants (Experimental)

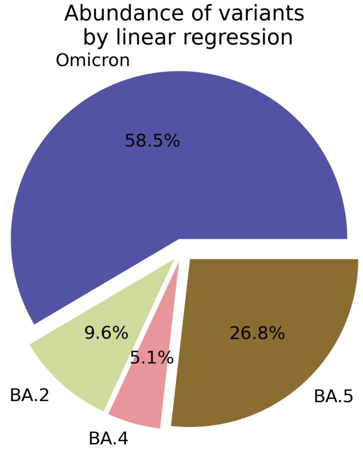


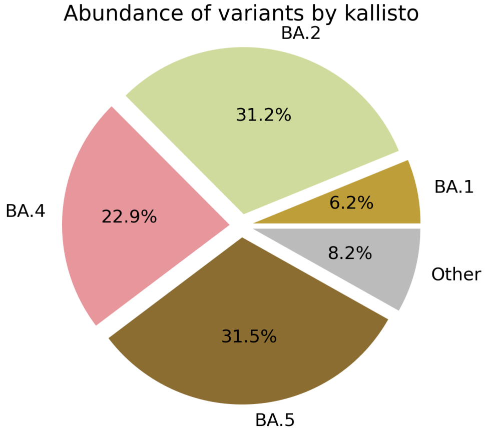


Based on deconvolution, B.1.1.529 is estimated to constitute 58.51% of the viral particles and hence is the most abundant variant in the sample. The $R^2$ for the linear regression was 0.60. Variants that were detected less than 5% were grouped under "Other"

Based on the consensus sequence of the observed reads, the "ensemble-averaged sequence" most closely resembles the BA.2 lineage. If this is a sample consisting of a single source of pathogens or an overwhelming majority of the different sources are infected with the same variant, the sample is dominated by this variant.

Based on mapping individual reads to the variant consensus sequences in the reference database, kallisto predicts that the sample is dominated by BA.5 lineage. Accuracy of this measure is expected to improve if the input data consists of long reads as opposed to convolution.

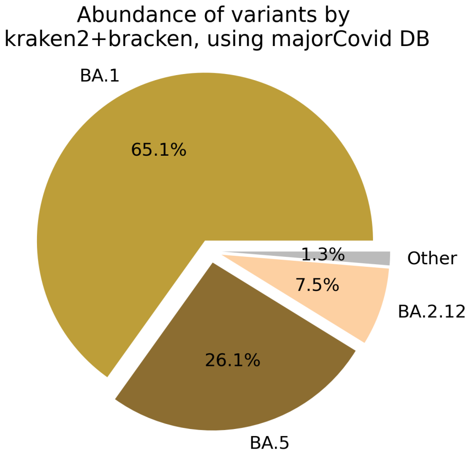


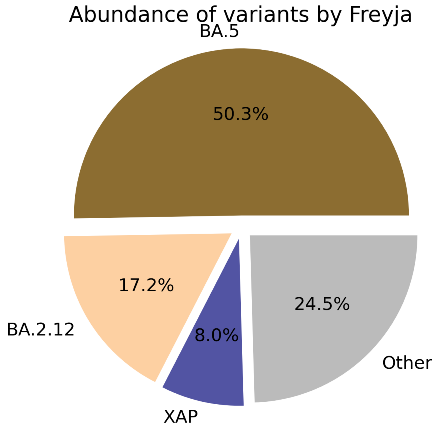

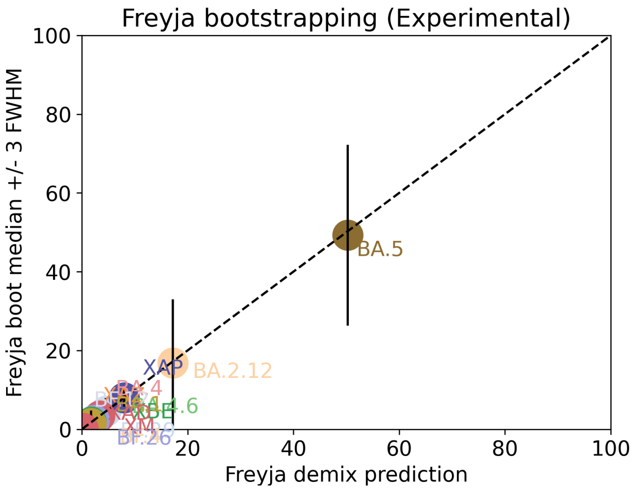


Under the assumption that the presence of a variant requires the detection of all respective mutations of the variant, the characteric mutations which support the presence of the respective variant are indicated in the respective column of the table. Numbers show the number of mutations detected, if any, and the number of mutations expected to be present based on the variant definitions.

| VOC | B.1.617.2 | BA.1 | BA.2 | BA.3 | BA.4 | BA.5 |
|---|---|---|---|---|---|---|
| Characteristic mutations detected | (4 of 13)<br>N:D377Y<br>S:G142D<br>S:L452R<br>S:T478K | (10 of 26)<br>NUC:C15240T<br>NUC:C25000T<br>NUC:C25584T<br>NUC:T13195C<br>ORF1AB:I3758V<br>ORF1AB:K856R<br>S:L981F<br>S:N856K<br>S:Q493R<br>S:T95I | (22 of 31)<br>N:S413R<br>NUC:A20055G<br>NUC:C10198T<br>NUC:C12880T<br>NUC:C15714T<br>NUC:C25000T<br>NUC:C25584T<br>NUC:C26858T<br>NUC:C4321T<br>NUC:G10447A<br>ORF1AB:G1307S<br>ORF1AB:L3027F<br>ORF1AB:L3201F<br>ORF1AB:S135R<br>ORF1AB:T3090I<br>ORF1AB:T842I<br>S:D405N<br>S:Q493R<br>S:R408S<br>S:S371F<br>S:T19I<br>S:T376A | (11 of 21)<br>N:S413R<br>NUC:C12880T<br>NUC:C15714T<br>NUC:C26858T<br>NUC:G10447A<br>ORF1AB:G1307S<br>ORF1AB:S135R<br>ORF1AB:T3090I<br>S:D405N<br>S:Q493R<br>S:S371F | (24 of 31)<br>N:P151S<br>N:S413R<br>NUC:A20055G<br>NUC:C10198T<br>NUC:C12880T<br>NUC:C15714T<br>NUC:C25000T<br>NUC:C25584T<br>NUC:C26858T<br>NUC:C4321T<br>NUC:G10447A<br>NUC:G12160A<br>NUC:G27788T<br>ORF1AB:G1307S<br>ORF1AB:S135R<br>ORF1AB:T3090I<br>ORF1AB:T842I<br>S:D405N<br>S:F486V<br>S:L452R<br>S:S371F<br>S:T19I<br>S:T376A<br>S:V213G | (22 of 28)<br>M:D3N<br>N:S413R<br>NUC:A20055G<br>NUC:C10198T<br>NUC:C12880T<br>NUC:C15714T<br>NUC:C25000T<br>NUC:C25584T<br>NUC:C4321T<br>NUC:G10447A<br>NUC:G12160A<br>ORF1AB:G1307S<br>ORF1AB:S135R<br>ORF1AB:T3090I<br>ORF1AB:T842I<br>S:D405N<br>S:F486V<br>S:L452R<br>S:S371F<br>S:T19I<br>S:T376A<br>S:V213G |

Jaccard Index is a measure of similarity between two sets A and B, reaching the maximum value of 1 if A=B and minimum value of 0 if A ∩ B = {}. In the c(d) representation below, c represents the Jaccard index of the set of mutations that were experimentally detected for this sample as listed above, whereas d refers to the ideal value of the Jaccard index expected from complete genome coverage without any sequencing errors.

| | B.1.617.2 | BA.1 | BA.2 | BA.3 | BA.4 | BA.5 |
|---|---|---|---|---|---|---|
| B.1.617.2 | 1.00 (1.00) | 0.00 (0.00) | 0.00 (0.00) | 0.00 (0.00) | 0.04 (0.02) | 0.04 (0.03) |
| BA.1 | 0.00 (0.00) | 1.00 (1.00) | 0.10 (0.10) | 0.05 (0.21) | 0.06 (0.08) | 0.07 (0.08) |
| BA.2 | 0.00 (0.00) | 0.10 (0.10) | 1.00 (1.00) | 0.50 (0.33) | 0.64 (0.63) | 0.63 (0.59) |
| BA.3 | 0.00 (0.00) | 0.05 (0.21) | 0.50 (0.33) | 1.00 (1.00) | 0.40 (0.30) | 0.38 (0.29) |

| | | | | | | |
|---|---|---|---|---|---|---|
| BA.4 | 0.04 (0.02) | 0.06 (0.08) | 0.64 (0.63) | 0.40 (0.30) | 1.00 (1.00) | 0.84 (0.84) |
| BA.5 | 0.04 (0.03) | 0.07 (0.08) | 0.63 (0.59) | 0.38 (0.29) | 0.84 (0.84) | 1.00 (1.00) |

#### Detected mutations

Excluded from this pdf version due to file size limitations.

CFSAN/OAO
BIOSTATISTICS AND BIOINFORMATICS STAFF

### WASTEWATER SARS-COV2 ANALYSIS REPORT

| | |
|---|---|
| Sample name: | CFSANSMP000119222 |
| Date generated: | 2023-02-01, 20:04:39 EST |
| Timestamp of C-WAP version used: | Tue Jan 31 11:49:22 2023 -0500 |
| Executed by: | Jasmine Amirzadegan |
| Executed on: | 172.20.44.127 (aka n127.raven.cfsan) |

#### Sequencing summary

| | |
|---|---|
| Sequencing chemistry: | Missing with Missing |
| Source site: | Missing (?,?) |
| Sampling date: | Missing |
| Collected by: | Missing |
| Sequenced by: | Missing |
| Total number of reads: | 2214544 |
| Reads aligned: | 1699294 (76%) |
| Average read quality: | 36.8 |
| Average read length: | 149 |
| Reads passing filter: | 1635964 (73%) |
| Average read quality passing filter: | 37.0 |
| Average read length passing filter: | 149 |
| Average coverage passing filter: | 8151X |

A read passes filter if the read length after adaptor trimming ≥ 30 and minimum read quality ≥ 20 within a sliding window of width 4.

#### Overall sequence characteristics

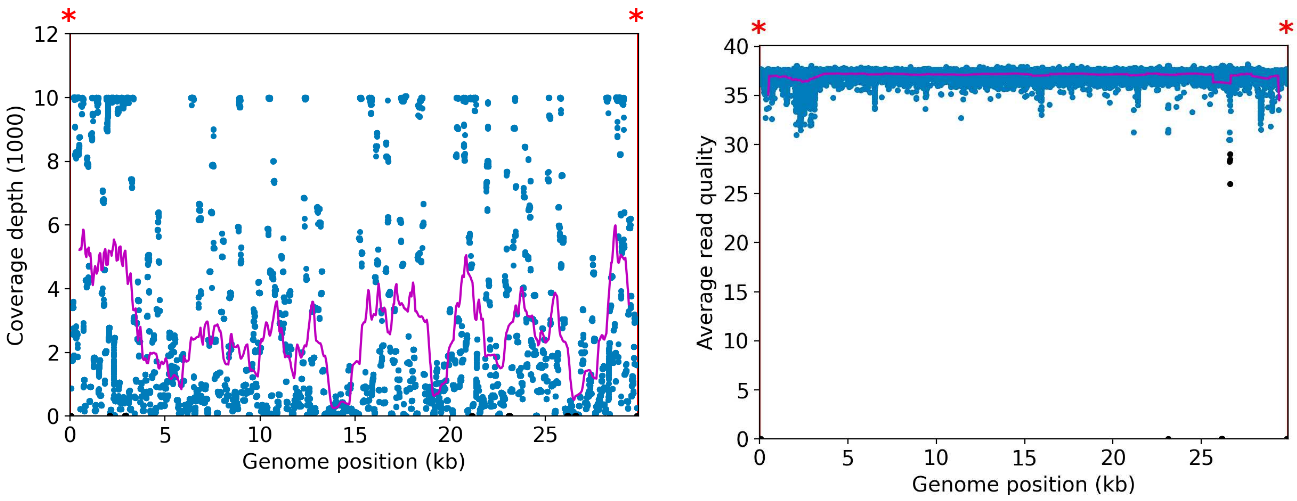


NOTE: The red shaded areas marked with a (*) are not covered by the design of the library preparation kit and hence excluded from analyses. Magenta curves represent moving average with a window width of 1kb.

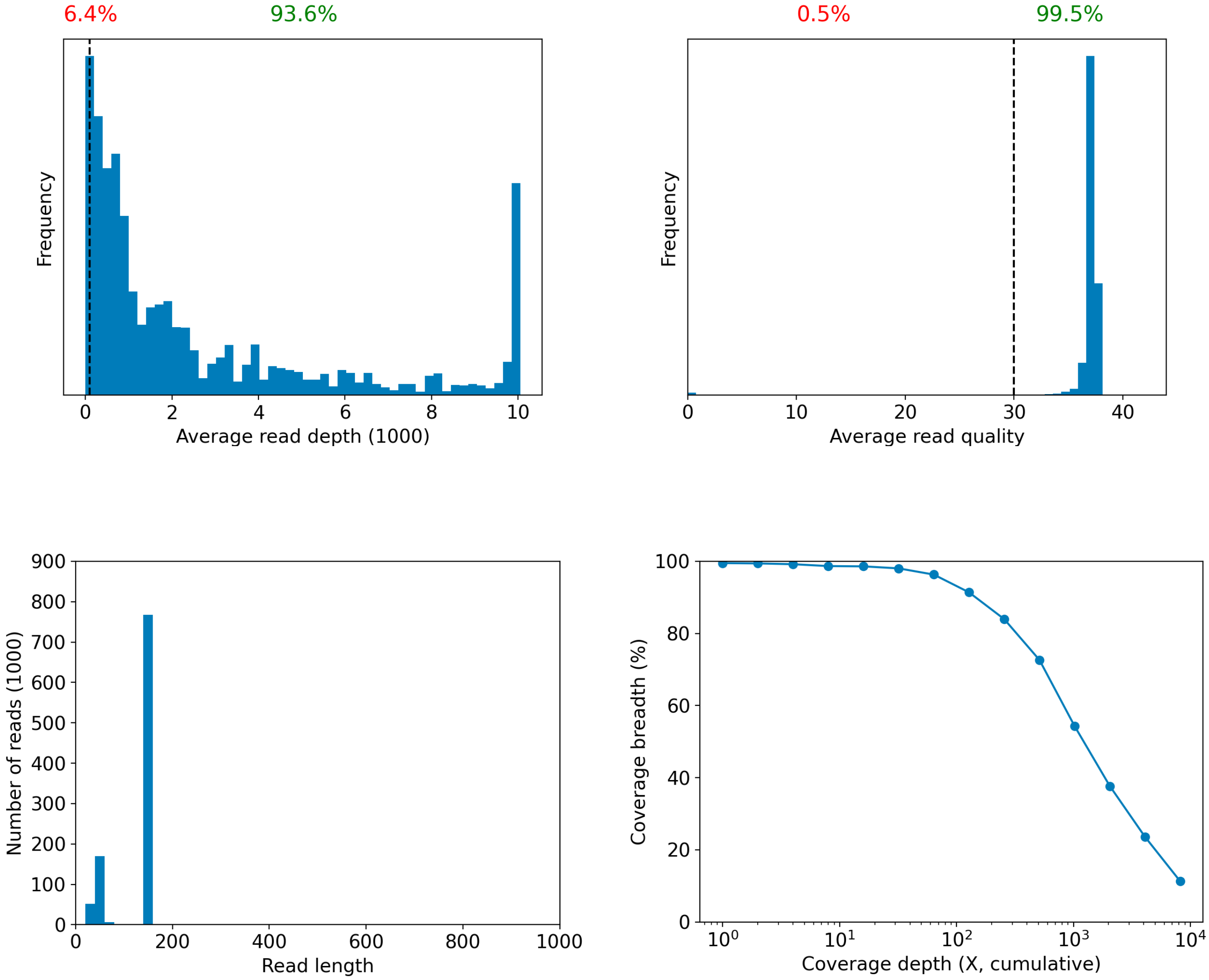

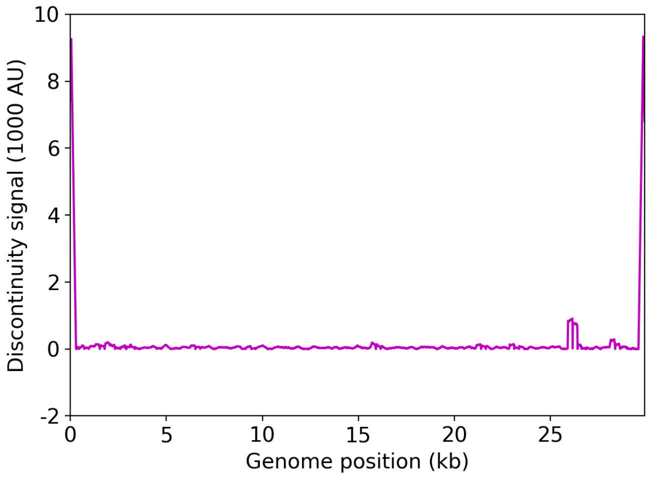


| | Uncovered coordinates (0X) | Poorly covered coordinates (<10X) |
|---|---|---|
| # Inaccessible genomic coordinates by kit design: | 121nt (0%) | 121nt (0%) |
| All genomic coordinates: | 144nt (0%) | 392nt (1%) |
| Common SNPs: | 0nt (0%) | 1nt (2%) |
| Diverse SNPs: | 29nt (12%) | 29nt (12%) |
| Rare SNPs: | 10nt (1%) | 47nt (5%) |

SNPs refer to the polymorphic sites currently in circulation that were detected out of recent GISAID entries. The sites that differ from the SC2 reference sequence are denoted as "common" if [90%, 100%] of the submissions carry this mutation, whereas those that are prevalent in [0%,10%] of the submissions are grouped under the "rare" category. The population is still diverse at the mutation sites that are observed in (10%,90%) of the entries and these coordinates are grouped under the "diverse" category.

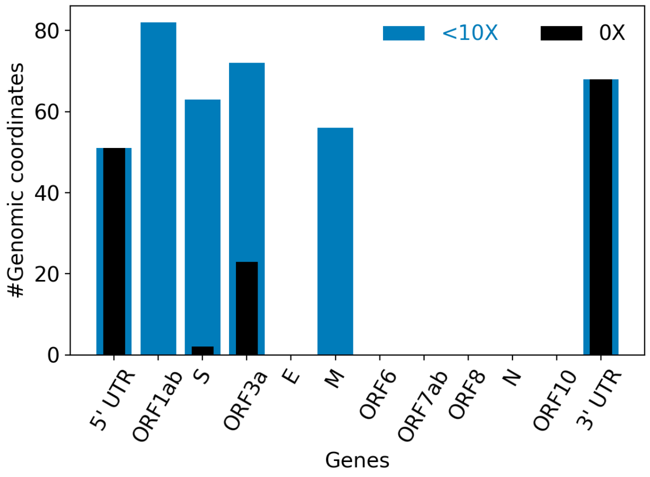


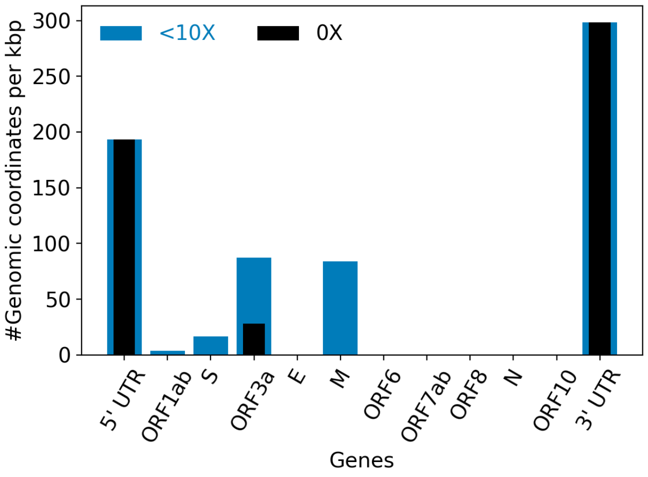


| | |
|---|---|
| Hits to SARS-Cov2 genome (kraken2): | 880953 reads (79.56%) |
| Hits to human genome (kraken2): | 515 reads (0.05%) |
| Hits to synthetic sequences (kraken2, taxid 28384): | 114 reads (0.01%) |
| Most abundant organisms (kraken2, family level): | Coronaviridae (79.56%)<br>Staphylococcaceae (0.29%)<br>Hominidae (0.05%) |

| | |
|---|---|
| | Mycobacteriaceae (0.01%)<br>Bacteroidaceae (0.01%) |

### Detected variants (Experimental)

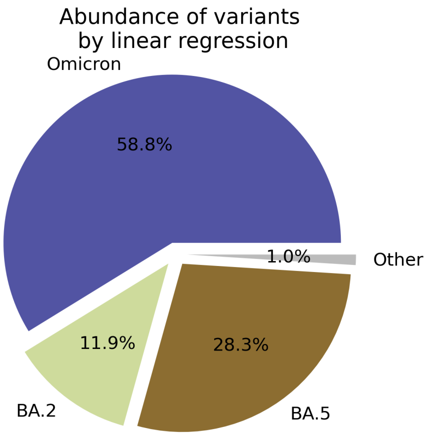


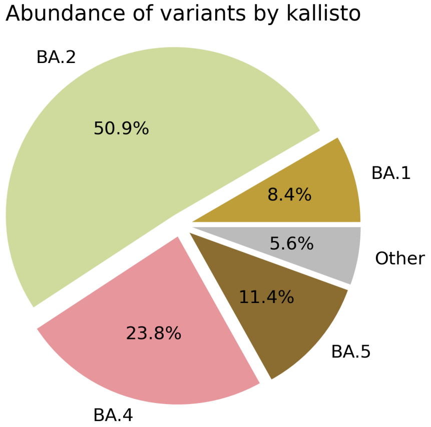


Based on deconvolution, B.1.1.529 is estimated to constitute 58.85% of the viral particles and hence is the most abundant variant in the sample. The $R^2$ for the linear regression was 0.59. Variants that were detected less than 5% were grouped under "Other"

Based on the consensus sequence of the observed reads, the "ensemble-averaged sequence" most closely resembles the BA.2 lineage. If this is a sample consisting of a single source of pathogens or an overwhelming majority of the different sources are infected with the same variant, the sample is dominated by this variant.

Based on mapping individual reads to the variant consensus sequences in the reference database, kallisto predicts that the sample is dominated by BA.2 lineage. Accuracy of this measure is expected to improve if the input data consists of long reads as opposed to convolution.

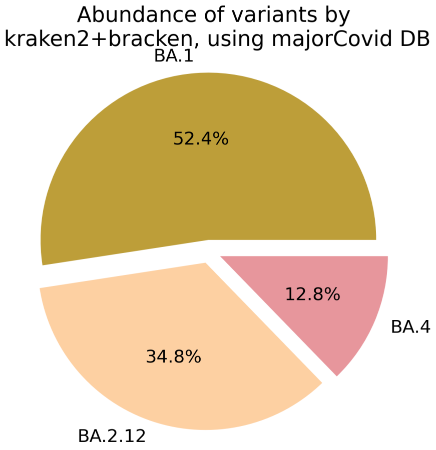


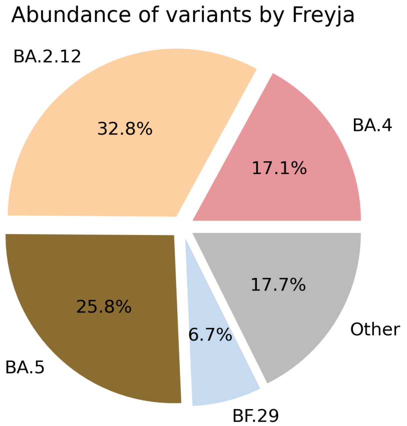

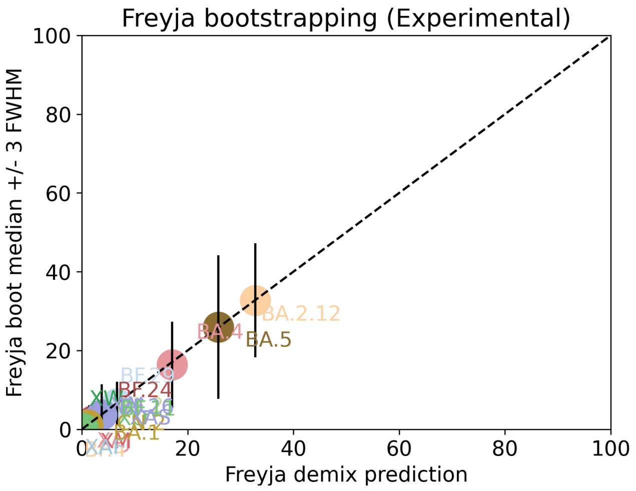


Under the assumption that the presence of a variant requires the detection of all respective mutations of the variant, the characteric mutations which support the presence of the respective variant are indicated in the respective column of the table. Numbers show the number of mutations detected, if any, and the number of mutations expected to be present based on the variant definitions.

| VOC | B.1.617.2 | BA.1 | BA.2 | BA.3 | BA.4 | BA.5 |
|---|---|---|---|---|---|---|
| Characteristic mutations detected | (3 of 13)<br>S:G142D<br>S:L452R<br>S:T478K | (10 of 26)<br>NUC:C25000T<br>NUC:C25584T<br>NUC:T13195C<br>ORF1AB:A2710T<br>ORF1AB:K856R<br>S:A67V<br>S:L981F<br>S:N856K<br>S:Q493R<br>S:T547K | (22 of 31)<br>N:S413R<br>NUC:A20055G<br>NUC:C10198T<br>NUC:C12880T<br>NUC:C15714T<br>NUC:C25000T<br>NUC:C25584T<br>NUC:C26858T<br>NUC:C4321T<br>NUC:G10447A<br>ORF1AB:G1307S<br>ORF1AB:L3027F<br>ORF1AB:L3201F<br>ORF1AB:S135R<br>ORF1AB:T3090I<br>ORF1AB:T842I<br>S:D405N<br>S:Q493R<br>S:R408S<br>S:S371F<br>S:T19I<br>S:T376A | (12 of 21)<br>N:S413R<br>NUC:C12880T<br>NUC:C15714T<br>NUC:C26858T<br>NUC:G10447A<br>ORF1AB:G1307S<br>ORF1AB:S135R<br>ORF1AB:T3090I<br>S:A67V<br>S:D405N<br>S:Q493R<br>S:S371F | (24 of 31)<br>N:P151S<br>N:S413R<br>NUC:A20055G<br>NUC:C10198T<br>NUC:C12880T<br>NUC:C15714T<br>NUC:C25000T<br>NUC:C25584T<br>NUC:C26858T<br>NUC:C4321T<br>NUC:G10447A<br>NUC:G12160A<br>NUC:G27788T<br>ORF1AB:G1307S<br>ORF1AB:S135R<br>ORF1AB:T3090I<br>ORF1AB:T842I<br>S:D405N<br>S:F486V<br>S:L452R<br>S:S371F<br>S:T19I<br>S:T376A<br>S:V213G | (22 of 28)<br>M:D3N<br>N:S413R<br>NUC:A20055G<br>NUC:C10198T<br>NUC:C12880T<br>NUC:C15714T<br>NUC:C25000T<br>NUC:C25584T<br>NUC:C4321T<br>NUC:G10447A<br>NUC:G12160A<br>ORF1AB:G1307S<br>ORF1AB:S135R<br>ORF1AB:T3090I<br>ORF1AB:T842I<br>S:D405N<br>S:F486V<br>S:L452R<br>S:S371F<br>S:T19I<br>S:T376A<br>S:V213G |

Jaccard Index is a measure of similarity between two sets A and B, reaching the maximum value of 1 if A=B and minimum value of 0 if A ∩ B = {}. In the c(d) representation below, c represents the Jaccard index of the set of mutations that were experimentally detected for this sample as listed above, whereas d refers to the ideal value of the Jaccard index expected from complete genome coverage without any sequencing errors.

| | B.1.617.2 | BA.1 | BA.2 | BA.3 | BA.4 | BA.5 |
|---|---|---|---|---|---|---|
| B.1.617.2 | 1.00 (1.00) | 0.00 (0.00) | 0.00 (0.00) | 0.00 (0.00) | 0.04 (0.02) | 0.04 (0.03) |
| BA.1 | 0.00 (0.00) | 1.00 (1.00) | 0.10 (0.10) | 0.10 (0.21) | 0.06 (0.08) | 0.07 (0.08) |
| BA.2 | 0.00 (0.00) | 0.10 (0.10) | 1.00 (1.00) | 0.48 (0.33) | 0.64 (0.63) | 0.63 (0.59) |
| BA.3 | 0.00 (0.00) | 0.10 (0.21) | 0.48 (0.33) | 1.00 (1.00) | 0.38 (0.30) | 0.36 (0.29) |

| | | | | | | |
|---|---|---|---|---|---|---|
| BA.4 | 0.04 (0.02) | 0.06 (0.08) | 0.64 (0.63) | 0.38 (0.30) | 1.00 (1.00) | 0.84 (0.84) |
| BA.5 | 0.04 (0.03) | 0.07 (0.08) | 0.63 (0.59) | 0.36 (0.29) | 0.84 (0.84) | 1.00 (1.00) |

#### Detected mutations

Excluded from this pdf version due to file size limitations.

FDA U.S. FOOD & DRUG ADMINISTRATION

CFSAN/OAO
BIOSTATISTICS AND BIOINFORMATICS STAFF

### WASTEWATER SARS-COV2 ANALYSIS REPORT

| | |
|---|---|
| Sample name: | CFSANSMP000119223 |
| Date generated: | 2023-02-01, 20:09:40 EST |
| Timestamp of C-WAP version used: | Tue Jan 31 11:49:22 2023 -0500 |
| Executed by: | Jasmine Amirzadegan |
| Executed on: | 172.20.44.108 (aka n108.raven.cfsan) |

#### Sequencing summary

| | |
|---|---|
| Sequencing chemistry: | Missing with Missing |
| Source site: | Missing (?,?) |
| Sampling date: | Missing |
| Collected by: | Missing |
| Sequenced by: | Missing |
| Total number of reads: | 4089396 |
| Reads aligned: | 3400167 (83%) |
| Average read quality: | 36.4 |
| Average read length: | 149 |
| Reads passing filter: | 3182172 (77%) |
| Average read quality passing filter: | 36.8 |
| Average read length passing filter: | 149 |
| Average coverage passing filter: | 15856X |

A read passes filter if the read length after adaptor trimming ≥ 30 and minimum read quality ≥ 20 within a sliding window of width 4.

#### Overall sequence characteristics

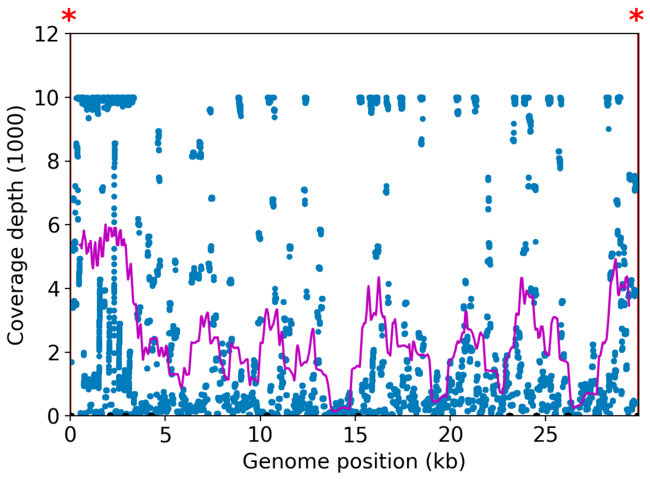


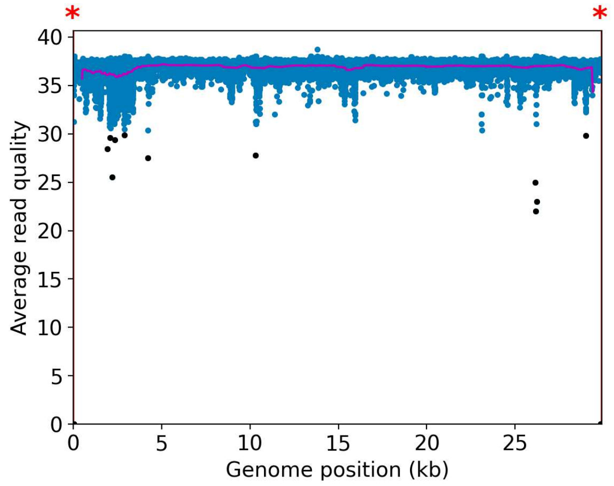


NOTE: The red shaded areas marked with a (*) are not covered by the design of the library preparation kit and hence excluded from analyses. Magenta curves represent moving average with a window width of 1kb.

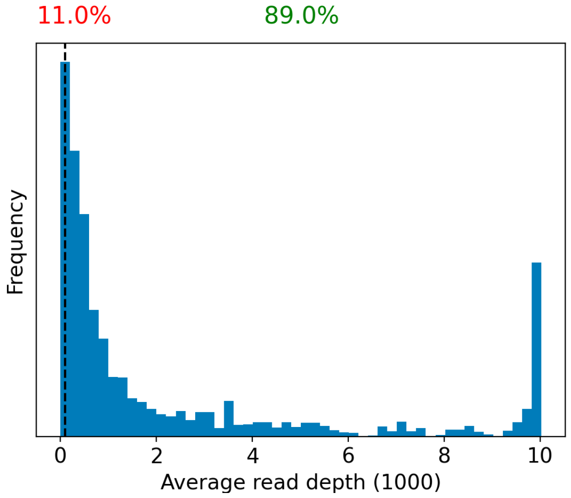


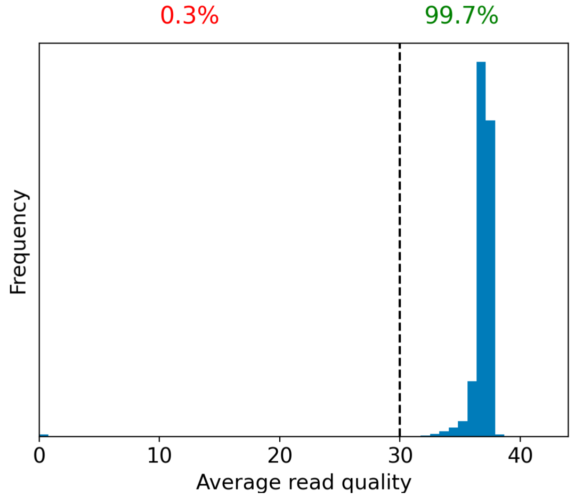


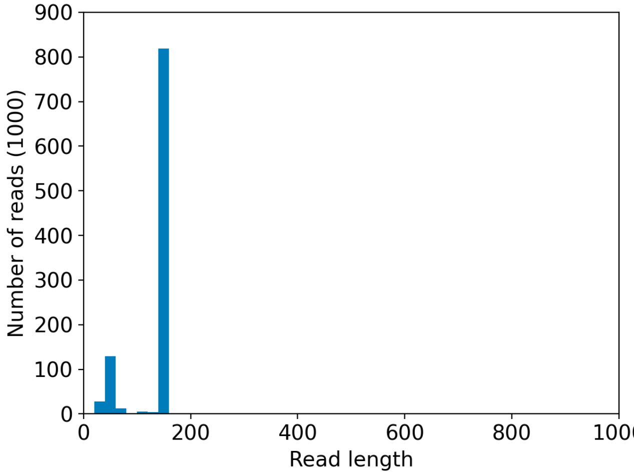


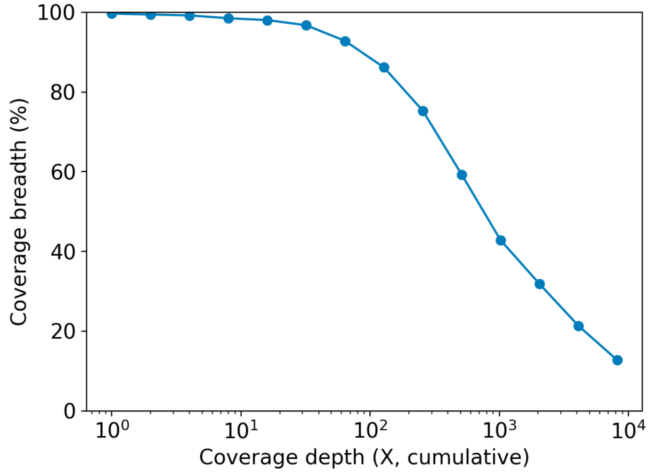

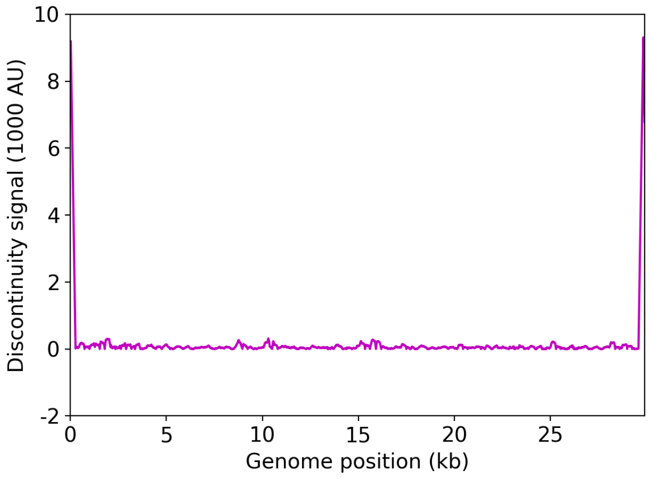


| | Uncovered coordinates (0X) | Poorly covered coordinates (<10X) |
|---|---|---|
| # Inaccessible genomic coordinates by kit design: | 121nt (0%) | 121nt (0%) |
| All genomic coordinates: | 92nt (0%) | 504nt (1%) |
| Common SNPs: | 0nt (0%) | 0nt (0%) |
| Diverse SNPs: | 28nt (12%) | 29nt (12%) |
| Rare SNPs: | 9nt (0%) | 46nt (4%) |

SNPs refer to the polymorphic sites currently in circulation that were detected out of recent GISAID entries. The sites that differ from the SC2 reference sequence are denoted as "common" if [90%, 100%] of the submissions carry this mutation, whereas those that are prevalent in [0%,10%] of the submissions are grouped under the "rare" category. The population is still diverse at the mutation sites that are observed in (10%,90%) of the entries and these coordinates are grouped under the "diverse" category.

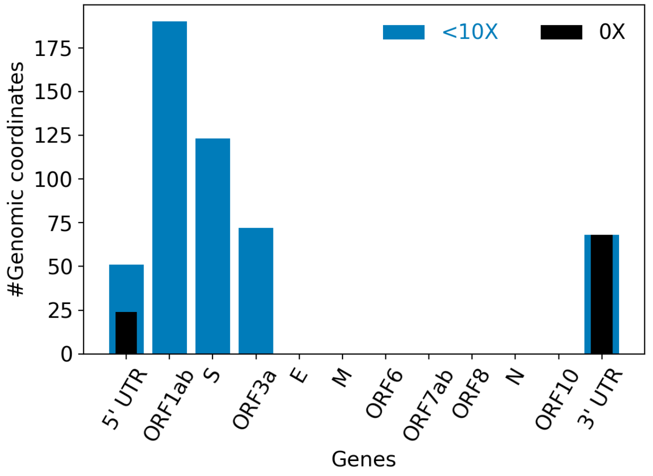


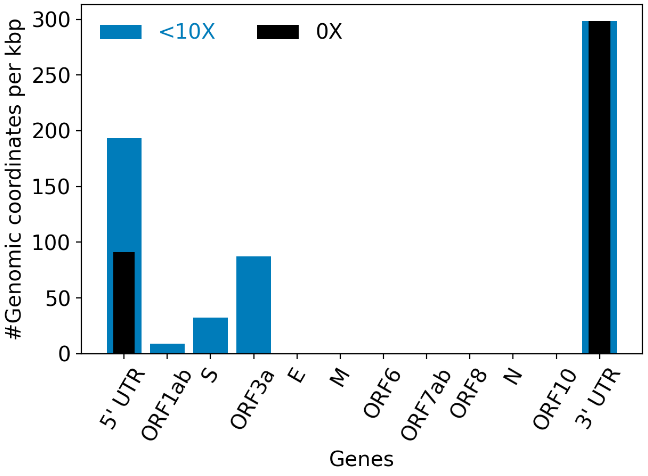


| | |
|---|---|
| Hits to SARS-Cov2 genome (kraken2): | 1770380 reads (86.58%) |
| Hits to human genome (kraken2): | 651 reads (0.03%) |
| Hits to synthetic sequences (kraken2, taxid 28384): | 294 reads (0.01%) |
| Most abundant organisms (kraken2, family level): | Coronaviridae (86.58%)<br>Enterococcaceae (0.18%)<br>Hominidae (0.03%) |

| | |
|---|---|
| | Bacteroidaceae (0.01%)<br>Staphylococcaceae (0.01%) |

### Detected variants (Experimental)

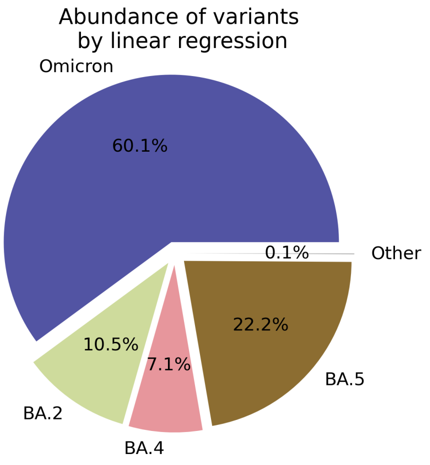


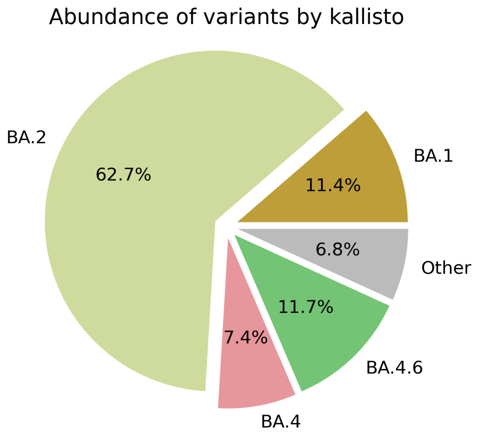


Based on deconvolution, B.1.1.529 is estimated to constitute 60.10% of the viral particles and hence is the most abundant variant in the sample. The $R^2$ for the linear regression was 0.61. Variants that were detected less than 5% were grouped under "Other"

Based on the consensus sequence of the observed reads, the "ensemble-averaged sequence" most closely resembles the BA.2 lineage. If this is a sample consisting of a single source of pathogens or an overwhelming majority of the different sources are infected with the same variant, the sample is dominated by this variant.

Based on mapping individual reads to the variant consensus sequences in the reference database, kallisto predicts that the sample is dominated by BA.2 lineage. Accuracy of this measure is expected to improve if the input data consists of long reads as opposed to convolution.

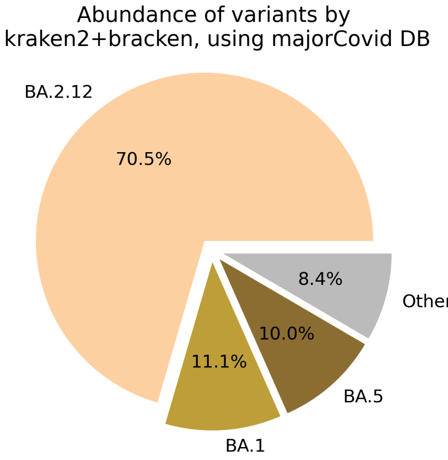


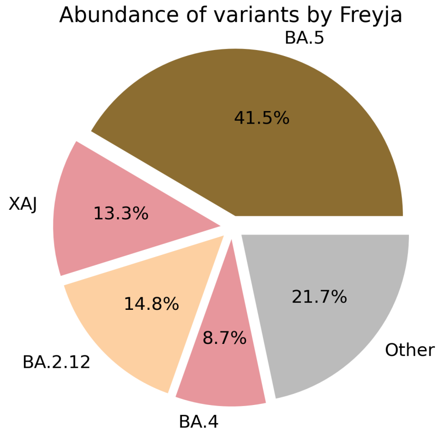

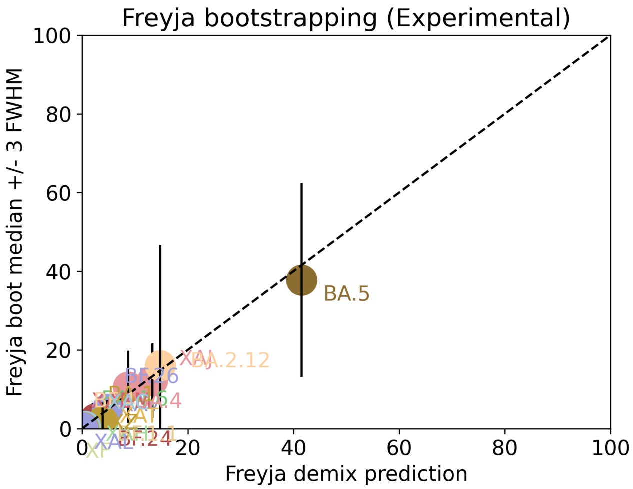


Under the assumption that the presence of a variant requires the detection of all respective mutations of the variant, the characteric mutations which support the presence of the respective variant are indicated in the respective column of the table. Numbers show the number of mutations detected, if any, and the number of mutations expected to be present based on the variant definitions.

| VOC | B.1.617.2 | BA.1 | BA.2 | BA.3 | BA.4 | BA.5 |
|---|---|---|---|---|---|---|
| Characteristic mutations detected | (3 of 13)<br>S:G142D<br>S:L452R<br>S:T478K | (14 of 26)<br>NUC:C15240T<br>NUC:C25000T<br>NUC:C25584T<br>NUC:T13195C<br>NUC:T5386G<br>ORF1AB:A2710T<br>ORF1AB:I3758V<br>ORF1AB:K856R<br>S:A67V<br>S:G446S<br>S:L981F<br>S:N856K<br>S:Q493R<br>S:T95I | (22 of 31)<br>N:S413R<br>NUC:A20055G<br>NUC:C10198T<br>NUC:C12880T<br>NUC:C15714T<br>NUC:C25000T<br>NUC:C25584T<br>NUC:C26858T<br>NUC:C4321T<br>NUC:G10447A<br>ORF1AB:G1307S<br>ORF1AB:L3027F<br>ORF1AB:L3201F<br>ORF1AB:S135R<br>ORF1AB:T3090I<br>ORF1AB:T842I<br>S:D405N<br>S:Q493R<br>S:R408S<br>S:S371F<br>S:T19I<br>S:T376A | (13 of 21)<br>N:S413R<br>NUC:C12880T<br>NUC:C15714T<br>NUC:C26858T<br>NUC:G10447A<br>ORF1AB:G1307S<br>ORF1AB:S135R<br>ORF1AB:T3090I<br>S:A67V<br>S:D405N<br>S:G446S<br>S:Q493R<br>S:S371F | (24 of 31)<br>N:P151S<br>N:S413R<br>NUC:A20055G<br>NUC:C10198T<br>NUC:C12880T<br>NUC:C15714T<br>NUC:C25000T<br>NUC:C25584T<br>NUC:C26858T<br>NUC:C4321T<br>NUC:G10447A<br>NUC:G12160A<br>NUC:G27788T<br>ORF1AB:G1307S<br>ORF1AB:S135R<br>ORF1AB:T3090I<br>ORF1AB:T842I<br>S:D405N<br>S:F486V<br>S:L452R<br>S:S371F<br>S:T19I<br>S:T376A<br>S:V213G | (22 of 28)<br>M:D3N<br>N:S413R<br>NUC:A20055G<br>NUC:C10198T<br>NUC:C12880T<br>NUC:C15714T<br>NUC:C25000T<br>NUC:C25584T<br>NUC:C4321T<br>NUC:G10447A<br>NUC:G12160A<br>ORF1AB:G1307S<br>ORF1AB:S135R<br>ORF1AB:T3090I<br>ORF1AB:T842I<br>S:D405N<br>S:F486V<br>S:L452R<br>S:S371F<br>S:T19I<br>S:T376A<br>S:V213G |

Jaccard Index is a measure of similarity between two sets A and B, reaching the maximum value of 1 if A=B and minimum value of 0 if A ∩ B = {}. In the c(d) representation below, c represents the Jaccard index of the set of mutations that were experimentally detected for this sample as listed above, whereas d refers to the ideal value of the Jaccard index expected from complete genome coverage without any sequencing errors.

| | B.1.617.2 | BA.1 | BA.2 | BA.3 | BA.4 | BA.5 |
|---|---|---|---|---|---|---|
| B.1.617.2 | 1.00 (1.00) | 0.00 (0.00) | 0.00 (0.00) | 0.00 (0.00) | 0.04 (0.02) | 0.04 (0.03) |
| BA.1 | 0.00 (0.00) | 1.00 (1.00) | 0.09 (0.10) | 0.12 (0.21) | 0.06 (0.08) | 0.06 (0.08) |
| BA.2 | 0.00 (0.00) | 0.09 (0.10) | 1.00 (1.00) | 0.46 (0.33) | 0.64 (0.63) | 0.63 (0.59) |
| BA.3 | 0.00 (0.00) | 0.12 (0.21) | 0.46 (0.33) | 1.00 (1.00) | 0.37 (0.30) | 0.35 (0.29) |

| | | | | | | |
|---|---|---|---|---|---|---|
| BA.4 | 0.04 (0.02) | 0.06 (0.08) | 0.64 (0.63) | 0.37 (0.30) | 1.00 (1.00) | 0.84 (0.84) |
| BA.5 | 0.04 (0.03) | 0.06 (0.08) | 0.63 (0.59) | 0.35 (0.29) | 0.84 (0.84) | 1.00 (1.00) |

#### Detected mutations

Excluded from this pdf version due to file size limitations.

CFSAN/OAO
BIOSTATISTICS AND BIOINFORMATICS STAFF

### WASTEWATER SARS-COV2 ANALYSIS REPORT

| | |
|---|---|
| Sample name: | CFSANSMP000119224 |
| Date generated: | 2023-02-01, 20:06:45 EST |
| Timestamp of C-WAP version used: | Tue Jan 31 11:49:22 2023 -0500 |
| Executed by: | Jasmine Amirzadegan |
| Executed on: | 172.20.44.132 (aka n132.raven.cfsan) |

#### Sequencing summary

| | |
|---|---|
| Sequencing chemistry: | Missing with Missing |
| Source site: | Missing (?,?) |
| Sampling date: | Missing |
| Collected by: | Missing |
| Sequenced by: | Missing |
| Total number of reads: | 6440258 |
| Reads aligned: | 2162483 (33%) |
| Average read quality: | 36.6 |
| Average read length: | 148 |
| Reads passing filter: | 2062962 (32%) |
| Average read quality passing filter: | 36.9 |
| Average read length passing filter: | 148 |
| Average coverage passing filter: | 10210X |

A read passes filter if the read length after adaptor trimming ≥ 30 and minimum read quality ≥ 20 within a sliding window of width 4.

#### Overall sequence characteristics

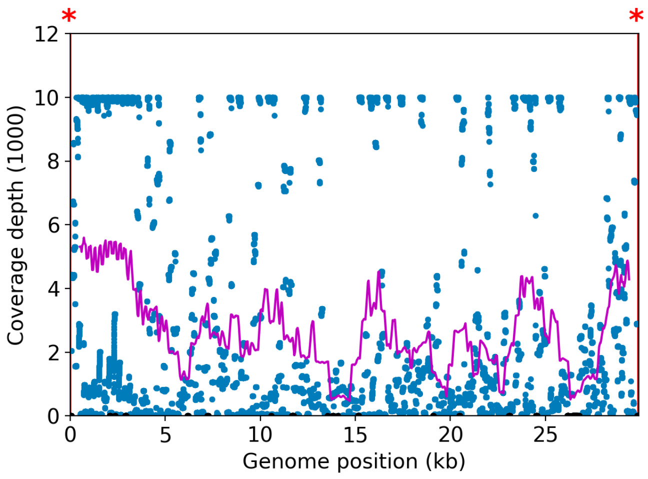


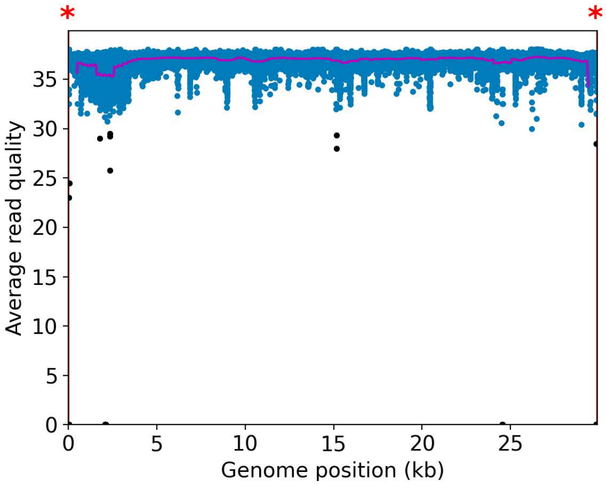


NOTE: The red shaded areas marked with a (*) are not covered by the design of the library preparation kit and hence excluded from analyses. Magenta curves represent moving average with a window width of 1kb.

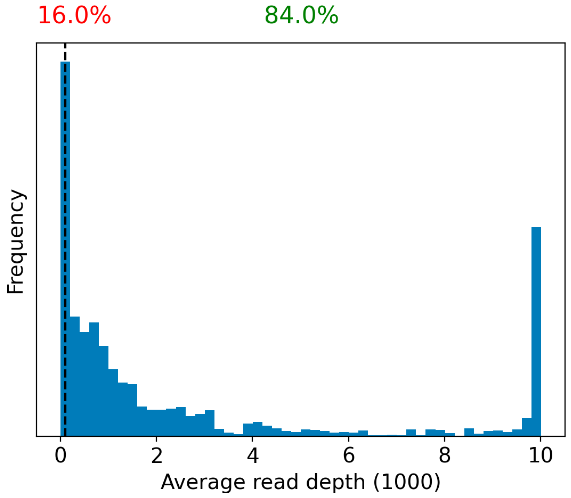


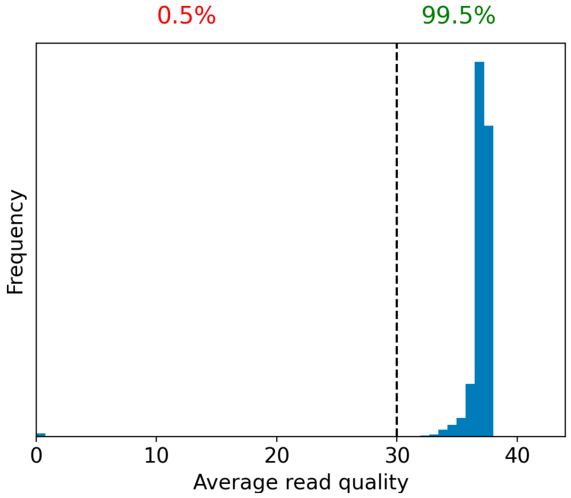


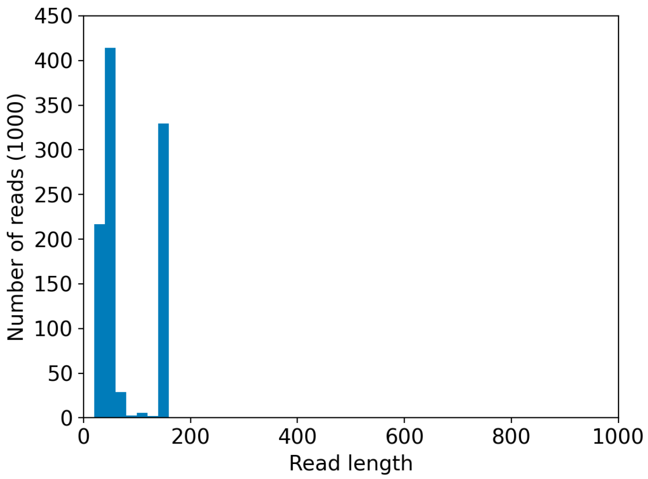


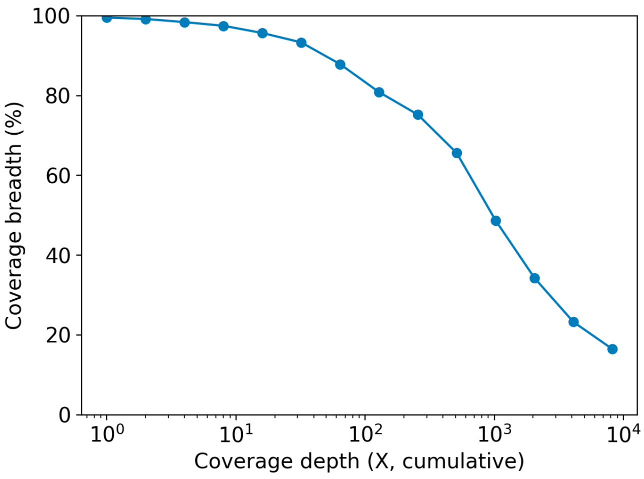

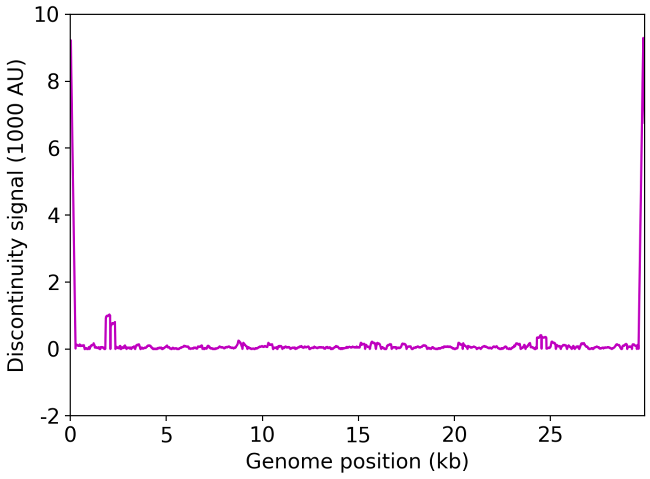


| | Uncovered coordinates (0X) | Poorly covered coordinates (<10X) |
|---|---|---|
| # Inaccessible genomic coordinates by kit design: | 121nt (0%) | 121nt (0%) |
| All genomic coordinates: | 127nt (0%) | 879nt (2%) |
| Common SNPs: | 0nt (0%) | 1nt (2%) |
| Diverse SNPs: | 28nt (12%) | 31nt (13%) |
| Rare SNPs: | 9nt (0%) | 47nt (5%) |

SNPs refer to the polymorphic sites currently in circulation that were detected out of recent GISAID entries. The sites that differ from the SC2 reference sequence are denoted as "common" if [90%, 100%] of the submissions carry this mutation, whereas those that are prevalent in [0%,10%] of the submissions are grouped under the "rare" category. The population is still diverse at the mutation sites that are observed in (10%,90%) of the entries and these coordinates are grouped under the "diverse" category.

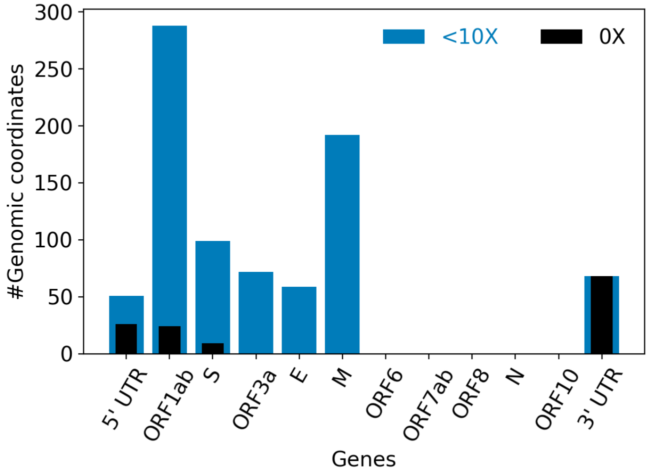


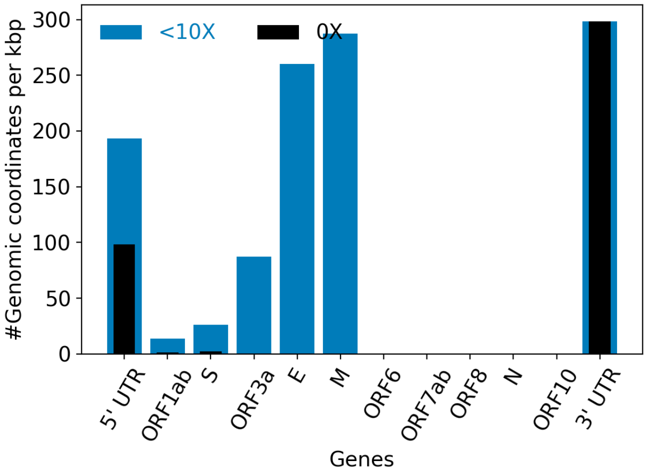


| | |
|---|---|
| Hits to SARS-Cov2 genome (kraken2): | 1342375 reads (41.69%) |
| Hits to human genome (kraken2): | 2944 reads (0.09%) |
| Hits to synthetic sequences (kraken2, taxid 28384): | 134 reads (0.00%) |
| Most abundant organisms (kraken2, family level): | Coronaviridae (41.69%)<br>Staphylococcaceae (0.86%)<br>Flavobacteriaceae (0.11%) |

| | |
|---|---|
| | Hominidae (0.09%)<br>Arcobacteraceae (0.08%) |

#### Detected variants (Experimental)

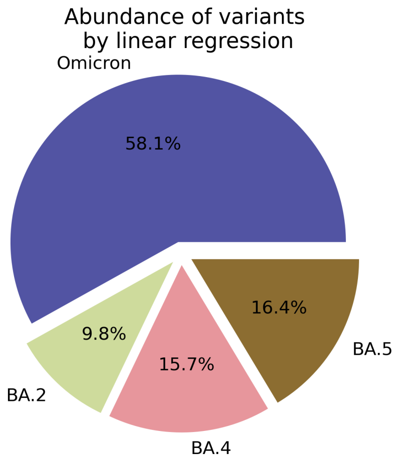


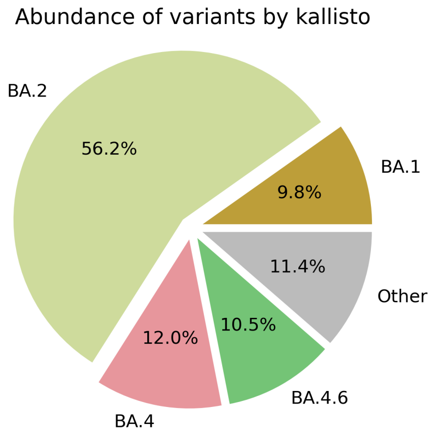


Based on deconvolution, B.1.1.529 is estimated to constitute 58.07% of the viral particles and hence is the most abundant variant in the sample. The $R^2$ for the linear regression was 0.59. Variants that were detected less than 5% were grouped under "Other"

Based on the consensus sequence of the observed reads, the "ensemble-averaged sequence" most closely resembles the BA.2 lineage. If this is a sample consisting of a single source of pathogens or an overwhelming majority of the different sources are infected with the same variant, the sample is dominated by this variant.

Based on mapping individual reads to the variant consensus sequences in the reference database, kallisto predicts that the sample is dominated by BA.2 lineage. Accuracy of this measure is expected to improve if the input data consists of long reads as opposed to convolution.

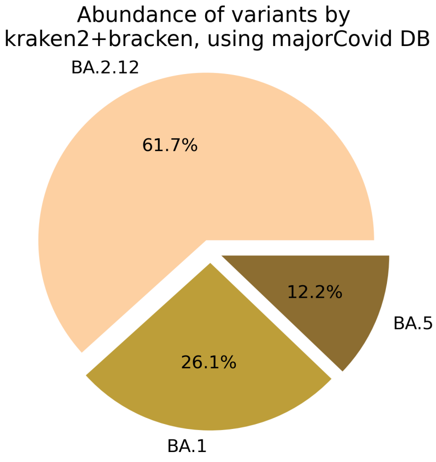


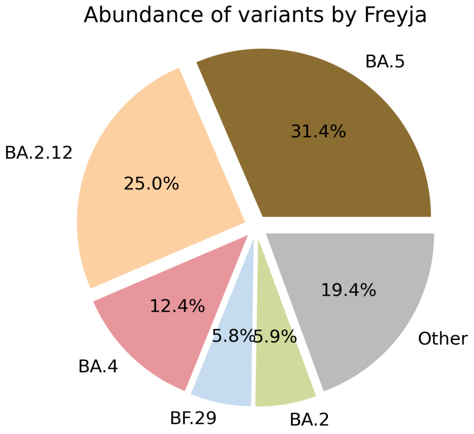

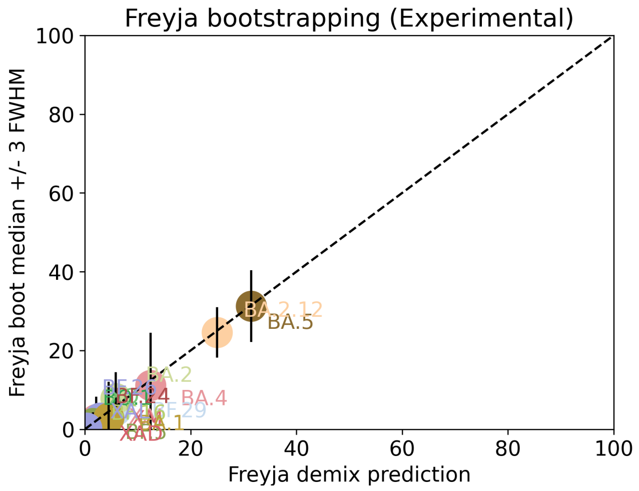


Under the assumption that the presence of a variant requires the detection of all respective mutations of the variant, the characteric mutations which support the presence of the respective variant are indicated in the respective column of the table. Numbers show the number of mutations detected, if any, and the number of mutations expected to be present based on the variant definitions.

| VOC | B.1.617.2 | BA.1 | BA.2 | BA.3 | BA.4 | BA.5 |
|---|---|---|---|---|---|---|
| Characteristic mutations detected | (3 of 13)<br>S:G142D<br>S:L452R<br>S:T478K | (9 of 26)<br>NUC:C15240T<br>NUC:C25000T<br>NUC:C25584T<br>NUC:T13195C<br>ORF1AB:A2710T<br>S:G446S<br>S:L981F<br>S:N856K<br>S:Q493R | (22 of 31)<br>N:S413R<br>NUC:A20055G<br>NUC:C10198T<br>NUC:C12880T<br>NUC:C15714T<br>NUC:C25000T<br>NUC:C25584T<br>NUC:C26858T<br>NUC:C4321T<br>NUC:G10447A<br>ORF1AB:G1307S<br>ORF1AB:L3027F<br>ORF1AB:L3201F<br>ORF1AB:S135R<br>ORF1AB:T3090I<br>ORF1AB:T842I<br>S:D405N<br>S:Q493R<br>S:R408S<br>S:S371F<br>S:T19I<br>S:T376A | (12 of 21)<br>N:S413R<br>NUC:C12880T<br>NUC:C15714T<br>NUC:C26858T<br>NUC:G10447A<br>ORF1AB:G1307S<br>ORF1AB:S135R<br>ORF1AB:T3090I<br>S:D405N<br>S:G446S<br>S:Q493R<br>S:S371F | (24 of 31)<br>N:P151S<br>N:S413R<br>NUC:A20055G<br>NUC:C10198T<br>NUC:C12880T<br>NUC:C15714T<br>NUC:C25000T<br>NUC:C25584T<br>NUC:C26858T<br>NUC:C4321T<br>NUC:G10447A<br>NUC:G12160A<br>NUC:G27788T<br>ORF1AB:G1307S<br>ORF1AB:S135R<br>ORF1AB:T3090I<br>ORF1AB:T842I<br>S:D405N<br>S:F486V<br>S:L452R<br>S:S371F<br>S:T19I<br>S:T376A<br>S:V213G | (21 of 28)<br>N:S413R<br>NUC:A20055G<br>NUC:C10198T<br>NUC:C12880T<br>NUC:C15714T<br>NUC:C25000T<br>NUC:C25584T<br>NUC:C4321T<br>NUC:G10447A<br>NUC:G12160A<br>ORF1AB:G1307S<br>ORF1AB:S135R<br>ORF1AB:T3090I<br>ORF1AB:T842I<br>S:D405N<br>S:F486V<br>S:L452R<br>S:S371F<br>S:T19I<br>S:T376A<br>S:V213G |

Jaccard Index is a measure of similarity between two sets A and B, reaching the maximum value of 1 if A=B and minimum value of 0 if A ∩ B = {}. In the c(d) representation below, c represents the Jaccard index of the set of mutations that were experimentally detected for this sample as listed above, whereas d refers to the ideal value of the Jaccard index expected from complete genome coverage without any sequencing errors.

| | B.1.617.2 | BA.1 | BA.2 | BA.3 | BA.4 | BA.5 |
|---|---|---|---|---|---|---|
| B.1.617.2 | 1.00 (1.00) | 0.00 (0.00) | 0.00 (0.00) | 0.00 (0.00) | 0.04 (0.02) | 0.04 (0.03) |
| BA.1 | 0.00 (0.00) | 1.00 (1.00) | 0.11 (0.10) | 0.11 (0.21) | 0.06 (0.08) | 0.07 (0.08) |
| BA.2 | 0.00 (0.00) | 0.11 (0.10) | 1.00 (1.00) | 0.48 (0.33) | 0.64 (0.63) | 0.65 (0.59) |
| BA.3 | 0.00 (0.00) | 0.11 (0.21) | 0.48 (0.33) | 1.00 (1.00) | 0.38 (0.30) | 0.38 (0.29) |

| | | | | | | |
|---|---|---|---|---|---|---|
| BA.4 | 0.04 (0.02) | 0.06 (0.08) | 0.64 (0.63) | 0.38 (0.30) | 1.00 (1.00) | 0.88 (0.84) |
| BA.5 | 0.04 (0.03) | 0.07 (0.08) | 0.65 (0.59) | 0.38 (0.29) | 0.88 (0.84) | 1.00 (1.00) |

#### Detected mutations

Excluded from this pdf version due to file size limitations.

CFSAN/OAO
BIOSTATISTICS AND BIOINFORMATICS STAFF

### WASTEWATER SARS-COV2 ANALYSIS REPORT

| | |
|---|---|
| Sample name: | CFSANSMP000119624 |
| Date generated: | 2023-02-01, 20:14:45 EST |
| Timestamp of C-WAP version used: | Tue Jan 31 11:49:22 2023 -0500 |
| Executed by: | Jasmine Amirzadegan |
| Executed on: | 172.20.44.108 (aka n108.raven.cfsan) |

#### Sequencing summary

| | |
|---|---|
| Sequencing chemistry: | Missing with Missing |
| Source site: | Missing (?,?) |
| Sampling date: | Missing |
| Collected by: | Missing |
| Sequenced by: | Missing |
| Total number of reads: | 3821038 |
| Reads aligned: | 3447227 (90%) |
| Average read quality: | 36.6 |
| Average read length: | 149 |
| Reads passing filter: | 3303434 (86%) |
| Average read quality passing filter: | 36.9 |
| Average read length passing filter: | 149 |
| Average coverage passing filter: | 16460X |

A read passes filter if the read length after adaptor trimming ≥ 30 and minimum read quality ≥ 20 within a sliding window of width 4.

#### Overall sequence characteristics

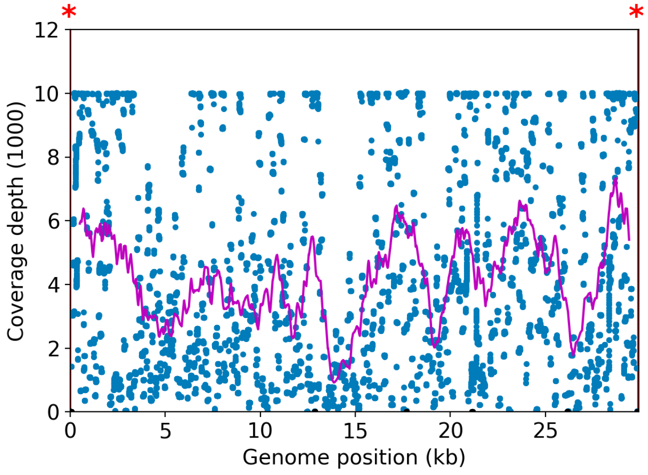


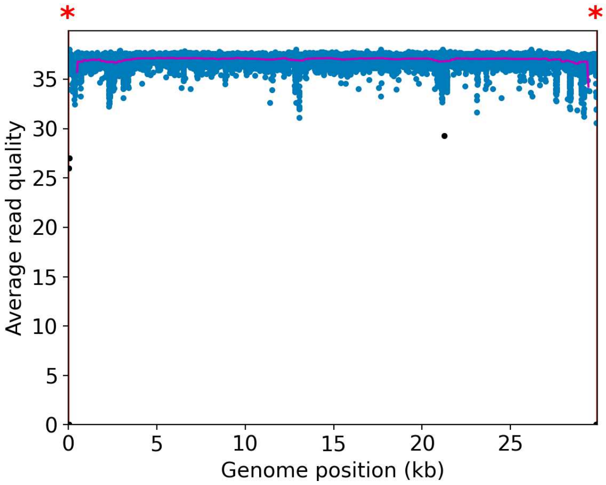


NOTE: The red shaded areas marked with a (*) are not covered by the design of the library preparation kit and hence excluded from analyses. Magenta curves represent moving average with a window width of 1kb.

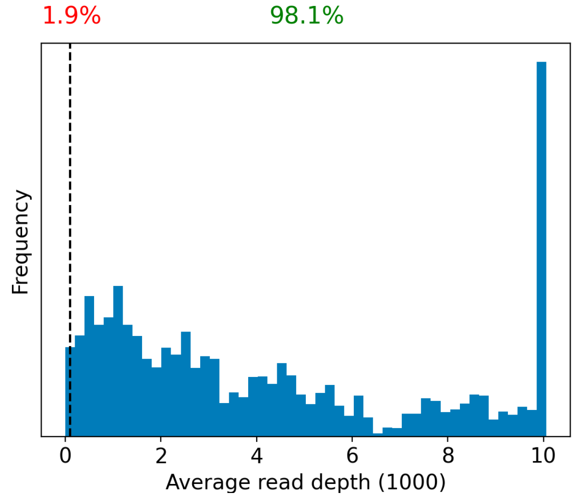


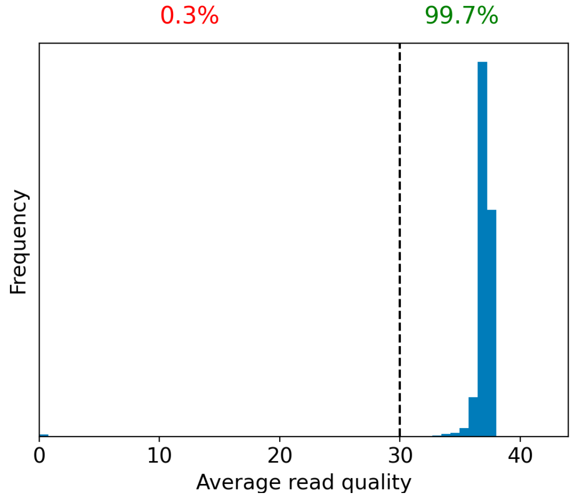


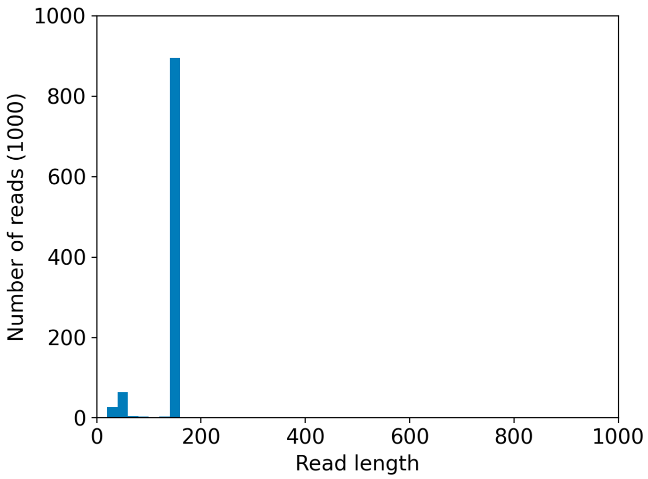


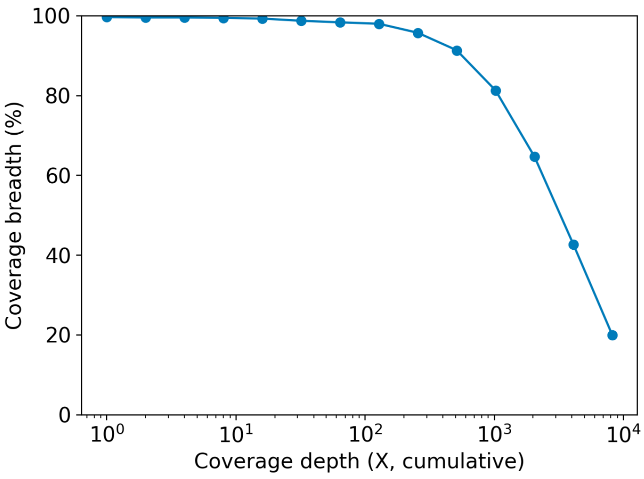

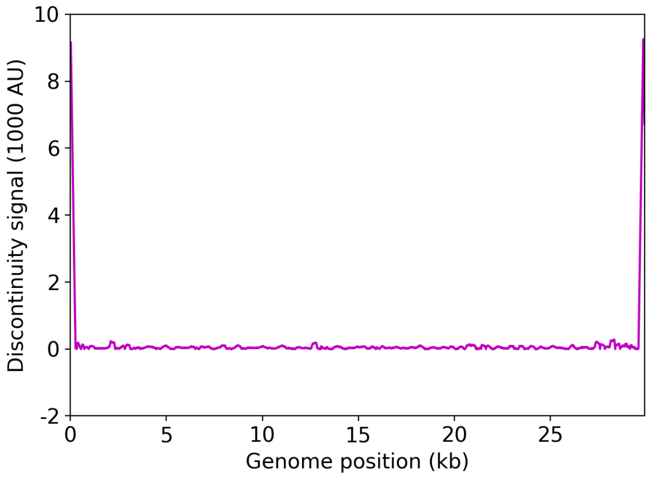


| | Uncovered coordinates (0X) | Poorly covered coordinates (<10X) |
|---|---|---|
| # Inaccessible genomic coordinates by kit design: | 121nt (0%) | 121nt (0%) |
| All genomic coordinates: | 94nt (0%) | 174nt (0%) |
| Common SNPs: | 0nt (0%) | 0nt (0%) |
| Diverse SNPs: | 28nt (12%) | 29nt (12%) |
| Rare SNPs: | 9nt (0%) | 10nt (1%) |

SNPs refer to the polymorphic sites currently in circulation that were detected out of recent GISAID entries. The sites that differ from the SC2 reference sequence are denoted as "common" if [90%, 100%] of the submissions carry this mutation, whereas those that are prevalent in [0%,10%] of the submissions are grouped under the "rare" category. The population is still diverse at the mutation sites that are observed in (10%,90%) of the entries and these coordinates are grouped under the "diverse" category.

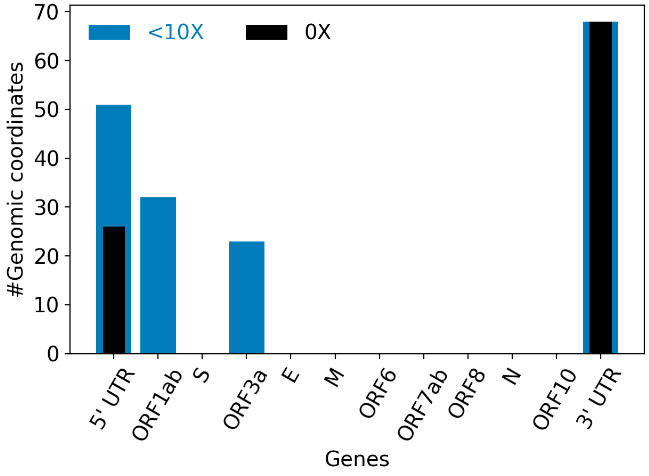


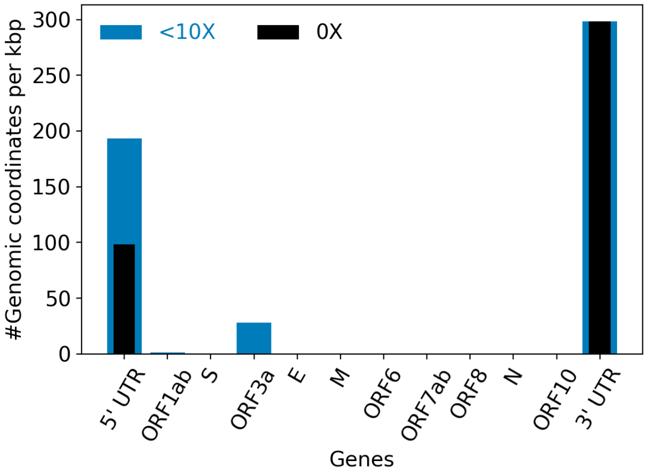


| | |
|---|---|
| Hits to SARS-Cov2 genome (kraken2): | 1746934 reads (91.44%) |
| Hits to human genome (kraken2): | 218 reads (0.01%) |
| Hits to synthetic sequences (kraken2, taxid 28384): | 1496 reads (0.08%) |
| Most abundant organisms (kraken2, family level): | Coronaviridae (91.44%)<br>Enterococcaceae (0.14%)<br>Halomonadaceae (0.03%) |

| | |
|---|---|
| | Arcobacteraceae (0.02%)<br>Bacteroidaceae (0.02%) |

### Detected variants (Experimental)

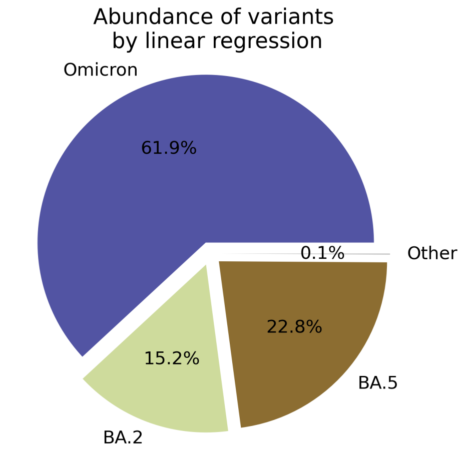


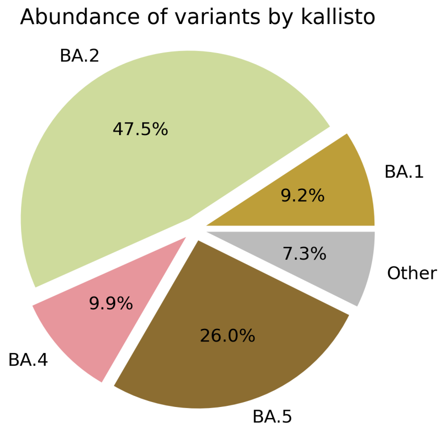


Based on deconvolution, B.1.1.529 is estimated to constitute 61.91% of the viral particles and hence is the most abundant variant in the sample. The $R^2$ for the linear regression was 0.62. Variants that were detected less than 5% were grouped under "Other"

Based on the consensus sequence of the observed reads, the "ensemble-averaged sequence" most closely resembles the BA.2 lineage. If this is a sample consisting of a single source of pathogens or an overwhelming majority of the different sources are infected with the same variant, the sample is dominated by this variant.

Based on mapping individual reads to the variant consensus sequences in the reference database, kallisto predicts that the sample is dominated by BA.2 lineage. Accuracy of this measure is expected to improve if the input data consists of long reads as opposed to convolution.

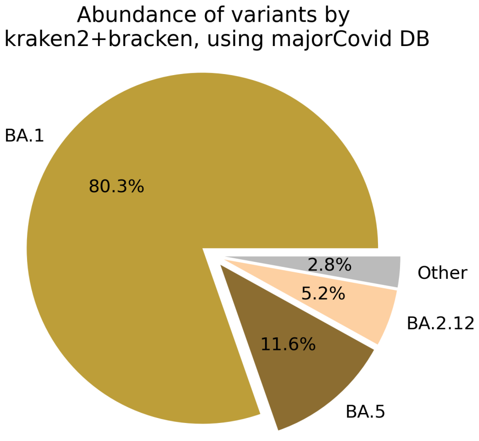


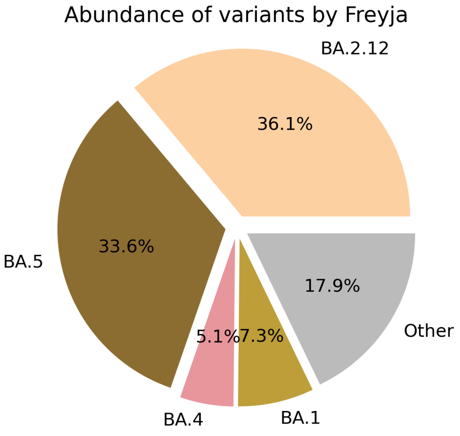

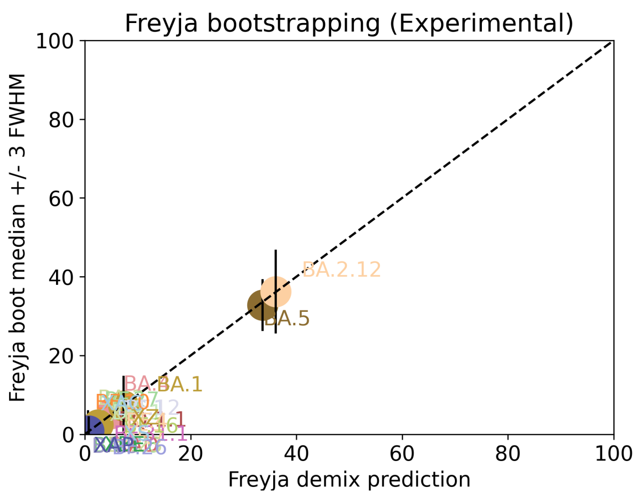


Under the assumption that the presence of a variant requires the detection of all respective mutations of the variant, the characteric mutations which support the presence of the respective variant are indicated in the respective column of the table. Numbers show the number of mutations detected, if any, and the number of mutations expected to be present based on the variant definitions.

| VOC | B.1.617.2 | BA.1 | BA.2 | BA.3 | BA.4 | BA.5 |
|---|---|---|---|---|---|---|
| Characteristic mutations detected | (3 of 13)<br>S:G142D<br>S:L452R<br>S:T478K | (12 of 26)<br>NUC:C15240T<br>NUC:C25000T<br>NUC:C25584T<br>NUC:T13195C<br>NUC:T5386G<br>ORF1AB:I3758V<br>S:G446S<br>S:G496S<br>S:L981F<br>S:N856K<br>S:Q493R<br>S:T95I | (23 of 31)<br>N:S413R<br>NUC:A20055G<br>NUC:A9424G<br>NUC:C10198T<br>NUC:C12880T<br>NUC:C15714T<br>NUC:C25000T<br>NUC:C25584T<br>NUC:C26858T<br>NUC:C4321T<br>NUC:G10447A<br>ORF1AB:G1307S<br>ORF1AB:L3027F<br>ORF1AB:L3201F<br>ORF1AB:S135R<br>ORF1AB:T3090I<br>ORF1AB:T842I<br>S:D405N<br>S:Q493R<br>S:R408S<br>S:S371F<br>S:T19I<br>S:T376A | (12 of 21)<br>N:S413R<br>NUC:C12880T<br>NUC:C15714T<br>NUC:C26858T<br>NUC:G10447A<br>ORF1AB:G1307S<br>ORF1AB:S135R<br>ORF1AB:T3090I<br>S:D405N<br>S:G446S<br>S:Q493R<br>S:S371F | (24 of 31)<br>N:P151S<br>N:S413R<br>NUC:A20055G<br>NUC:C10198T<br>NUC:C12880T<br>NUC:C15714T<br>NUC:C25000T<br>NUC:C25584T<br>NUC:C26858T<br>NUC:C4321T<br>NUC:G10447A<br>NUC:G12160A<br>NUC:G27788T<br>ORF1AB:G1307S<br>ORF1AB:S135R<br>ORF1AB:T3090I<br>ORF1AB:T842I<br>S:D405N<br>S:F486V<br>S:L452R<br>S:S371F<br>S:T19I<br>S:T376A<br>S:V213G | (22 of 28)<br>M:D3N<br>N:S413R<br>NUC:A20055G<br>NUC:C10198T<br>NUC:C12880T<br>NUC:C15714T<br>NUC:C25000T<br>NUC:C25584T<br>NUC:C4321T<br>NUC:G10447A<br>NUC:G12160A<br>ORF1AB:G1307S<br>ORF1AB:S135R<br>ORF1AB:T3090I<br>ORF1AB:T842I<br>S:D405N<br>S:F486V<br>S:L452R<br>S:S371F<br>S:T19I<br>S:T376A<br>S:V213G |

Jaccard Index is a measure of similarity between two sets A and B, reaching the maximum value of 1 if A=B and minimum value of 0 if A ∩ B = {}. In the c(d) representation below, c represents the Jaccard index of the set of mutations that were experimentally detected for this sample as listed above, whereas d refers to the ideal value of the Jaccard index expected from complete genome coverage without any sequencing errors.

| | B.1.617.2 | BA.1 | BA.2 | BA.3 | BA.4 | BA.5 |
|---|---|---|---|---|---|---|
| B.1.617.2 | 1.00 (1.00) | 0.00 (0.00) | 0.00 (0.00) | 0.00 (0.00) | 0.04 (0.02) | 0.04 (0.03) |
| BA.1 | 0.00 (0.00) | 1.00 (1.00) | 0.09 (0.10) | 0.09 (0.21) | 0.06 (0.08) | 0.06 (0.08) |
| BA.2 | 0.00 (0.00) | 0.09 (0.10) | 1.00 (1.00) | 0.46 (0.33) | 0.62 (0.63) | 0.61 (0.59) |
| BA.3 | 0.00 (0.00) | 0.09 (0.21) | 0.46 (0.33) | 1.00 (1.00) | 0.38 (0.30) | 0.36 (0.29) |

| | | | | | | |
|---|---|---|---|---|---|---|
| BA.4 | 0.04 (0.02) | 0.06 (0.08) | 0.62 (0.63) | 0.38 (0.30) | 1.00 (1.00) | 0.84 (0.84) |
| BA.5 | 0.04 (0.03) | 0.06 (0.08) | 0.61 (0.59) | 0.36 (0.29) | 0.84 (0.84) | 1.00 (1.00) |

#### Detected mutations

Excluded from this pdf version due to file size limitations.

U.S. FOOD & DRUG ADMINISTRATION

CFSAN/OAO
BIOSTATISTICS AND BIOINFORMATICS STAFF

### WASTEWATER SARS-COV2 ANALYSIS REPORT

| | |
|---|---|
| Sample name: | CFSANSMP000119626 |
| Date generated: | 2023-02-01, 20:07:25 EST |
| Timestamp of C-WAP version used: | Tue Jan 31 11:49:22 2023 -0500 |
| Executed by: | Jasmine Amirzadegan |
| Executed on: | 172.20.44.127 (aka n127.raven.cfsan) |

#### Sequencing summary

| | |
|---|---|
| Sequencing chemistry: | Missing with Missing |
| Source site: | Missing (?,?) |
| Sampling date: | Missing |
| Collected by: | Missing |
| Sequenced by: | Missing |
| Total number of reads: | 3761430 |
| Reads aligned: | 1935752 (51%) |
| Average read quality: | 36.6 |
| Average read length: | 148 |
| Reads passing filter: | 1843202 (49%) |
| Average read quality passing filter: | 36.9 |
| Average read length passing filter: | 149 |
| Average coverage passing filter: | 9184X |

A read passes filter if the read length after adaptor trimming ≥ 30 and minimum read quality ≥ 20 within a sliding window of width 4.

#### Overall sequence characteristics

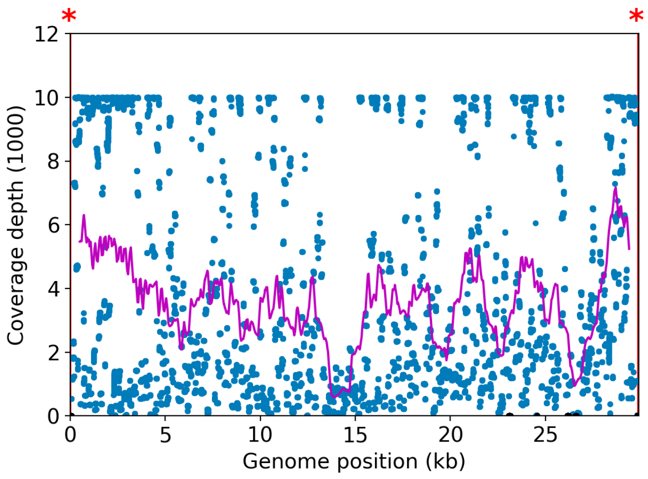


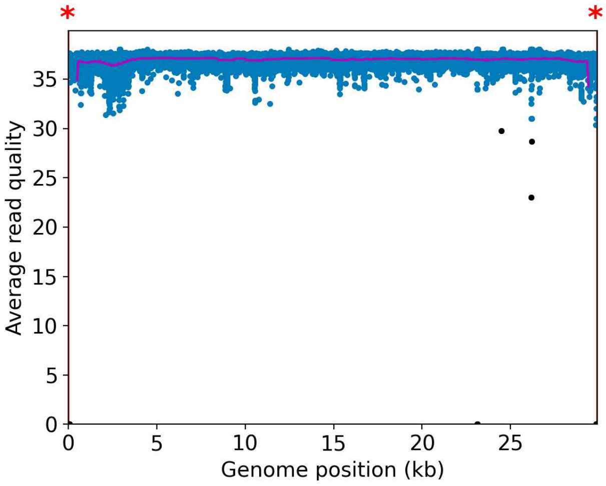


NOTE: The red shaded areas marked with a (*) are not covered by the design of the library preparation kit and hence excluded from analyses. Magenta curves represent moving average with a window width of 1kb.

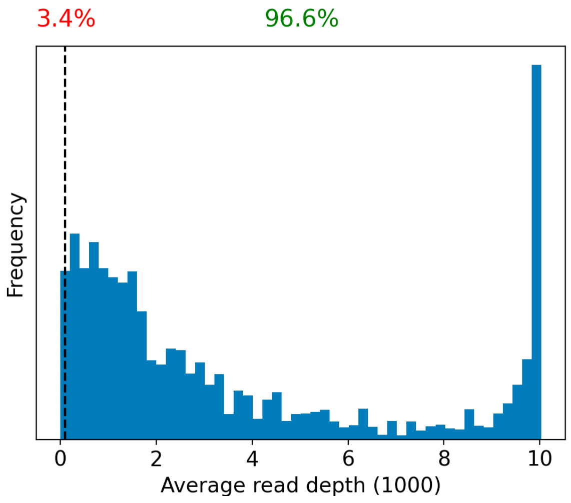


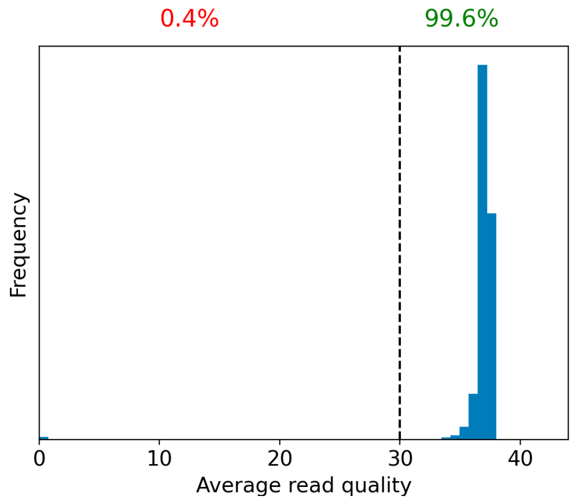


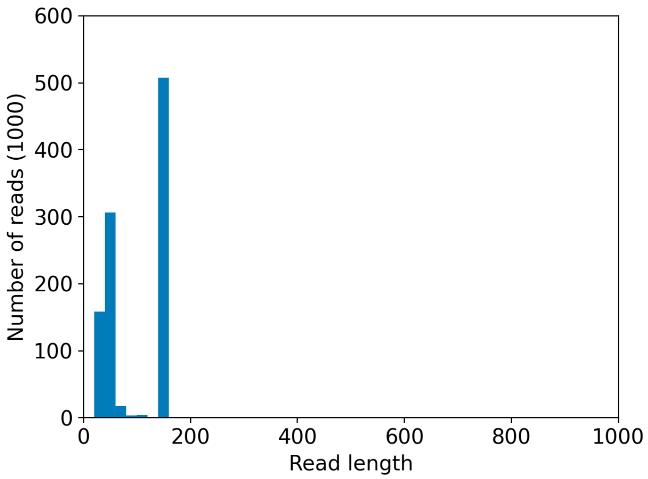


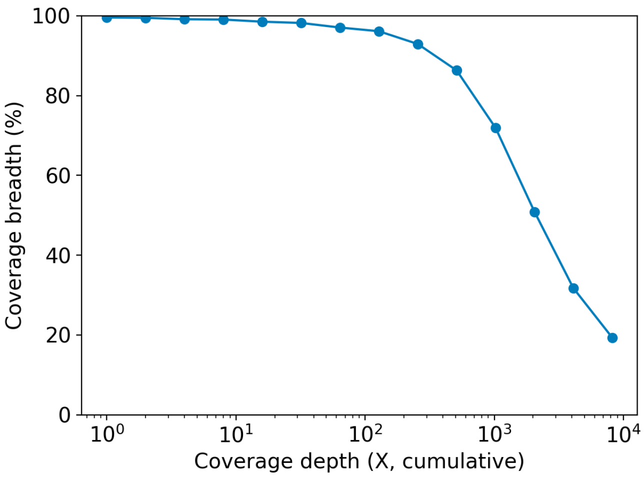

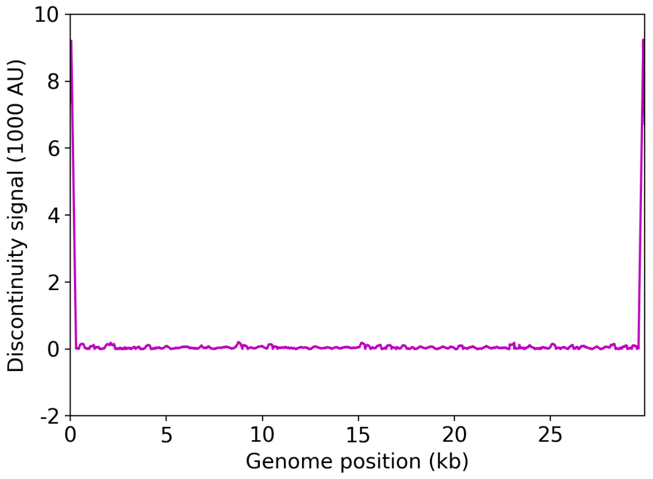


| | Uncovered coordinates (0X) | Poorly covered coordinates (<10X) |
|---|---|---|
| # Inaccessible genomic coordinates by kit design: | 121nt (0%) | 121nt (0%) |
| All genomic coordinates: | 122nt (0%) | 277nt (0%) |
| Common SNPs: | 0nt (0%) | 0nt (0%) |
| Diverse SNPs: | 29nt (12%) | 29nt (12%) |
| Rare SNPs: | 10nt (1%) | 46nt (4%) |

SNPs refer to the polymorphic sites currently in circulation that were detected out of recent GISAID entries. The sites that differ from the SC2 reference sequence are denoted as "common" if [90%, 100%] of the submissions carry this mutation, whereas those that are prevalent in [0%,10%] of the submissions are grouped under the "rare" category. The population is still diverse at the mutation sites that are observed in (10%,90%) of the entries and these coordinates are grouped under the "diverse" category.

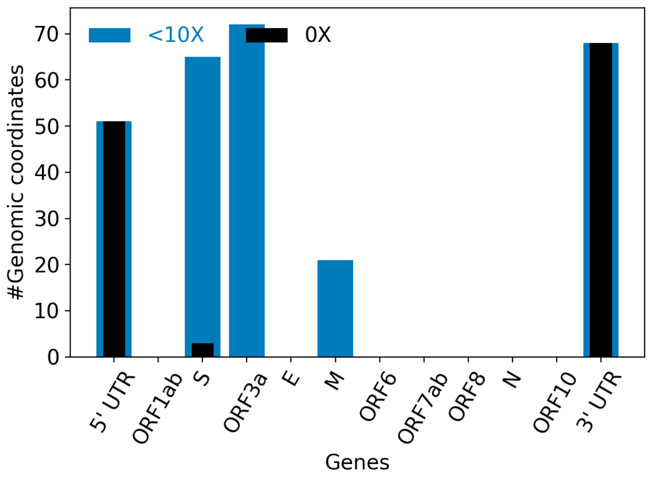


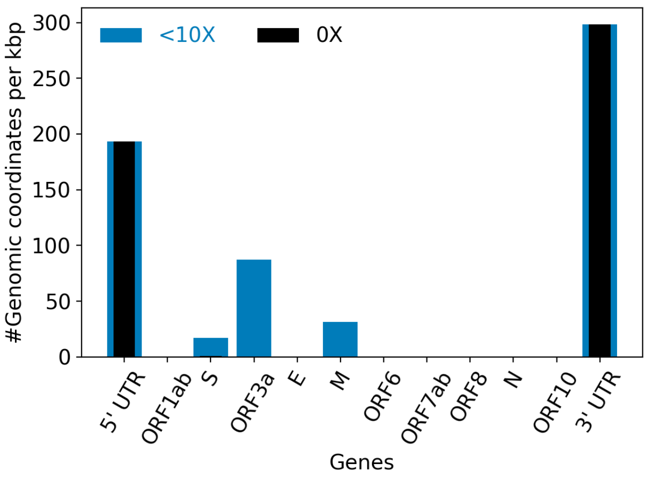


| | |
|---|---|
| Hits to SARS-Cov2 genome (kraken2): | 1124635 reads (59.80%) |
| Hits to human genome (kraken2): | 1126 reads (0.06%) |
| Hits to synthetic sequences (kraken2, taxid 28384): | 176 reads (0.01%) |
| Most abundant organisms (kraken2, family level): | Coronaviridae (59.80%)<br>Pseudomonadaceae (0.28%)<br>Enterobacteriaceae (0.17%) |

| | |
|---|---|
| | Hominidae (0.06%)<br>Morganellaceae (0.06%) |

### Detected variants (Experimental)

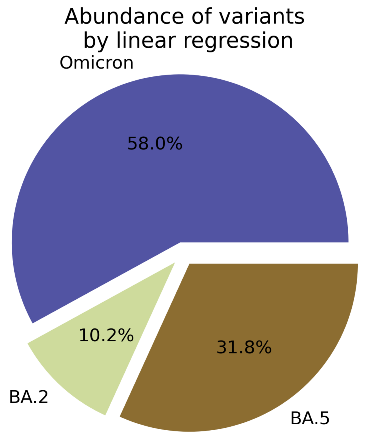


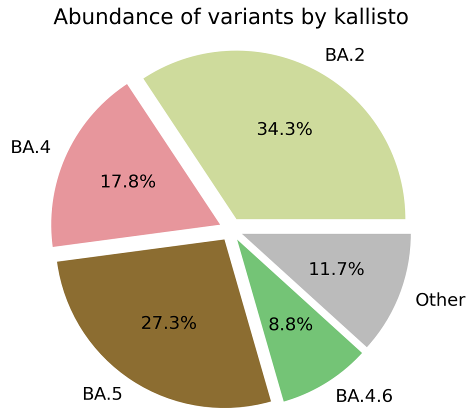


Based on deconvolution, B.1.1.529 is estimated to constitute 58.00% of the viral particles and hence is the most abundant variant in the sample. The $R^2$ for the linear regression was 0.60. Variants that were detected less than 5% were grouped under "Other"

Based on the consensus sequence of the observed reads, the "ensemble-averaged sequence" most closely resembles the BA.2 lineage. If this is a sample consisting of a single source of pathogens or an overwhelming majority of the different sources are infected with the same variant, the sample is dominated by this variant.

Based on mapping individual reads to the variant consensus sequences in the reference database, kallisto predicts that the sample is dominated by BA.2 lineage. Accuracy of this measure is expected to improve if the input data consists of long reads as opposed to convolution.

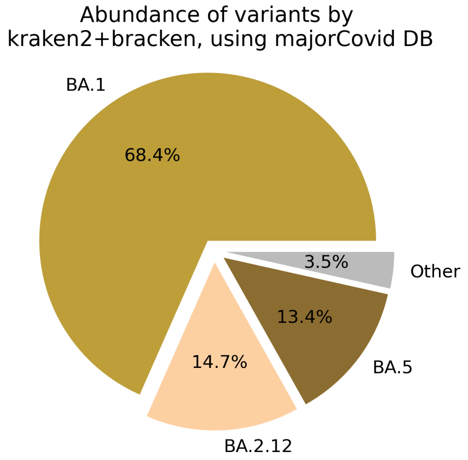


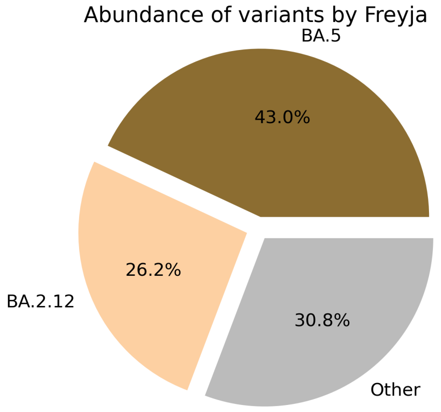

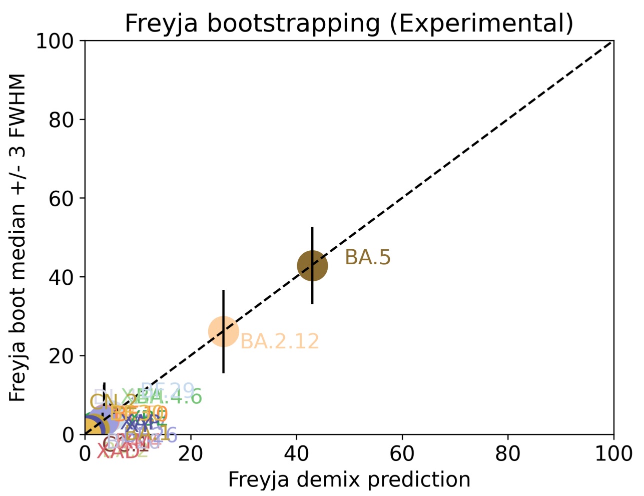


Under the assumption that the presence of a variant requires the detection of all respective mutations of the variant, the characteric mutations which support the presence of the respective variant are indicated in the respective column of the table. Numbers show the number of mutations detected, if any, and the number of mutations expected to be present based on the variant definitions.

| VOC | B.1.617.2 | BA.1 | BA.2 | BA.3 | BA.4 | BA.5 |
|---|---|---|---|---|---|---|
| Characteristic mutations detected | (4 of 13)<br>N:D377Y<br>S:G142D<br>S:L452R<br>S:T478K | (9 of 26)<br>NUC:C15240T<br>NUC:C25000T<br>NUC:C25584T<br>NUC:T13195C<br>S:G446S<br>S:G496S<br>S:L981F<br>S:N856K<br>S:Q493R | (22 of 31)<br>N:S413R<br>NUC:A20055G<br>NUC:C10198T<br>NUC:C12880T<br>NUC:C15714T<br>NUC:C25000T<br>NUC:C25584T<br>NUC:C26858T<br>NUC:C4321T<br>NUC:G10447A<br>ORF1AB:G1307S<br>ORF1AB:L3027F<br>ORF1AB:L3201F<br>ORF1AB:S135R<br>ORF1AB:T3090I<br>ORF1AB:T842I<br>S:D405N<br>S:Q493R<br>S:R408S<br>S:S371F<br>S:T19I<br>S:T376A | (12 of 21)<br>N:S413R<br>NUC:C12880T<br>NUC:C15714T<br>NUC:C26858T<br>NUC:G10447A<br>ORF1AB:G1307S<br>ORF1AB:S135R<br>ORF1AB:T3090I<br>S:D405N<br>S:G446S<br>S:Q493R<br>S:S371F | (24 of 31)<br>N:P151S<br>N:S413R<br>NUC:A20055G<br>NUC:C10198T<br>NUC:C12880T<br>NUC:C15714T<br>NUC:C25000T<br>NUC:C25584T<br>NUC:C26858T<br>NUC:C4321T<br>NUC:G10447A<br>NUC:G12160A<br>NUC:G27788T<br>ORF1AB:G1307S<br>ORF1AB:S135R<br>ORF1AB:T3090I<br>ORF1AB:T842I<br>S:D405N<br>S:F486V<br>S:L452R<br>S:S371F<br>S:T19I<br>S:T376A<br>S:V213G | (22 of 28)<br>M:D3N<br>N:S413R<br>NUC:A20055G<br>NUC:C10198T<br>NUC:C12880T<br>NUC:C15714T<br>NUC:C25000T<br>NUC:C25584T<br>NUC:C4321T<br>NUC:G10447A<br>NUC:G12160A<br>ORF1AB:G1307S<br>ORF1AB:S135R<br>ORF1AB:T3090I<br>ORF1AB:T842I<br>S:D405N<br>S:F486V<br>S:L452R<br>S:S371F<br>S:T19I<br>S:T376A<br>S:V213G |

Jaccard Index is a measure of similarity between two sets A and B, reaching the maximum value of 1 if A=B and minimum value of 0 if A ∩ B = {}. In the c(d) representation below, c represents the Jaccard index of the set of mutations that were experimentally detected for this sample as listed above, whereas d refers to the ideal value of the Jaccard index expected from complete genome coverage without any sequencing errors.

| | B.1.617.2 | BA.1 | BA.2 | BA.3 | BA.4 | BA.5 |
|---|---|---|---|---|---|---|
| B.1.617.2 | 1.00 (1.00) | 0.00 (0.00) | 0.00 (0.00) | 0.00 (0.00) | 0.04 (0.02) | 0.04 (0.03) |
| BA.1 | 0.00 (0.00) | 1.00 (1.00) | 0.11 (0.10) | 0.11 (0.21) | 0.06 (0.08) | 0.07 (0.08) |
| BA.2 | 0.00 (0.00) | 0.11 (0.10) | 1.00 (1.00) | 0.48 (0.33) | 0.64 (0.63) | 0.63 (0.59) |
| BA.3 | 0.00 (0.00) | 0.11 (0.21) | 0.48 (0.33) | 1.00 (1.00) | 0.38 (0.30) | 0.36 (0.29) |

| | | | | | | |
|---|---|---|---|---|---|---|
| BA.4 | 0.04 (0.02) | 0.06 (0.08) | 0.64 (0.63) | 0.38 (0.30) | 1.00 (1.00) | 0.84 (0.84) |
| BA.5 | 0.04 (0.03) | 0.07 (0.08) | 0.63 (0.59) | 0.36 (0.29) | 0.84 (0.84) | 1.00 (1.00) |

#### Detected mutations

Excluded from this pdf version due to file size limitations.

CFSAN/OAO
BIOSTATISTICS AND BIOINFORMATICS STAFF

### WASTEWATER SARS-COV2 ANALYSIS REPORT

| | |
|---|---|
| Sample name: | CFSANSMP000119627 |
| Date generated: | 2023-02-01, 20:05:40 EST |
| Timestamp of C-WAP version used: | Tue Jan 31 11:49:22 2023 -0500 |
| Executed by: | Jasmine Amirzadegan |
| Executed on: | 172.20.44.127 (aka n127.raven.cfsan) |

#### Sequencing summary

| | |
|---|---|
| Sequencing chemistry: | Missing with Missing |
| Source site: | Missing (?,?) |
| Sampling date: | Missing |
| Collected by: | Missing |
| Sequenced by: | Missing |
| Total number of reads: | 3257848 |
| Reads aligned: | 1842055 (56%) |
| Average read quality: | 36.6 |
| Average read length: | 149 |
| Reads passing filter: | 1756675 (53%) |
| Average read quality passing filter: | 36.9 |
| Average read length passing filter: | 149 |
| Average coverage passing filter: | 8753X |

A read passes filter if the read length after adaptor trimming ≥ 30 and minimum read quality ≥ 20 within a sliding window of width 4.

#### Overall sequence characteristics

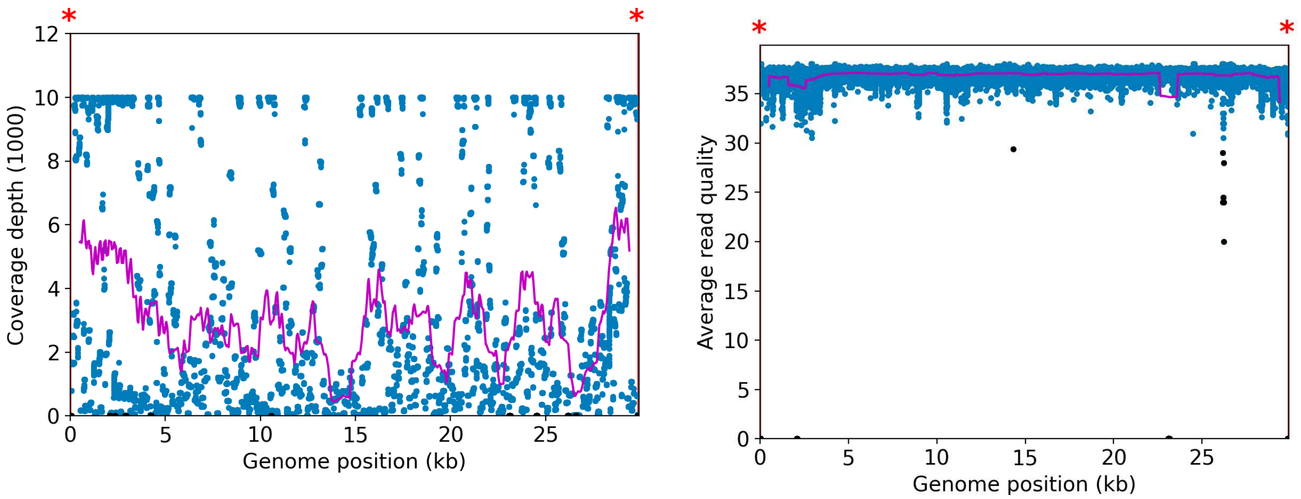


NOTE: The red shaded areas marked with a (*) are not covered by the design of the library preparation kit and hence excluded from analyses. Magenta curves represent moving average with a window width of 1kb.

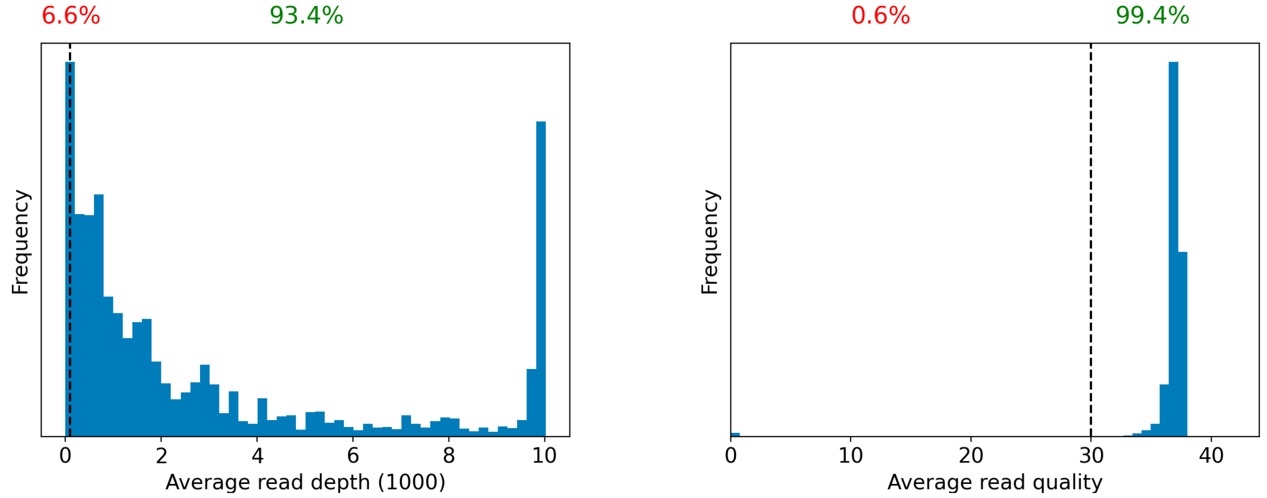


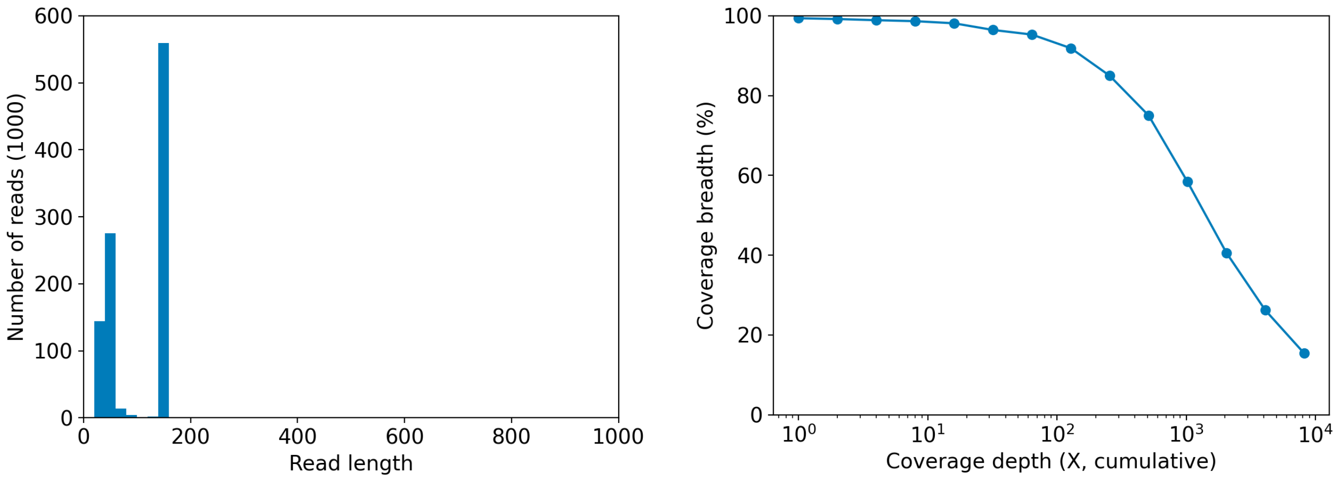

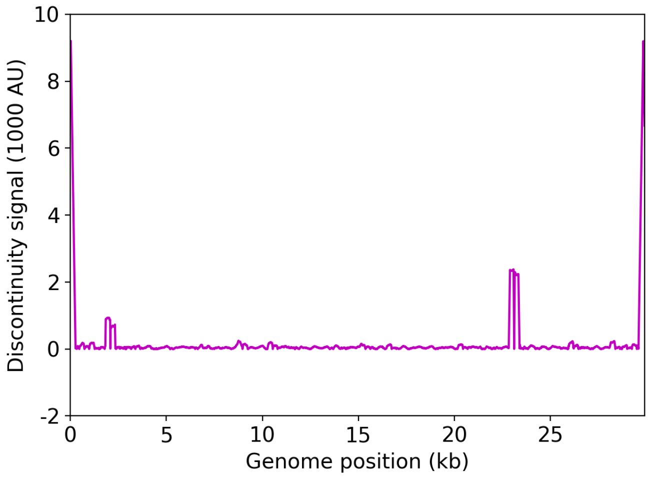


| | Uncovered coordinates (0X) | Poorly covered coordinates (<10X) |
|---|---|---|
| # Inaccessible genomic coordinates by kit design: | 121nt (0%) | 121nt (0%) |
| All genomic coordinates: | 178nt (0%) | 464nt (1%) |
| Common SNPs: | 0nt (0%) | 0nt (0%) |
| Diverse SNPs: | 28nt (12%) | 29nt (12%) |
| Rare SNPs: | 45nt (4%) | 46nt (4%) |

SNPs refer to the polymorphic sites currently in circulation that were detected out of recent GISAID entries. The sites that differ from the SC2 reference sequence are denoted as "common" if [90%, 100%] of the submissions carry this mutation, whereas those that are prevalent in [0%,10%] of the submissions are grouped under the "rare" category. The population is still diverse at the mutation sites that are observed in (10%,90%) of the entries and these coordinates are grouped under the "diverse" category.

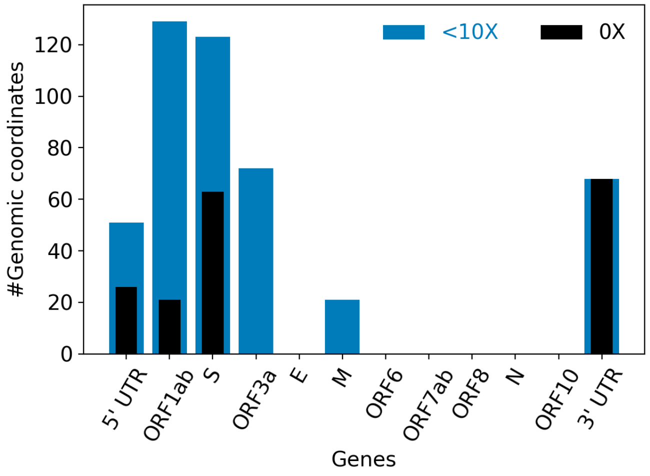


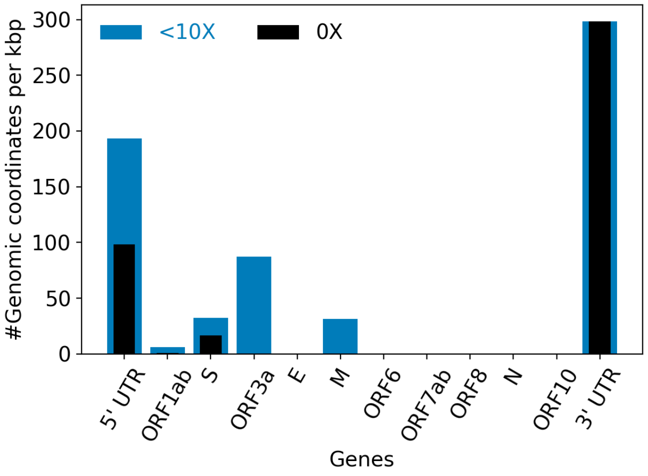


| | |
|---|---|
| Hits to SARS-Cov2 genome (kraken2): | 1024906 reads (62.92%) |
| Hits to human genome (kraken2): | 1328 reads (0.08%) |
| Hits to synthetic sequences (kraken2, taxid 28384): | 416 reads (0.03%) |
| Most abundant organisms (kraken2, family level): | Coronaviridae (62.92%)<br>Hominidae (0.08%)<br>Staphylococcaceae (0.06%) |

| | |
|---|---|
| | Bdellovibrionaceae (0.03%)<br>Mycobacteriaceae (0.01%) |

#### Detected variants (Experimental)

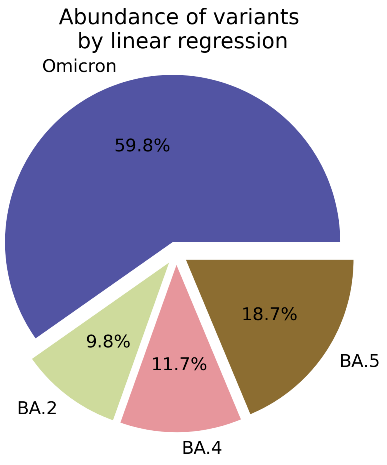


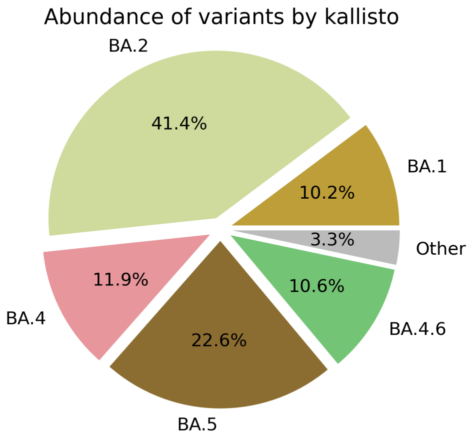


Based on deconvolution, B.1.1.529 is estimated to constitute 59.71% of the viral particles and hence is the most abundant variant in the sample. The $R^2$ for the linear regression was 0.60. Variants that were detected less than 5% were grouped under "Other"

Based on the consensus sequence of the observed reads, the "ensemble-averaged sequence" most closely resembles the BA.2 lineage. If this is a sample consisting of a single source of pathogens or an overwhelming majority of the different sources are infected with the same variant, the sample is dominated by this variant.

Based on mapping individual reads to the variant consensus sequences in the reference database, kallisto predicts that the sample is dominated by BA.2 lineage. Accuracy of this measure is expected to improve if the input data consists of long reads as opposed to convolution.

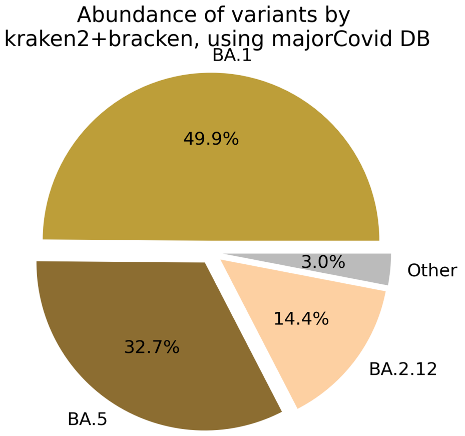


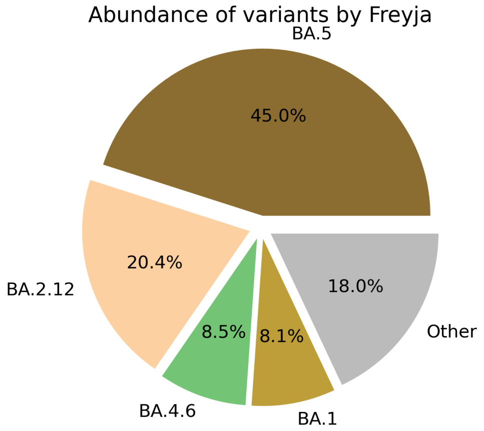

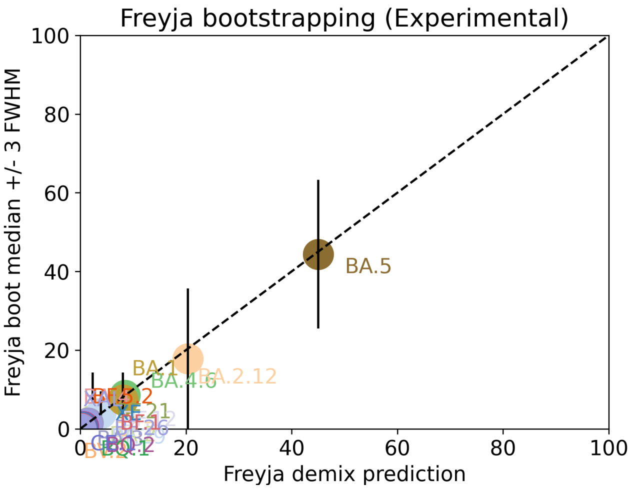


Under the assumption that the presence of a variant requires the detection of all respective mutations of the variant, the characteric mutations which support the presence of the respective variant are indicated in the respective column of the table. Numbers show the number of mutations detected, if any, and the number of mutations expected to be present based on the variant definitions.

| VOC | B.1.617.2 | BA.1 | BA.2 | BA.3 | BA.4 | BA.5 |
|---|---|---|---|---|---|---|
| Characteristic mutations detected | (4 of 13)<br>N:D377Y<br>S:G142D<br>S:L452R<br>S:T478K | (13 of 26)<br>NUC:C15240T<br>NUC:C25000T<br>NUC:C25584T<br>NUC:T13195C<br>NUC:T5386G<br>ORF1AB:A2710T<br>S:G446S<br>S:G496S<br>S:L981F<br>S:N856K<br>S:Q493R<br>S:T547K<br>S:T95I | (22 of 31)<br>N:S413R<br>NUC:A20055G<br>NUC:C10198T<br>NUC:C12880T<br>NUC:C15714T<br>NUC:C25000T<br>NUC:C25584T<br>NUC:C26858T<br>NUC:C4321T<br>NUC:G10447A<br>ORF1AB:G1307S<br>ORF1AB:L3027F<br>ORF1AB:L3201F<br>ORF1AB:S135R<br>ORF1AB:T3090I<br>ORF1AB:T842I<br>S:D405N<br>S:Q493R<br>S:R408S<br>S:S371F<br>S:T19I<br>S:T376A | (12 of 21)<br>N:S413R<br>NUC:C12880T<br>NUC:C15714T<br>NUC:C26858T<br>NUC:G10447A<br>ORF1AB:G1307S<br>ORF1AB:S135R<br>ORF1AB:T3090I<br>S:D405N<br>S:G446S<br>S:Q493R<br>S:S371F | (24 of 31)<br>N:P151S<br>N:S413R<br>NUC:A20055G<br>NUC:C10198T<br>NUC:C12880T<br>NUC:C15714T<br>NUC:C25000T<br>NUC:C25584T<br>NUC:C26858T<br>NUC:C4321T<br>NUC:G10447A<br>NUC:G12160A<br>NUC:G27788T<br>ORF1AB:G1307S<br>ORF1AB:S135R<br>ORF1AB:T3090I<br>ORF1AB:T842I<br>S:D405N<br>S:F486V<br>S:L452R<br>S:S371F<br>S:T19I<br>S:T376A<br>S:V213G | (21 of 28)<br>N:S413R<br>NUC:A20055G<br>NUC:C10198T<br>NUC:C12880T<br>NUC:C15714T<br>NUC:C25000T<br>NUC:C25584T<br>NUC:C4321T<br>NUC:G10447A<br>NUC:G12160A<br>ORF1AB:G1307S<br>ORF1AB:S135R<br>ORF1AB:T3090I<br>ORF1AB:T842I<br>S:D405N<br>S:F486V<br>S:L452R<br>S:S371F<br>S:T19I<br>S:T376A<br>S:V213G |

Jaccard Index is a measure of similarity between two sets A and B, reaching the maximum value of 1 if A=B and minimum value of 0 if A ∩ B = {}. In the c(d) representation below, c represents the Jaccard index of the set of mutations that were experimentally detected for this sample as listed above, whereas d refers to the ideal value of the Jaccard index expected from complete genome coverage without any sequencing errors.

| | B.1.617.2 | BA.1 | BA.2 | BA.3 | BA.4 | BA.5 |
|---|---|---|---|---|---|---|
| B.1.617.2 | 1.00 (1.00) | 0.00 (0.00) | 0.00 (0.00) | 0.00 (0.00) | 0.04 (0.02) | 0.04 (0.03) |
| BA.1 | 0.00 (0.00) | 1.00 (1.00) | 0.09 (0.10) | 0.09 (0.21) | 0.06 (0.08) | 0.06 (0.08) |
| BA.2 | 0.00 (0.00) | 0.09 (0.10) | 1.00 (1.00) | 0.48 (0.33) | 0.64 (0.63) | 0.65 (0.59) |
| BA.3 | 0.00 (0.00) | 0.09 (0.21) | 0.48 (0.33) | 1.00 (1.00) | 0.38 (0.30) | 0.38 (0.29) |

| | | | | | | |
|---|---|---|---|---|---|---|
| BA.4 | 0.04 (0.02) | 0.06 (0.08) | 0.64 (0.63) | 0.38 (0.30) | 1.00 (1.00) | 0.88 (0.84) |
| BA.5 | 0.04 (0.03) | 0.06 (0.08) | 0.65 (0.59) | 0.38 (0.29) | 0.88 (0.84) | 1.00 (1.00) |

#### Detected mutations

Excluded from this pdf version due to file size limitations.

U.S. FOOD & DRUG ADMINISTRATION

**CFSAN/OAO**
**BIOSTATISTICS AND BIOINFORMATICS STAFF**

### WASTEWATER SARS-COV2 ANALYSIS REPORT

| | |
|---|---|
| Sample name: | CFSANSMP000119629 |
| Date generated: | 2023-02-01, 20:05:19 EST |
| Timestamp of C-WAP version used: | Tue Jan 31 11:49:22 2023 -0500 |
| Executed by: | Jasmine Amirzadegan |
| Executed on: | 172.20.44.127 (aka n127.raven.cfsan) |

#### Sequencing summary

| | |
|---|---|
| Sequencing chemistry: | Missing with Missing |
| Source site: | Missing (?,?) |
| Sampling date: | Missing |
| Collected by: | Missing |
| Sequenced by: | Missing |
| Total number of reads: | 3144978 |
| Reads aligned: | 1455316 (46%) |
| Average read quality: | 36.6 |
| Average read length: | 149 |
| Reads passing filter: | 1390136 (44%) |
| Average read quality passing filter: | 36.9 |
| Average read length passing filter: | 149 |
| Average coverage passing filter: | 6926X |

A read passes filter if the read length after adaptor trimming ≥ 30 and minimum read quality ≥ 20 within a sliding window of width 4.

#### Overall sequence characteristics

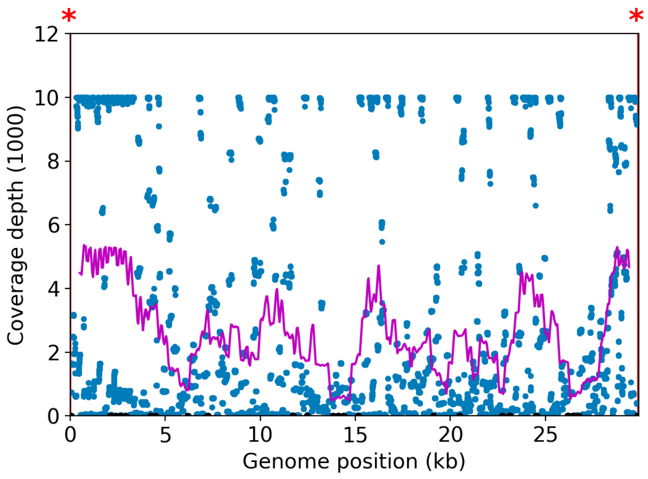


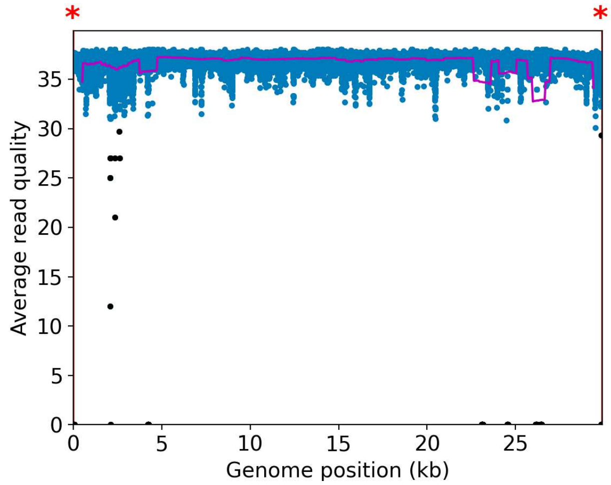


NOTE: The red shaded areas marked with a (*) are not covered by the design of the library preparation kit and hence excluded from analyses. Magenta curves represent moving average with a window width of 1kb.

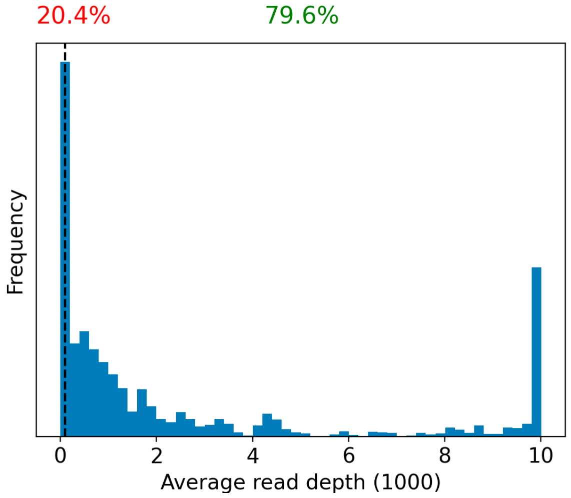


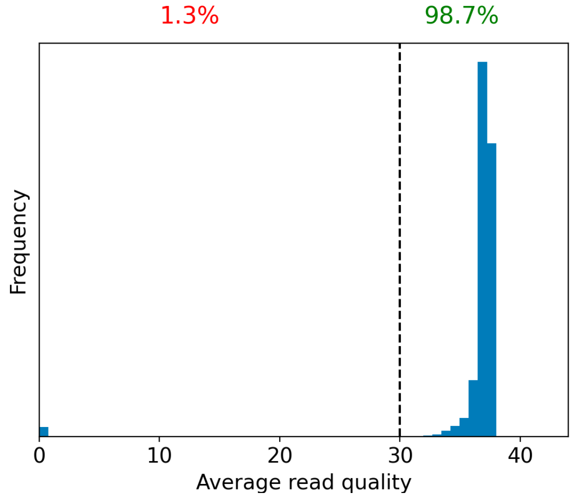


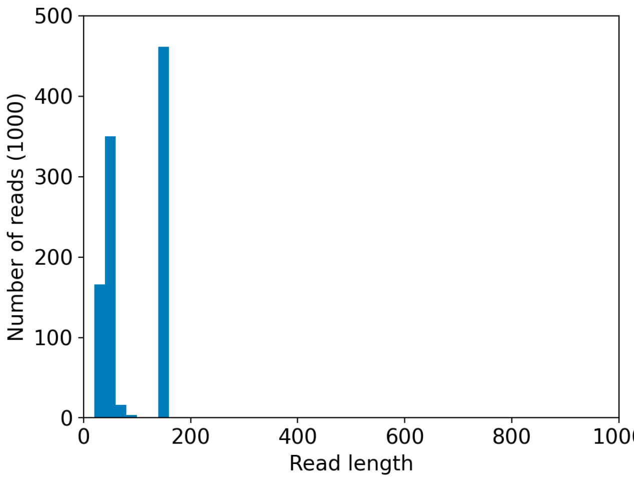


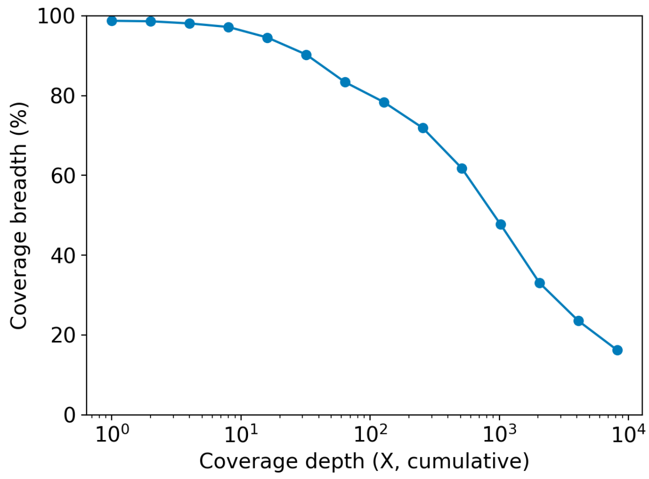

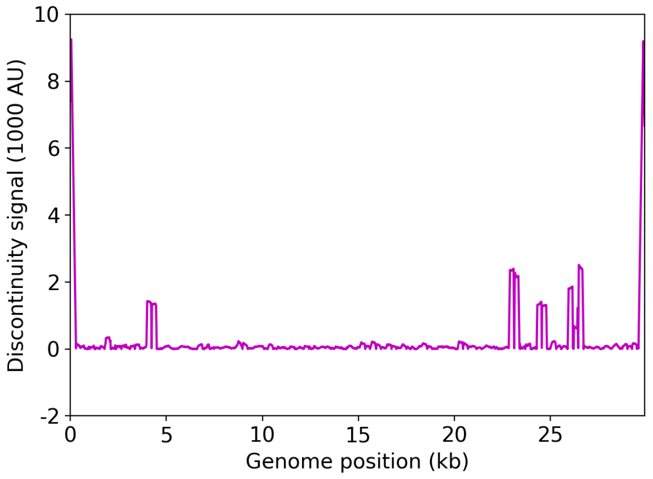


| | Uncovered coordinates (0X) | Poorly covered coordinates (<10X) |
|---|---|---|
| # Inaccessible genomic coordinates by kit design: | 121nt (0%) | 121nt (0%) |
| All genomic coordinates: | 369nt (1%) | 998nt (3%) |
| Common SNPs: | 0nt (0%) | 0nt (0%) |
| Diverse SNPs: | 29nt (12%) | 29nt (12%) |
| Rare SNPs: | 46nt (4%) | 87nt (9%) |

SNPs refer to the polymorphic sites currently in circulation that were detected out of recent GISAID entries. The sites that differ from the SC2 reference sequence are denoted as "common" if [90%, 100%] of the submissions carry this mutation, whereas those that are prevalent in [0%,10%] of the submissions are grouped under the "rare" category. The population is still diverse at the mutation sites that are observed in (10%,90%) of the entries and these coordinates are grouped under the "diverse" category.

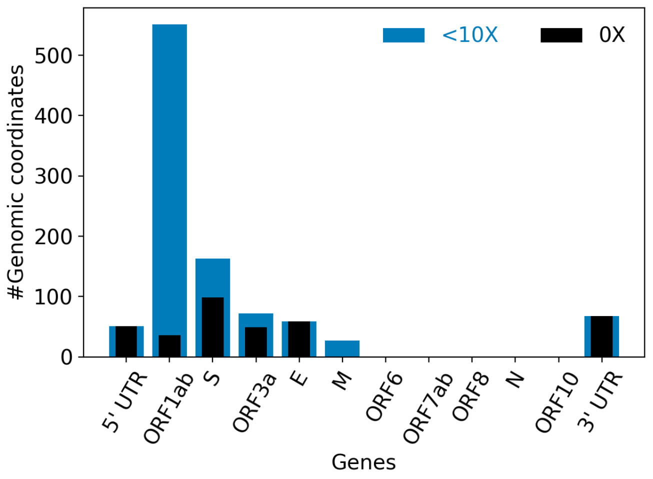


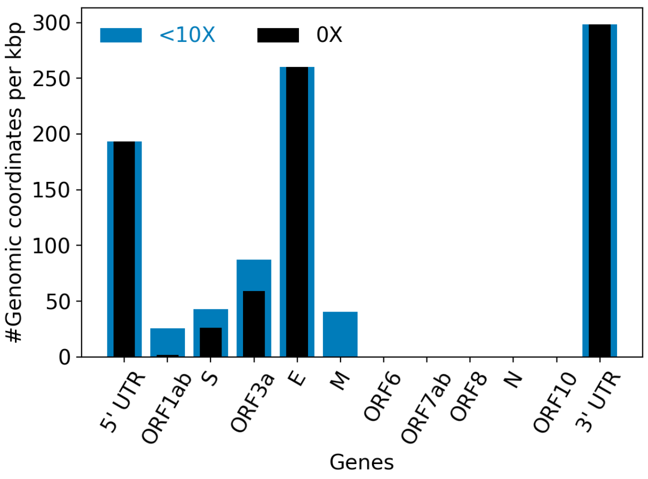


| | |
|---|---|
| Hits to SARS-Cov2 genome (kraken2): | 848972 reads (53.99%) |
| Hits to human genome (kraken2): | 1534 reads (0.10%) |
| Hits to synthetic sequences (kraken2, taxid 28384): | 2196 reads (0.14%) |
| Most abundant organisms (kraken2, family level): | Coronaviridae (53.99%)<br>Hominidae (0.10%)<br>Staphylococcaceae (0.09%) |

| | |
|---|---|
| | Arcobacteraceae (0.07%)<br>Weeksellaceae (0.04%) |

### Detected variants (Experimental)

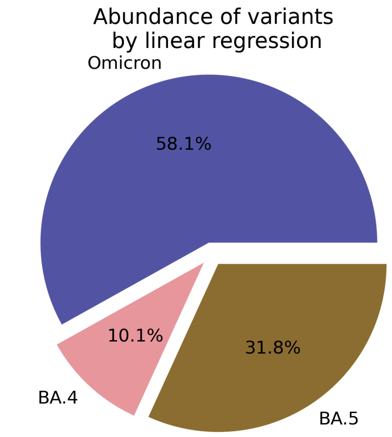


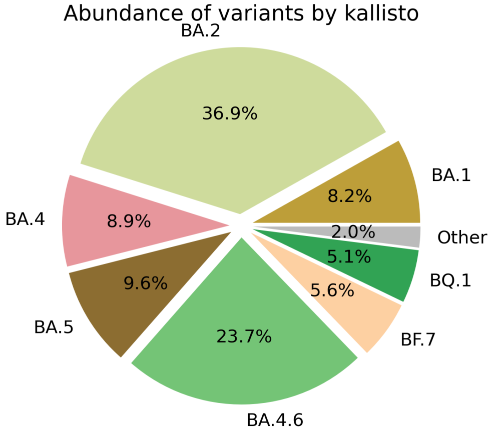


Based on deconvolution, B.1.1.529 is estimated to constitute 58.10% of the viral particles and hence is the most abundant variant in the sample. The $R^2$ for the linear regression was 0.61. Variants that were detected less than 5% were grouped under "Other"

Based on the consensus sequence of the observed reads, the "ensemble-averaged sequence" most closely resembles the BA.2 lineage. If this is a sample consisting of a single source of pathogens or an overwhelming majority of the different sources are infected with the same variant, the sample is dominated by this variant.

Based on mapping individual reads to the variant consensus sequences in the reference database, kallisto predicts that the sample is dominated by BA.2 lineage. Accuracy of this measure is expected to improve if the input data consists of long reads as opposed to convolution.

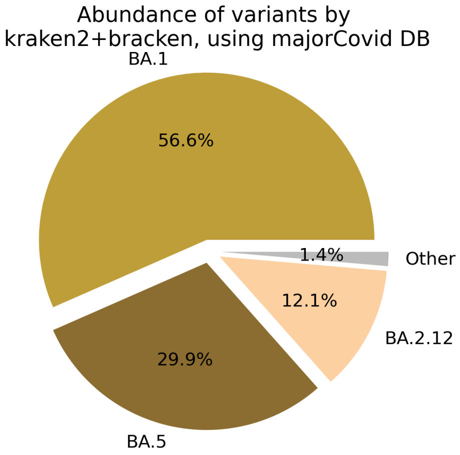


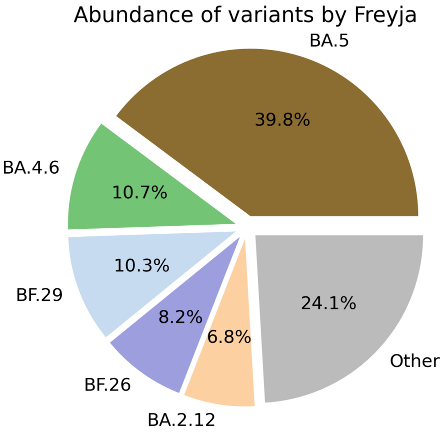

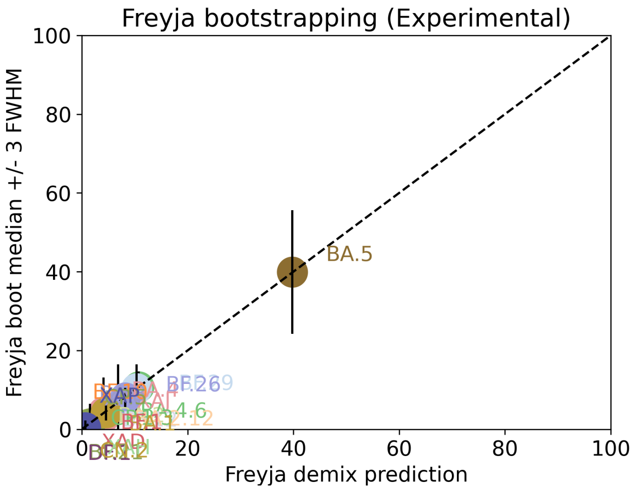


Under the assumption that the presence of a variant requires the detection of all respective mutations of the variant, the characteric mutations which support the presence of the respective variant are indicated in the respective column of the table. Numbers show the number of mutations detected, if any, and the number of mutations expected to be present based on the variant definitions.

| VOC | B.1.617.2 | BA.1 | BA.2 | BA.3 | BA.4 | BA.5 |
|---|---|---|---|---|---|---|
| Characteristic mutations detected | (4 of 13)<br>N:D377Y<br>S:G142D<br>S:L452R<br>S:T478K | (10 of 26)<br>NUC:C15240T<br>NUC:C25000T<br>NUC:C25584T<br>NUC:T13195C<br>ORF1AB:I3758V<br>ORF1AB:K856R<br>S:G446S<br>S:L981F<br>S:N856K<br>S:Q493R | (23 of 31)<br>N:S413R<br>NUC:A20055G<br>NUC:A9424G<br>NUC:C10198T<br>NUC:C12880T<br>NUC:C15714T<br>NUC:C25000T<br>NUC:C25584T<br>NUC:C26858T<br>NUC:C4321T<br>NUC:G10447A<br>ORF1AB:G1307S<br>ORF1AB:L3027F<br>ORF1AB:L3201F<br>ORF1AB:S135R<br>ORF1AB:T3090I<br>ORF1AB:T842I<br>S:D405N<br>S:Q493R<br>S:R408S<br>S:S371F<br>S:T19I<br>S:T376A | (12 of 21)<br>N:S413R<br>NUC:C12880T<br>NUC:C15714T<br>NUC:C26858T<br>NUC:G10447A<br>ORF1AB:G1307S<br>ORF1AB:S135R<br>ORF1AB:T3090I<br>S:D405N<br>S:G446S<br>S:Q493R<br>S:S371F | (24 of 31)<br>N:P151S<br>N:S413R<br>NUC:A20055G<br>NUC:C10198T<br>NUC:C12880T<br>NUC:C15714T<br>NUC:C25000T<br>NUC:C25584T<br>NUC:C26858T<br>NUC:C4321T<br>NUC:G10447A<br>NUC:G12160A<br>NUC:G27788T<br>ORF1AB:G1307S<br>ORF1AB:S135R<br>ORF1AB:T3090I<br>ORF1AB:T842I<br>S:D405N<br>S:F486V<br>S:L452R<br>S:S371F<br>S:T19I<br>S:T376A<br>S:V213G | (22 of 28)<br>M:D3N<br>N:S413R<br>NUC:A20055G<br>NUC:C10198T<br>NUC:C12880T<br>NUC:C15714T<br>NUC:C25000T<br>NUC:C25584T<br>NUC:C4321T<br>NUC:G10447A<br>NUC:G12160A<br>ORF1AB:G1307S<br>ORF1AB:S135R<br>ORF1AB:T3090I<br>ORF1AB:T842I<br>S:D405N<br>S:F486V<br>S:L452R<br>S:S371F<br>S:T19I<br>S:T376A<br>S:V213G |

Jaccard Index is a measure of similarity between two sets A and B, reaching the maximum value of 1 if A=B and minimum value of 0 if A ∩ B = {}. In the c(d) representation below, c represents the Jaccard index of the set of mutations that were experimentally detected for this sample as listed above, whereas d refers to the ideal value of the Jaccard index expected from complete genome coverage without any sequencing errors.

| | B.1.617.2 | BA.1 | BA.2 | BA.3 | BA.4 | BA.5 |
|---|---|---|---|---|---|---|
| B.1.617.2 | 1.00 (1.00) | 0.00 (0.00) | 0.00 (0.00) | 0.00 (0.00) | 0.04 (0.02) | 0.04 (0.03) |
| BA.1 | 0.00 (0.00) | 1.00 (1.00) | 0.10 (0.10) | 0.10 (0.21) | 0.06 (0.08) | 0.07 (0.08) |
| BA.2 | 0.00 (0.00) | 0.10 (0.10) | 1.00 (1.00) | 0.46 (0.33) | 0.62 (0.63) | 0.61 (0.59) |
| BA.3 | 0.00 (0.00) | 0.10 (0.21) | 0.46 (0.33) | 1.00 (1.00) | 0.38 (0.30) | 0.36 (0.29) |

| | | | | | | |
|---|---|---|---|---|---|---|
| BA.4 | 0.04 (0.02) | 0.06 (0.08) | 0.62 (0.63) | 0.38 (0.30) | 1.00 (1.00) | 0.84 (0.84) |
| BA.5 | 0.04 (0.03) | 0.07 (0.08) | 0.61 (0.59) | 0.36 (0.29) | 0.84 (0.84) | 1.00 (1.00) |

#### Detected mutations

Excluded from this pdf version due to file size limitations.

CFSAN/OAO
BIOSTATISTICS AND BIOINFORMATICS STAFF

### WASTEWATER SARS-COV2 ANALYSIS REPORT

| | |
|---|---|
| Sample name: | Undetermined |
| Date generated: | 2023-02-01, 20:11:09 EST |
| Timestamp of C-WAP version used: | Tue Jan 31 11:49:22 2023 -0500 |
| Executed by: | Jasmine Amirzadegan |
| Executed on: | 172.20.44.108 (aka n108.raven.cfsan) |

#### Sequencing summary

| | |
|---|---|
| Sequencing chemistry: | Missing with Missing |
| Source site: | Missing (?,?) |
| Sampling date: | Missing |
| Collected by: | Missing |
| Sequenced by: | Missing |
| Total number of reads: | 4361554 |
| Reads aligned: | 2917865 (66%) |
| Average read quality: | 36.1 |
| Average read length: | 149 |
| Reads passing filter: | 2691770 (61%) |
| Average read quality passing filter: | 36.7 |
| Average read length passing filter: | 149 |
| Average coverage passing filter: | 13412X |

A read passes filter if the read length after adaptor trimming ≥ 30 and minimum read quality ≥ 20 within a sliding window of width 4.

#### Overall sequence characteristics

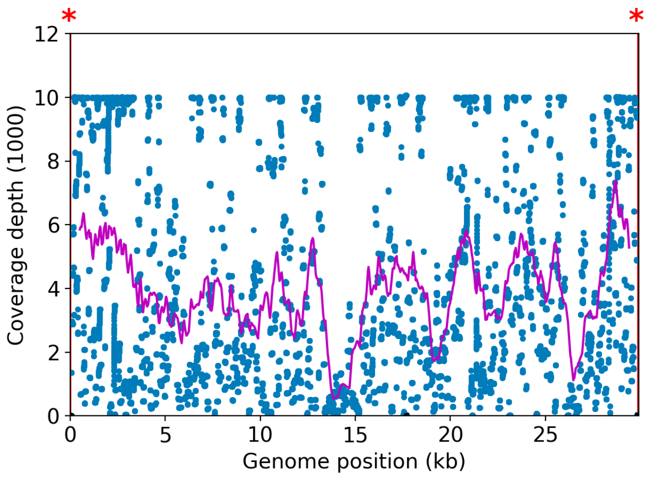


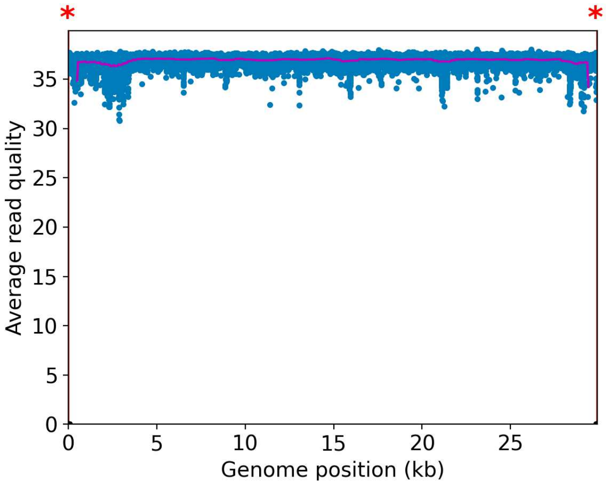


NOTE: The red shaded areas marked with a (*) are not covered by the design of the library preparation kit and hence excluded from analyses. Magenta curves represent moving average with a window width of 1kb.

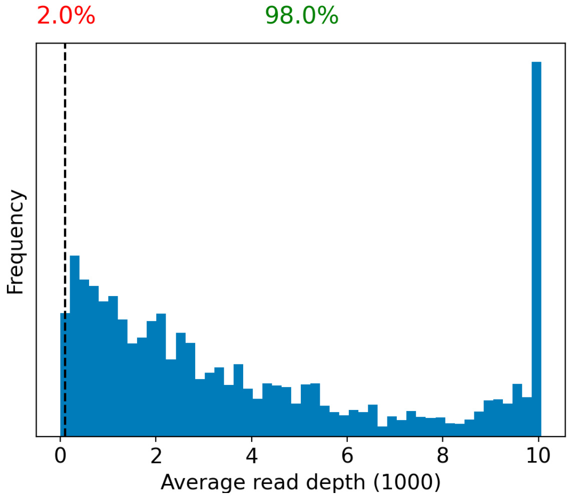


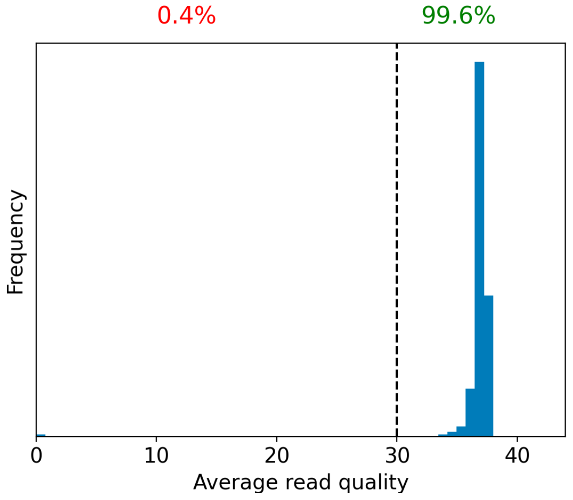


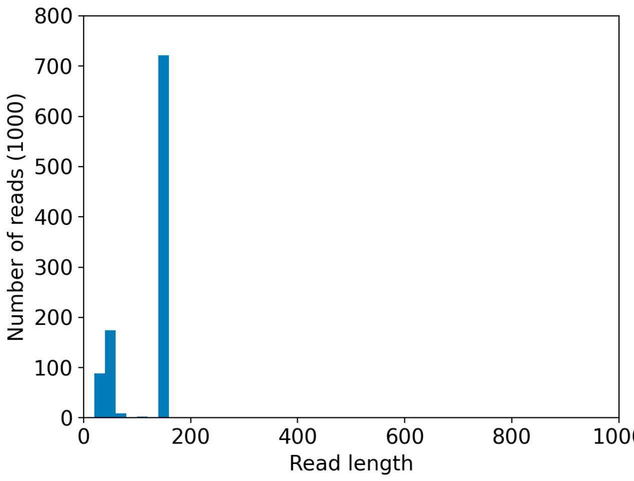


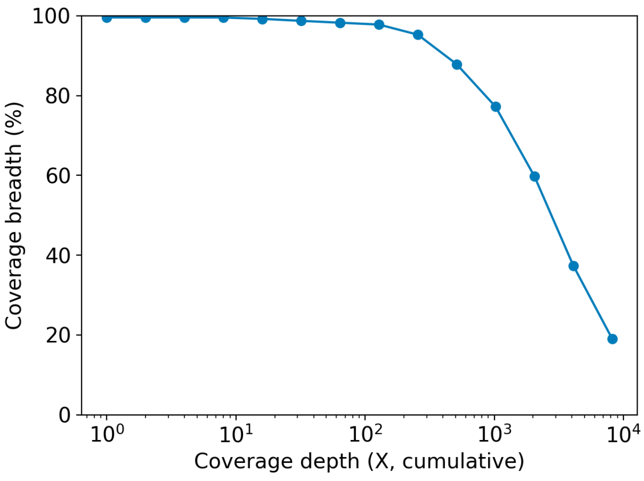

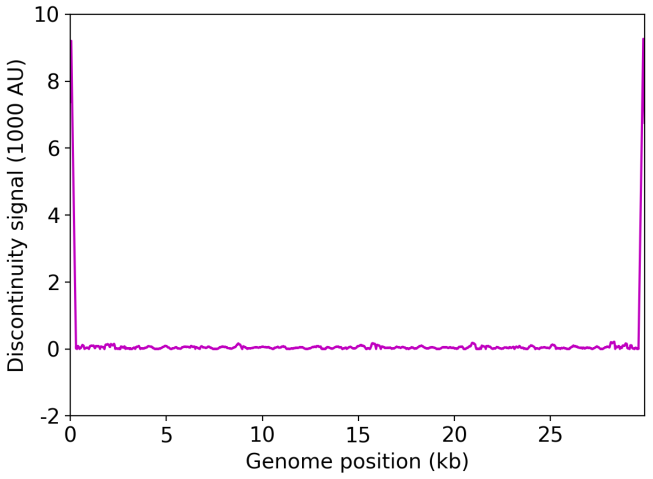


| | Uncovered coordinates (0X) | Poorly covered coordinates (<10X) |
|---|---|---|
| # Inaccessible genomic coordinates by kit design: | 121nt (0%) | 121nt (0%) |
| All genomic coordinates: | 119nt (0%) | 126nt (0%) |
| Common SNPs: | 0nt (0%) | 0nt (0%) |
| Diverse SNPs: | 29nt (12%) | 29nt (12%) |
| Rare SNPs: | 10nt (1%) | 10nt (1%) |

SNPs refer to the polymorphic sites currently in circulation that were detected out of recent GISAID entries. The sites that differ from the SC2 reference sequence are denoted as "common" if [90%, 100%] of the submissions carry this mutation, whereas those that are prevalent in [0%,10%] of the submissions are grouped under the "rare" category. The population is still diverse at the mutation sites that are observed in (10%,90%) of the entries and these coordinates are grouped under the "diverse" category.

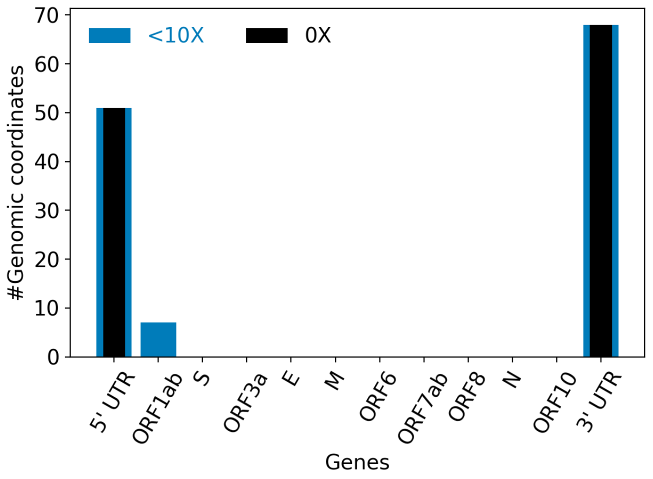


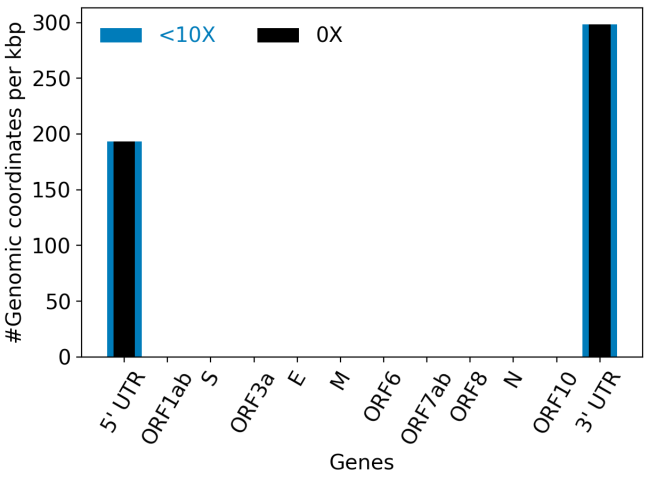


| | |
|---|---|
| Hits to SARS-Cov2 genome (kraken2): | 1574189 reads (72.18%) |
| Hits to human genome (kraken2): | 33636 reads (1.54%) |
| Hits to synthetic sequences (kraken2, taxid 28384): | 1667 reads (0.08%) |
| Most abundant organisms (kraken2, family level): | Coronaviridae (72.18%)<br>Hominidae (1.54%)<br>Staphylococcaceae (0.25%) |

| | |
|---|---|
| | Enterobacteriaceae (0.08%)<br>Enterococcaceae (0.06%) |

### Detected variants (Experimental)

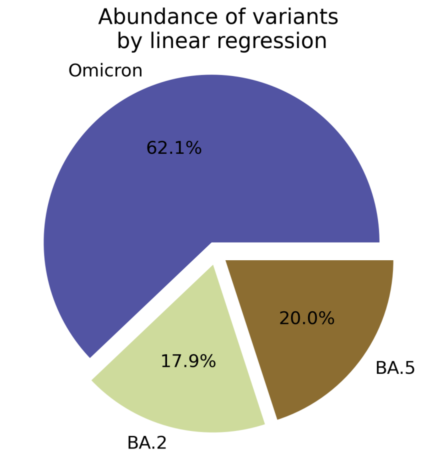


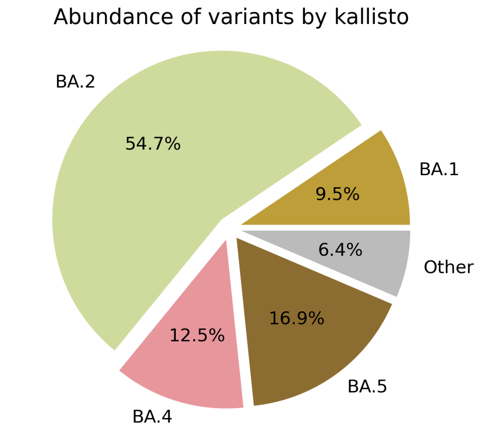


Based on deconvolution, B.1.1.529 is estimated to constitute 62.10% of the viral particles and hence is the most abundant variant in the sample. The $R^2$ for the linear regression was 0.63. Variants that were detected less than 5% were grouped under "Other"

Based on the consensus sequence of the observed reads, the "ensemble-averaged sequence" most closely resembles the BA.2 lineage. If this is a sample consisting of a single source of pathogens or an overwhelming majority of the different sources are infected with the same variant, the sample is dominated by this variant.

Based on mapping individual reads to the variant consensus sequences in the reference database, kallisto predicts that the sample is dominated by BA.2 lineage. Accuracy of this measure is expected to improve if the input data consists of long reads as opposed to convolution.

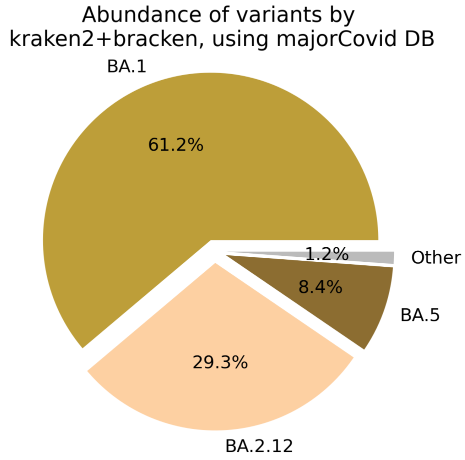


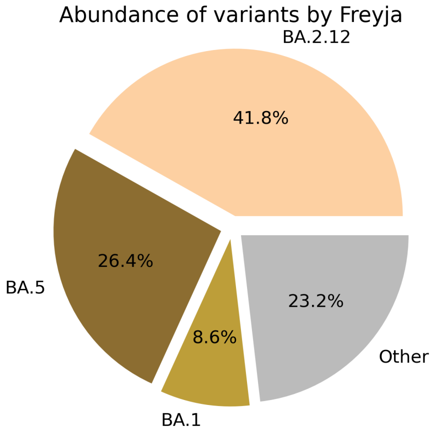

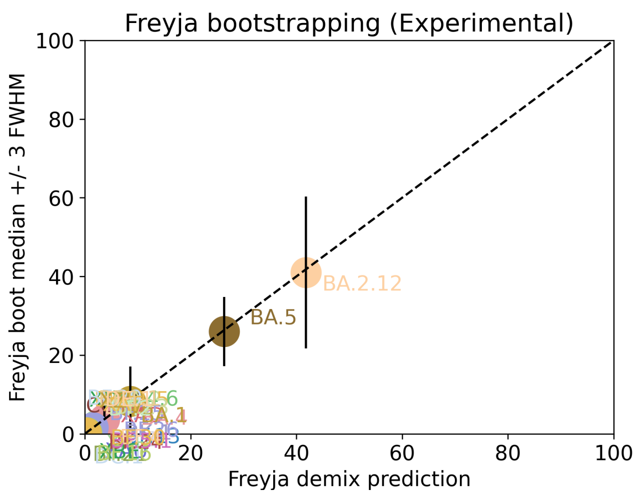


Under the assumption that the presence of a variant requires the detection of all respective mutations of the variant, the characteric mutations which support the presence of the respective variant are indicated in the respective column of the table. Numbers show the number of mutations detected, if any, and the number of mutations expected to be present based on the variant definitions.

| VOC | B.1.617.2 | BA.1 | BA.2 | BA.3 | BA.4 | BA.5 |
|---|---|---|---|---|---|---|
| Characteristic mutations detected | (4 of 13)<br>N:D377Y<br>S:G142D<br>S:L452R<br>S:T478K | (16 of 26)<br>NUC:C15240T<br>NUC:C25000T<br>NUC:C25584T<br>NUC:T13195C<br>NUC:T5386G<br>ORF1AB:A2710T<br>ORF1AB:I3758V<br>ORF1AB:K856R<br>S:A67V<br>S:G446S<br>S:G496S<br>S:L981F<br>S:N856K<br>S:Q493R<br>S:T547K<br>S:T95I | (23 of 31)<br>N:S413R<br>NUC:A20055G<br>NUC:A9424G<br>NUC:C10198T<br>NUC:C12880T<br>NUC:C15714T<br>NUC:C25000T<br>NUC:C25584T<br>NUC:C26858T<br>NUC:C4321T<br>NUC:G10447A<br>ORF1AB:G1307S<br>ORF1AB:L3027F<br>ORF1AB:L3201F<br>ORF1AB:S135R<br>ORF1AB:T3090I<br>ORF1AB:T842I<br>S:D405N<br>S:Q493R<br>S:R408S<br>S:S371F<br>S:T19I<br>S:T376A | (13 of 21)<br>N:S413R<br>NUC:C12880T<br>NUC:C15714T<br>NUC:C26858T<br>NUC:G10447A<br>ORF1AB:G1307S<br>ORF1AB:S135R<br>ORF1AB:T3090I<br>S:A67V<br>S:D405N<br>S:G446S<br>S:Q493R<br>S:S371F | (24 of 31)<br>N:P151S<br>N:S413R<br>NUC:A20055G<br>NUC:C10198T<br>NUC:C12880T<br>NUC:C15714T<br>NUC:C25000T<br>NUC:C25584T<br>NUC:C26858T<br>NUC:C4321T<br>NUC:G10447A<br>NUC:G12160A<br>NUC:G27788T<br>ORF1AB:G1307S<br>ORF1AB:S135R<br>ORF1AB:T3090I<br>ORF1AB:T842I<br>S:D405N<br>S:F486V<br>S:L452R<br>S:S371F<br>S:T19I<br>S:T376A<br>S:V213G | (22 of 28)<br>M:D3N<br>N:S413R<br>NUC:A20055G<br>NUC:C10198T<br>NUC:C12880T<br>NUC:C15714T<br>NUC:C25000T<br>NUC:C25584T<br>NUC:C4321T<br>NUC:G10447A<br>NUC:G12160A<br>ORF1AB:G1307S<br>ORF1AB:S135R<br>ORF1AB:T3090I<br>ORF1AB:T842I<br>S:D405N<br>S:F486V<br>S:L452R<br>S:S371F<br>S:T19I<br>S:T376A<br>S:V213G |

Jaccard Index is a measure of similarity between two sets A and B, reaching the maximum value of 1 if A=B and minimum value of 0 if A ∩ B = {}. In the c(d) representation below, c represents the Jaccard index of the set of mutations that were experimentally detected for this sample as listed above, whereas d refers to the ideal value of the Jaccard index expected from complete genome coverage without any sequencing errors.

| | B.1.617.2 | BA.1 | BA.2 | BA.3 | BA.4 | BA.5 |
|---|---|---|---|---|---|---|
| B.1.617.2 | 1.00 (1.00) | 0.00 (0.00) | 0.00 (0.00) | 0.00 (0.00) | 0.04 (0.02) | 0.04 (0.03) |
| BA.1 | 0.00 (0.00) | 1.00 (1.00) | 0.08 (0.10) | 0.12 (0.21) | 0.05 (0.08) | 0.06 (0.08) |
| BA.2 | 0.00 (0.00) | 0.08 (0.10) | 1.00 (1.00) | 0.44 (0.33) | 0.62 (0.63) | 0.61 (0.59) |
| BA.3 | 0.00 (0.00) | 0.12 (0.21) | 0.44 (0.33) | 1.00 (1.00) | 0.37 (0.30) | 0.35 (0.29) |

| | | | | | | |
|---|---|---|---|---|---|---|
| BA.4 | 0.04 (0.02) | 0.05 (0.08) | 0.62 (0.63) | 0.37 (0.30) | 1.00 (1.00) | 0.84 (0.84) |
| BA.5 | 0.04 (0.03) | 0.06 (0.08) | 0.61 (0.59) | 0.35 (0.29) | 0.84 (0.84) | 1.00 (1.00) |

#### Detected mutations

Excluded from this pdf version due to file size limitations.

CFSAN/OAO
BIOSTATISTICS AND BIOINFORMATICS STAFF

### WASTEWATER SARS-COV2 ANALYSIS REPORT

| | |
|---|---|
| Sample name: | Water |
| Date generated: | 2023-02-01, 19:42:14 EST |
| Timestamp of C-WAP version used: | Tue Jan 31 11:49:22 2023 -0500 |
| Executed by: | Jasmine Amirzadegan |
| Executed on: | 172.20.44.134 (aka n134.raven.cfsan) |

#### Sequencing summary

| | |
|---|---|
| Sequencing chemistry: | Missing with Missing |
| Source site: | Missing (?,?) |
| Sampling date: | Missing |
| Collected by: | Missing |
| Sequenced by: | Missing |
| Total number of reads: | 45196 |
| Reads aligned: | 1655 (3%) |
| Average read quality: | 36.5 |
| Average read length: | 144 |
| Reads passing filter: | 1515 (3%) |
| Average read quality passing filter: | 36.9 |
| Average read length passing filter: | 148 |
| Average coverage passing filter: | 7X |

A read passes filter if the read length after adaptor trimming ≥ 30 and minimum read quality ≥ 20 within a sliding window of width 4.

#### Overall sequence characteristics

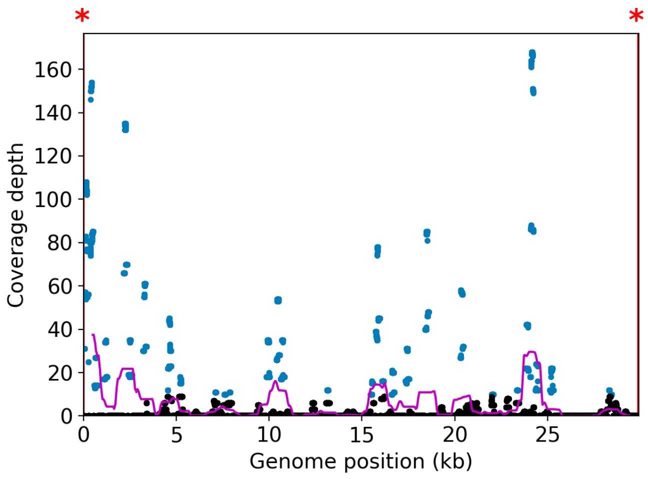


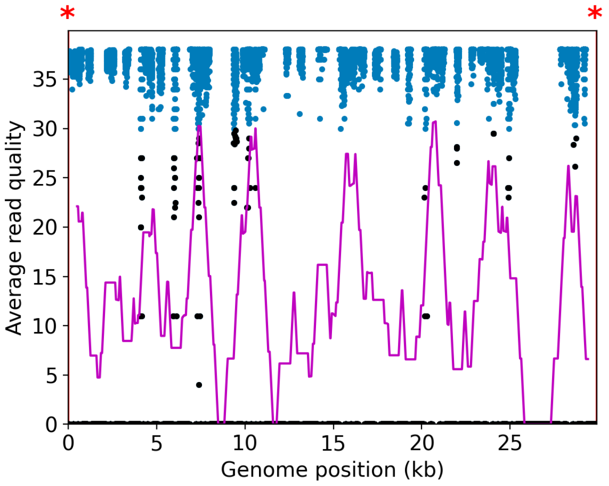


NOTE: The red shaded areas marked with a (*) are not covered by the design of the library preparation kit and hence excluded from analyses. Magenta curves represent moving average with a window width of 1kb.

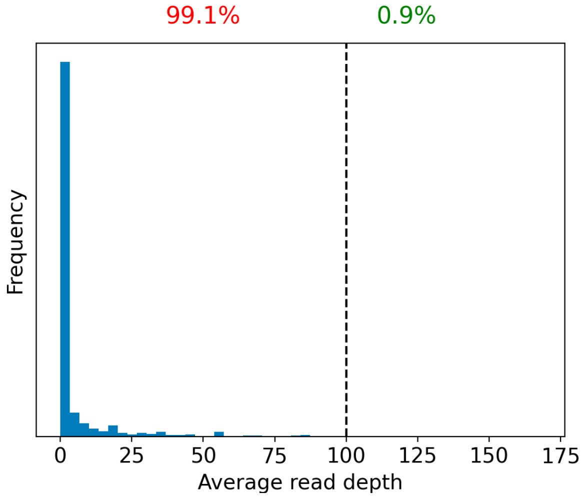


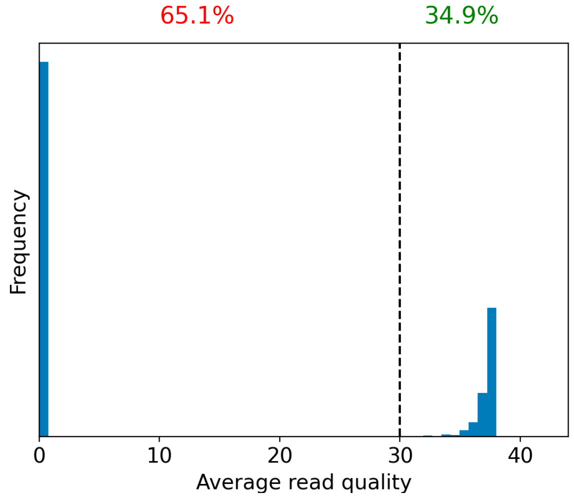


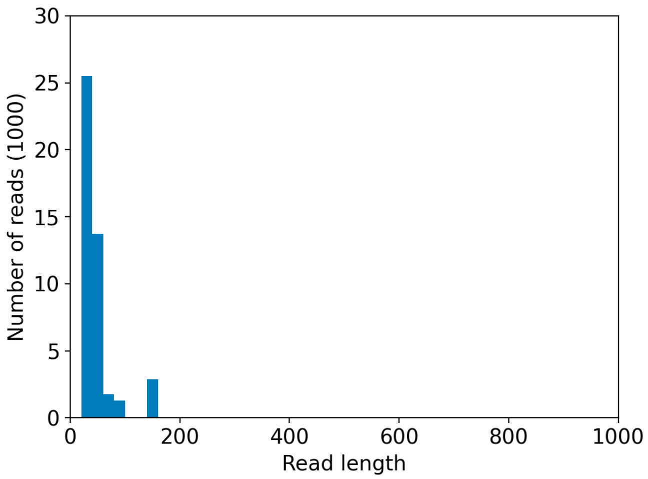


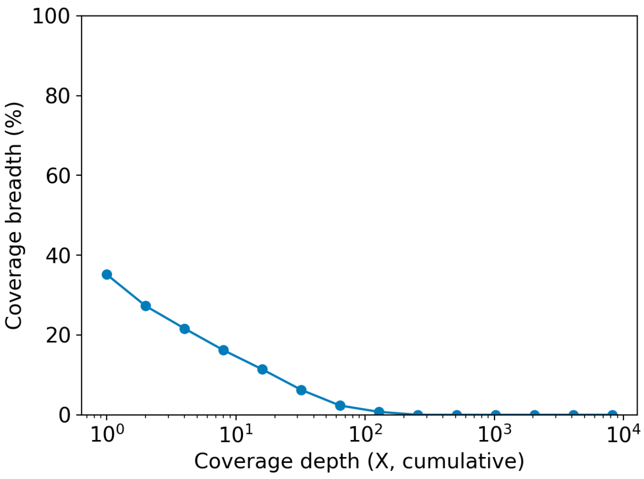

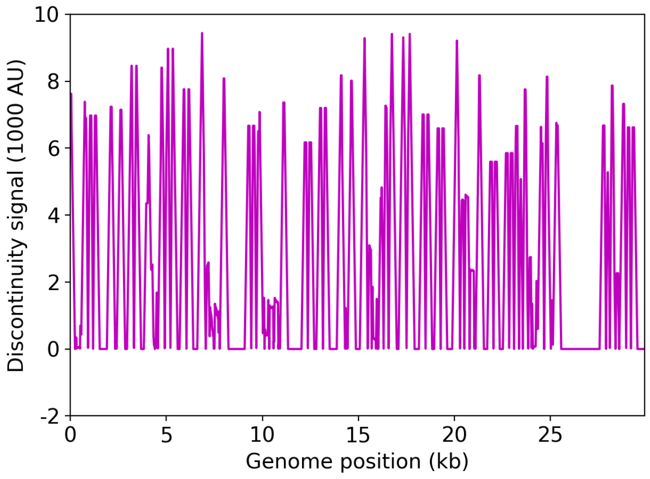


WARNING: The sequence coverage is very low (7X)

| | **Uncovered coordinates (0X)** | **Poorly covered coordinates (<10X)** |
|---|---|---|
| # Inaccessible genomic coordinates by kit design: | 121nt (0%) | 121nt (0%) |
| All genomic coordinates: | 19387nt (64%) | 25466nt (85%) |
| Common SNPs: | 22nt (61%) | 27nt (75%) |
| Diverse SNPs: | 194nt (86%) | 218nt (96%) |
| Rare SNPs: | 603nt (65%) | 862nt (93%) |

SNPs refer to the polymorphic sites currently in circulation that were detected out of recent GISAID entries. The sites that differ from the SC2 reference sequence are denoted as "common" if [90%, 100%] of the submissions carry this mutation, whereas those that are prevalent in [0%,10%] of the submissions are grouped under the "rare" category. The population is still diverse at the mutation sites that are observed in (10%,90%) of the entries and these coordinates are grouped under the "diverse" category.

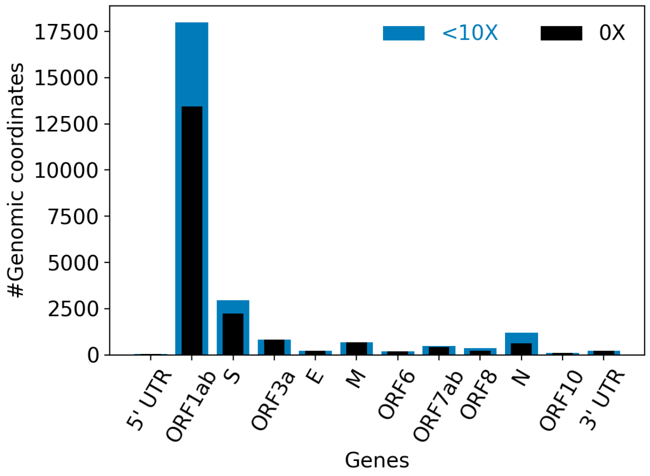


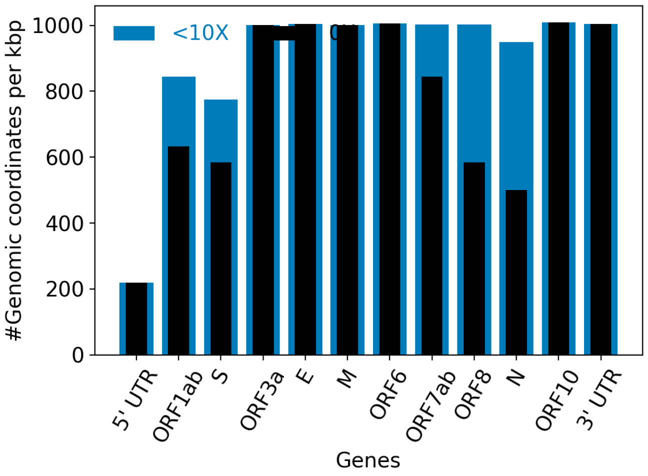


| | |
|---|---|
| Hits to SARS-Cov2 genome (kraken2): | 4973 reads (22.01%) |
| Hits to human genome (kraken2): | 41 reads (0.18%) |
| Hits to synthetic sequences (kraken2, taxid 28384): | 7 reads (0.03%) |
| | Coronaviridae (22.01%)<br>Hominidae (0.18%) |

| | |
|---|---|
| Most abundant organisms (kraken2, family level): | Mycobacteriaceae (0.12%)<br>Staphylococcaceae (0.11%)<br>Enterobacteriaceae (0.06%) |

#### Detected variants (Experimental)

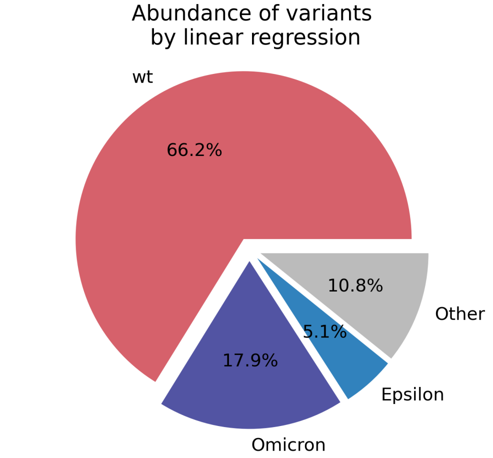


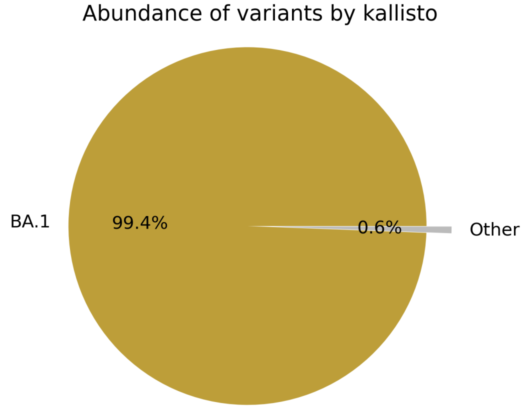


Based on deconvolution, wt is estimated to constitute 65.94% of the viral particles and hence is the most abundant variant in the sample. The $R^2$ for the linear regression was 0.13. Variants that were detected less than 5% were grouped under "Other"

Based on the consensus sequence of the observed reads, the "ensemble-averaged sequence" most closely resembles the B.1 lineage. If this is a sample consisting of a single source of pathogens or an overwhelming majority of the different sources are infected with the same variant, the sample is dominated by this variant.

Based on mapping individual reads to the variant consensus sequences in the reference database, kallisto predicts that the sample is dominated by BA.1 lineage. Accuracy of this measure is expected to improve if the input data consists of long reads as opposed to convolution.

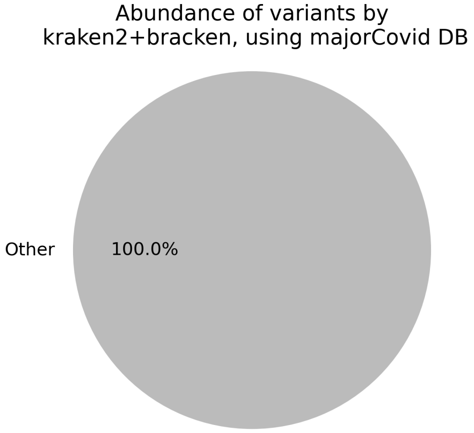


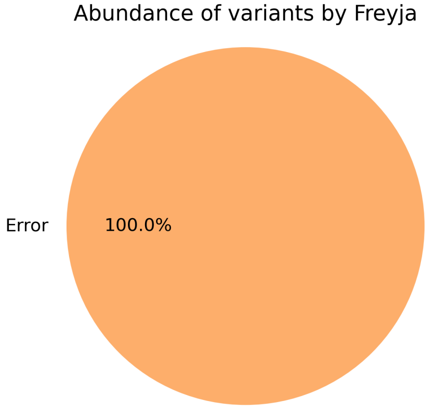


Freyja bootstrapping

Under the assumption that the presence of a variant requires the detection of all respective mutations of the variant, the characteric mutations which support the presence of the respective variant are indicated in the respective column of the table. Numbers show the number of mutations detected, if any, and the number of mutations expected to be present based on the variant definitions.

| VOC | B.1.617.2 | BA.1 | BA.2 | BA.3 | BA.4 | BA.5 |
|---|---|---|---|---|---|---|
| Characteristic mutations detected | (0 of 13) | (1 of 26) S:L981F | (0 of 31) | (0 of 21) | (0 of 31) | (0 of 28) |

Jaccard Index is a measure of similarity between two sets A and B, reaching the maximum value of 1 if A=B and minimum value of 0 if A ∩ B = {}. In the c(d) representation below, c represents the Jaccard index of the set of mutations that were experimentally detected for this sample as listed above, whereas d refers to the ideal value of the Jaccard index expected from complete genome coverage without any sequencing errors.

| | B.1.617.2 | BA.1 | BA.2 | BA.3 | BA.4 | BA.5 |
|---|---|---|---|---|---|---|
| B.1.617.2 | -1.00 (1.00) | 0.00 (0.00) | -1.00 (0.00) | -1.00 (0.00) | -1.00 (0.02) | -1.00 (0.03) |
| BA.1 | 0.00 (0.00) | 1.00 (1.00) | 0.00 (0.10) | 0.00 (0.21) | 0.00 (0.08) | 0.00 (0.08) |
| BA.2 | -1.00 (0.00) | 0.00 (0.10) | -1.00 (1.00) | -1.00 (0.33) | -1.00 (0.63) | -1.00 (0.59) |
| BA.3 | -1.00 (0.00) | 0.00 (0.21) | -1.00 (0.33) | -1.00 (1.00) | -1.00 (0.30) | -1.00 (0.29) |
| BA.4 | -1.00 (0.02) | 0.00 (0.08) | -1.00 (0.63) | -1.00 (0.30) | -1.00 (1.00) | -1.00 (0.84) |
| BA.5 | -1.00 (0.03) | 0.00 (0.08) | -1.00 (0.59) | -1.00 (0.29) | -1.00 (0.84) | -1.00 (1.00) |

#### Detected mutations

Excluded from this pdf version due to file size limitations.
