## Supplementary figures and images for "Harnessing methods, data analysis, and near-real-time wastewater monitoring for enhanced public health response using high throughput sequencing"

### Supplementary figure 1

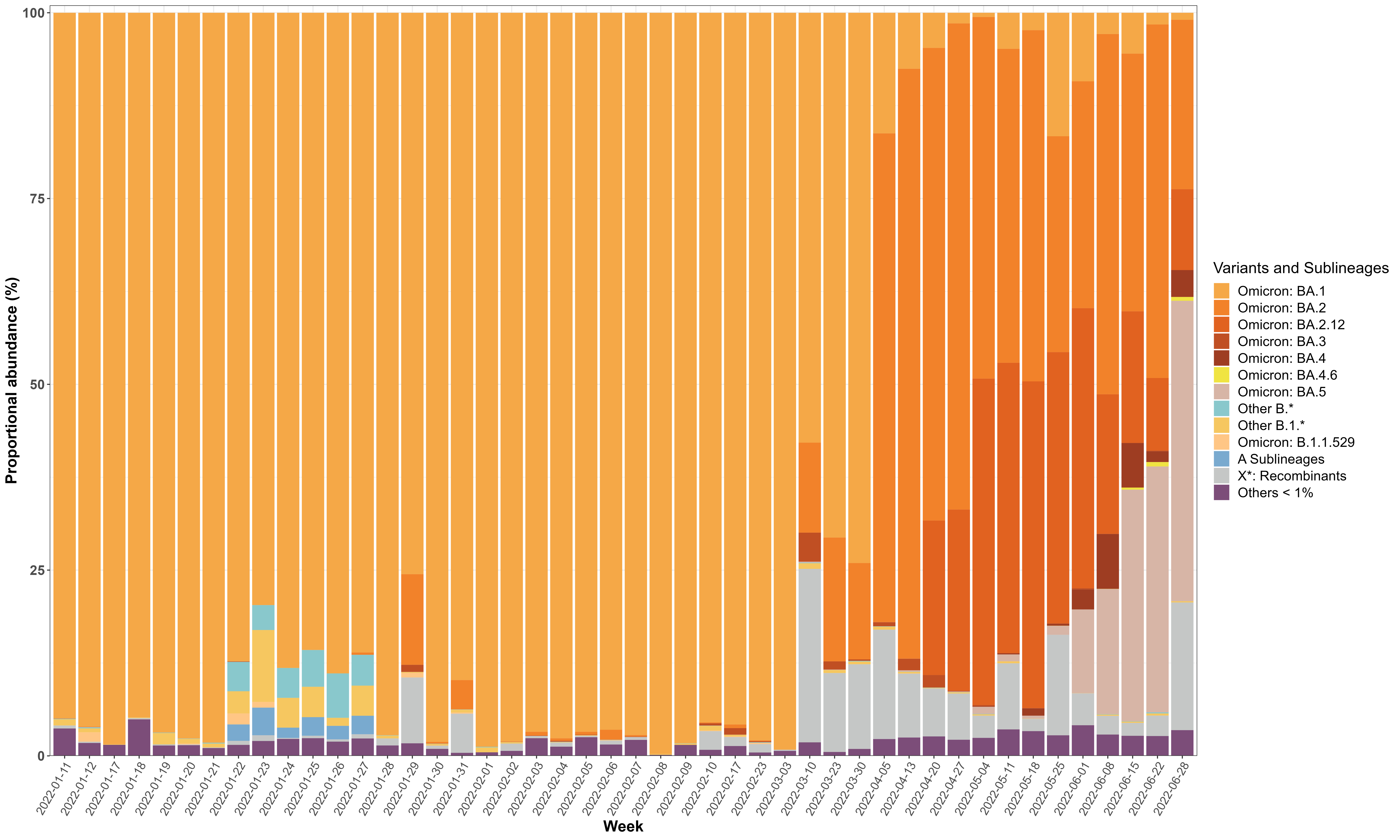
